# Excess years of life lost to COVID-19 and other causes of death by sex, neighbourhood deprivation and region in England & Wales during 2020

**DOI:** 10.1101/2021.07.05.21259786

**Authors:** Evangelos Kontopantelis, Mamas A. Mamas, Roger T. Webb, Ana Castro-Avila, Martin K. Rutter, Chris P. Gale, Darren M. Ashcroft, Matthias Pierce, Kathryn M. Abel, Gareth Price, Corinne Faivre-Finn, Harriette G.C. Van Spall, Michelle M. Graham, Marcello Morciano, Glen P. Martin, Matt Sutton, Tim Doran

## Abstract

**Background:** Deaths in the first year of the COVID-19 pandemic in England & Wales have been shown to be unevenly distributed socioeconomically and geographically. However, the full scale of inequalities may have been underestimated as most measures of excess mortality do not adequately account for varying age profiles of deaths between social groups. We measured years of life lost (YLL) attributable to the pandemic, directly or indirectly, comparing mortality across geographic and socioeconomic groups.

**Methods:** YLL for registered deaths in England & Wales, from 27^th^ December 2014 until 25^th^ December 2020, were calculated using 2019 single year sex-specific life tables for England & Wales. Panel time-series models were used to estimate expected YLL by sex, geographical region, and deprivation quintile between 7^th^ March 2020 and 25^th^ December 2020 by cause: direct deaths (COVID-19 and other respiratory diseases), cardiovascular disease & diabetes, cancer, and other indirect deaths - all other causes). Excess YLL during the pandemic period were calculated by subtracting observed from expected values. Additional analyses focused on excess deaths for region and deprivation strata, by age-group.

**Findings:** Between 7^th^ March 2020 and 25^th^ December 2020 there were an estimated 763,550 (95% CI: 696,826 to 830,273) excess YLL in England & Wales, equivalent to a 15% (95% CI: 14 to 16) increase in YLL compared to the equivalent time period in 2019. There was a strong deprivation gradient in all-cause excess YLL, with rates per 100,000 population ranging from (916; 95% CI: 820 to 1,012) for the least deprived quintile to (1,645; 95% CI: 1,472 to 1,819) for the most deprived. The differences in excess YLL between deprivation quintiles were greatest in younger age groups; for all-cause deaths, an average of 9.1 years per death (95% CI: 8.2 to 10.0) were lost in the least deprived quintile, compared to 10.8 (95% CI: 10.0 to 11.6) in the most deprived; for COVID-19 and other respiratory deaths, an average of 8.9 years per death (95% CI: 8.7 to 9.1) were lost in the least deprived quintile, compared to 11.2 (95% CI: 11.0 to 11.5) in the most deprived. There was marked variability in both all-cause and direct excess YLL by region, with the highest rates in both in the North West.

**Interpretation:** During 2020, the first calendar year of the COVID-19 pandemic, longstanding socioeconomic and geographical health inequalities in England & Wales were exacerbated, with the most deprived areas suffering the greatest losses in potential years of life lost.

**Funding:** None

## Introduction

Estimating excess deaths, by comparing observed deaths with the number expected based on historical trends, is an informative way of quantifying the potential impact of a pandemic on different population groups. There were an estimated 126,658 excess deaths in England in the first year of the COVID-19 pandemic,^1^ and these deaths were unevenly distributed both socioeconomically and geographically. Socioeconomic factors underly much of the geographic variation, mediating their effects through population health status, living and work circumstances, environmental conditions, and infection prevention and mitigation measures. Although all English regions and all social groups have been detrimentally affected by the pandemic, age-standardised rates in the first wave were 67% higher in the West Midlands than in the South West, and 33% higher in the most deprived than the most affluent quintile.^2^

The count of all-cause excess deaths includes those directly attributable to the virus and additional indirect deaths resulting from public health measures and the wider societal response, which for some causes will be lower than predicted. In the early part of the current pandemic, a quarter of deaths were attributable to causes other than COVID-19 infection, in particular cardiovascular disease and dementia.^1^ These indirect excess deaths followed different regional and socioeconomic patterns depending on underlying cause,^2^ and resulted from changes in primary and secondary care capacity,^3 4^ and in social behaviour, including decisions on seeking care.^5 6^

The risk of dying following infection with COVID-19 increased sharply with age.^7^ However, susceptibility in younger age groups may have also varied across social groups, giving rise to inequalities that are not adequately captured by a simple count of deaths. Additionally, indirect deaths are unlikely to follow the same age-related patterns as deaths directly attributable to COVID-19, and may result in substantial numbers of deaths in younger age groups. Years of life lost (YLL) is an alternative measure of premature mortality that allows for comparisons between causes of death and across population groups, because it accounts for both the number of deaths and the age at which those deaths occurred (being higher for deaths that occur at a younger age). In this study we measure YLL attributable to the pandemic by cause of death, comparing mortality across genders and geographic and socioeconomic groups in England & Wales.

## Methods

### Data

Deaths registered in England & Wales from 27^th^ December 2014 until 1^st^ March 2021 were accessed using the secure Amazon Workspaces environment setup by NHS Digital, which is the national provider of information, data and IT systems within England’s National Health Service (NHS). To minimise the effect of delayed death registration, the observation period was from the 1^st^ week of 2015 (ending 2^nd^ January) to the last week of 2020 (ending 25^th^ December). For each death, the underlying cause (defined as the disease or injury that initiated the series of events leading directly to death, or the circumstances of the accident or violence that produced the fatal injury^8^) and up to 15 contributory causes are recorded on the death certificate. Additional information in the database include: unique NHS identifier, age, sex, registered date of death, residential postcode and place of death category. For each death, potential years of life lost (YLL) were calculated using 2019 single year sex-specific life tables for England & Wales.^9 10^ Those aged over 100 were assumed to have the life expectancy of centenarians of the same sex.

Using residential postcodes, the database was linked to Lower Super Output Areas (LSOAs), which are small geographical areas containing a median of around 1,500 residents. Additional information was linked through LSOAs, including region, population sizes by age and sex strata, and area-level socioeconomic deprivation information. Deprivation was measured using the Indices of Multiple Deprivation (IMD), a composite score across seven domains: income, employment, health, education & skills, housing, crime and environment. Deprivation rankings and scores often remain relatively constant over time,^11^ and so we used the latest available IMDs reported as scores (for 2019 in England and 2014 in Wales). Population estimates at the LSOA level were available from the Office for National Statistics (ONS) up to 2019,^12^ and were extrapolated to 2020 using simple linear regression (year modelled as continuous predictor). Regional information pertained to the 10 former Strategic Health Authorities in England & Wales, which were responsible for management of performance, enacting directives and implementation of health policy locally as required by the Department of Health & Social Care.

Deaths were organised in two main categories: direct or indirect. Direct deaths were those where the underlying cause of death was attributed to COVID-19 (ICD-10 code U071 virus identified, or U072 virus not identified), plus respiratory deaths (J00–J99). This aggregation was used because coding of COVID-19 varied over the study’s observation period, with an unknown number of COVID-19 deaths being attributed to other respiratory diseases, especially in the early stages of the pandemic and in care homes.^13^ For comparison, we also present and model non-COVID-19 respiratory deaths separately. We examined three subcategories of deaths indirectly related to COVID-19: cardiovascular and diabetes (ICD-10 codes: I00–I99 and E10-E14, except for I426 alcoholic cardiomyopathy), since these are closely related disorders; cancer (ICD-10 codes: C00–D48); and other indirect deaths (including drug-related and alcohol-specific deaths, suicides, accidents and all other causes). More details on the selected groups have been provided elsewhere.^2^

### Analyses

Data were imported, cleaned and formatted as a time series in Stata v16. Total years of life lost for each category were aggregated, initially without stratification and then across the following strata: by sex, by region and by deprivation quintile. Sex-specific age-standardised mortality rates (ASMRs) per 100,000 population (and 95% confidence intervals) were computed using the World Health Organization (WHO) World Standard Population to allow for global comparisons,^14^ and are presented graphically across the examined strata. Aggregated datasets were extracted from the secure environment and analysed locally.

Time series of aggregate YLL for each examined death category were analysed in similar models. To model YLL, data from week 9 in 2020 (22-28 February), two weeks before the first officially confirmed COVID-19 related deaths (7-13 March), were set to ‘missing’. A linear regression model with Newey-West standard errors was used on the aggregated England & Wales data, while linear regression models with Driscoll-Kray standard errors were used for all the panel time series (with both error structures allowing for time autocorrelation, with the latter also allowing correlation between panels).^15^ We used the built-in *newey* and the user-written *xtscc* time-series analysis commands in Stata,^16^ with the maximum lag order being set to 52 weeks (a year). As well as fitting time as a continuous covariate, the models included week number (1-52) as a categorical covariate, to account for potential seasonality and to obtain more accurate predictions for the investigated time-period. Each of the panel time series models included a categorical term of the respective panel (i.e. sex, region, or deprivation quintile). For weeks 11 (7 to 13 March) to 52 (19 to 25 Dec 2020), we used the *margins* post-estimation command to obtain the linear prediction from each model (and its standard error, computed using the delta-method), which provides an estimate of the weekly expected YLL. This estimate was then subtracted from the respective weekly observed YLL to give an estimate of weekly excess YLL. To better quantify the YLL respective to the size of the population, we also computed YLL per 100,000 population, and repeated all analyses. Additional figures by each categorisation of interest are provided in the supplementary file. We also provide the sex-specific age-standardised mortality ratios (ASMRs) in data files, to enable international comparisons. Our previously reported analyses on excess all-cause and direct deaths were updated for this study period, allowing us to quantify excess mortality across deprivation quintiles and regions, by age group.^17^ This allowed us to quantify the years of life lost per excess death, and report excess number of deaths across age-groups. These interactions were not analysed in other death categories due to low cell counts.

## Results

### All-cause

We estimated 763,550 (95% CI: 696,826 to 830,273) excess YLL during the study period across England & Wales, equivalent to 15% (95% CI: 14 to 16) of all-cause YLL observed during the equivalent time period in 2019. Excess YLL were higher for males (461,919; 95% CI: 426,408 to 497,430) compared to females (301,631; 95% CI: 267,557 to 335,706) and were highest in London (126,761; 95% CI: 121,370 to 132,152) and the North West (121,017; 95% CI: 110,647 to 131,387), and lowest in the South Central (39,953; 95% CI: 33,469 to 46,436) and the South West of England (35,032; 95% CI: 27,142 to 42,923). Excess YLL were lowest in the least deprived quintile (118,457; 95% CI: 106,554 to 130,360) and highest in the most deprived quintile (220,798; 95% CI: 204,705 to 236,891). This inequality is also captured in the sex-specific ASMRs, across both sexes (Figure 1). Excess all-cause deaths across age-groups, by deprivation quintile and region are presented in Appendix Table 5, with a larger ratio of excess deaths in the most deprived over the least deprived areas in younger age groups (45-64: 3150/1050; 65-74: 3317/1722; 75-84: 5916/4279; 85+: 5771/6094). The North West and London had the highest number of excess deaths in people aged 74 or below.

**Figure 1:**
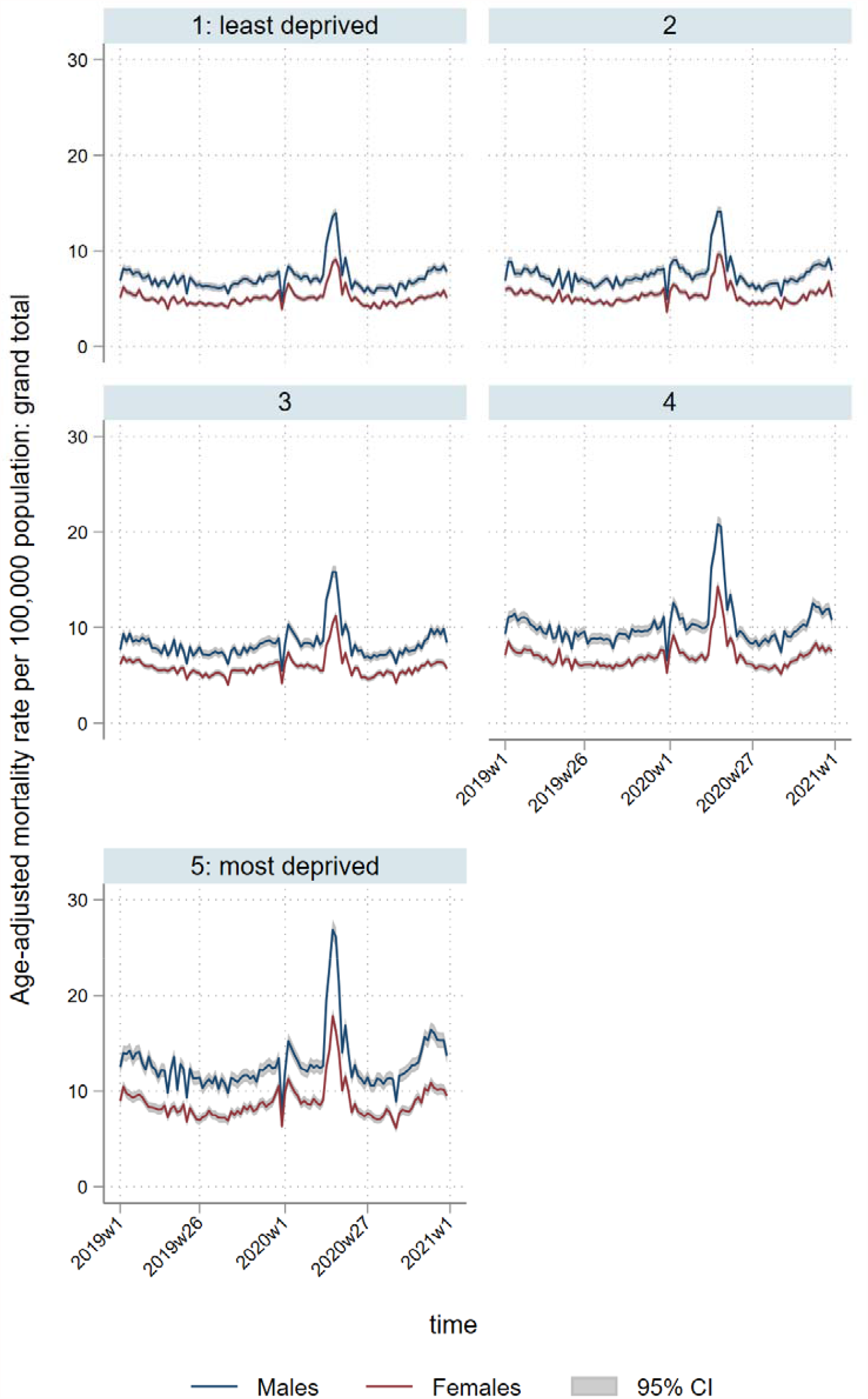
Temporal patterns in age-standardised mortality rates per 100,000 population for all deaths, by sex and deprivation quintile

There were 1,125 (95% CI: 997 to 1,252) excess YLL per 100,000 nationally, ranging from 490 (95% CI: 319 to 661) per 100,000 in the South West to 1,550 (95% CI: 1,384 to 1,716) per 100,000 in the North West. Excess YLL per 100,000 population were unequally distributed across deprivation quintiles, with similar rates in quintiles 1 (916; 95% CI: 820 to 1,012) to 3 (977; 95% CI: 869 to 1,085), but sharply increasing for quintiles 4 (1,218; 95% CI: 1,089 to 1,346) and 5 (1,645; 95% CI: 1,472 to 1,819). Patterns were broadly similar to those for deaths caused directly by COVID-19 or other respiratory causes, which comprised the majority of excess deaths over the study period (Figure 2). Excess all-cause deaths across age-groups per 100,000 population, by deprivation quintile and region are presented in Appendix Table 6, with a similar pattern of higher excess deaths for younger people in more deprived areas, but less pronounced due to the known socioeconomic gap between the young and the old.^18^ Assuming estimated excess all-cause YLL were only distributed amongst estimated excess all-cause deaths, an average of 9.1 years per death (95% CI: 8.2 to 10.0) were lost in the least deprived quintile, compared to 10.8 (95% CI: 10.0 to 11.6) in the most deprived. The North West and London continued to have the highest rate of excess deaths, closely followed by West Midlands.

**Figure 2:**
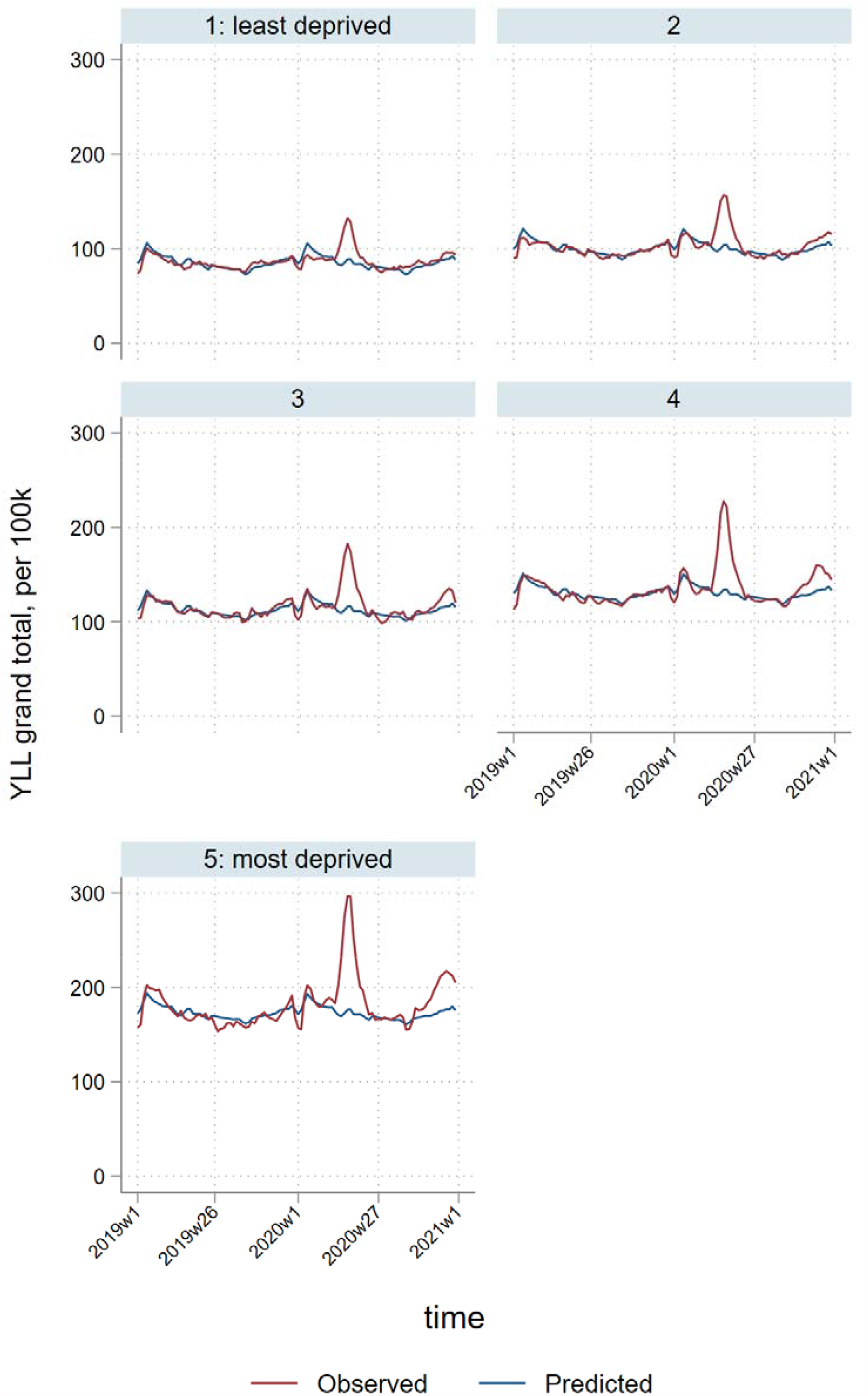
Temporal patterns in years of life lost per 100,000 population due to all-cause deaths, by deprivation quintile^*^ ^*^3-week smoothed

### Respiratory disease

Between 7^th^ March 2020 and 25^th^ December 2020 there were an estimated 646,518 (95% CI: 632,925 to 660,111) excess YLL in England & Wales attributed to COVID-19 or another underlying respiratory cause (direct, Table 1), representing an 123% increase (95% CI: 120% to 126%) compared to the equivalent time period in 2019. Of these, 387,406 (95% CI: 380,649 to 394,163) were in males and 259,112 (95% CI: 251,991 to 266,233) in females. There was a steep deprivation gradient, ranging from 92,782 (95% CI: 90,595 to 94,968) excess YLL in the least deprived quintile to 181,298 (95% CI: 177,509 to 185,086) in the most deprived. This inequality is reflected in the sex-specific ASMRs, with rates for males in the most deprived areas approximately double those of females in the most deprived areas or of males in the least deprived areas (Figure 3). Excess respiratory deaths across age-groups, by deprivation quintile and region are presented in Appendix Table 5, with a larger ratio of excess deaths in the most deprived over the least deprived areas in younger age groups (45-64: 2008/681; 65-74: 2774/1377; 75-84: 5069/3655; 85+: 5302/5636). The North West and London had the highest numbers of excess respiratory deaths for people aged 74 or below.

**Table 1:**
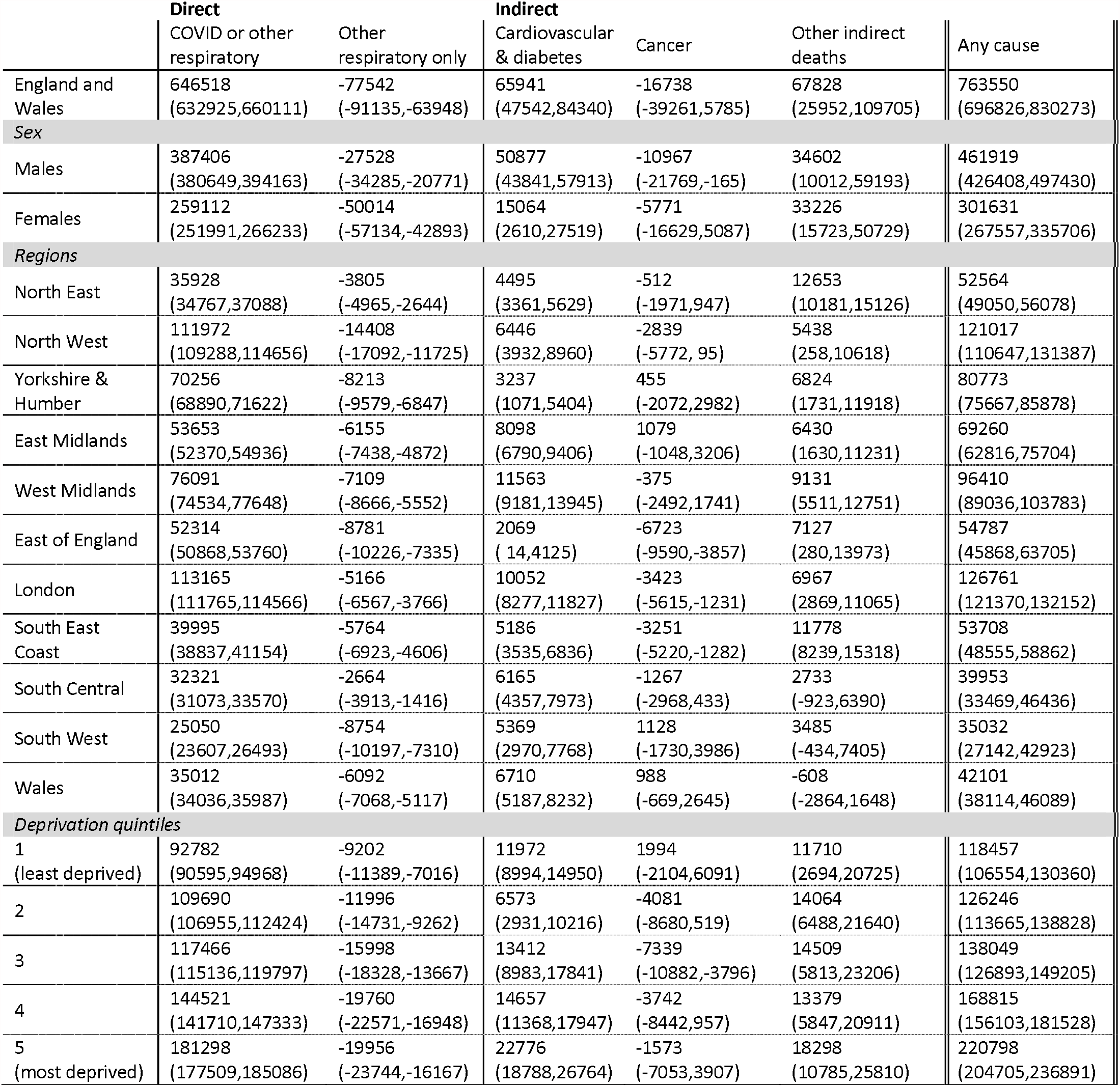
Estimated excess years of life lost, of direct, indirect and total excess deaths, weeks 11 to 52 (7 Mar 2020 to 25 Dec 2020)

**Figure 3:**
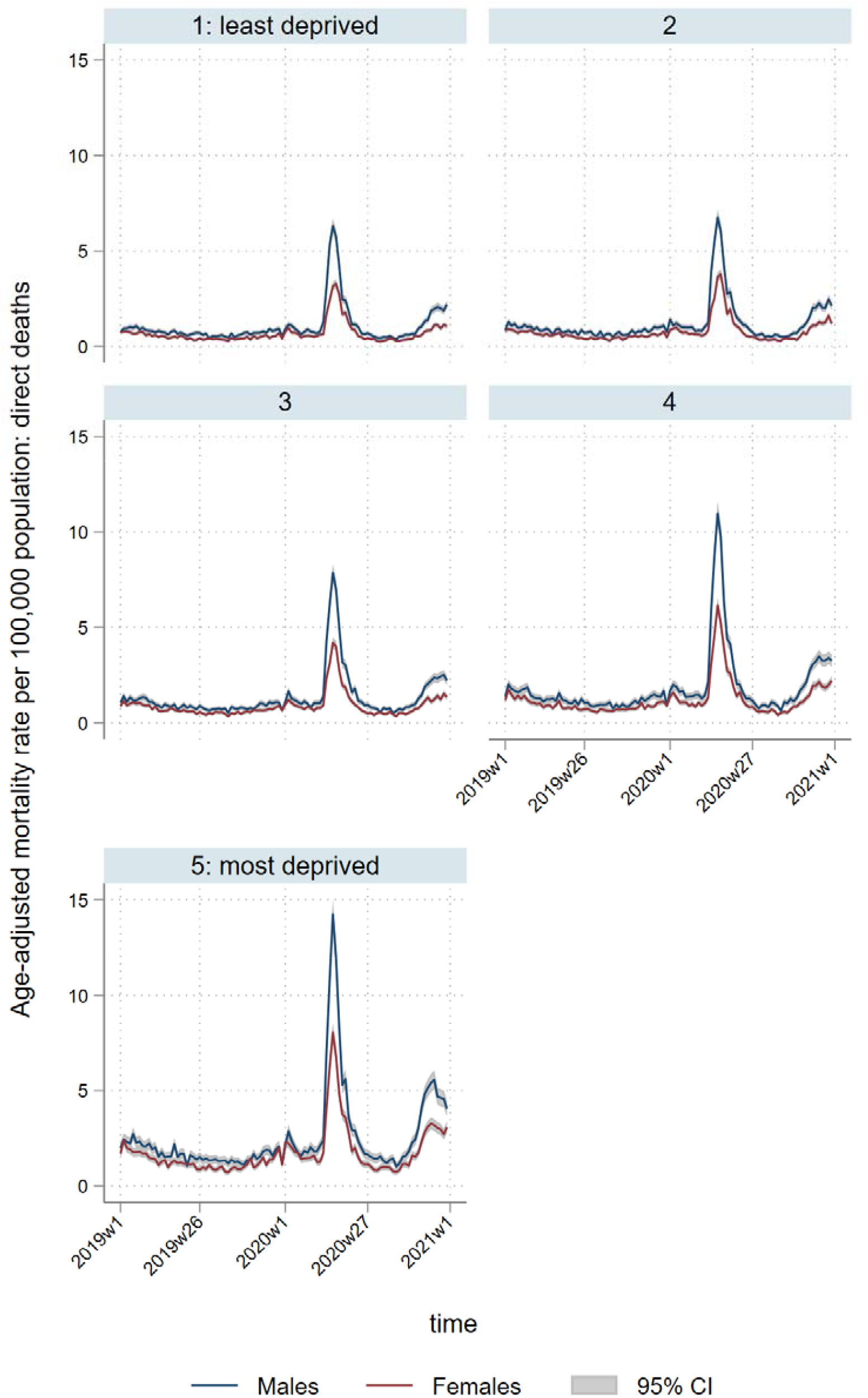
Temporal patterns in age-standardised mortality rates per 100,000 population for direct deaths, by sex and deprivation quintile

The overall excess YLL for England & Wales directly attributed to COVID-19 infection or respiratory causes was 1,066 (95% CI: 1,041 to 1,091) per 100,000 population (Table 2), but there was substantial regional variation: rates ranged from 438 (95% CI: 408 to 468) per 100,000 in the South West of England to 1,529 (95% CI: 1,494 to 1,564) per 100,000 in the North West. For males, excess YLL per 100,000 population were 1293 (95% CI: 1,269 to 1,317) for males and, for females, 844 (95% CI: 817 to 870). In terms of area-level deprivation, rates ranged from 789 (95% CI: 770 to 809) per 100,000 for the least deprived quintile to 1,493 (95% CI: 1,459 to 1,527) per 100,000 for the most deprived, reflecting higher spikes in more deprived regions and localities during the first and second phases of the pandemic (April-July and November-December 2020; Figures 3 & 4). In addition to having higher baseline YLL, areas in the most deprived quintiles experienced the highest proportional increases in YLL during the pandemic; there was an 18% increase in the most deprived quintile compared to 15% in the least deprived (Appendix Table 4). Per 100,000 population, we estimated more excess respiratory deaths for younger people in more deprived areas, but the differences were less pronounced compared to the population-unadjusted analyses, due to the larger older population in the less deprived areas (Appendix Table 6).^18^ Assuming estimated excess respiratory YLL were only distributed amongst estimated excess respiratory deaths, an average of 8.9 years per death (95% CI: 8.7 to 9.1) were lost in the least deprived quintile, compared to 11.2 (95% CI: 11.0 to 11.5) in the most deprived. London had the highest rate of excess deaths in people aged 74 or below, followed by the North West (Appendix Table 6).

**Table 2:** Estimated excess years of life lost, of direct, indirect and total excess deaths per 100,000 population, weeks 11 to 52 (7 Mar 2020 to 25 Dec 2020)

|  | Direct |  | Indirect |  |  |  |
| --- | --- | --- | --- | --- | --- | --- |
|  | COVID or other respiratory | Other respiratory only | Cardiovascular & diabetes | Cancer | Other indirect deaths | Any cause |
| England and Wales | 1066<br>(1041,1091) | -146<br>(-171,-120) | 75<br>(31,119) | -79<br>(-131,-27) | 63<br>(6,119) | 1125<br>(997,1252) |
| **Sex** |  |  |  |  |  |  |
| Males | 1293<br>(1269,1317) | -111<br>(-135,-87) | 128<br>(95,162) | -92<br>(-143,-41) | 58<br>(-6,123) | 1387<br>(1276,1499) |
| Females | 844<br>(817,870) | -179<br>(-206,-153) | 23<br>(-30,75) | -66<br>(-115,-18) | 66<br>(13,118) | 866<br>(724,1007) |
| **Regions** |  |  |  |  |  |  |
| North East | 1214<br>(1174,1253) | -268<br>(-307,-228) | 55<br>(21,88) | -98<br>(-159,-37) | 348<br>(264,432) | 1519<br>(1414,1624) |
| North West | 1529<br>(1494,1564) | -181<br>(-216,-146) | 71<br>(31,111) | -87<br>(-144,-29) | 37<br>(-36,109) | 1550<br>(1384,1716) |
| Yorkshire & Humber | 1265<br>(1239,1290) | -155<br>(-181,-130) | 32<br>(-17,80) | -34<br>(-92,24) | 85<br>(8,162) | 1347<br>(1264,1429) |
| East Midlands | 1074<br>(1046,1102) | -155<br>(-183,-127) | 114<br>(70,157) | -47<br>(-96,2) | 63<br>(-17,142) | 1203<br>(1089,1318) |
| West Midlands | 1272<br>(1245,1300) | -122<br>(-150,-95) | 163<br>(112,213) | -61<br>(-109,-13) | 101<br>(42,161) | 1475<br>(1324,1626) |
| East of England | 840<br>(814,865) | -135<br>(-160,-109) | 13<br>(-25,52) | -153<br>(-214,-93) | 80<br>(-15,175) | 779<br>(644,914) |
| London | 1294<br>(1266,1321) | -19<br>(-47,9) | 117<br>(83,151) | -67<br>(-110,-25) | 68<br>(25,110) | 1411<br>(1300,1521) |
| South East Coast | 808<br>(782,833) | -146<br>(-172,-121) | 64<br>(16,113) | -127<br>(-186,-68) | 187<br>(124,250) | 932<br>(802,1062) |
| South Central | 699<br>(673,726) | -90<br>(-117,-64) | 99<br>(53,144) | -74<br>(-113,-35) | 11<br>(-66,88) | 735<br>(593,877) |
| South West | 438<br>(408,468) | -160<br>(-190,-130) | 65<br>(9,122) | -31<br>(-95,33) | 18<br>(-44,80) | 490<br>(319,661) |
| Wales | 1014<br>(986,1042) | -284<br>(-311,-256) | 123<br>(64,182) | -35<br>(-83,12) | -115<br>(-178,-53) | 986<br>(871,1102) |
| **Deprivation quintiles** |  |  |  |  |  |  |
| 1<br>(least deprived) | 789<br>(770,809) | -92<br>(-111,-72) | 79<br>(47,111) | -21<br>(-59,17) | 69<br>(-2,139) | 916<br>(820,1012) |
| 2 | 913<br>(888,938) | -116<br>(-141,-91) | 23<br>(-20,65) | -85<br>(-133,-37) | 74<br>(18,130) | 925<br>(804,1045) |
| 3 | 952<br>(929,974) | -147<br>(-169,-124) | 74<br>(23,124) | -116<br>(-161,-70) | 68<br>(12,124) | 977<br>(869,1085) |
| 4 | 1163<br>(1137,1189) | -177<br>(-202,-151) | 83<br>(43,123) | -83<br>(-140,-26) | 55<br>(7,102) | 1218<br>(1089,1346) |
| 5<br>(most deprived) | 1493<br>(1459,1527) | -192<br>(-226,-158) | 144<br>(97,191) | -72<br>(-138,-5) | 80<br>(21,139) | 1645<br>(1472,1819) |

**Figure 4:**
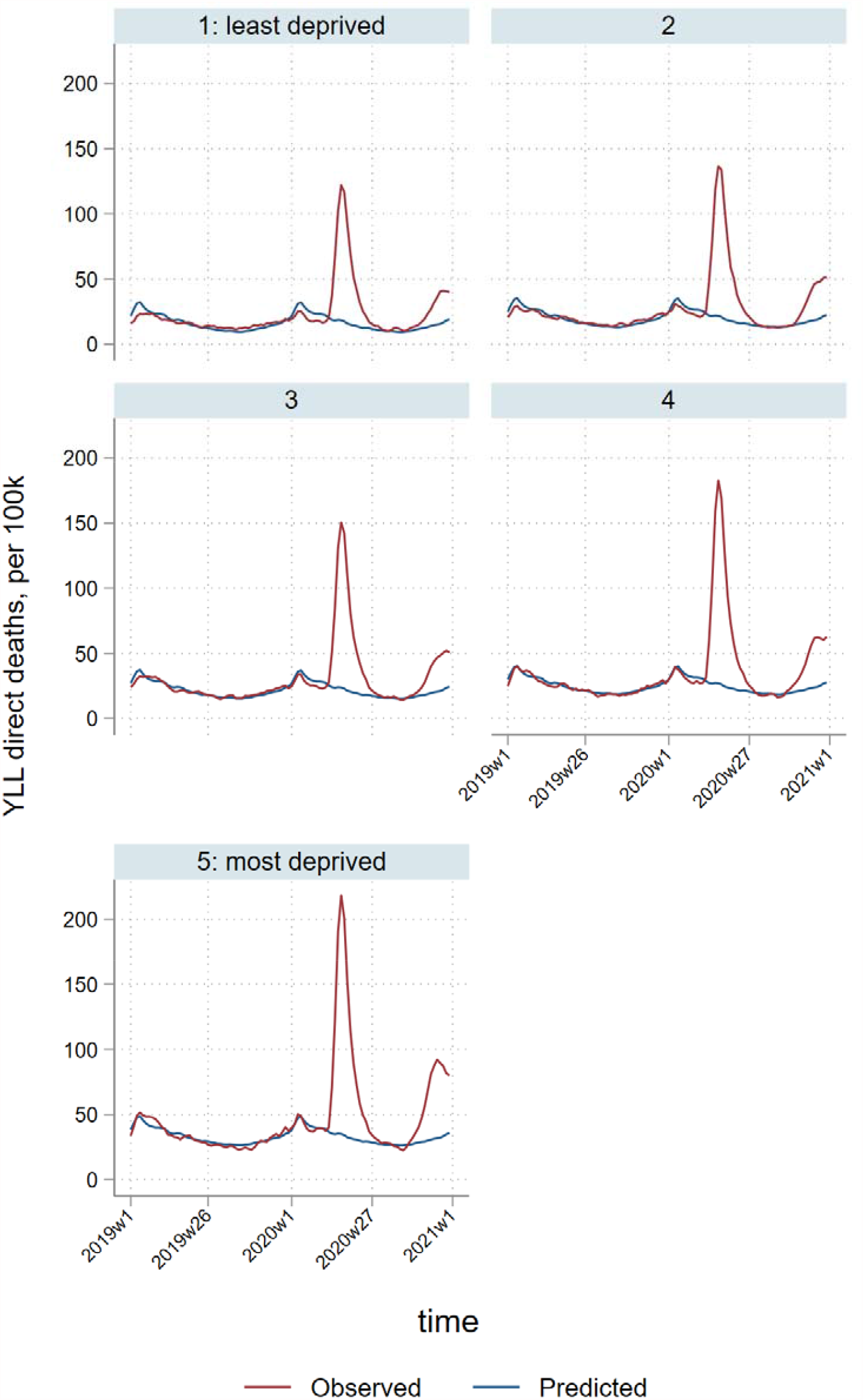
Temporal patterns in years of life lost per 100,000 population due to direct deaths, by deprivation quintile^*^ ^*^3-week smoothed

### Cardiovascular disease & diabetes

We observed an increase in YLL due to cardiovascular and diabetes deaths, with an estimated 65,941 (95% CI: 47,542 to 84,340) excess YLL nationally, reflecting a 6% (95% CI: 4% to 7%) increase compared to the equivalent time period in 2019. Furthermore, YLL attributable to cardiovascular and diabetes deaths was over three times greater for males, compared to females (50,877; 95%CI: 43,841 to 57,913 vs 15,064; 95%CI: 2,610 to 27,519). There was an overall socioeconomic gradient evident in these excess YLL; 11,927 (95% CI: 8,994 to 14,950) and 22,776 (95% CI: 18,788 to 26,764) excess YLL were estimated in the least and most deprived areas, respectively.

Nationally, most of the excess YLL due to cardiovascular disease/diabetes occurred during the first half of the first wave of the pandemic (April to mid-May), but observed YLL exceeded predictions until the end of 2020 (Figure 5). There were 75 (95% CI: 31 to 119) excess YLL per 100,000 of the population, ranging from 13 (95% CI: -25 to 55) per 100,000 in the East of England to 163 (95% CI: 112 to 213) per 100,000 in the West Midlands. Rates ranged from 23 (95% CI: -20 to 65) per 100,000 in the second least deprived to 144 (95% CI: 97 to 191) per 100,000 in the most deprived quintile.

**Figure 5:**
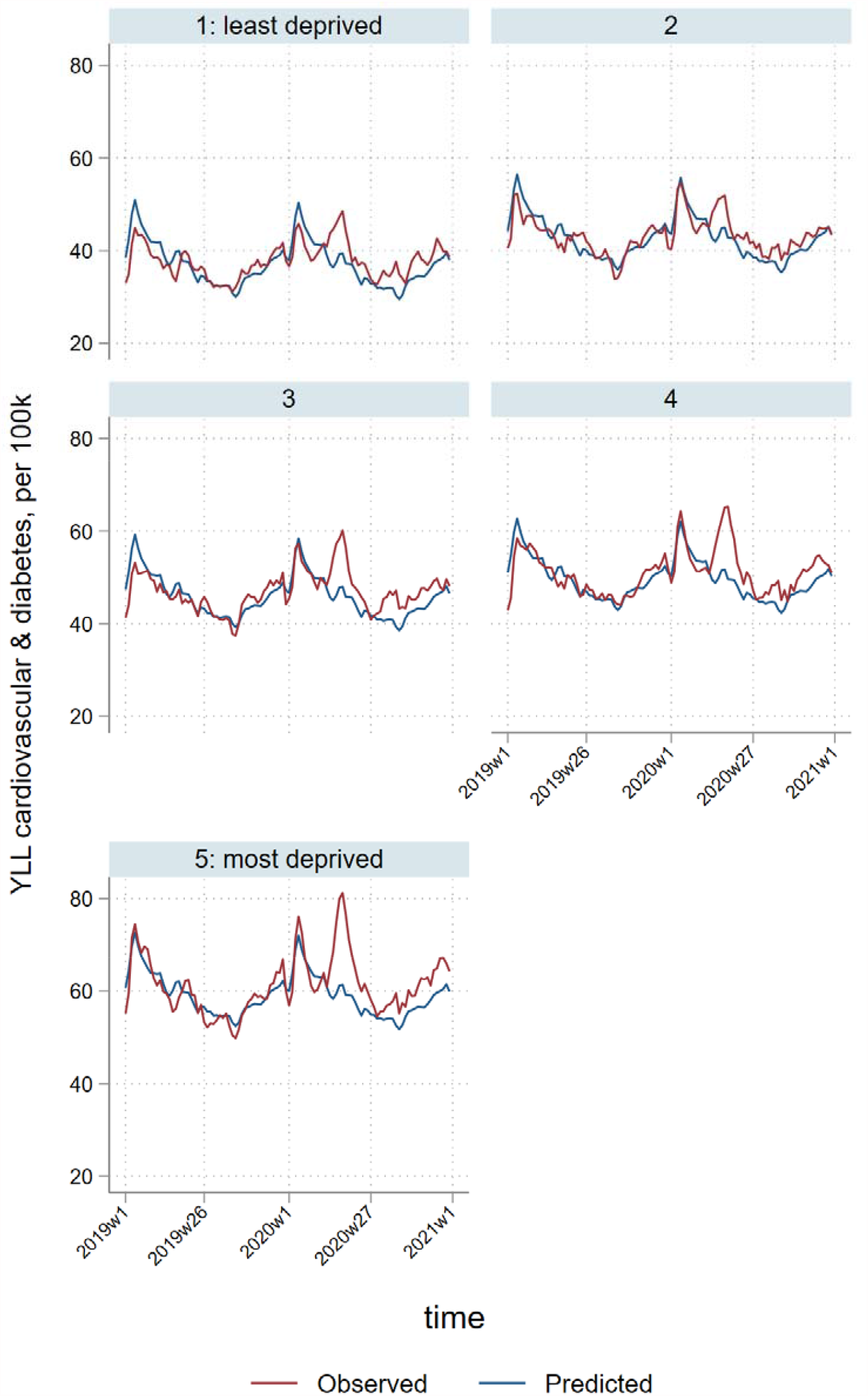
Temporal patterns in years of life lost per 100,000 population due to cardiovascular & diabetes deaths, by deprivation quintile^*^ ^*^3-week smoothed

### Cancer and other indirect deaths (including drug-related, alcohol-specific, suicides, fatal accidents, and all other causes)

We observed no significant pandemic-related changes in YLL due to cancer. Nationally, there were -16,738 (95% CI: -39,261 to 5,785) excess YLL due to cancer deaths over the study period. Regional and deprivation quintile estimates were small, and in some cases not statistically significant. The highest rates per 100,000 population were in the least deprived quintile and in the South West region, but even here estimates were negative, albeit non-significant.

Nationally, there were 67,828 (95% CI: 25,952 to 109,705) excess YLL attributable to other indirect deaths, with wide regional variability, from 12,653 (95% CI: 10,181 to 15,126) in the North East to -608 (95% CI: -2,864 to 1,648) in Wales. Excess YLL ranged from 11,710 (95% CI: 2,694 to 20,725) in the least deprived quintile to 18,298 (95% CI: 10,754 to 25,810) in the most deprived. Estimates of rates of excess YLL were non-significant for some regions, with substantially higher rates in the North East of England (348 per 100,000). There was no clear socioeconomic pattern, although the highest YLL excess rates were observed in the most deprived quintile (80 per 100,000).

Supplementary file Table 1 presents total observed YLL across each category, and Table 2 presents and percentages of excess YLL in relation to these totals. In the supplementary file we also provide YLL denominators for the equivalent weeks in 2019 (Table 3), and the percentages of excess YLL in 2020, in relation to the 2019 baselines (Table 4). Persistent socioeconomic and regional health inequalities exist in England & Wales, as indicated by YLL. That is, in the last 42 weeks of 2019 (the equivalent time-period we investigated in 2020 to quantify the impact of the COVID-19 pandemic), there was a steep socioeconomic gradient in YLL, with 804,312 in the least deprived quintile, increased to 1,255,588 in the most deprived. The ratio of YLL in the most deprived over the least deprived quintile was 1.56. For the equivalent time period in 2020, this ratio increased to 1.64 (1,501,323/913,418).

## Discussion

Before the COVID-19 pandemic, there were clear and long-standing socioeconomic and geographic patterns in years of life lost (YLL) in England & Wales, such that years lost increased with material deprivation and were persistently higher in northern English regions and in Wales, especially for males.^19 20^ This is reflected in pre-pandemic differences in the ASMRs across deprivation quintiles (Figure 1 and supplementary file). In the first 42 weeks of the pandemic, these underlying inequalities widened as the most deprived areas experienced the greatest losses, with total excess YLL nearly twice as high in the most deprived quintile, compared to the most affluent. Similarly, the most deprived areas experienced the greatest proportional increase in excess YLL, compared with the least deprived. Higher rates of YLL occurred in northern regions, although the West Midlands and London were also badly affected, most likely reflecting poverty in the major conurbations. Patterns for indirect deaths, i.e. those not directly attributed to COVID-19, varied by cause. Excess YLL from deaths attributable to cardiovascular disease and diabetes followed a similar socioeconomic pattern, and no obvious pattern was apparent for other deaths.

### Strengths and Limitations

Our analysis provides a comprehensive picture of excess years of life lost during the first 42 weeks of pandemic, including major causes both related and unrelated to COVID-19 infection. This enabled us to estimate the total impact of the pandemic on years of life lost, including regional and area-level socioeconomic patterns. There are some limitations to our study. For deaths not referred for investigation and adjudication by a coroner, data on cause of death rely on accurate diagnosis and recording by clinicians. During a pandemic, assessing the contribution of COVID-19 to some deaths can be challenging.^21^ We defined COVID-19 deaths as those for which the underlying cause was attributed to the virus on the death certificate. Inevitably, there will have been some misclassification, particularly in the early part of the pandemic when COVID-19 testing was not widespread and doctors’ awareness may have been more limited.^22 23^ This is also problematic for estimating years of life lost by deaths caused indirectly by the pandemic, so in addition to excluding COVID-19 deaths from the total excess deaths, we also excluded other respiratory causes of death (the most likely source of ‘missed’ COVID-19 diagnoses). Furthermore, this approach enabled us to apply data from earlier years to create a reference ‘baseline’, in the absence of COVID-19 deaths occurring before 2020. Consequently, our estimates of excess years of life lost that occurred as an indirect effect of the pandemic do not include respiratory deaths not attributable to COVID-19, and are unlikely to include substantial numbers caused directly by COVID-19 infection. The aggregates of weekly estimates that we have reported included negative estimates (i.e. they were not set to zero), to allow for more conservative estimates. We did not control for population trends, assuming small increases over time, that might have a small impact on our estimates. There was also relatively high variability in the weekly time-trends of some subgroups, due to relatively small numbers, but that uncertainty is reflected in the estimated confidence intervals. The survival tables we used were not region specific, to highlight baseline differences across regions, and the choice of survival tables, national or regional, would not have greatly affected our estimates of excess YLL since they are based on historical trends. Finally, time-lags for coroners’ verdicts will not have been sufficiently long for many cases, particularly for external causes of death such as accidents and suicides, and we are therefore unable to determine excess years of life lost specific to these categories. We categorised groups to minimise the impact of these time-lags, with the most affected causes contributing to a single group along with those whose causes remained undetermined (‘other indirect deaths’).

### Clinical interpretation and implications of findings

We estimate that there were 763,550 excess years of life lost during 2020 in England & Wales since the start of the pandemic, of which 646,518 (84.7%) were directly attributed to COVID-19 or other respiratory diseases, and 117,032 (15.3%) were attributed to other causes. The greatest contributors to indirect deaths were cardiovascular disease and diabetes (65,941 YLL), with three quarters of these lost years occurring in males. This may partially reflect undiagnosed COVID-19 infection, but it is also consistent with reduced capacity for investigation and elective treatment of cardiovascular disease and hospital avoidance for acute cardiovascular events.^3^ Our observations around the relatively small negative estimates for YLL as a result of cancer may relate to decreases in the diagnosis of cancer during the pandemic, where patients were reluctant to seek medical advice for cancer symptoms and elective investigations (particularly cancer imaging) were delayed to protect patients and staff from exposure to COVID-19, and therefore undiagnosed cancer deaths were assigned to other causes.^24^ In addition, the impact of the pandemic on cancer mortality is likely to be longer term. We have previously estimated that 26% of excess deaths during the pandemic were attributable to non-respiratory causes,^2^ which suggests that these indirect deaths had an older age profile than deaths directly attributed to COVID-19, and therefore comprised a lower proportion of the total years of life lost.

The socioeconomic gradient in total years of life lost was steeper than that previously found for excess deaths.^2^ Over 50% of excess YLL were in the two most deprived quintiles, indicating that a disproportionate number of deaths in these areas, of which there were more than in more affluent areas, were in younger age groups. Given that the most deprived parts of the country had a far higher pre-pandemic level of YLL, this means that the already wide health inequalities in England & Wales were magnified during the 2020 phase of the COVID-19 pandemic, in both absolute and relative terms. In our previous work we observed that in the first 30 weeks of the pandemic mortality rates in the most deprived quintile were 1.13 (72,595/64,109) times that in the least.^2^ However, for the first 42 weeks of the pandemic, we found that YLL were 1.64 higher (1,501,323/913,418; supplementary file Table 1), indicating that a higher proportion of total deaths were in younger age groups in more deprived areas. Similarly, regions of England & Wales with persistently high YLL such as the North East and North West of England had the greatest increases in YLL. For the North East, rates of excess direct YLL were substantially lower than for the North West, and rates of ‘other’ YLL were far higher, suggesting that there may have been lower rates of recording of COVID-19 on the death certificates of younger people in the North East. However, factors other than deprivation will also have contributed, with relatively high rates of YLL in London and the West Midlands and relatively low rates in the South West and Wales, given these regions’ health, ethnicity and deprivation profiles.

## Conclusion

The COVID-19 pandemic has widened pre-existing health inequalities across England & Wales: regions and social groups with the highest baseline mortality rates experienced the greatest excesses in years of life lost. Our findings support the notion that YLL can be more informative for determining unmet needs and are in line with several of the recommendations made by the British Medical Association for mitigating the effects of COVID-19. These include prioritising vaccine delivery and providing targeted financial and social support for groups severely impacted by both the pandemic disease and the public health measures implemented to reduce transmission,^25^ as well as increased support for primary care services in the areas most affected. Many of these groups were already compromised in financial and health terms by the 2007-2009 Great Recession and subsequent years of austerity, and face further economic uncertainty following the pandemic.^26^ Successive economic shocks place constraints on policymakers, but if health inequalities are to be attenuated, and ideally significantly reduced, a robust policy response is required, including substantial investment in future pandemic preparedness and to support chronically stretched health services.^27^ Finally, more effective public health actions must continue at the national level to address the full range of socioeconomic determinants of ill health.

## Data Availability

Data are freely and publicly available from the Office of National Statistics website

https://www.ons.gov.uk/peoplepopulationandcommunity/birthsdeathsandmarriages/deaths/datasets/weeklyprovisionalfiguresondeathsregisteredinenglandandwales

## Contributors

EK, TD, MM and RW designed the study. EK extracted the data from the secure environment. EK and GM conducted the statistical analyses. EK and TD drafted the paper and all co-authors critically revised it.

## Declaration of interests

CG declares receiving support from Abbot and BMS towards this work, consulting fees from Amgen and AstraZeneca, honoraria from AstraZeneca, participation in Data Safety Monitoring Boards for several trials (TACTIC, DUAL-ACS, DANBLOCK, PROFID, RAPID NSTEMI, STEEER-AF), stock options with the European Heart Journal Quality of Care and Clinical Outcomes as Deputy Editor, and other financial interests with Wondr Medical. None of the other authors have anything relevant to declare.

## Acknowledgements

This work would have not been possible without the support of NHS Digital, in making all mortality data in England and Wales available to us, through a secure data environment.

## Data availability statement

Data are freely and publicly available from the Office of National Statistics website: https://www.ons.gov.uk/peoplepopulationandcommunity/birthsdeathsandmarriages/deaths/datasets/weeklyprovisionalfiguresondeathsregisteredinenglandandwales

## Notes

### Funding Statement

No external funding was received

