## Appendix for "Excess years of life lost to COVID-19 and other causes of death by sex, neighbourhood deprivation and region in England & Wales during 2020"

June 29, 2021

#### Contents

**1 Notes** **3**

**2 Tables** **4**

**3 Tables** **8**

**4 Tables** **9**

**5 Total** **10**
5.1 AASMRs . . . . . . . . . . 10
5.2 YLLs . . . . . . . . . . 15
5.2.1 England-Wales aggregate . . . . . . . . . . 15
5.2.2 By sex . . . . . . . . . . 18
5.2.3 By deprivation quintile . . . . . . . . . . 23
5.2.4 By Strategic Health Authority . . . . . . . . . . 28
5.3 YLLs per $100,000$ population . . . . . . . . . . 33
5.3.1 England-Wales aggregate . . . . . . . . . . 33
5.3.2 By sex . . . . . . . . . . 36
5.3.3 By deprivation quintile . . . . . . . . . . 41
5.3.4 By Strategic Health Authority . . . . . . . . . . 46

**6 Direct** **51**
6.1 AASMRs . . . . . . . . . . 51
6.2 YLLs . . . . . . . . . . 56
6.2.1 England-Wales aggregate . . . . . . . . . . 56
6.2.2 By sex . . . . . . . . . . 59
6.2.3 By deprivation quintile . . . . . . . . . . 64
6.2.4 By Strategic Health Authority . . . . . . . . . . 69

6.3 YLLs per $100,000$ population . . . 74
6.3.1 England-Wales aggregate . . . 74
6.3.2 By sex . . . 77
6.3.3 By deprivation quintile . . . 82
6.3.4 By Strategic Health Authority . . . 87

**7 Cardiovascular & diabetes** **92**
7.1 AASMRs . . . 92
7.2 YLLs . . . 97
7.2.1 England-Wales aggregate . . . 97
7.2.2 By sex . . . 100
7.2.3 By deprivation quintile . . . 105
7.2.4 By Strategic Health Authority . . . 110
7.3 YLLs per $100,000$ population . . . 115
7.3.1 England-Wales aggregate . . . 115
7.3.2 By sex . . . 118
7.3.3 By deprivation quintile . . . 123
7.3.4 By Strategic Health Authority . . . 128

**8 Cancer** **133**
8.1 AASMRs . . . 133
8.2 YLLs . . . 138
8.2.1 England-Wales aggregate . . . 138
8.2.2 By sex . . . 141
8.2.3 By deprivation quintile . . . 146
8.2.4 By Strategic Health Authority . . . 151
8.3 YLLs per $100,000$ population . . . 156
8.3.1 England-Wales aggregate . . . 156
8.3.2 By sex . . . 159
8.3.3 By deprivation quintile . . . 164
8.3.4 By Strategic Health Authority . . . 169

**9 All other indirect** **174**
9.1 AASMRs . . . 174
9.2 YLLs . . . 179
9.2.1 England-Wales aggregate . . . 179
9.2.2 By sex . . . 182
9.2.3 By deprivation quintile . . . 187
9.2.4 By Strategic Health Authority . . . 192
9.3 YLLs per $100,000$ population . . . 197
9.3.1 England-Wales aggregate . . . 197
9.3.2 By sex . . . 200
9.3.3 By deprivation quintile . . . 205
9.3.4 By Strategic Health Authority . . . 210

### 1 Notes

On years of life lost (YLLs) and age-standardised mortality rates (ASMRs):

1. YLLs calculated against 2019 life expectancy, which was 79.4 for males and 83.1 for females

2. Any death over those age thresholds, for males and females respectively does not contribute to the YLL total for the particular stratum (i.e. England, deprivation quintile or region)

3. Linear regression models using data from Jan 2015 are used to estimate excess YLLs following the start of the pandemic, in a monthly time series

4. ASMRs are age-standardised using 5-year band (plus 90 or over) extrapolated estimates from the 2011 census (available from the ONS), and the 2020-2025 WHO reference population (see this WHO link)

5. ASMRs are mortality rates per $100,000$ population

6. ASMRs are reported separately for males and females (And hence no ASMRs figures are reported under 'by sex' subsections)

7. YLLs and ASMRs are available for these groups of deaths:

   - Direct (COVID-19 identified or suspected, or respiratory)
   - Respiratory only (not reported in this file, to save some space)
   - Cardiovascular & diabetes
   - Cancer
   - All other indirect (including a few cases where no underlying cause of death, or preliminary)
   - Total (i.e. direct, cardiovascular & diabetes, cancer, and all other indirect)

8. Sums of subgroups not necessarily adding up to England-Wales aggregate (especially for deprivation when residence postcode is not available or missing)

9. Cardiovascular excludes alcoholic cardiomyopathy, which is included in all other indirect

10. Other groupings are available but not reported here, for example deprivation by region

#### 2 Tables

Table 1: YLL denominators by group for weeks 11-52 (week ending 13 Mar to week ending 25 Dec 2020)

| | **Direct** | | **Indirect** | | | |
|---|---|---|---|---|---|---|
| | COVID or respiratory | Respiratory only | Cardio-vascular & diabetes | Neoplasm | Other indirect deaths | Any cause |
| England | 1173914 | 449854 | 1231248 | 1674052 | 1781211 | 5860425 |
| *Sex* | | | | | | |
| male | 648478 | 233544 | 723654 | 836099 | 946400 | 3154631 |
| female | 525435 | 216310 | 507594 | 837953 | 834811 | 2705794 |
| *Regions* | | | | | | |
| North East | 64755 | 25023 | 63946 | 90602 | 104999 | 324303 |
| North West | 199250 | 72870 | 171953 | 226771 | 253822 | 851796 |
| Yorkshire & Humber | 125860 | 47392 | 123552 | 166875 | 178431 | 594719 |
| East Midlands | 97674 | 37866 | 106935 | 145066 | 148402 | 498077 |
| West Midlands | 131248 | 48048 | 134615 | 172769 | 197101 | 635733 |
| East of England | 105300 | 44205 | 119013 | 171563 | 178251 | 574128 |
| London | 161599 | 43268 | 134467 | 169811 | 191390 | 657267 |
| South East Coast | 80225 | 34465 | 92515 | 135823 | 141547 | 450110 |
| South Central | 61527 | 26541 | 78500 | 114260 | 112393 | 366680 |
| South West | 73469 | 39666 | 120054 | 172764 | 168367 | 534655 |
| Wales | 71080 | 29976 | 82150 | 104389 | 101881 | 359500 |
| *Deprivation quintiles* | | | | | | |
| 1 (least deprived) | 160413 | 58428 | 187051 | 307000 | 258954 | 913418 |
| 2 | 195872 | 74186 | 214563 | 327606 | 304527 | 1042569 |
| 3 | 217610 | 84146 | 243931 | 334083 | 342801 | 1138425 |
| 4 | 262227 | 97946 | 266398 | 337917 | 384693 | 1251235 |
| 5 (most deprived) | 335866 | 134613 | 315758 | 364088 | 485610 | 1501323 |

[a] Using single ICD-10 code reported in underlying cause field (mutually exclusive)
[b] Sums of subgroups not necessarily adding up to England-Wales aggregate
[c] Direct deaths: where underlying cause was COVID (confirmed or suspected) or respiratory

Table 2: Percentage of estimated excess YLL over the total of YLLs (and 95% CI) for weeks 11-52 (week ending 13 Mar to week ending 25 Dec 2020)

| | **Direct** | | **Indirect** | | | |
|---|---|---|---|---|---|---|
| | COVID or respiratory | Respiratory only | Cardio-vascular & diabetes | Neoplasm | Other indirect deaths | Any cause |
| England | 55(54,56) | -17(-20,-14) | 5(4,7) | -1(-2,0) | 4(1,6) | 13(12,14) |
| *Sex* | | | | | | |
| male | 60(59,61) | -12(-15,-9) | 7(6,8) | -1(-3,-0) | 4(1,6) | 15(14,16) |
| female | 49(48,51) | -23(-26,-20) | 3(1,5) | -1(-2,1) | 4(2,6) | 11(10,12) |
| *Regions* | | | | | | |
| North East | 55(54,57) | -15(-20,-11) | 7(5,9) | -1(-2,1) | 12(10,14) | 16(15,17) |
| North West | 56(55,58) | -20(-23,-16) | 4(2,5) | -1(-3,0) | 2(0,4) | 14(13,15) |
| Yorkshire & Humber | 56(55,57) | -17(-20,-14) | 3(1,4) | 0(-1,2) | 4(1,7) | 14(13,14) |
| East Midlands | 55(54,56) | -16(-20,-13) | 8(6,9) | 1(-1,2) | 4(1,8) | 14(13,15) |
| West Midlands | 58(57,59) | -15(-18,-12) | 9(7,10) | -0(-1,1) | 5(3,6) | 15(14,16) |
| East of England | 50(48,51) | -20(-23,-17) | 2(0,3) | -4(-6,-2) | 4(0,8) | 10(8,11) |
| London | 70(69,71) | -12(-15,-9) | 7(6,9) | -2(-3,-1) | 4(1,6) | 19(18,20) |
| South East Coast | 50(48,51) | -17(-20,-13) | 6(4,7) | -2(-4,-1) | 8(6,11) | 12(11,13) |
| South Central | 53(51,55) | -10(-15,-5) | 8(6,10) | -1(-3,0) | 2(-1,6) | 11(9,13) |
| South West | 34(32,36) | -22(-26,-18) | 4(2,6) | 1(-1,2) | 2(-0,4) | 7(5,8) |
| Wales | 49(48,51) | -20(-24,-17) | 8(6,10) | 1(-1,3) | -1(-3,2) | 12(11,13) |
| *Deprivation quintiles* | | | | | | |
| 1 (least deprived) | 58(56,59) | -16(-19,-12) | 6(5,8) | 1(-1,2) | 5(1,8) | 13(12,14) |
| 2 | 56(55,57) | -16(-20,-12) | 3(1,5) | -1(-3,0) | 5(2,7) | 12(11,13) |
| 3 | 54(53,55) | -19(-22,-16) | 5(4,7) | -2(-3,-1) | 4(2,7) | 12(11,13) |
| 4 | 55(54,56) | -20(-23,-17) | 6(4,7) | -1(-2,0) | 3(2,5) | 13(12,15) |
| 5 (most deprived) | 54(53,55) | -15(-18,-12) | 7(6,8) | -0(-2,1) | 4(2,5) | 15(14,16) |

[a] Using single ICD-10 code reported in underlying cause field (mutually exclusive)
[b] Sums of subgroups not necessarily adding up to England-Wales aggregate
[c] Direct deaths: where underlying cause was COVID (confirmed or suspected) or respiratory

Table 3: YLL denominators by group for weeks 11-52 in 2019

| | **Direct** | | **Indirect** | | | |
|---|---|---|---|---|---|---|
| | COVID or respiratory | Respiratory only | Cardio-vascular & diabetes | Neoplasm | Other indirect deaths | Any cause |
| England | 525747 | 525747 | 1146098 | 1669307 | 1734200 | 5075352 |
| *Sex* | | | | | | |
| male | 265988 | 265988 | 665188 | 834610 | 933391 | 2699178 |
| female | 259758 | 259758 | 480910 | 834697 | 800809 | 2376174 |
| *Regions* | | | | | | |
| North East | 31466 | 31466 | 60709 | 91497 | 94984 | 278656 |
| North West | 85058 | 85058 | 162036 | 228128 | 247841 | 723065 |
| Yorkshire & Humber | 54909 | 54909 | 115093 | 162543 | 178288 | 510833 |
| East Midlands | 44231 | 44231 | 98092 | 144400 | 146067 | 432790 |
| West Midlands | 55149 | 55149 | 118099 | 169146 | 185740 | 528133 |
| East of England | 51182 | 51182 | 115136 | 173473 | 172102 | 511893 |
| London | 46769 | 46769 | 124503 | 170577 | 186141 | 527991 |
| South East Coast | 40294 | 40294 | 84757 | 134269 | 133940 | 393260 |
| South Central | 31289 | 31289 | 72630 | 116695 | 112621 | 333235 |
| South West | 47225 | 47225 | 111190 | 167657 | 163310 | 489382 |
| Wales | 37016 | 37016 | 76188 | 104899 | 104532 | 322635 |
| *Deprivation quintiles* | | | | | | |
| 1 (least deprived) | 72092 | 72092 | 173528 | 302562 | 256129 | 804312 |
| 2 | 89389 | 89389 | 203652 | 330742 | 294091 | 917874 |
| 3 | 100110 | 100110 | 225879 | 338356 | 338398 | 1002744 |
| 4 | 114623 | 114623 | 249117 | 333735 | 373880 | 1071355 |
| 5 (most deprived) | 148373 | 148373 | 286257 | 357890 | 463068 | 1255588 |

[a] Using single ICD-10 code reported in underlying cause field (mutually exclusive)
[b] Sums of subgroups not necessarily adding up to England-Wales aggregate
[c] Direct deaths: where underlying cause was COVID (confirmed or suspected) or respiratory

Table 4: Percentage of estimated excess YLL for weeks 11-52 in 2020 over the total of YLLs for equivalent weeks in 2019 (and 95% CI)

| | **Direct** | | **Indirect** | | | |
|---|---|---|---|---|---|---|
| | COVID or respiratory | Respiratory only | Cardio-vascular & diabetes | Neoplasm | Other indirect deaths | Any cause |
| England | 123(120,126) | -15(-17,-12) | 6(4,7) | -1(-2,0) | 4(1,6) | 15(14,16) |
| *Sex* | | | | | | |
| male | 146(143,148) | -10(-13,-8) | 8(7,9) | -1(-3,-0) | 4(1,6) | 17(16,18) |
| female | 100(97,102) | -19(-22,-17) | 3(1,6) | -1(-2,1) | 4(2,6) | 13(11,14) |
| *Regions* | | | | | | |
| North East | 114(110,118) | -12(-16,-8) | 7(6,9) | -1(-2,1) | 13(11,16) | 19(18,20) |
| North West | 132(128,135) | -17(-20,-14) | 4(2,6) | -1(-3,0) | 2(0,4) | 17(15,18) |
| Yorkshire & Humber | 128(125,130) | -15(-17,-12) | 3(1,5) | 0(-1,2) | 4(1,7) | 16(15,17) |
| East Midlands | 121(118,124) | -14(-17,-11) | 8(7,10) | 1(-1,2) | 4(1,8) | 16(15,17) |
| West Midlands | 138(135,141) | -13(-16,-10) | 10(8,12) | -0(-1,1) | 5(3,7) | 18(17,20) |
| East of England | 102(99,105) | -17(-20,-14) | 2(0,4) | -4(-6,-2) | 4(0,8) | 11(9,12) |
| London | 242(239,245) | -11(-14,-8) | 8(7,9) | -2(-3,-1) | 4(2,6) | 24(23,25) |
| South East Coast | 99(96,102) | -14(-17,-11) | 6(4,8) | -2(-4,-1) | 9(6,11) | 14(12,15) |
| South Central | 103(99,107) | -9(-13,-5) | 8(6,11) | -1(-3,0) | 2(-1,6) | 12(10,14) |
| South West | 53(50,56) | -19(-22,-15) | 5(3,7) | 1(-1,2) | 2(-0,5) | 7(6,9) |
| Wales | 95(92,97) | -16(-19,-14) | 9(7,11) | 1(-1,3) | -1(-3,2) | 13(12,14) |
| *Deprivation quintiles* | | | | | | |
| 1 (least deprived) | 129(126,132) | -13(-16,-10) | 7(5,9) | 1(-1,2) | 5(1,8) | 15(13,16) |
| 2 | 123(120,126) | -13(-16,-10) | 3(1,5) | -1(-3,0) | 5(2,7) | 14(12,15) |
| 3 | 117(115,120) | -16(-18,-14) | 6(4,8) | -2(-3,-1) | 4(2,7) | 14(13,15) |
| 4 | 126(124,129) | -17(-20,-15) | 6(5,7) | -1(-3,0) | 4(2,6) | 16(15,17) |
| 5 (most deprived) | 122(120,125) | -13(-16,-11) | 8(7,9) | -0(-2,1) | 4(2,6) | 18(16,19) |

[a] Using single ICD-10 code reported in underlying cause field (mutually exclusive)
[b] Sums of subgroups not necessarily adding up to England-Wales aggregate
[c] Direct deaths: where underlying cause was COVID (confirmed or suspected) or respiratory

### 3 Tables

Table 5: Excess deaths (and 95% CI) by age-group for weeks 11-52 (week ending 13 Mar to week ending 25 Dec 2020)

COVID/repiratory

| | 15-44 | 45-64 | 65-74 | 75-84 | 85+ |
|---|---|---|---|---|---|
| IMD | | | | | |
| 1(least) | 51(46,57) | 681(658,703) | 1377(1317,1436) | 3655(3515,3795) | 5636(5389,5883) |
| 2 | 75(69,81) | 856(827,884) | 1538(1467,1609) | 3801(3646,3957) | 5376(5143,5609) |
| 3 | 87(79,95) | 1087(1051,1123) | 1694(1612,1777) | 4059(3908,4211) | 4902(4684,5120) |
| 4 | 153(142,164) | 1599(1546,1653) | 2122(2021,2224) | 4613(4444,4781) | 5015(4806,5223) |
| 5(most) | 268(250,285) | 2008(1927,2090) | 2774(2648,2900) | 5069(4896,5242) | 5302(5125,5479) |
| Regions | | | | | |
| NE | 9(5,13) | 237(220,254) | 519(487,550) | 1312(1260,1365) | 1799(1734,1864) |
| NW | 108(99,116) | 1081(1042,1120) | 1757(1685,1830) | 3879(3756,4003) | 4106(3965,4248) |
| York&H | 41(35,48) | 610(583,637) | 1050(999,1102) | 2481(2396,2566) | 3104(3004,3204) |
| EastM | 40(35,45) | 477(455,498) | 807(766,847) | 1766(1695,1836) | 2314(2225,2404) |
| WestM | 92(85,99) | 730(706,754) | 1113(1065,1160) | 2514(2427,2602) | 2922(2811,3034) |
| East | 43(37,48) | 500(479,522) | 827(784,870) | 1862(1779,1944) | 2304(2183,2425) |
| London | 205(199,211) | 1490(1465,1514) | 1615(1575,1655) | 2844(2779,2909) | 3098(3006,3189) |
| SECoast | 30(26,35) | 333(315,351) | 564(528,601) | 1353(1286,1419) | 2123(2020,2227) |
| SouthC | 25(21,29) | 278(264,292) | 410(381,438) | 1051(998,1104) | 1564(1484,1643) |
| SW | 12(8,17) | 231(210,251) | 366(326,406) | 933(856,1010) | 1563(1448,1679) |
| Wales | 30(26,34) | 283(264,302) | 519(483,555) | 1243(1181,1304) | 1408(1329,1486) |

Total

| | 15-44 | 45-64 | 65-74 | 75-84 | 85+ |
|---|---|---|---|---|---|
| IMD | | | | | |
| 1(least) | 42(-13,98) | 1050(862,1237) | 1722(1419,2024) | 4279(3669,4890) | 6094(5092,7096) |
| 2 | 104(43,165) | 1268(1060,1476) | 1795(1471,2118) | 3883(3281,4486) | 5717(4776,6659) |
| 3 | 29(-49,107) | 1710(1477,1942) | 2236(1909,2563) | 4535(3961,5109) | 5772(4905,6639) |
| 4 | 273(176,369) | 2361(2080,2643) | 2393(2037,2748) | 5435(4869,6001) | 5471(4702,6241) |
| 5(most) | 480(345,615) | 3150(2786,3515) | 3317(2929,3705) | 5916(5382,6450) | 5771(5127,6416) |
| Regions | | | | | |
| NE | 165(135,195) | 500(421,580) | 577(472,683) | 1401(1243,1560) | 1703(1488,1918) |
| NW | 127(70,184) | 1757(1583,1930) | 2124(1896,2352) | 4137(3768,4506) | 4270(3803,4738) |
| York&H | 137(93,182) | 903(778,1029) | 1077(907,1248) | 2462(2178,2747) | 3150(2779,3522) |
| EastM | 29(-9,68) | 833(725,941) | 1000(851,1149) | 2087(1839,2334) | 2379(2046,2712) |
| WestM | 139(91,187) | 1169(1042,1296) | 1553(1381,1726) | 3306(3007,3605) | 3509(3095,3923) |
| East | 43(2,85) | 478(347,609) | 966(789,1144) | 1842(1540,2145) | 2782(2324,3240) |
| London | 273(216,330) | 2002(1858,2147) | 1805(1648,1962) | 3356(3120,3592) | 3338(2993,3682) |
| SECoast | 57(23,91) | 632(534,730) | 807(671,944) | 1762(1522,2001) | 2637(2252,3022) |
| SouthC | 29(-3,61) | 392(304,479) | 562(449,674) | 1188(1002,1375) | 1704(1410,1999) |
| SW | 1(-39,42) | 493(375,611) | 600(434,765) | 1330(1046,1614) | 2313(1853,2773) |
| Wales | -39(-71,-7) | 512(420,604) | 563(445,682) | 1422(1233,1611) | 1391(1135,1647) |

### 4 Tables

Table 6: Excess deaths (and 95% CI) by age-group for weeks 11-52 (week ending 13 Mar to week ending 25 Dec 2020), per 100,000 population

COVID/repiratory

| | 15-44 | 45-64 | 65-74 | 75-84 | 85+ |
|---|---|---|---|---|---|
| IMD | | | | | |
| 1(least) | 1(1,2) | 22(21,23) | 107(102,112) | 407(386,427) | 1545(1462,1627) |
| 2 | 2(2,2) | 28(27,29) | 120(114,126) | 435(411,458) | 1567(1485,1649) |
| 3 | 2(2,2) | 36(34,37) | 140(133,148) | 513(488,538) | 1546(1463,1628) |
| 4 | 3(3,3) | 55(53,57) | 204(193,216) | 723(690,756) | 1933(1837,2028) |
| 5(most) | 5(5,6) | 73(69,76) | 306(288,323) | 989(947,1030) | 2524(2423,2625) |
| Regions | | | | | |
| NE | 1(0,1) | 35(32,37) | 177(166,189) | 762(729,795) | 2650(2542,2757) |
| NW | 4(3,4) | 57(54,59) | 235(224,246) | 828(796,861) | 2295(2201,2389) |
| York&H | 2(2,2) | 44(42,46) | 191(181,201) | 724(695,753) | 2311(2226,2396) |
| EastM | 2(2,3) | 38(36,39) | 160(151,168) | 545(518,572) | 1912(1829,1994) |
| WestM | 4(4,4) | 49(47,51) | 196(187,205) | 644(617,671) | 1978(1893,2063) |
| East | 2(2,2) | 31(29,32) | 129(122,136) | 444(421,468) | 1307(1229,1385) |
| London | 5(5,5) | 72(70,73) | 272(265,280) | 829(808,849) | 2016(1949,2083) |
| SECoast | 2(2,2) | 26(24,27) | 113(105,121) | 401(377,426) | 1474(1393,1556) |
| SouthC | 2(1,2) | 24(23,26) | 99(92,106) | 374(352,396) | 1329(1254,1404) |
| SW | 1(0,1) | 15(14,17) | 56(49,63) | 207(185,229) | 888(814,961) |
| Wales | 3(2,3) | 35(32,37) | 148(137,159) | 557(526,588) | 1651(1549,1754) |

Total

| | 15-44 | 45-64 | 65-74 | 75-84 | 85+ |
|---|---|---|---|---|---|
| IMD | | | | | |
| 1(least) | 3(2,5) | 47(40,54) | 189(163,215) | 454(367,541) | 1713(1397,2029) |
| 2 | 4(3,6) | 53(46,61) | 190(161,218) | 403(313,493) | 1725(1413,2036) |
| 3 | 2(-0,4) | 65(56,75) | 217(185,249) | 538(445,631) | 1884(1576,2191) |
| 4 | 6(3,8) | 84(72,96) | 236(194,277) | 826(715,937) | 2174(1842,2505) |
| 5(most) | 8(4,11) | 106(89,123) | 331(276,386) | 1166(1041,1290) | 2789(2442,3137) |
| Regions | | | | | |
| NE | 17(14,21) | 85(72,97) | 204(164,245) | 861(758,963) | 2447(2097,2798) |
| NW | 4(2,7) | 97(86,108) | 303(268,338) | 849(754,944) | 2381(2090,2672) |
| York&H | 7(5,10) | 72(62,82) | 216(182,251) | 720(624,816) | 2349(2046,2651) |
| EastM | 2(-0,4) | 72(62,81) | 226(193,258) | 562(466,657) | 2068(1771,2364) |
| WestM | 6(4,8) | 83(73,93) | 307(273,341) | 765(670,861) | 2425(2115,2735) |
| East | 3(1,5) | 34(25,43) | 179(148,209) | 405(319,492) | 1597(1314,1880) |
| London | 8(6,9) | 96(88,104) | 306(276,337) | 1053(977,1129) | 2165(1916,2414) |
| SECoast | 4(1,6) | 49(40,58) | 182(152,213) | 455(367,543) | 1897(1606,2189) |
| SouthC | 4(2,6) | 42(33,50) | 174(146,202) | 401(322,479) | 1530(1263,1797) |
| SW | 1(-1,3) | 37(28,46) | 109(81,138) | 221(138,304) | 1456(1171,1740) |
| Wales | -3(-6,-0) | 70(58,82) | 181(144,217) | 593(495,690) | 1634(1312,1956) |

### 5 Total

#### 5.1 AASMRs

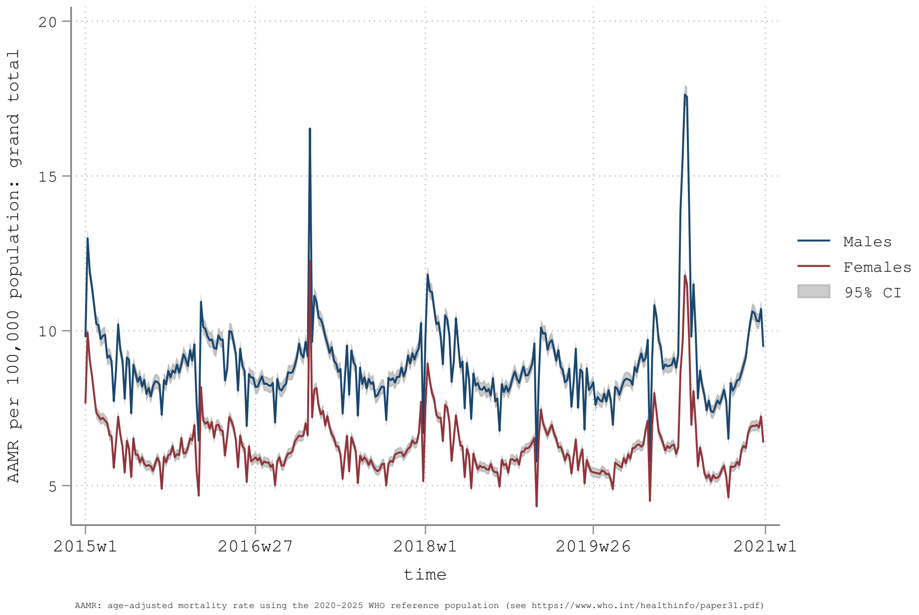


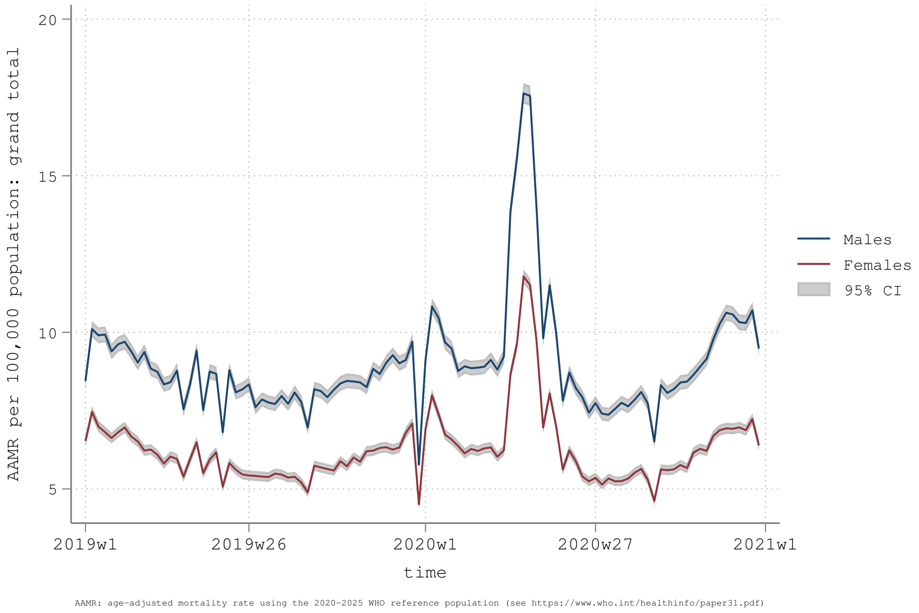

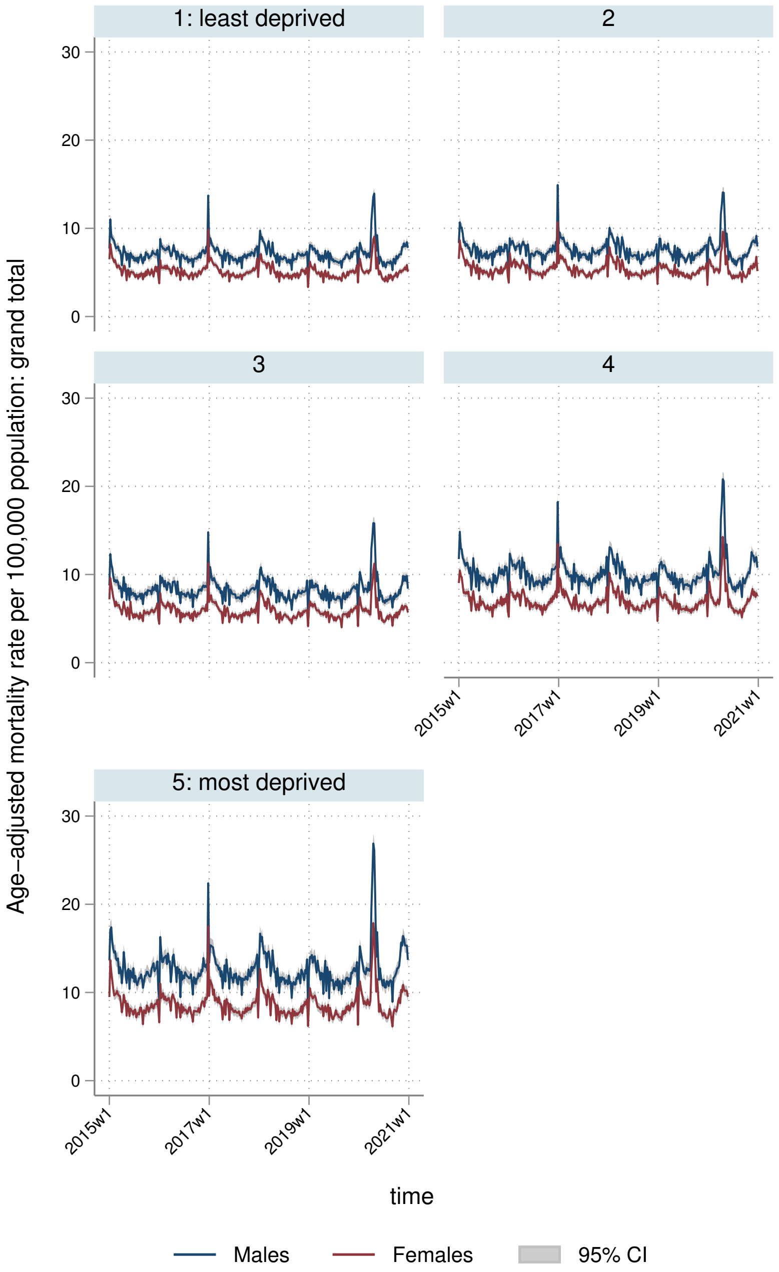
1: least deprived
2
3
4
5: most deprived
Age−adjusted mortality rate per 100,000 population: grand total
30
20
10
0
2015w1
2017w1
2019w1
2021w1
time
Males
Females
95% CI

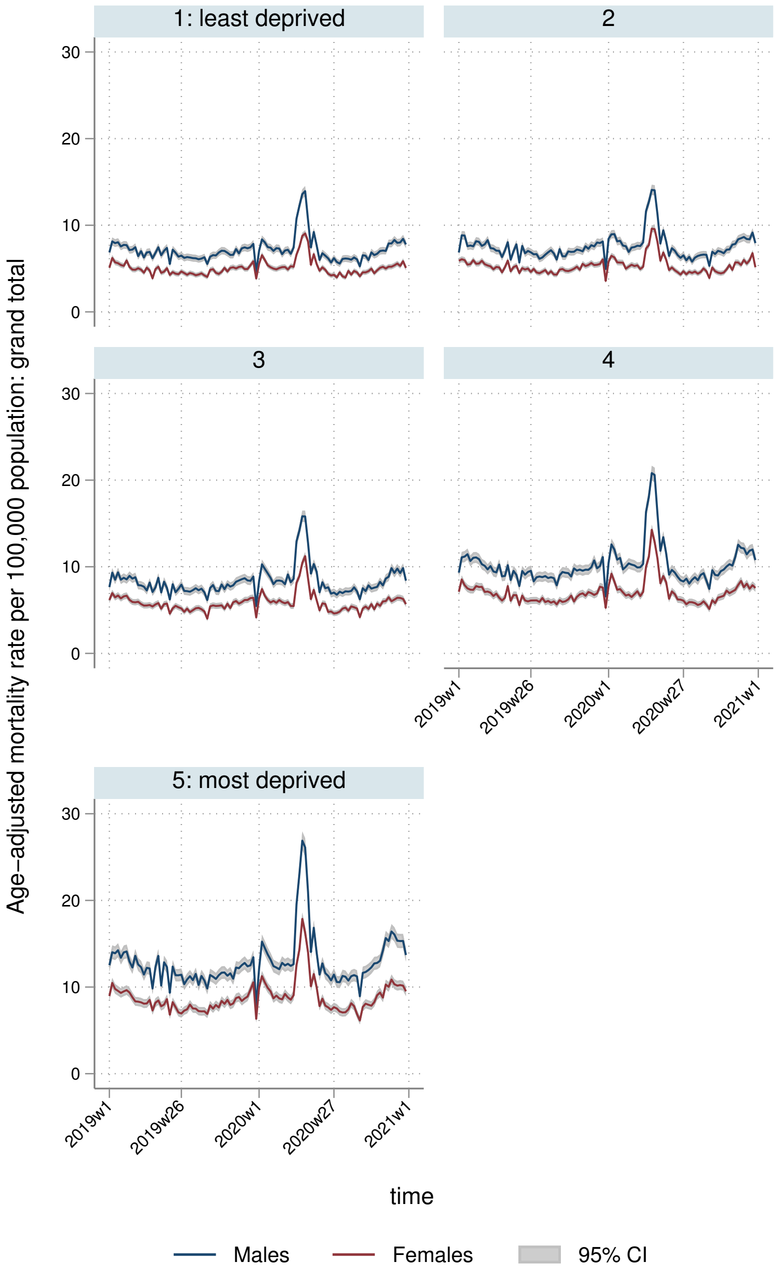
1: least deprived
2
3
4
5: most deprived
Age–adjusted mortality rate per 100,000 population: grand total
0
10
20
30
2019w1
2019w26
2020w1
2020w27
2021w1
time
Males
Females
95% CI

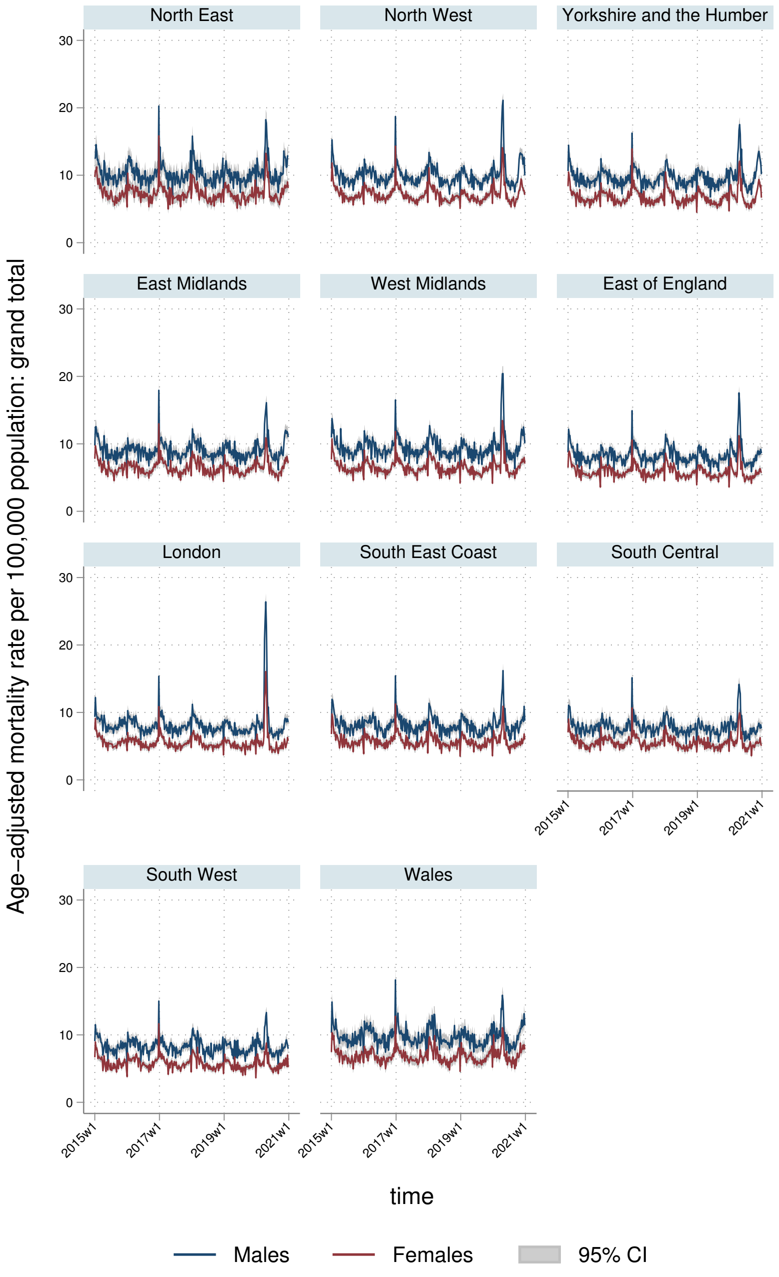
North East
North West
Yorkshire and the Humber
East Midlands
West Midlands
East of England
London
South East Coast
South Central
South West
Wales
Age-adjusted mortality rate per 100,000 population: grand total
0
10
20
30
2015w1
2017w1
2019w1
2021w1
time
Males
Females
95% CI

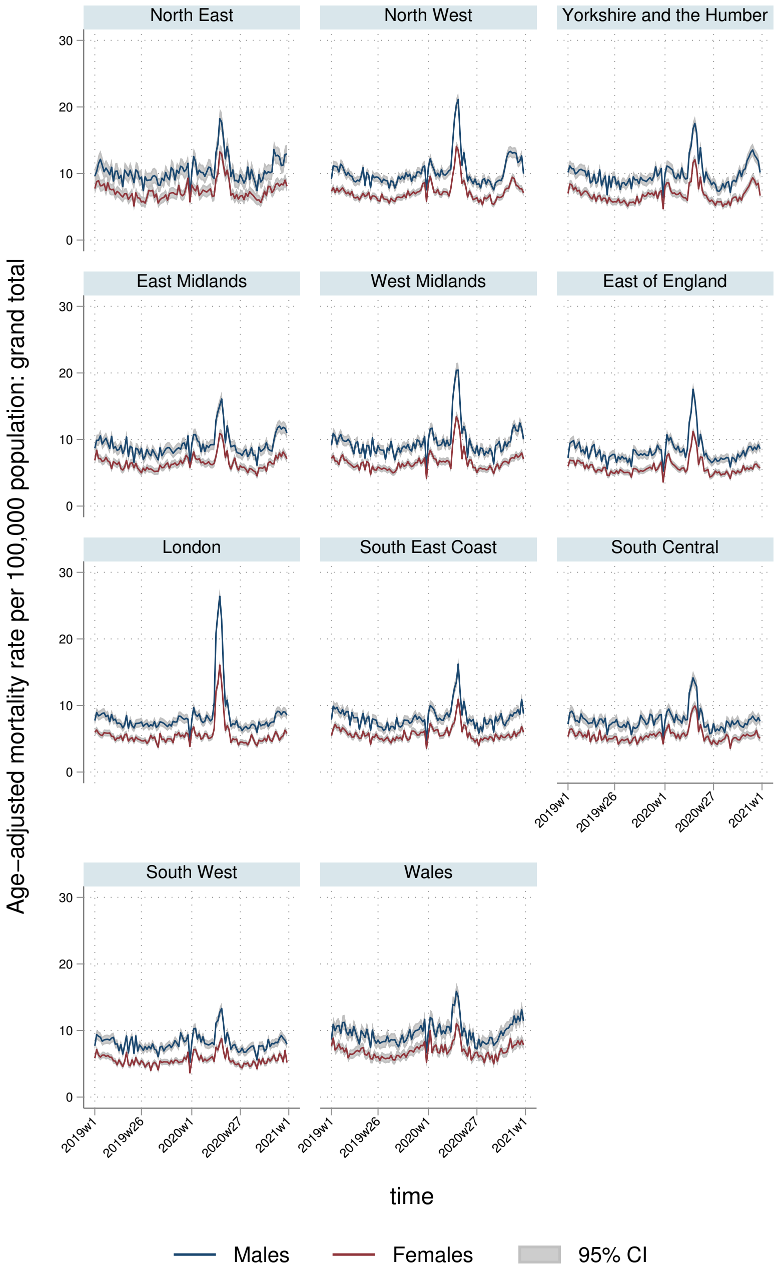
North East
North West
Yorkshire and the Humber
East Midlands
West Midlands
East of England
London
South East Coast
South Central
South West
Wales
Age-adjusted mortality rate per 100,000 population: grand total
30
20
10
0
2019w1
2019w26
2020w1
2020w27
2021w1
time
Males
Females
95% CI

#### 5.2 YLLs

##### 5.2.1 England-Wales aggregate

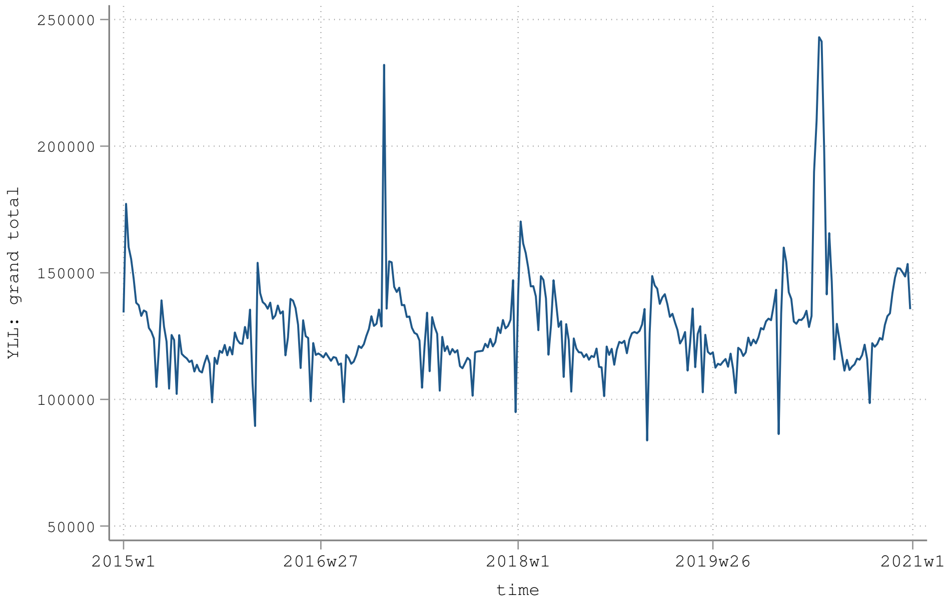


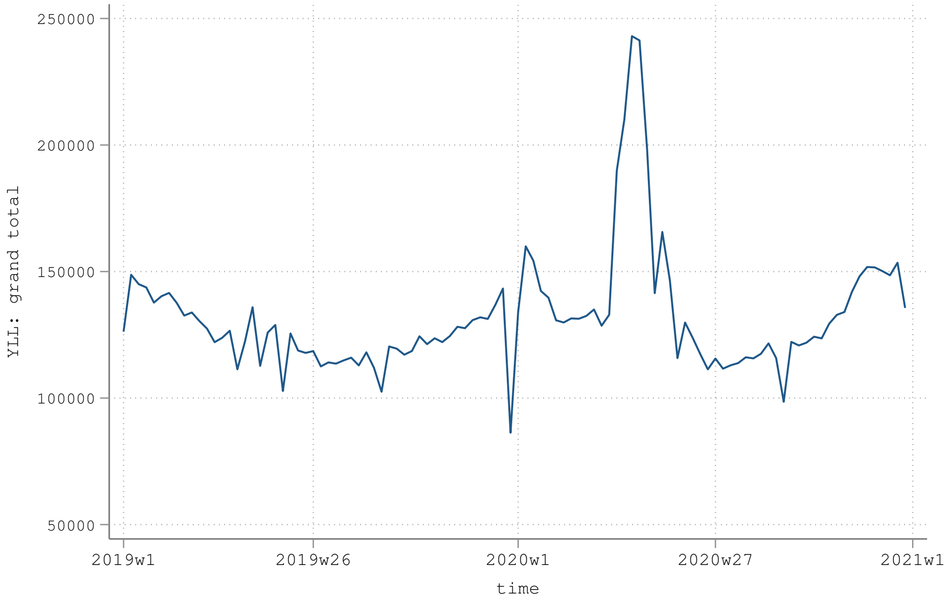

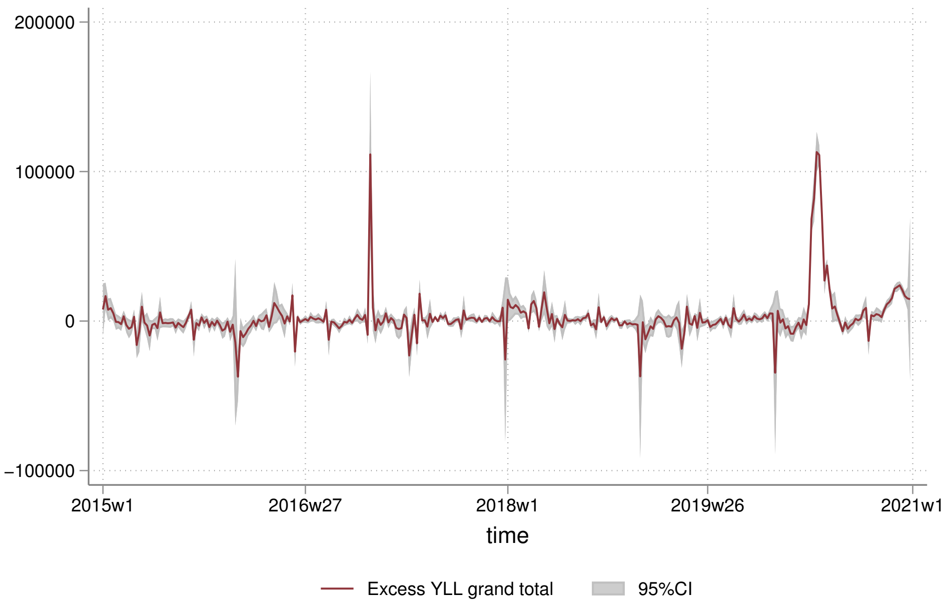

200000
100000
0
−100000
2015w1
2016w27
2018w1
2019w26
2021w1
time
Excess YLL grand total
95%CI


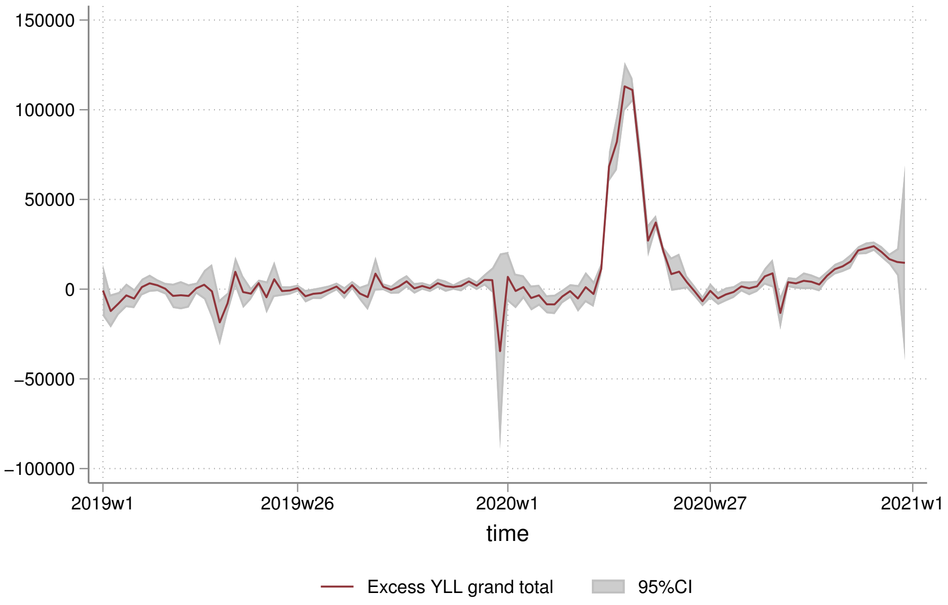

150000
100000
50000
0
−50000
−100000
2019w1
2019w26
2020w1
2020w27
2021w1
time
Excess YLL grand total
95%CI

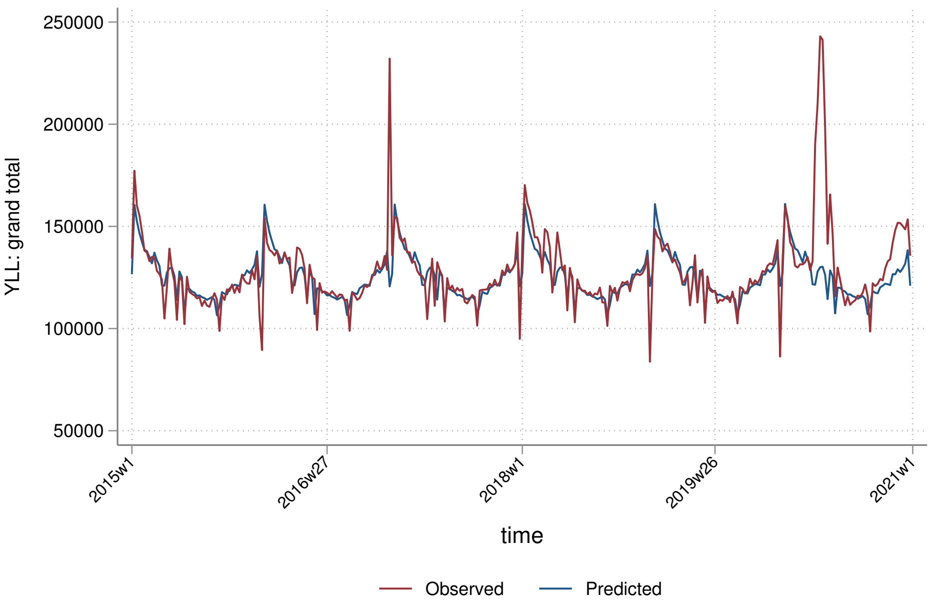
250000
200000
150000
100000
50000
YLL: grand total
2015w1
2016w27
2018w1
2019w26
2021w1
time
Observed
Predicted

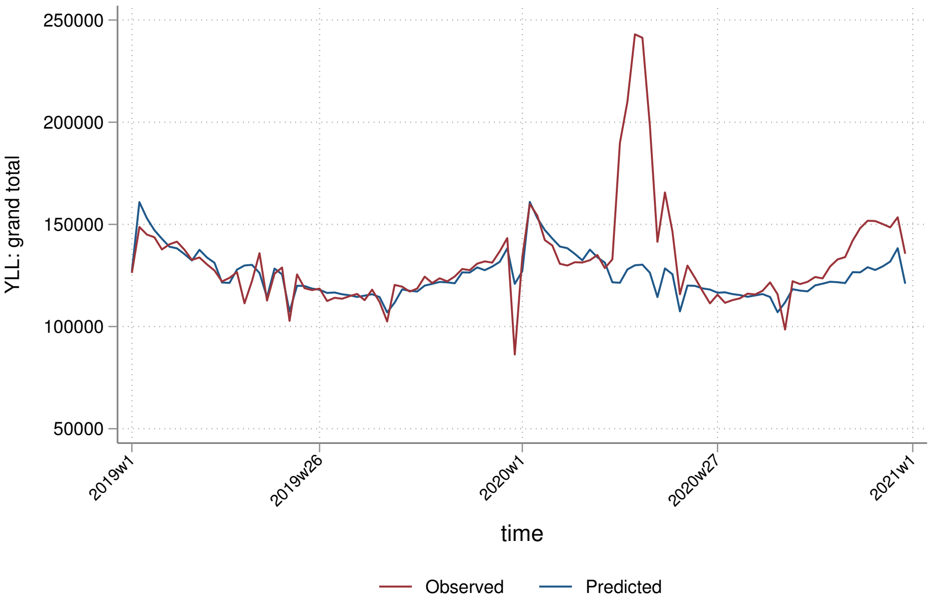
250000
200000
150000
100000
50000
YLL: grand total
2019w1
2019w26
2020w1
2020w27
2021w1
time
Observed
Predicted

##### 5.2.2 By sex

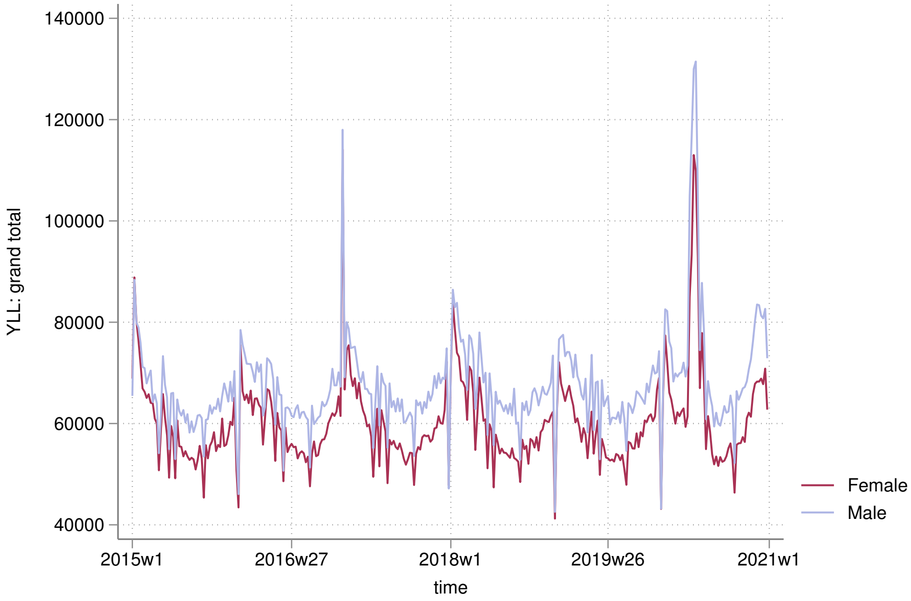

140000
120000
100000
80000
60000
40000
YLL: grand total
2015w1
2016w27
2018w1
2019w26
2021w1
time
Female
Male


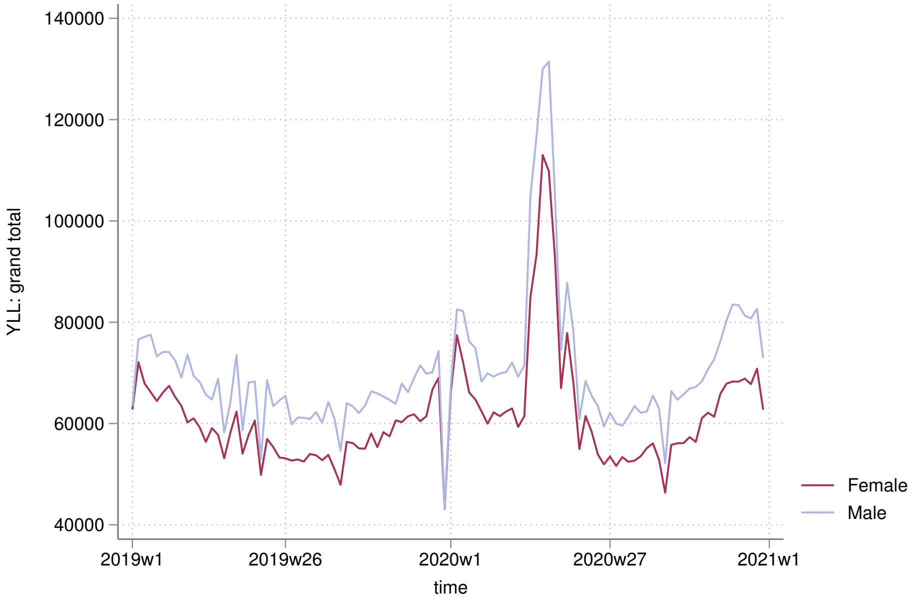

140000
120000
100000
80000
60000
40000
YLL: grand total
2019w1
2019w26
2020w1
2020w27
2021w1
time
Female
Male

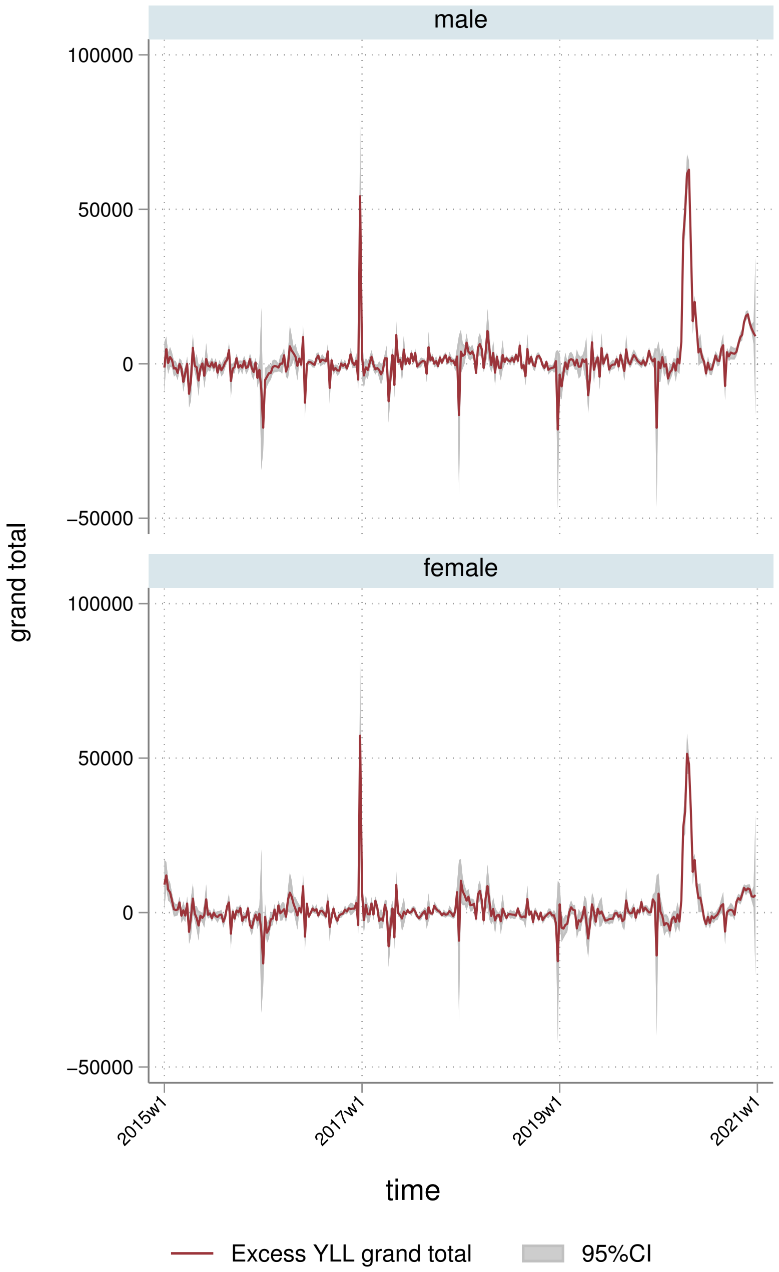
male
female
100000
50000
0
−50000
grand total
2015w1
2017w1
2019w1
2021w1
time
Excess YLL grand total
95%CI

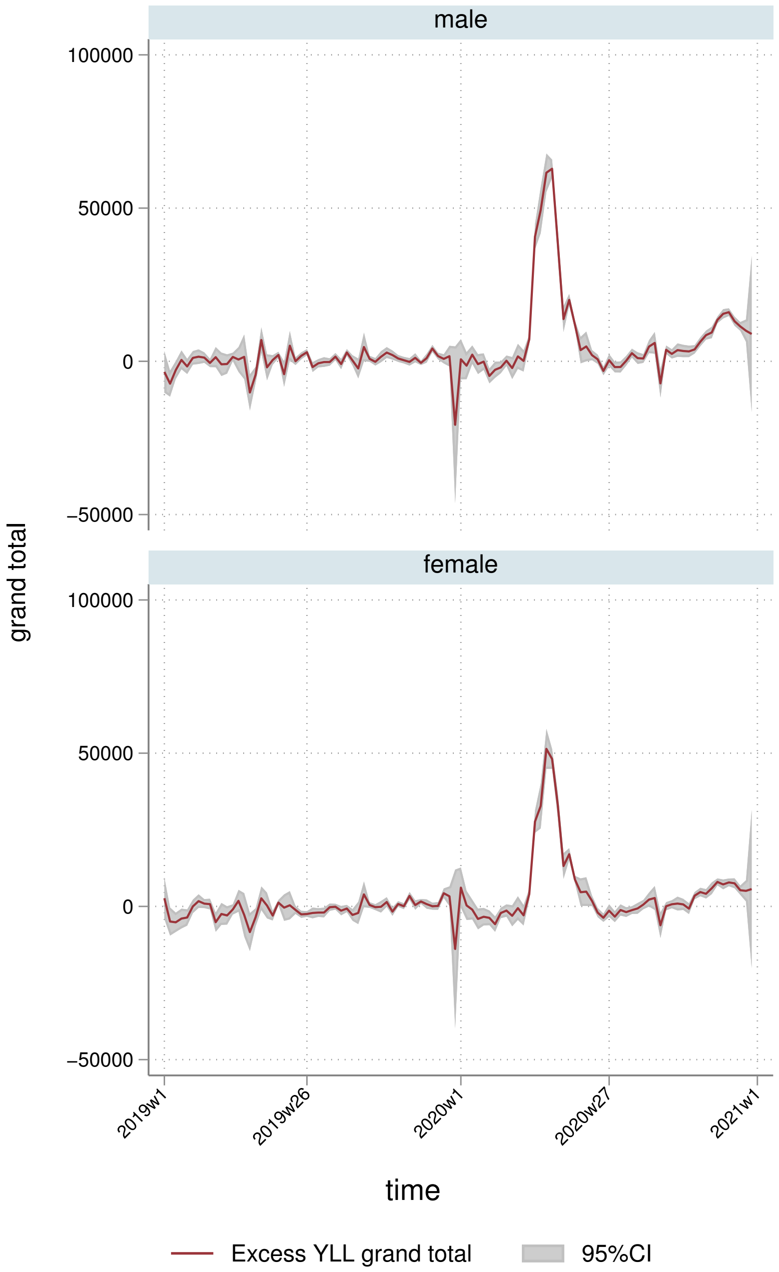
male
female
100000
50000
0
−50000
grand total
2019w1
2019w26
2020w1
2020w27
2021w1
time
Excess YLL grand total
95%CI

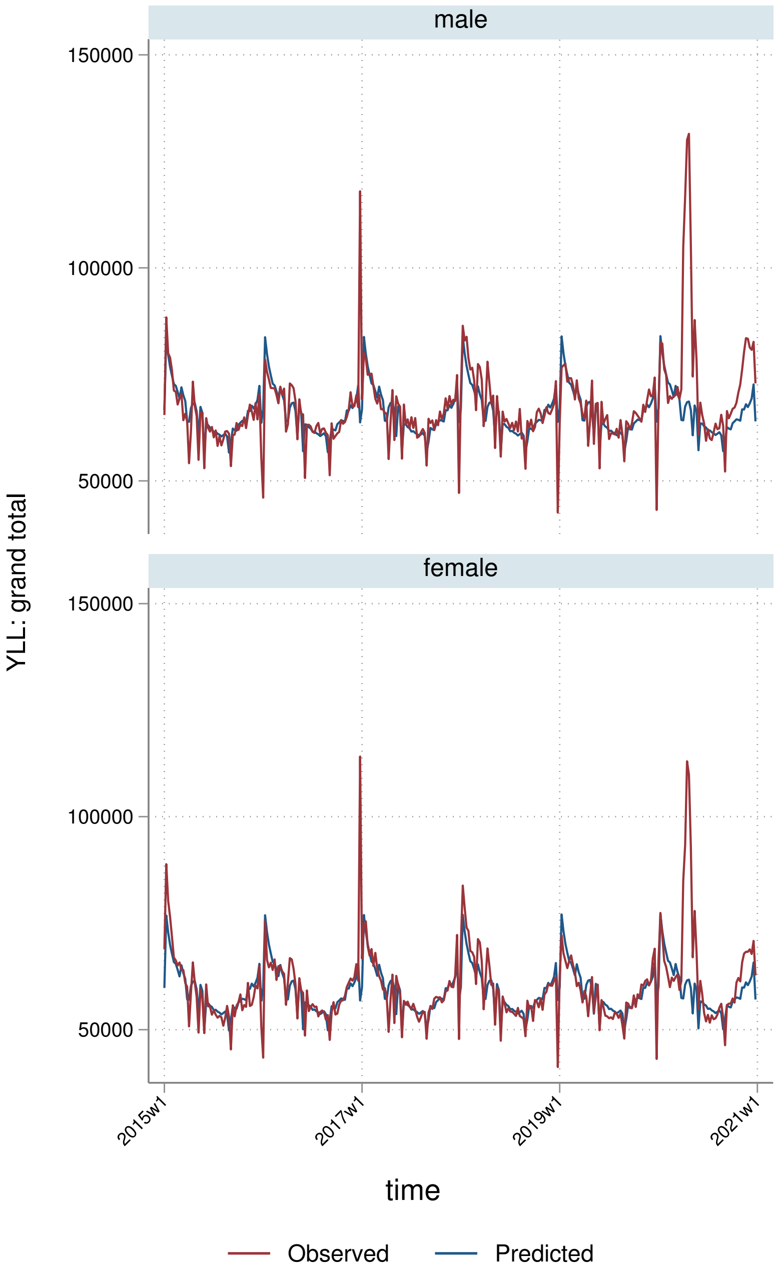

male
female
150000
100000
50000
150000
100000
50000
YLL: grand total
2015w1
2017w1
2019w1
2021w1
time
Observed
Predicted

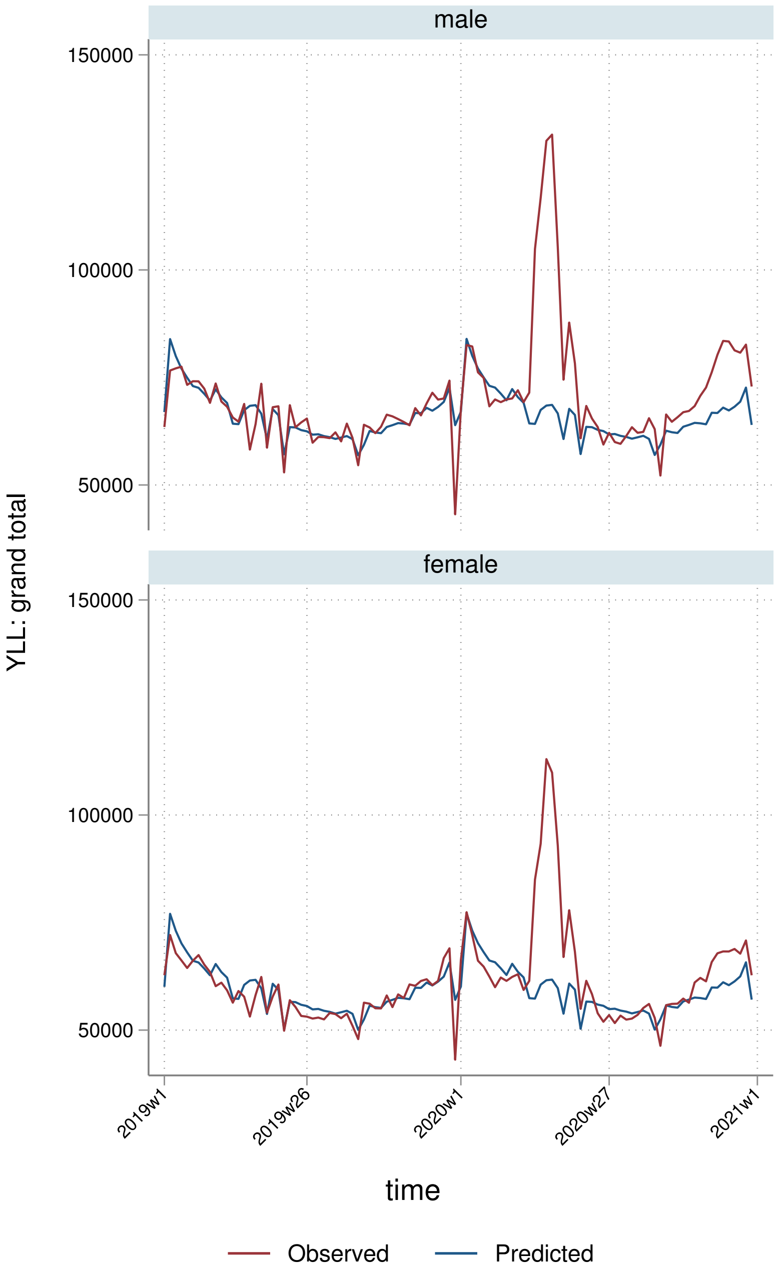
male
female
150000
100000
50000
YLL: grand total
2019w1
2019w26
2020w1
2020w27
2021w1
time
Observed
Predicted

##### 5.2.3 By deprivation quintile

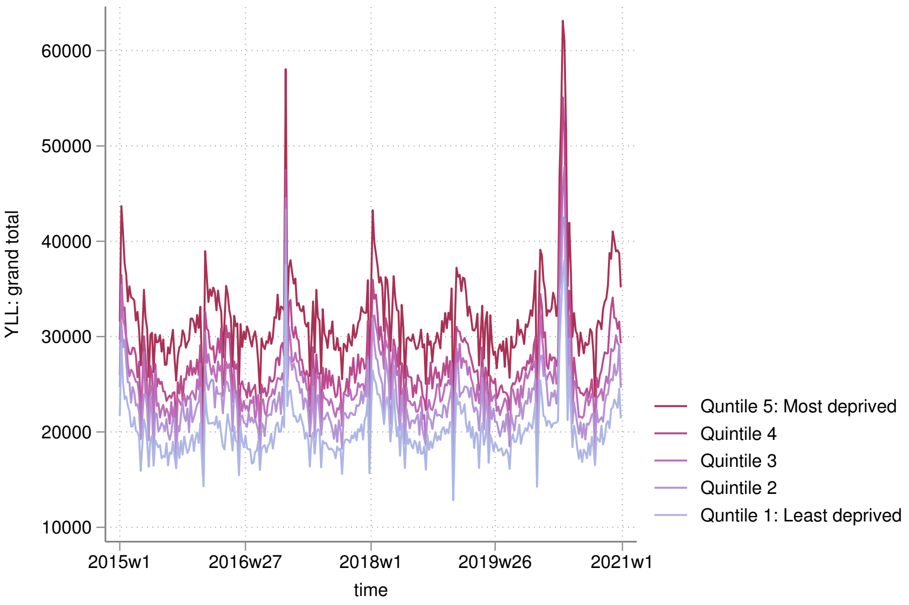


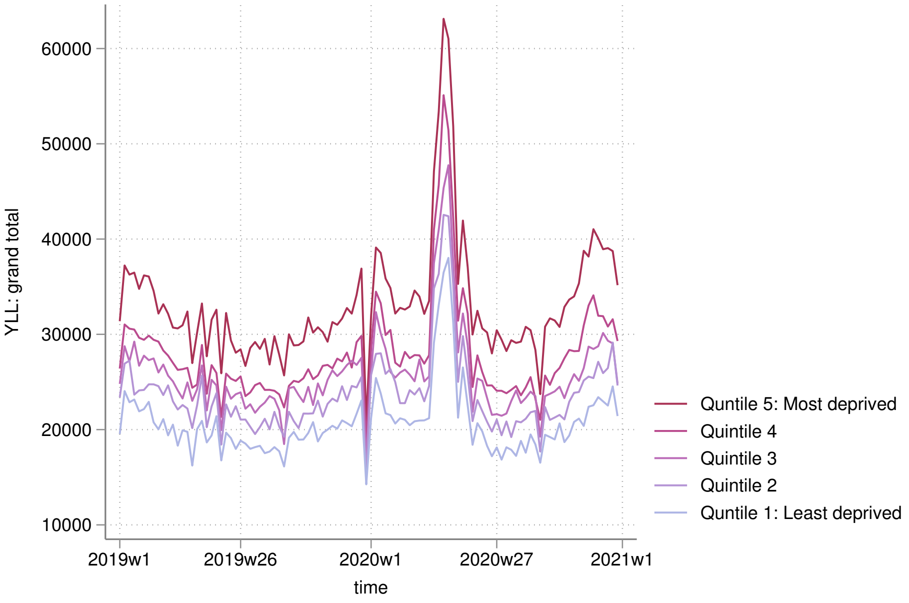

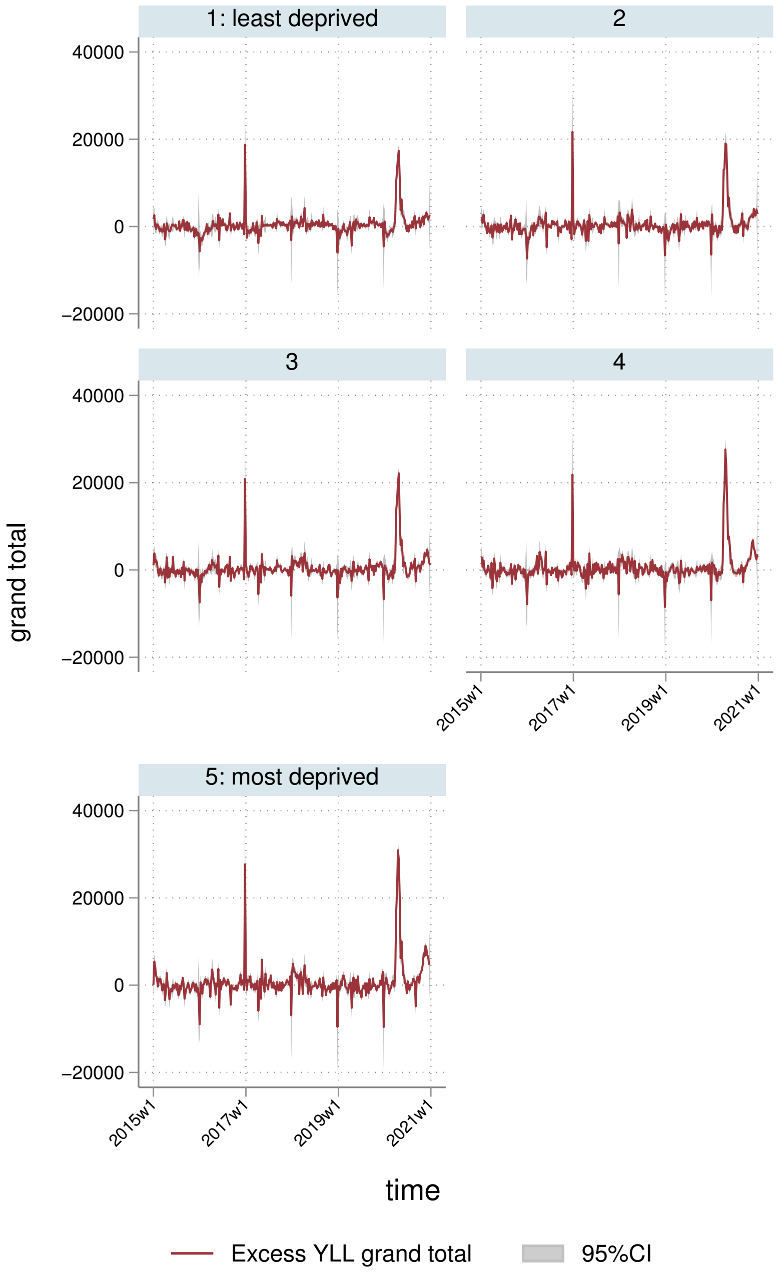
1: least deprived
2
3
4
5: most deprived
40000
20000
0
−20000
2015w1
2017w1
2019w1
2021w1
grand total
time
Excess YLL grand total
95%CI

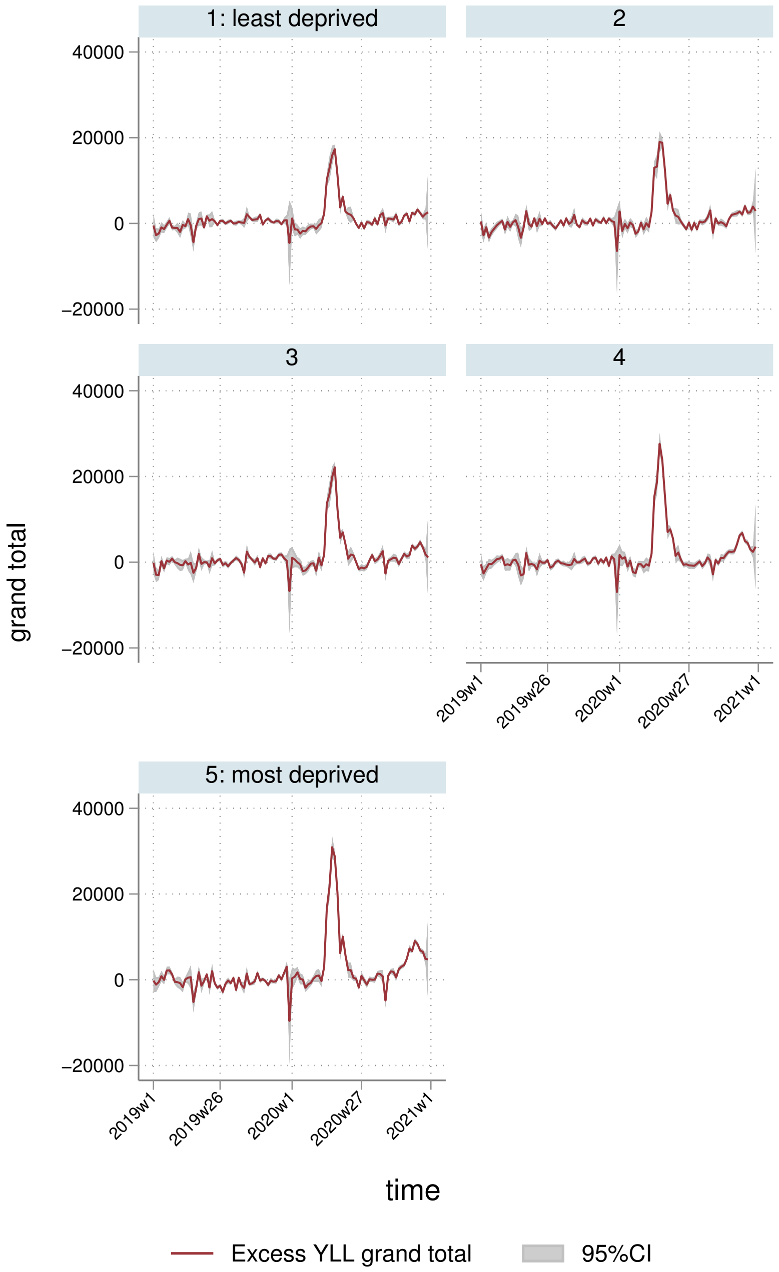

1: least deprived
2
3
4
5: most deprived
40000
20000
0
−20000
grand total
2019w1
2019w26
2020w1
2020w27
2021w1
time
Excess YLL grand total
95%CI

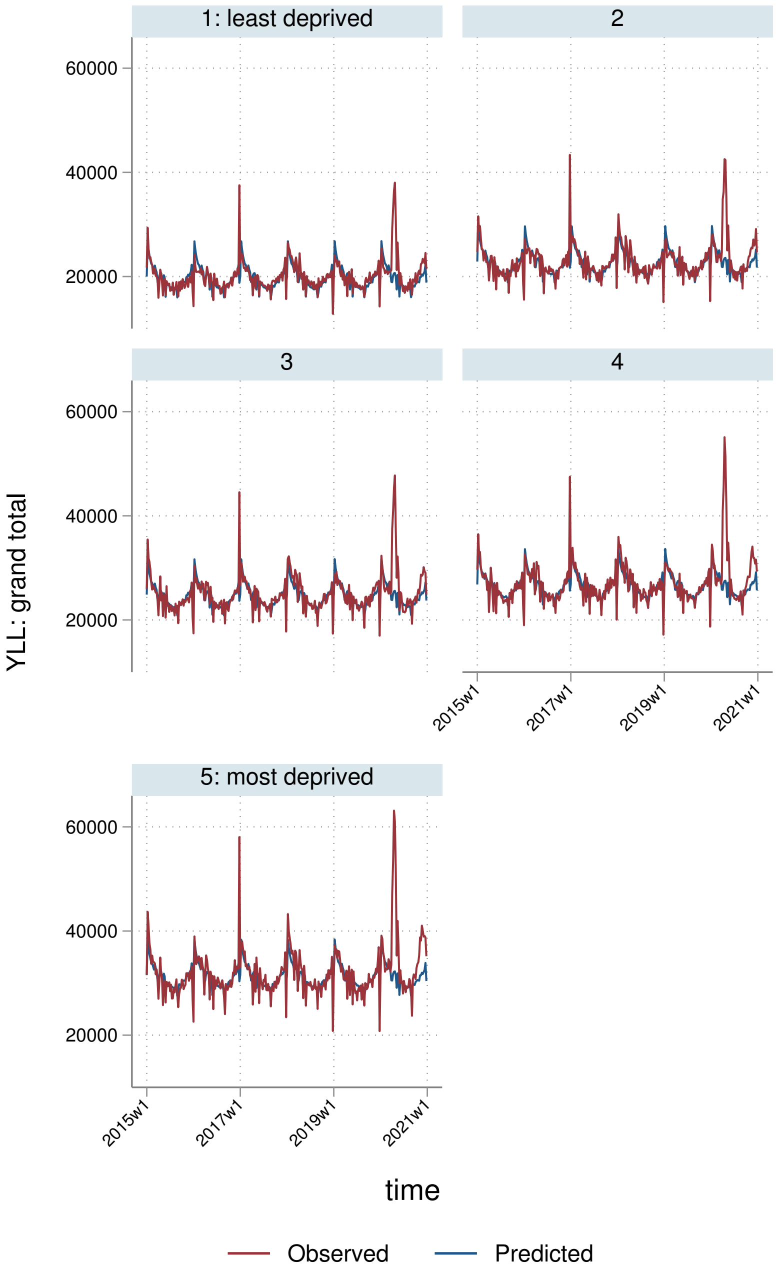
1: least deprived
2
3
4
5: most deprived
60000
40000
20000
YLL: grand total
2015w1
2017w1
2019w1
2021w1
time
Observed
Predicted

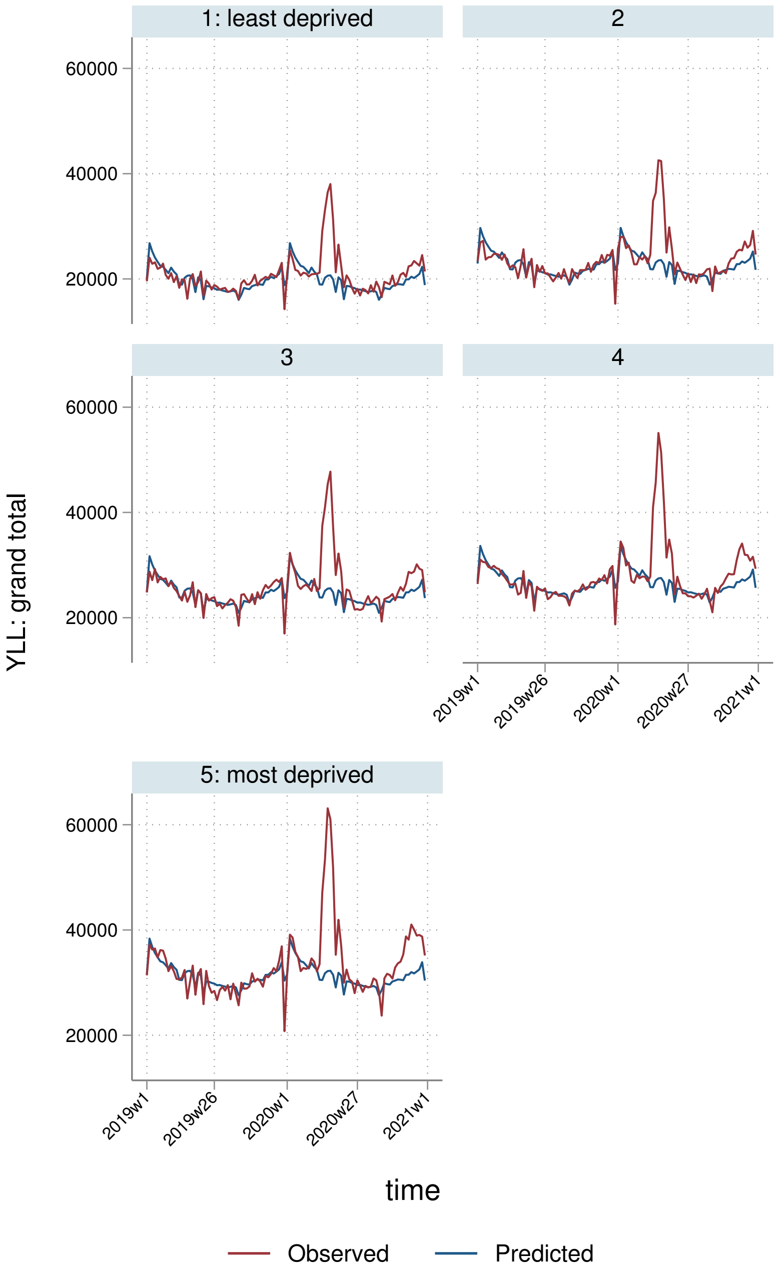
1: least deprived
2
3
4
5: most deprived
60000
40000
20000
YLL: grand total
2019w1
2019w26
2020w1
2020w27
2021w1
time
Observed
Predicted

##### 5.2.4 By Strategic Health Authority

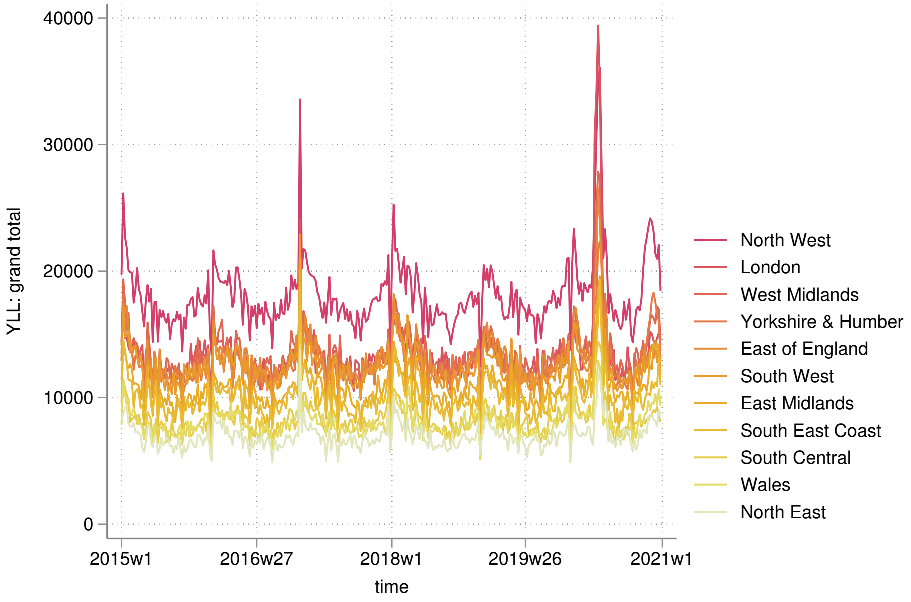


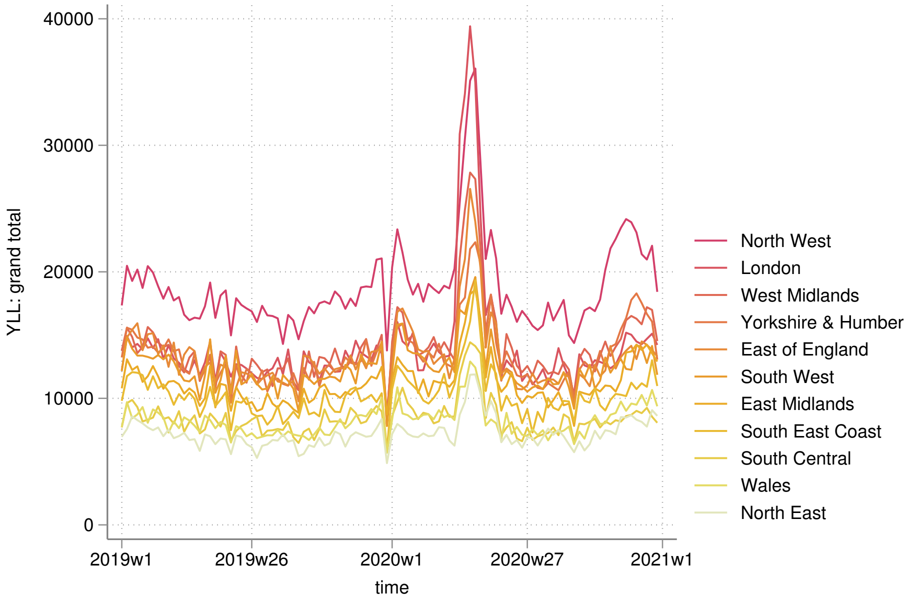

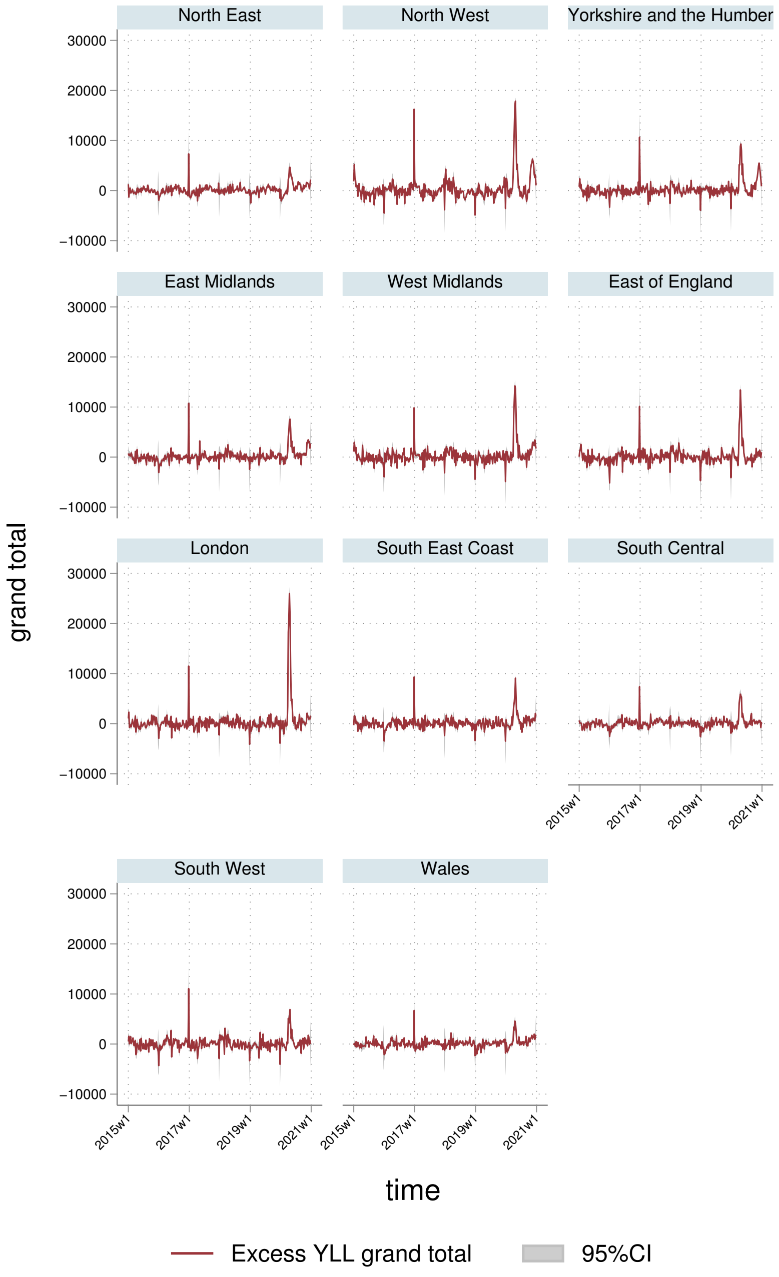
North East
North West
Yorkshire and the Humber
East Midlands
West Midlands
East of England
London
South East Coast
South Central
South West
Wales
30000
20000
10000
0
−10000
grand total
2015w1
2017w1
2019w1
2021w1
time
Excess YLL grand total
95%CI

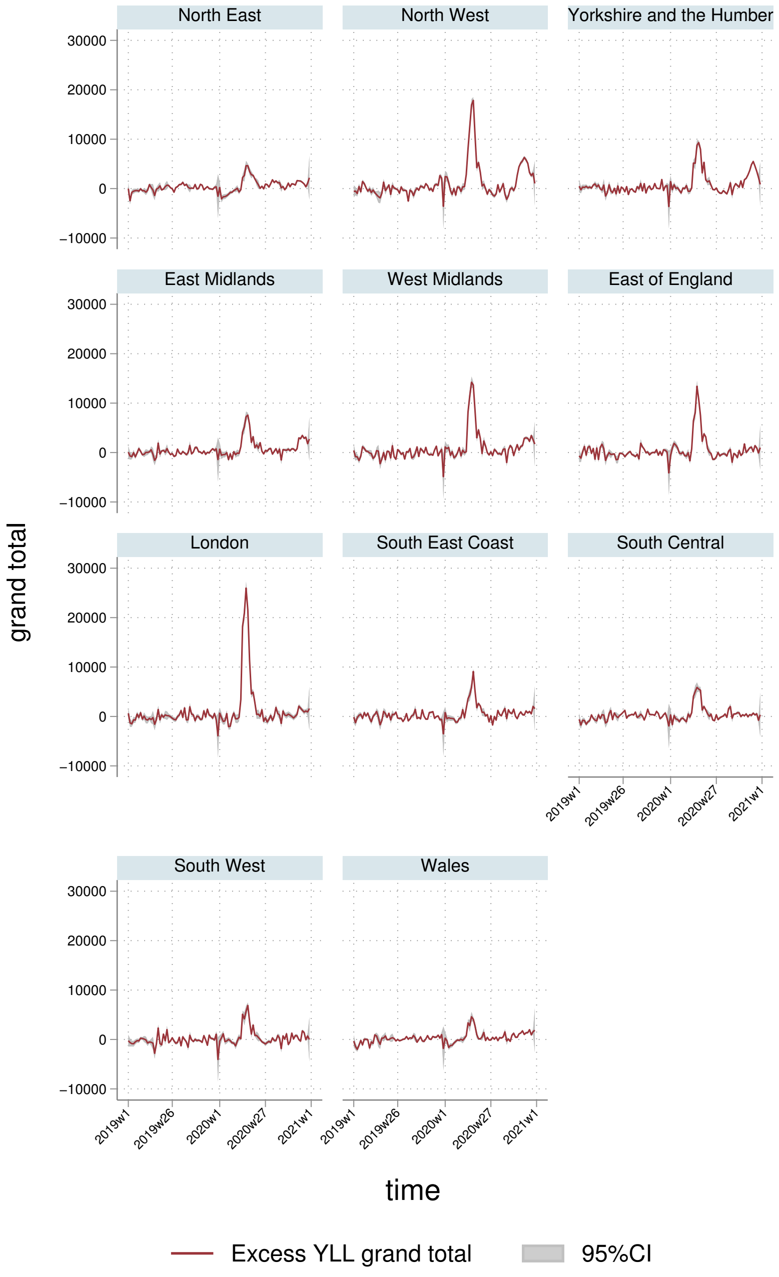
North East
North West
Yorkshire and the Humber
East Midlands
West Midlands
East of England
London
South East Coast
South Central
South West
Wales
30000
20000
10000
0
−10000
grand total
2019w1
2019w26
2020w1
2020w27
2021w1
time
Excess YLL grand total
95%CI

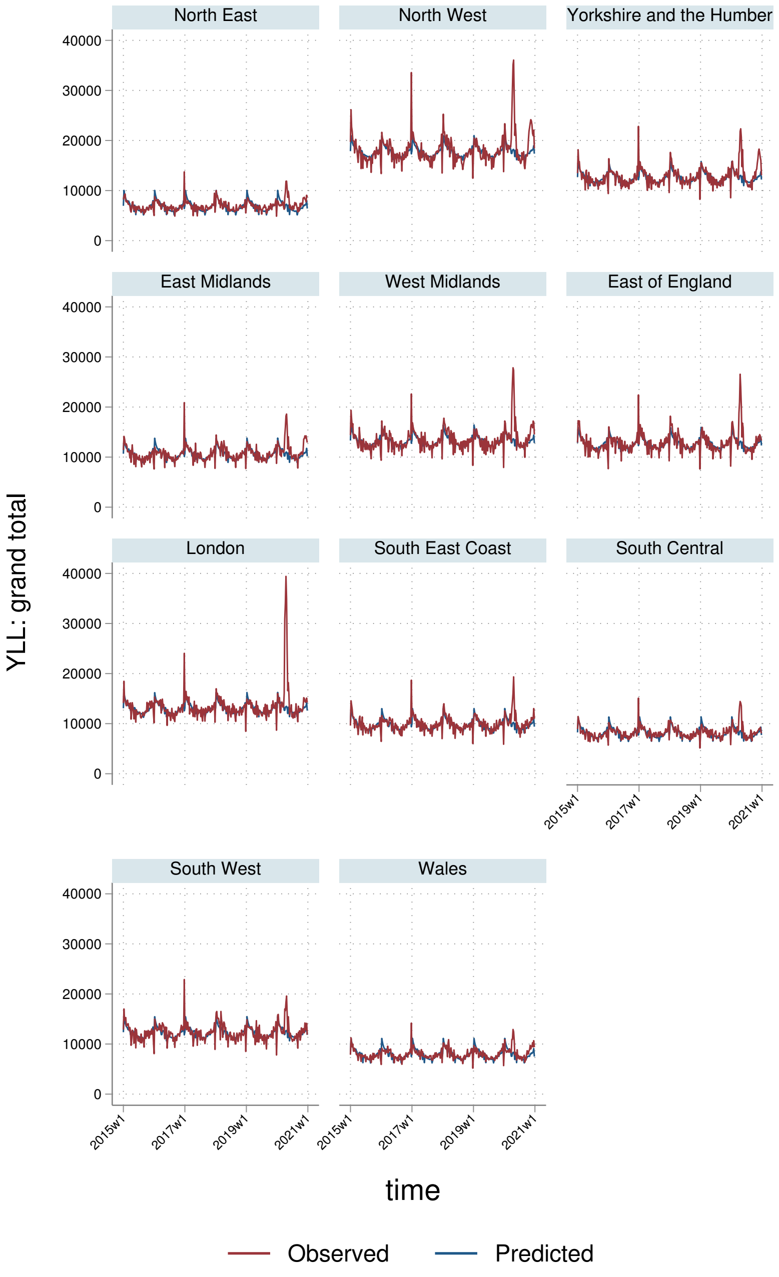
North East
North West
Yorkshire and the Humber
East Midlands
West Midlands
East of England
London
South East Coast
South Central
South West
Wales
YLL: grand total
0
10000
20000
30000
40000
2015w1
2017w1
2019w1
2021w1
time
Observed
Predicted

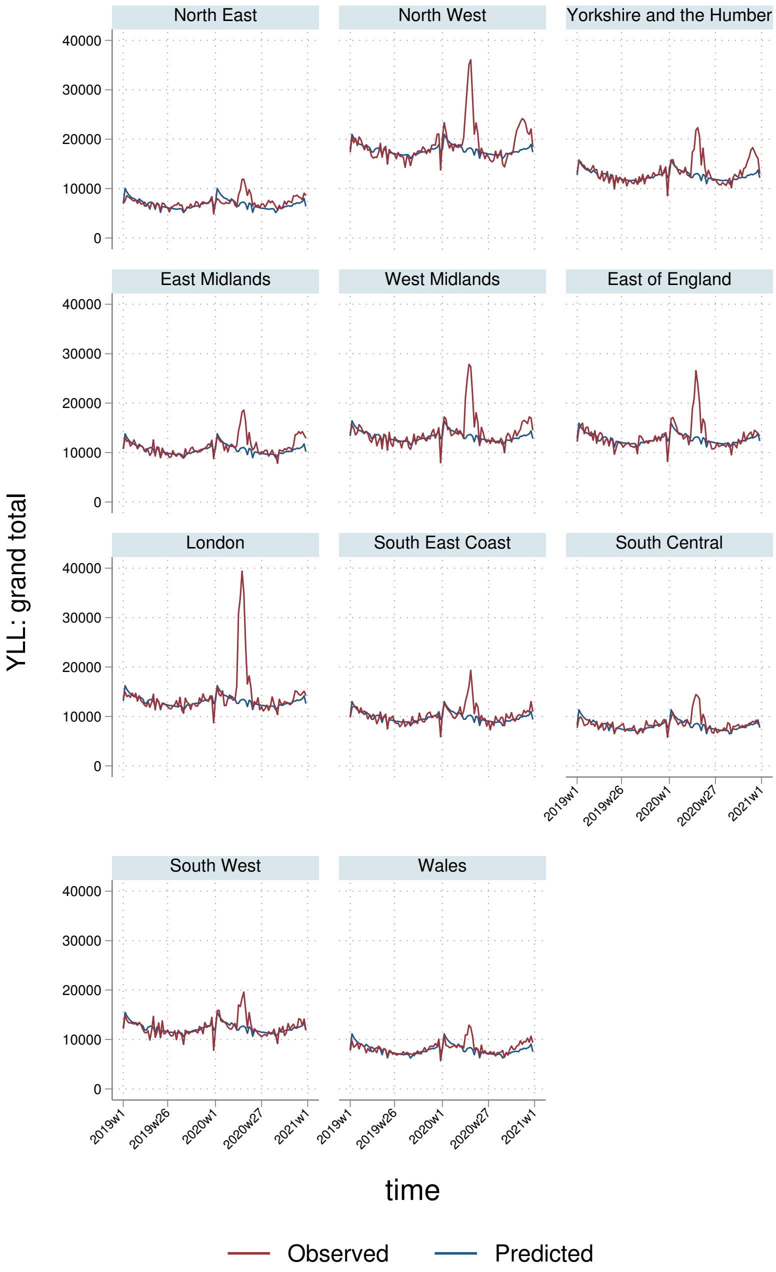

North East
North West
Yorkshire and the Humber
East Midlands
West Midlands
East of England
London
South East Coast
South Central
South West
Wales
YLL: grand total
0
10000
20000
30000
40000
2019w1
2019w26
2020w1
2020w27
2021w1
time
Observed
Predicted

#### 5.3 YLLs per $100,000$ population

##### 5.3.1 England-Wales aggregate

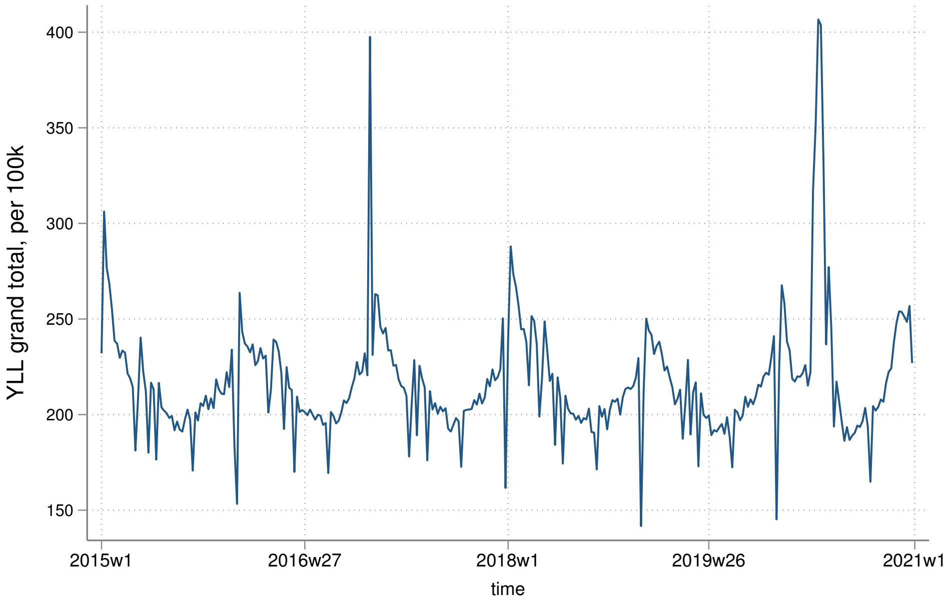


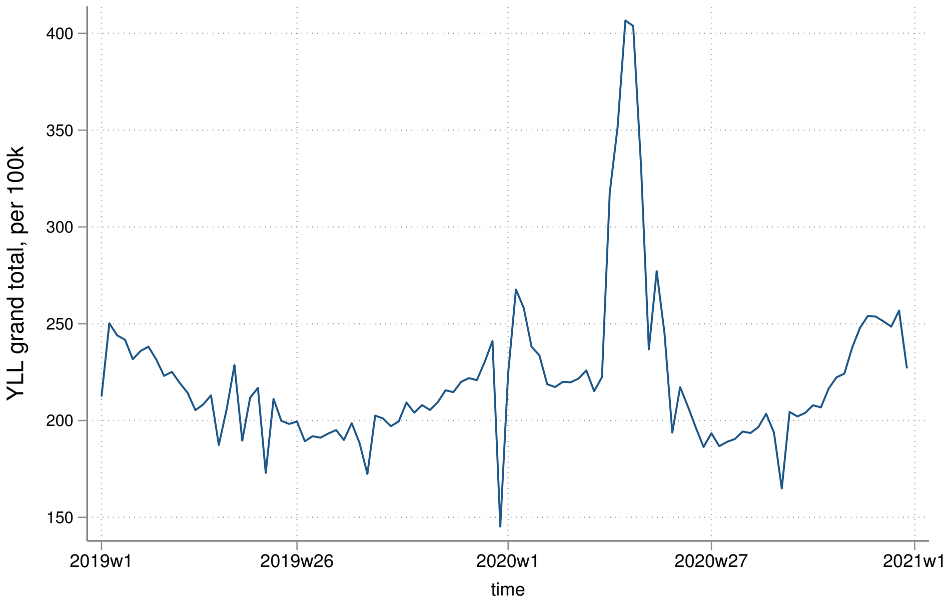

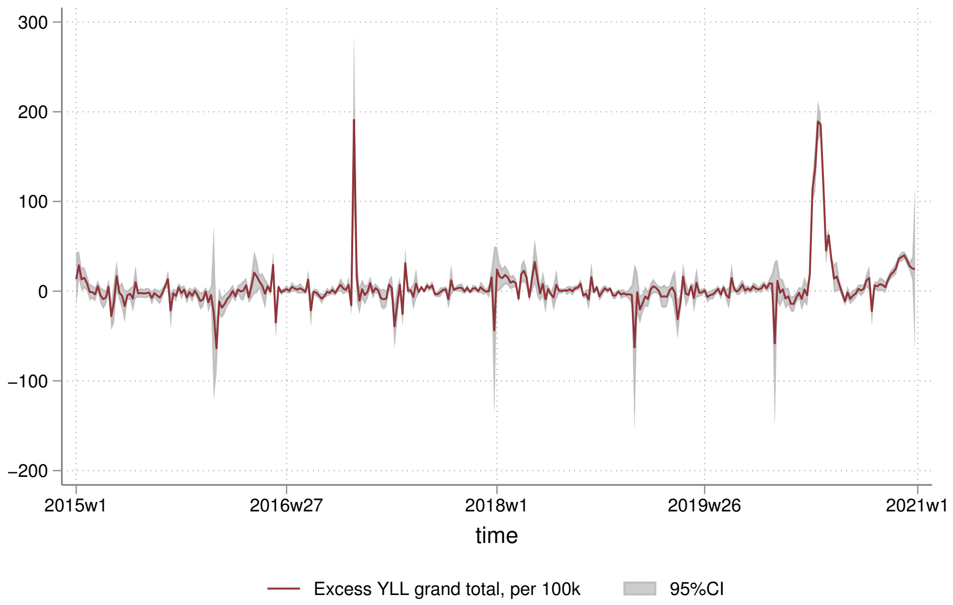
300
200
100
0
−100
−200
2015w1
2016w27
2018w1
2019w26
2021w1
time
Excess YLL grand total, per 100k
95%CI

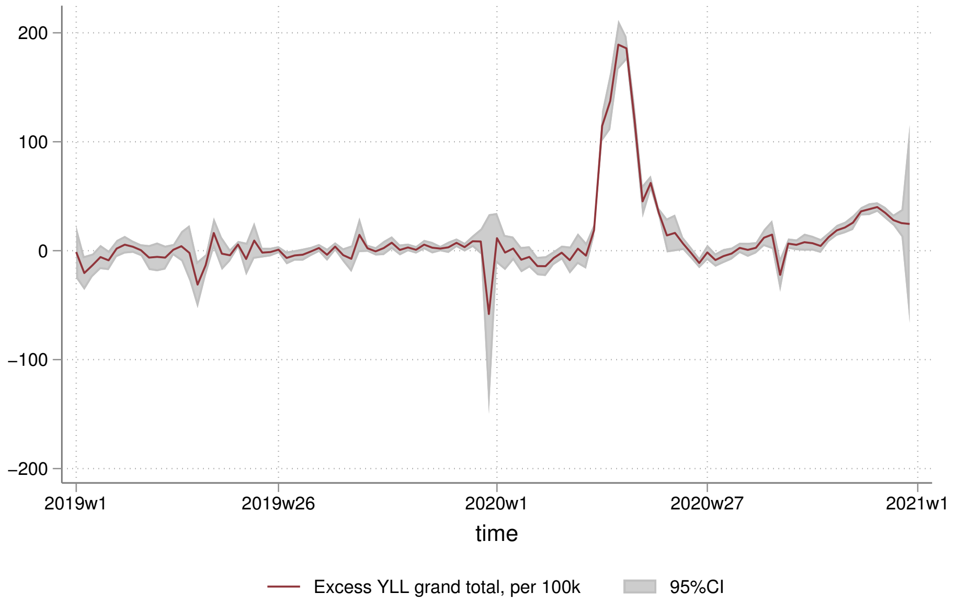
200
100
0
−100
−200
2019w1
2019w26
2020w1
2020w27
2021w1
time
Excess YLL grand total, per 100k
95%CI

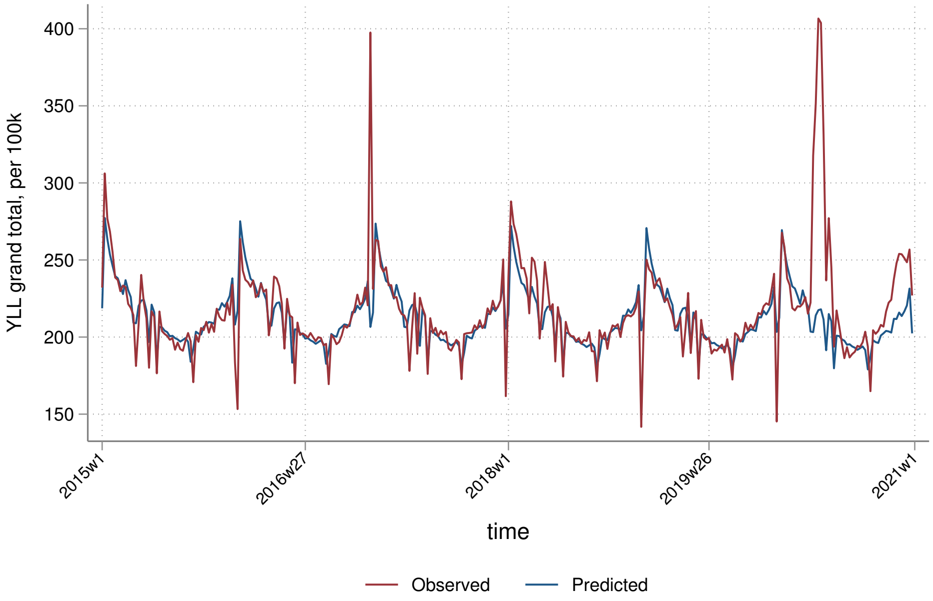
YLL grand total, per 100k
400
350
300
250
200
150
2015w1
2016w27
2018w1
2019w26
2021w1
time
Observed
Predicted

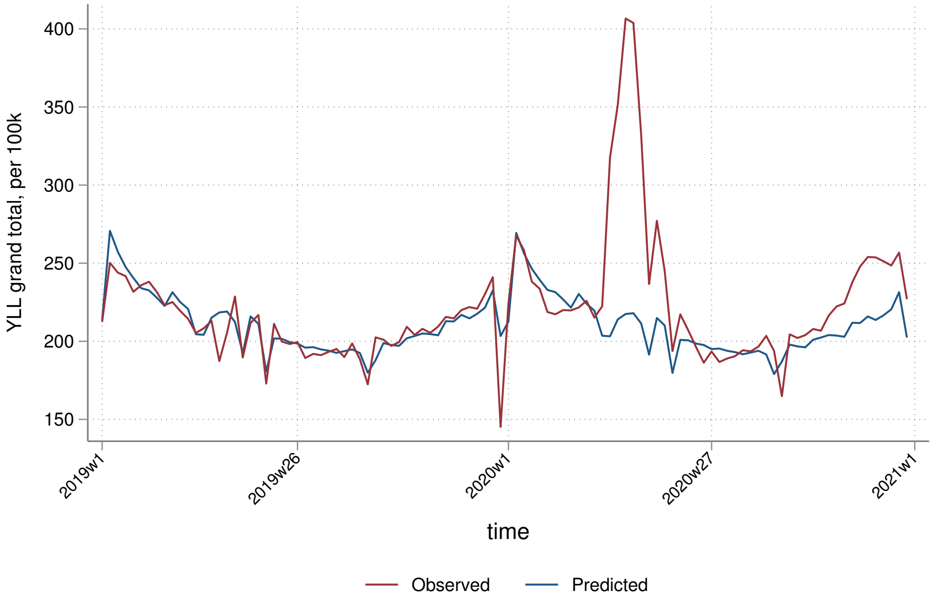
YLL grand total, per 100k
400
350
300
250
200
150
2019w1
2019w26
2020w1
2020w27
2021w1
time
Observed
Predicted

##### 5.3.2 By sex

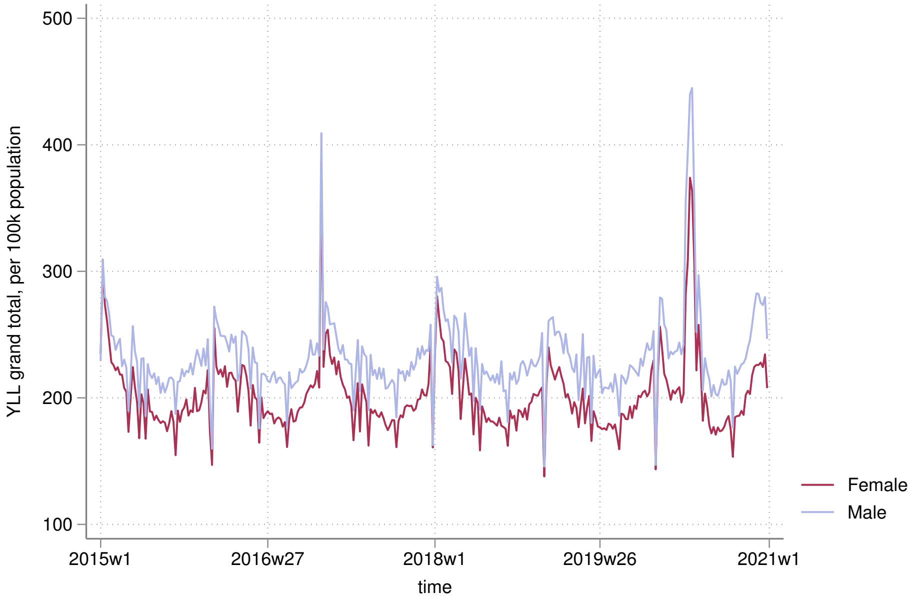

500
400
300
200
100
YLL grand total, per 100k population
2015w1
2016w27
2018w1
2019w26
2021w1
time
Female
Male


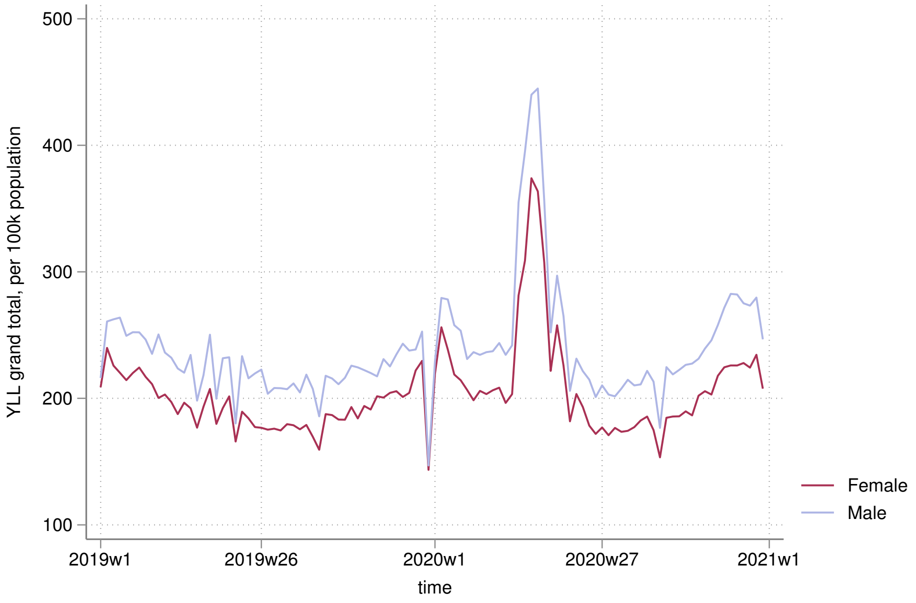

500
400
300
200
100
YLL grand total, per 100k population
2019w1
2019w26
2020w1
2020w27
2021w1
time
Female
Male

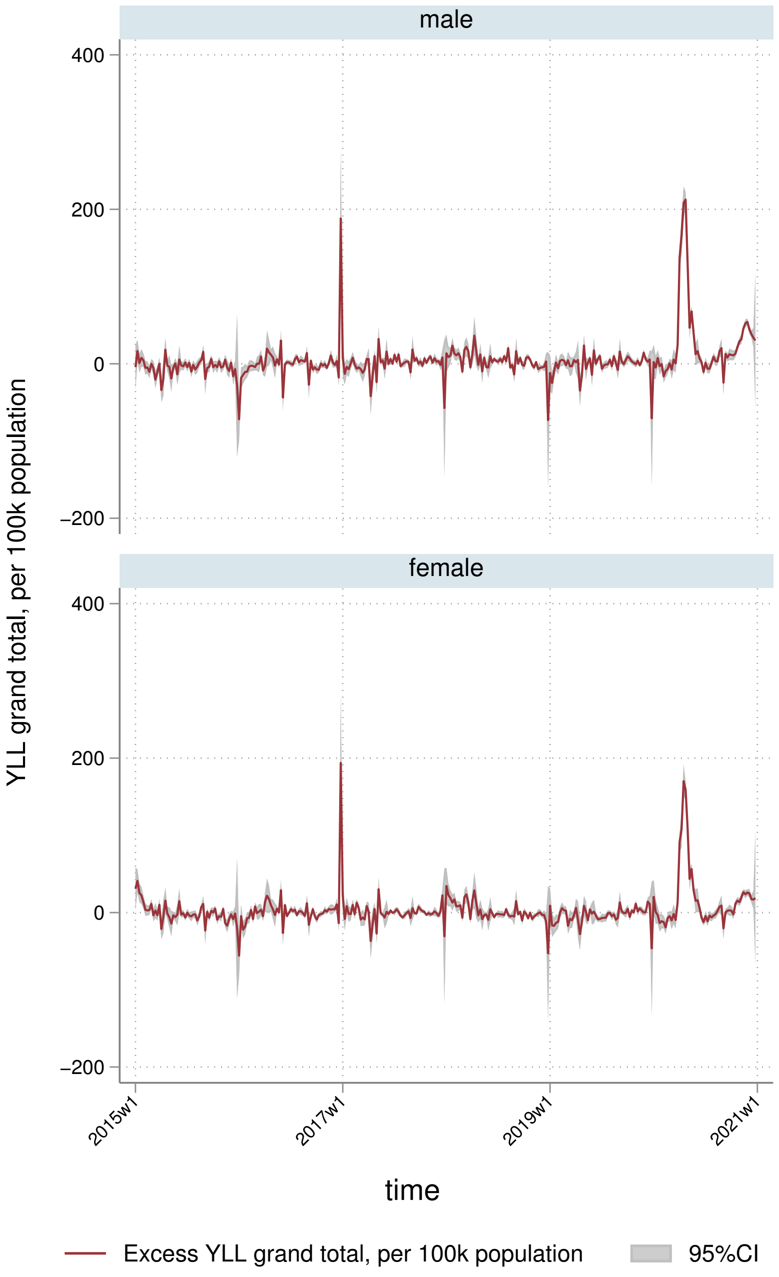
male
female
400
200
0
−200
YLL grand total, per 100k population
2015w1
2017w1
2019w1
2021w1
time
Excess YLL grand total, per 100k population
95%CI

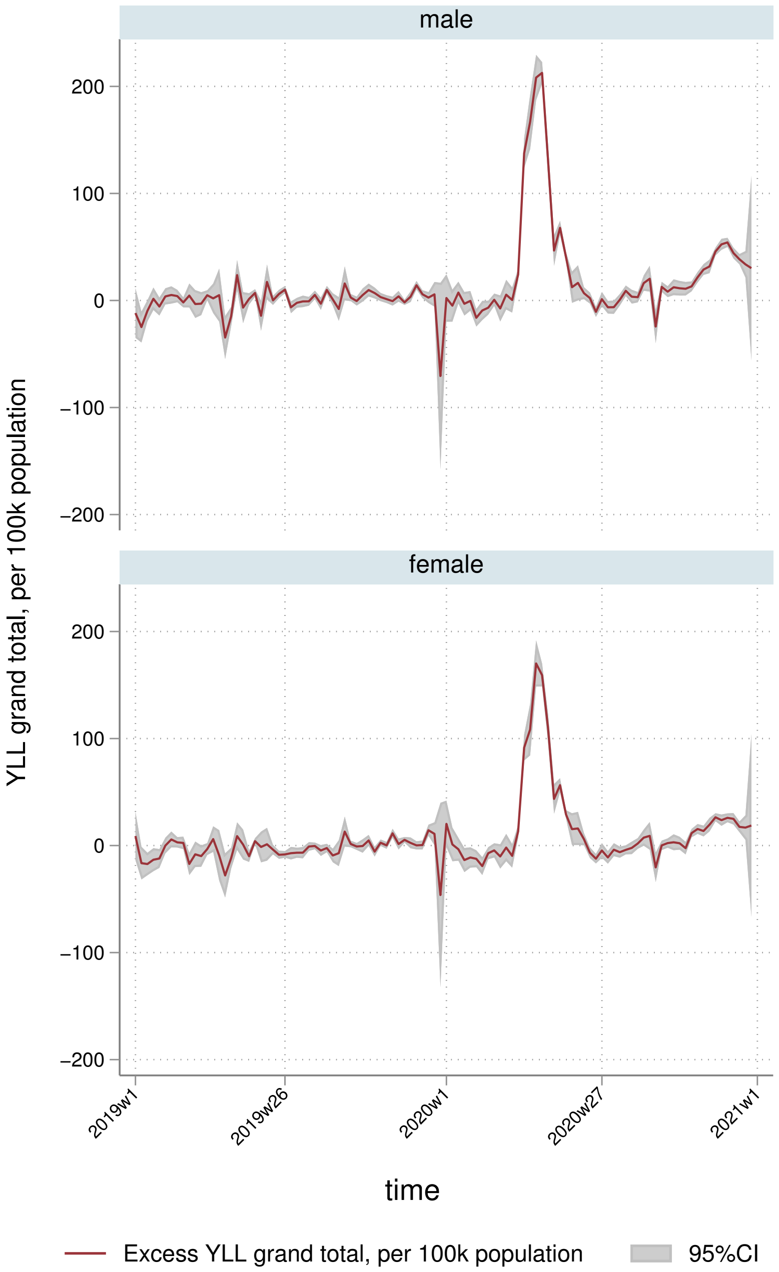
male
female
200
100
0
−100
−200
YLL grand total, per 100k population
2019w1
2019w26
2020w1
2020w27
2021w1
time
Excess YLL grand total, per 100k population
95%CI

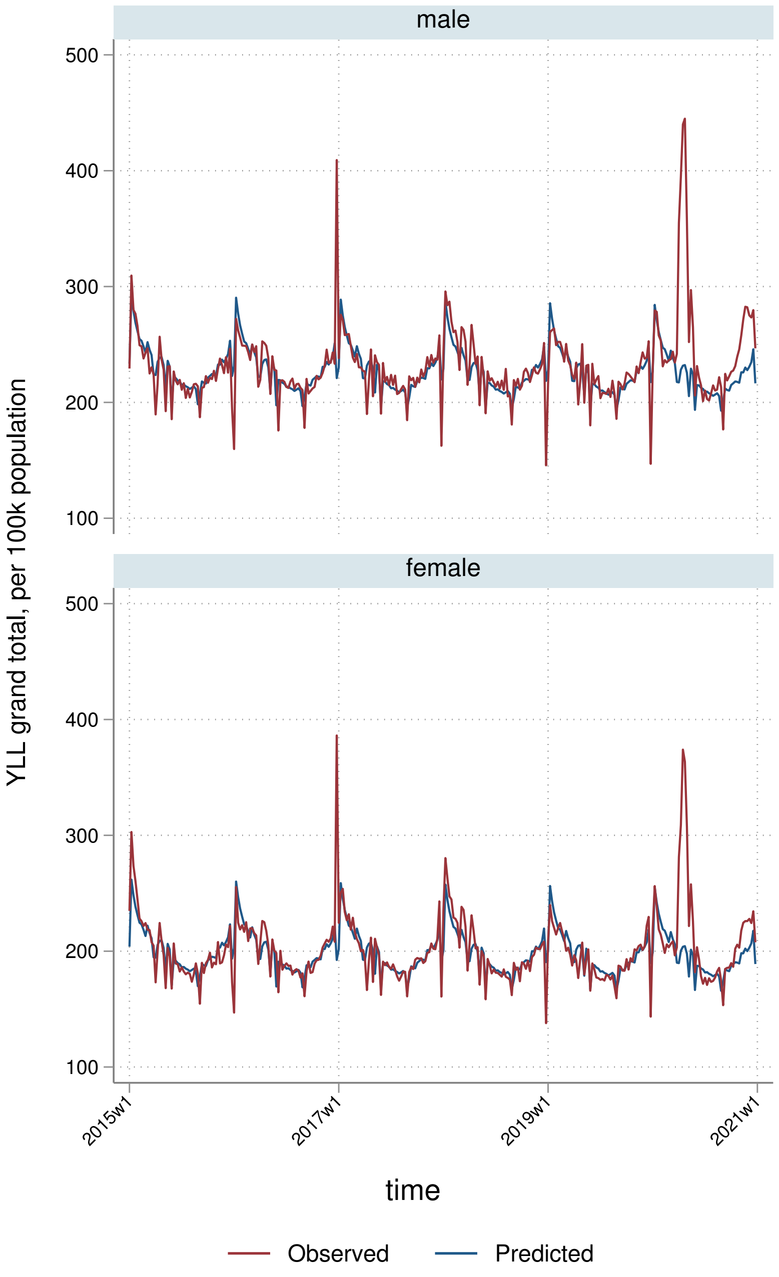

male
female
500
400
300
200
100
YLL grand total, per 100k population
2015w1
2017w1
2019w1
2021w1
time
Observed
Predicted

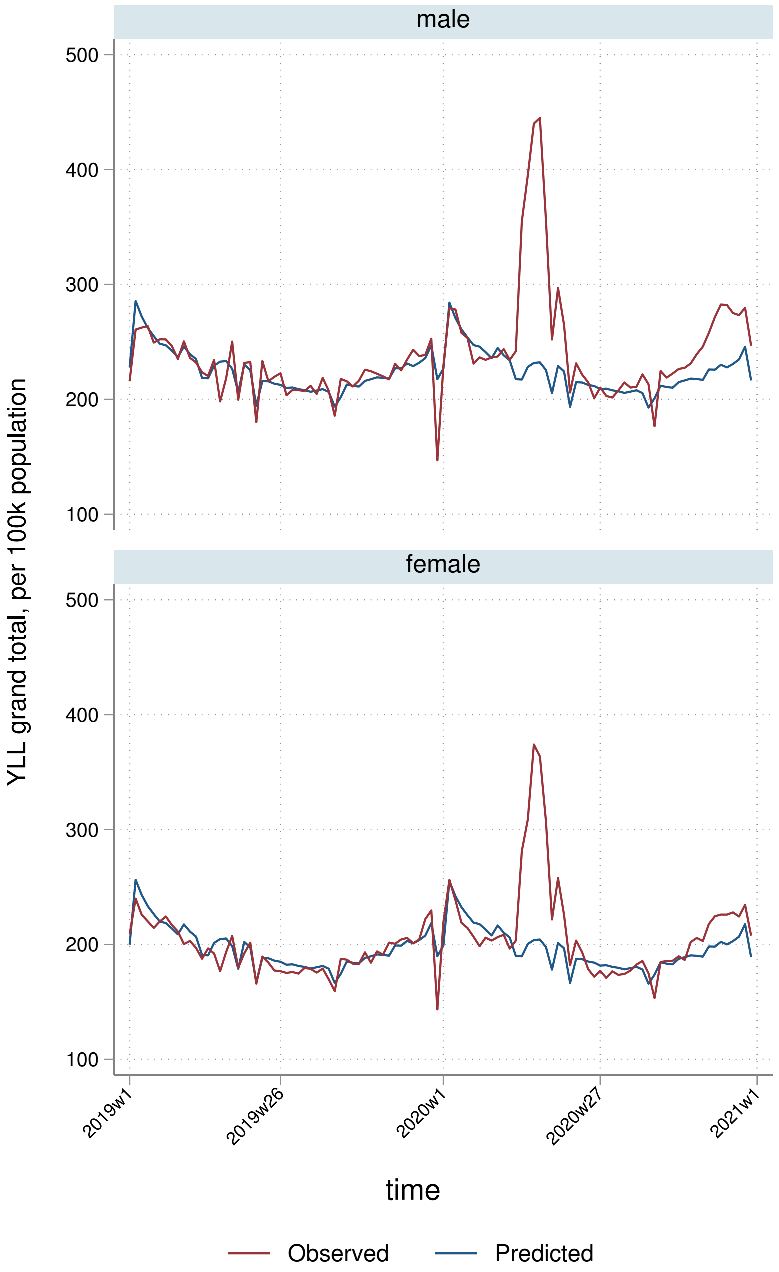
male
female
500
400
300
200
100
YLL grand total, per 100k population
2019w1
2019w26
2020w1
2020w27
2021w1
time
Observed
Predicted

##### 5.3.3 By deprivation quintile

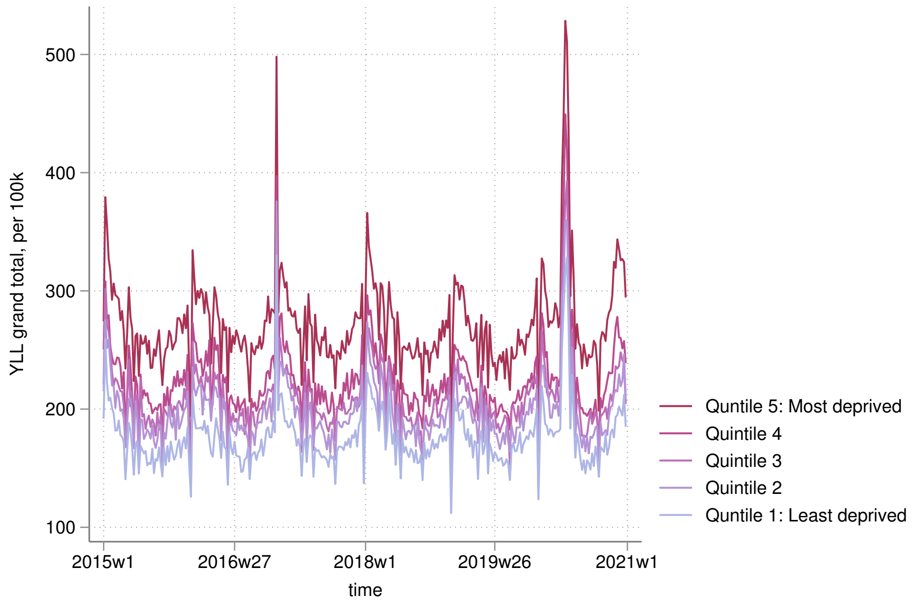


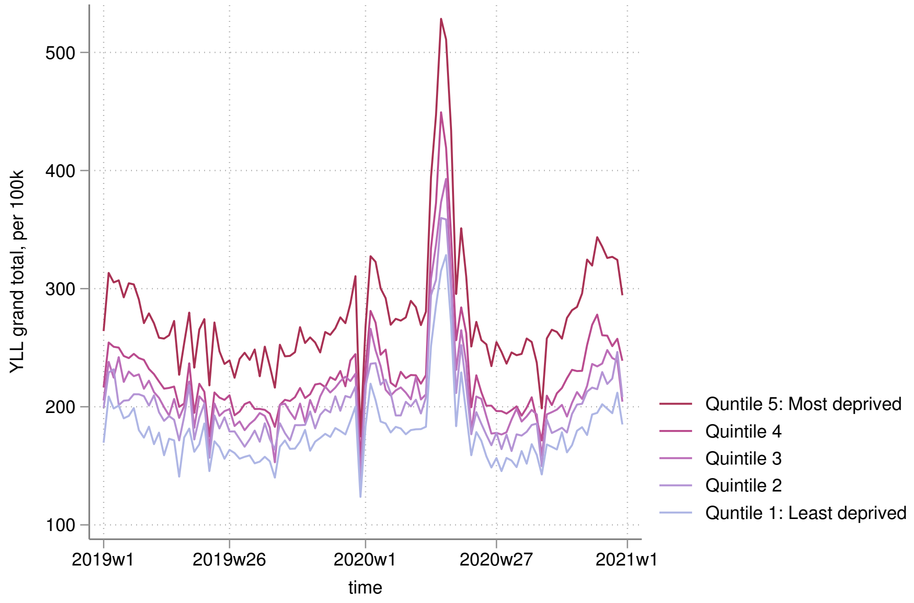

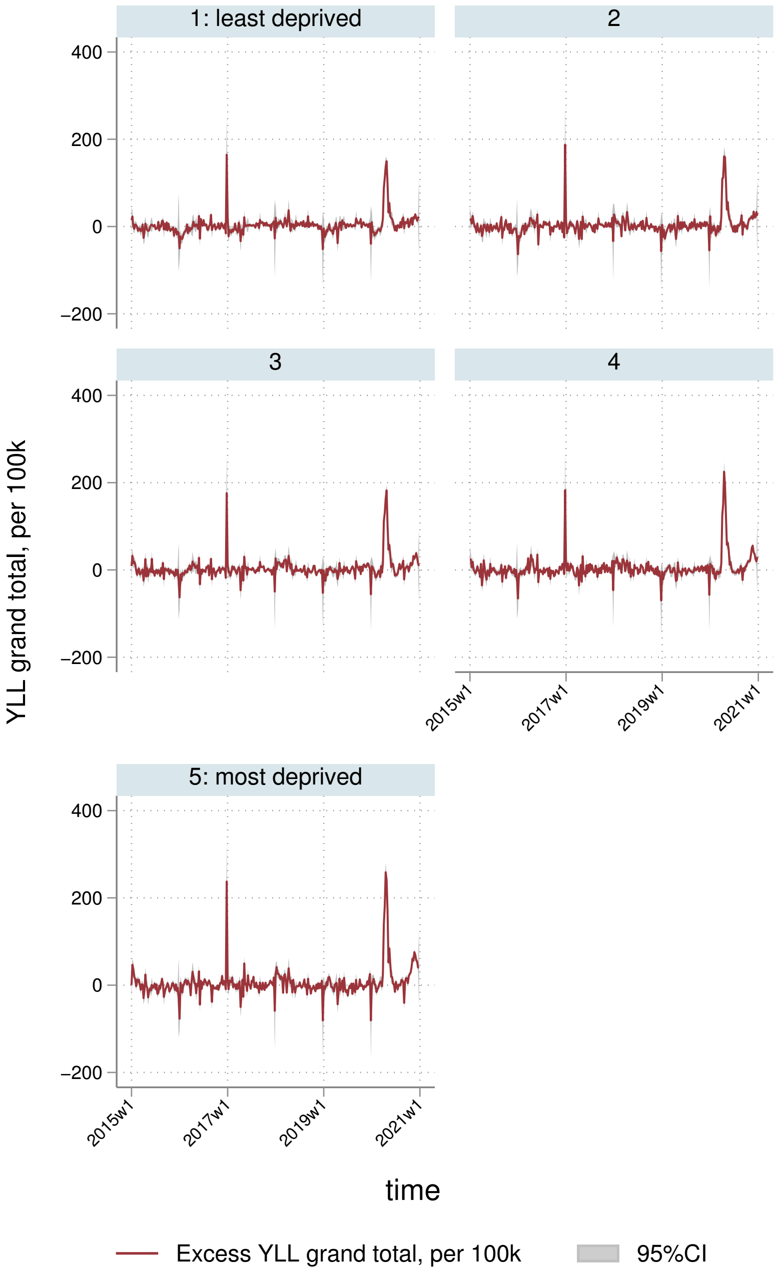
1: least deprived
2
3
4
5: most deprived
400
200
0
−200
YLL grand total, per 100k
2015w1
2017w1
2019w1
2021w1
time
Excess YLL grand total, per 100k
95%CI

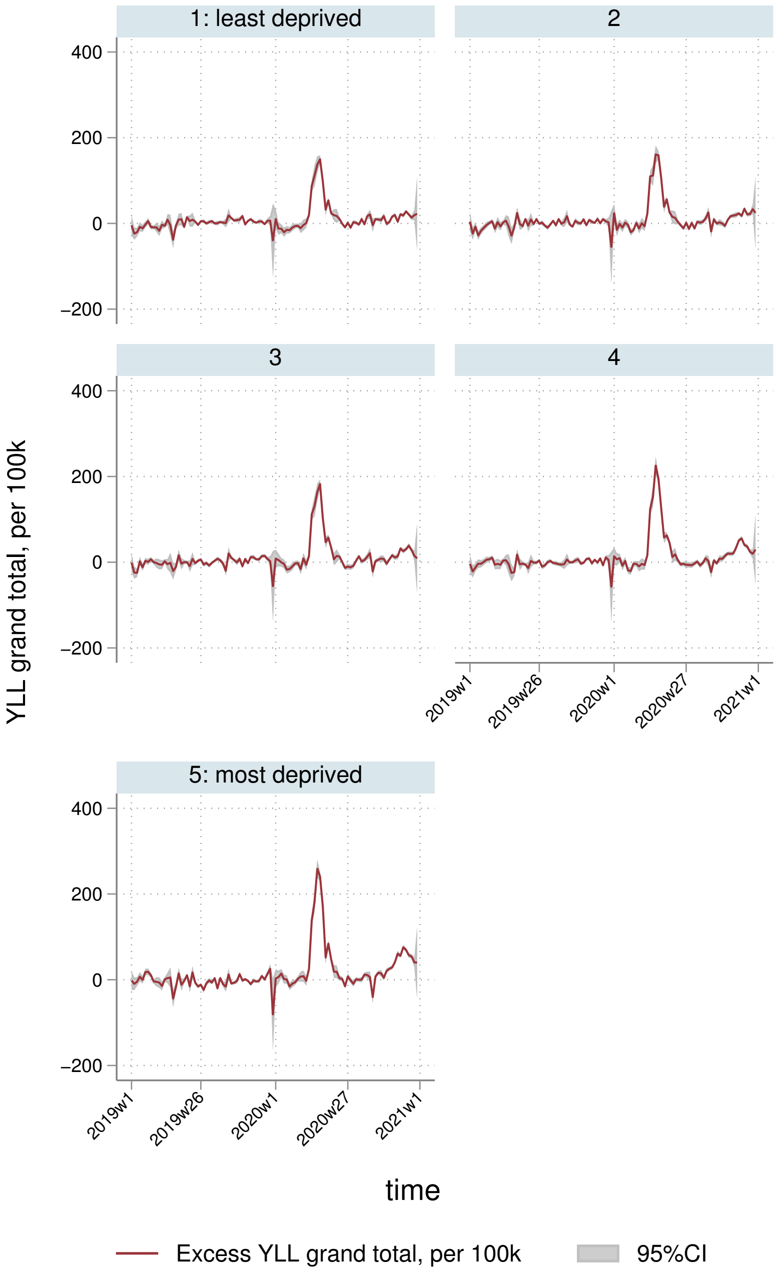
1: least deprived
2
3
4
5: most deprived
400
200
0
−200
YLL grand total, per 100k
2019w1
2019w26
2020w1
2020w27
2021w1
time
Excess YLL grand total, per 100k
95%CI

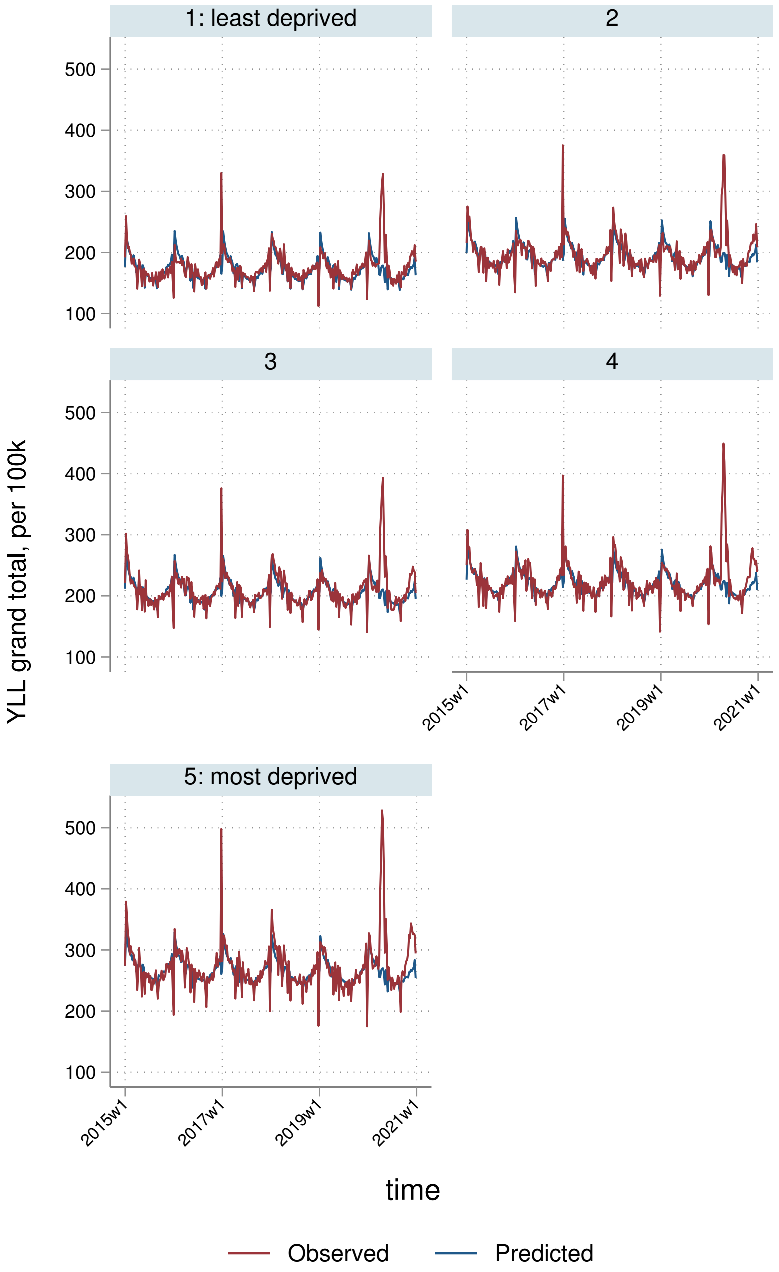
1: least deprived
2
3
4
5: most deprived
YLL grand total, per 100k
100
200
300
400
500
2015w1
2017w1
2019w1
2021w1
time
Observed
Predicted

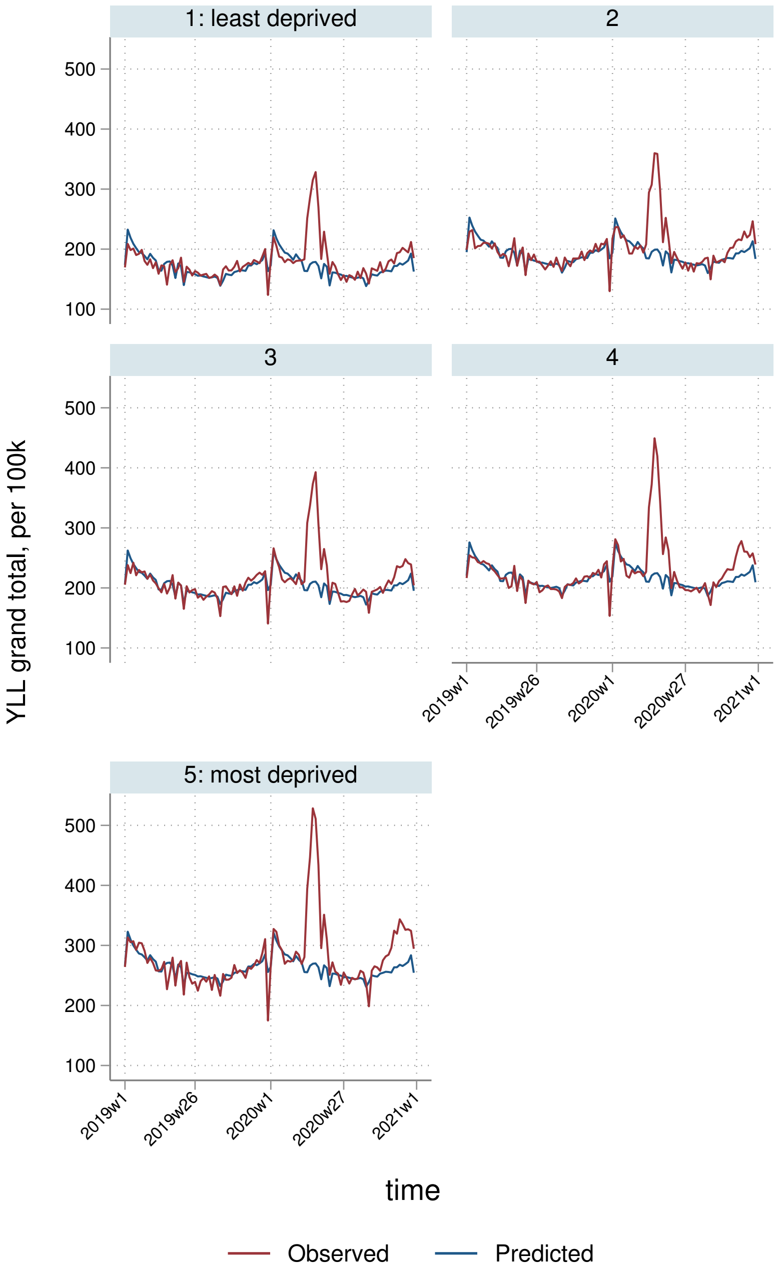
1: least deprived
2
3
4
5: most deprived
YLL grand total, per 100k
500
400
300
200
100
2019w1
2019w26
2020w1
2020w27
2021w1
time
Observed
Predicted

##### 5.3.4 By Strategic Health Authority

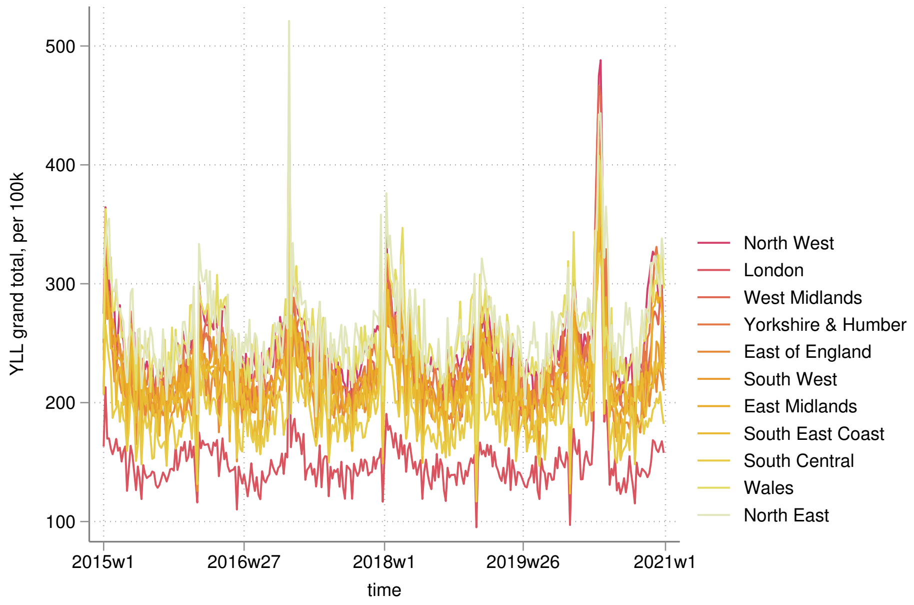


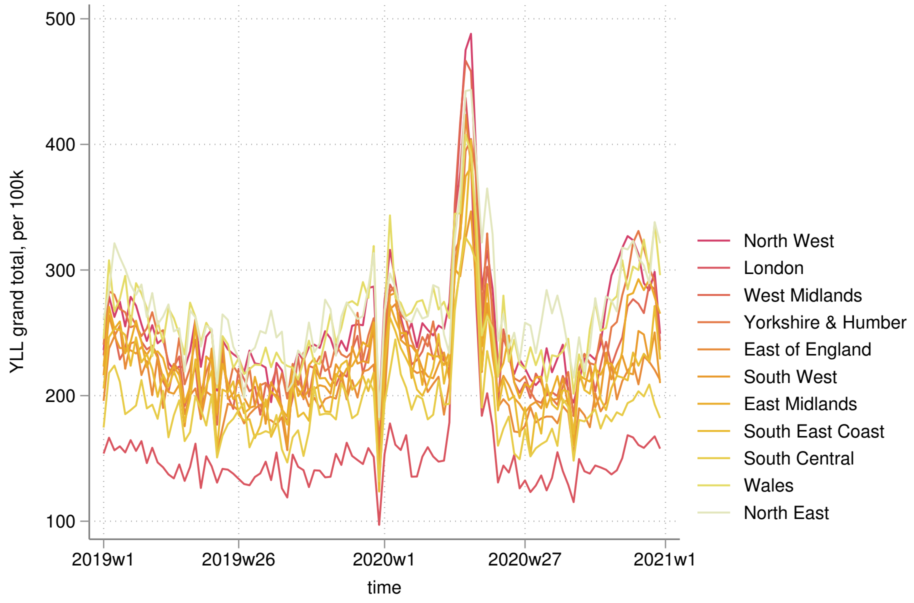

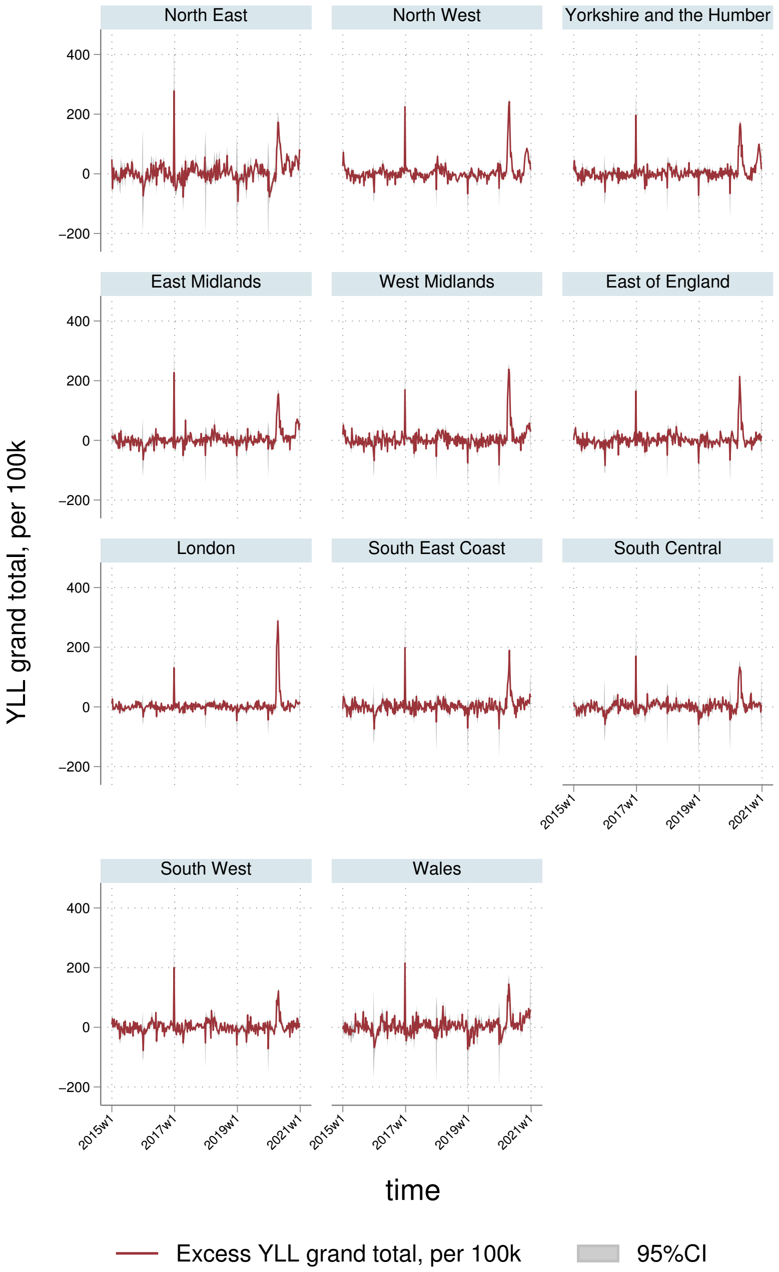
North East
North West
Yorkshire and the Humber
East Midlands
West Midlands
East of England
London
South East Coast
South Central
South West
Wales
400
200
0
−200
2015w1
2017w1
2019w1
2021w1
YLL grand total, per 100k
time
Excess YLL grand total, per 100k
95%CI

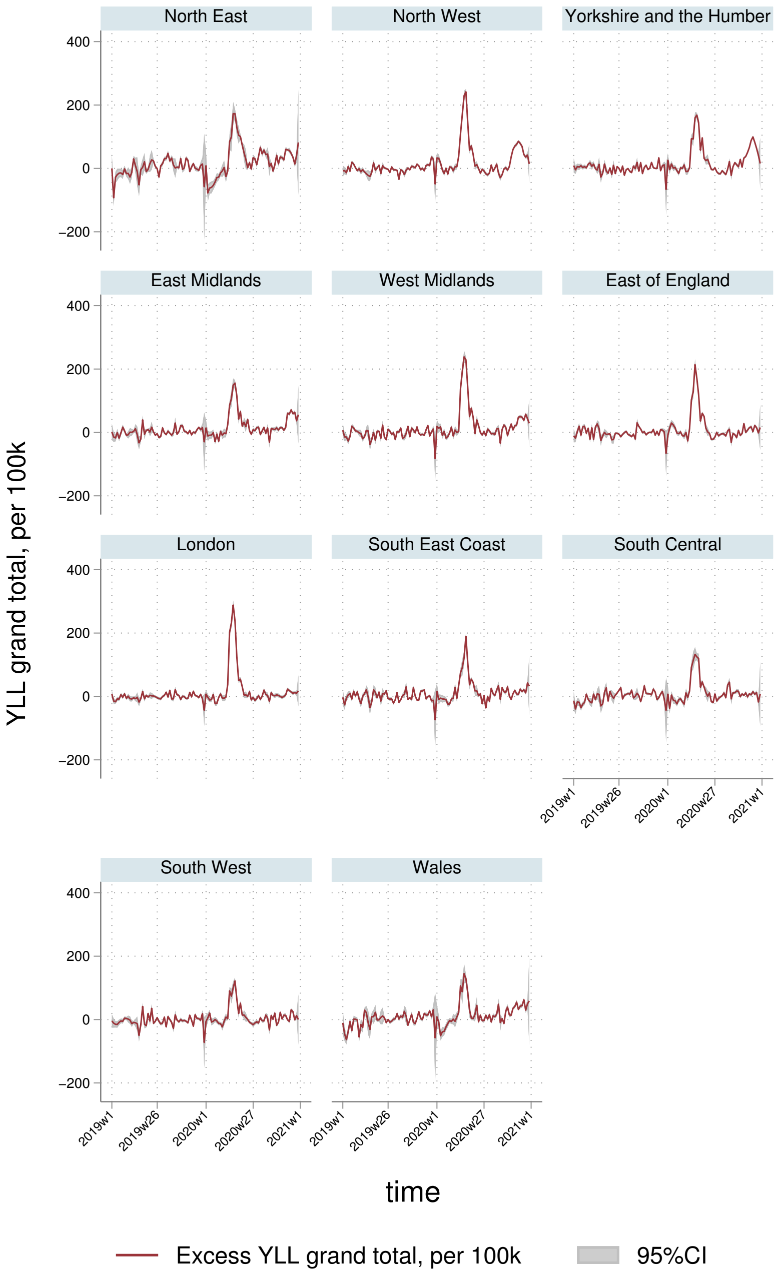

North East
North West
Yorkshire and the Humber
East Midlands
West Midlands
East of England
London
South East Coast
South Central
South West
Wales
400
200
0
−200
YLL grand total, per 100k
2019w1
2019w26
2020w1
2020w27
2021w1
time
Excess YLL grand total, per 100k
95%CI

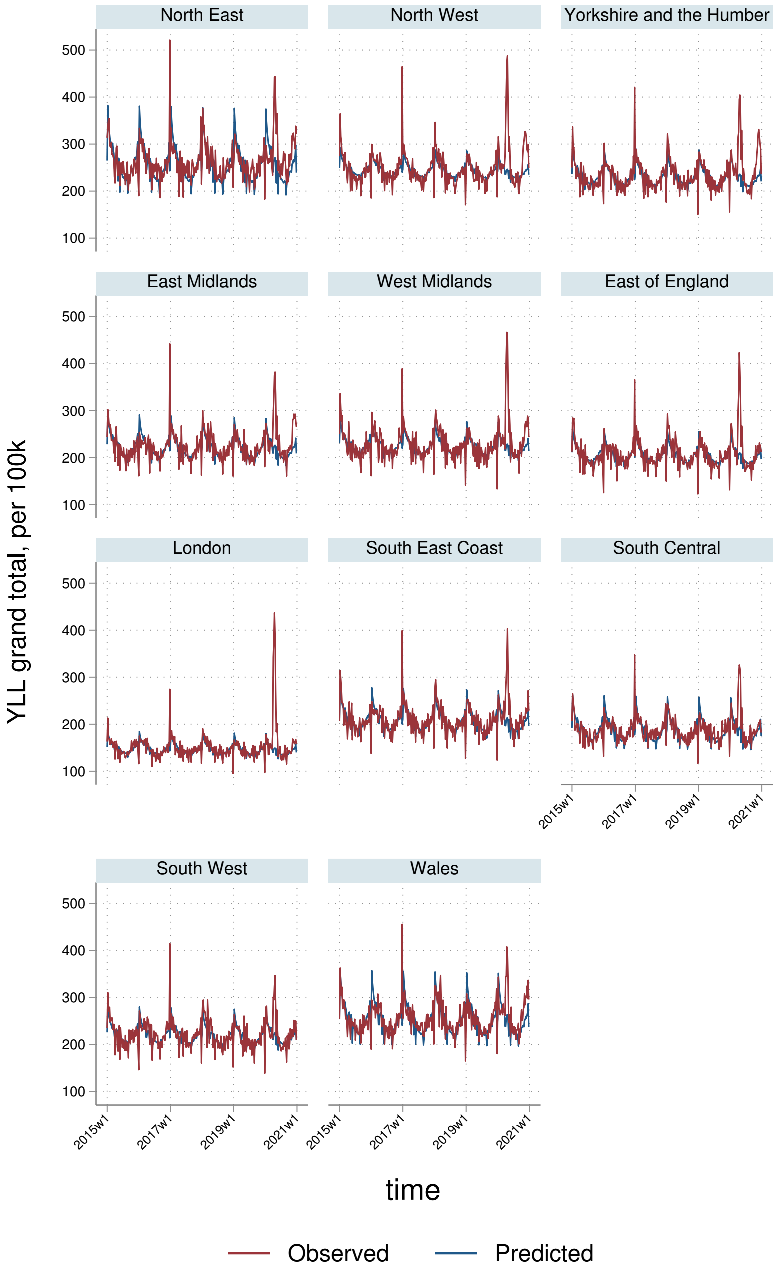
North East
North West
Yorkshire and the Humber
East Midlands
West Midlands
East of England
London
South East Coast
South Central
South West
Wales
YLL grand total, per 100k
100
200
300
400
500
2015w1
2017w1
2019w1
2021w1
time
Observed
Predicted

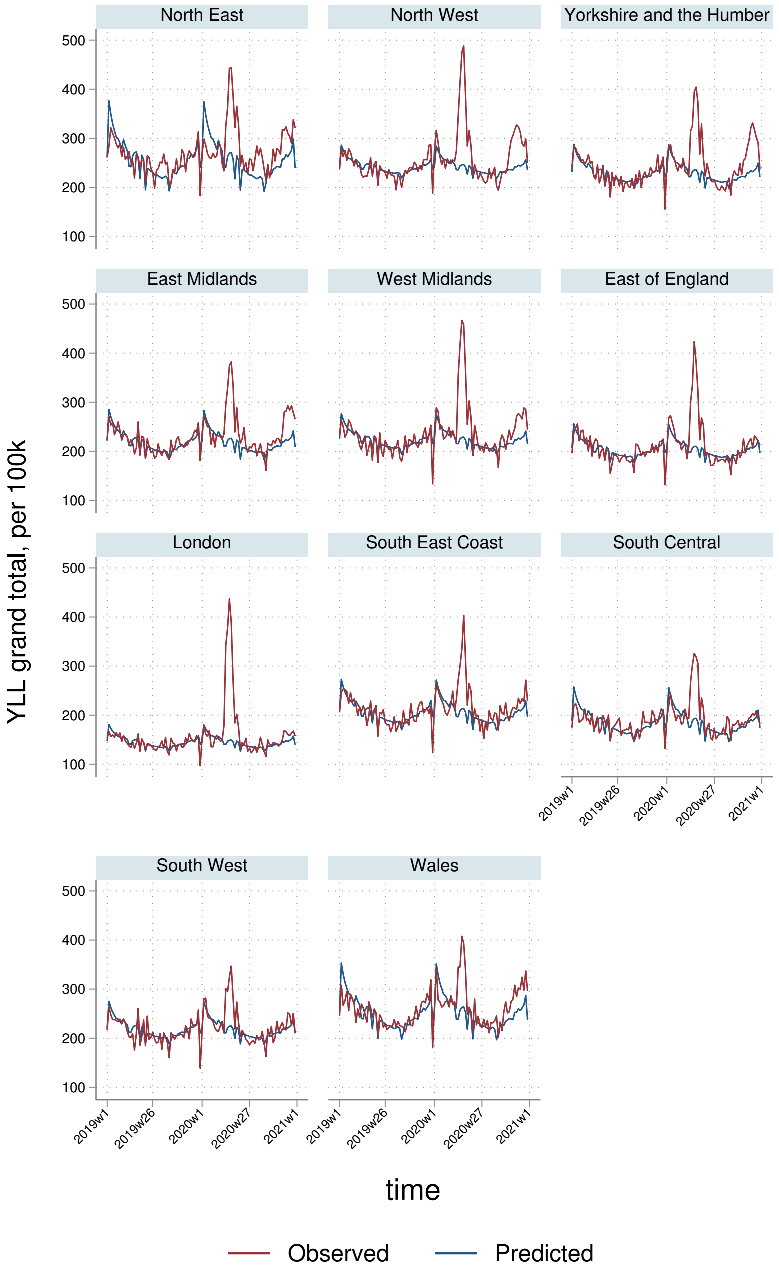
North East
North West
Yorkshire and the Humber
East Midlands
West Midlands
East of England
London
South East Coast
South Central
South West
Wales
500
400
300
200
100
2019w1
2019w26
2020w1
2020w27
2021w1
YLL grand total, per 100k
time
Observed
Predicted

### 6 Direct

#### 6.1 AASMRs

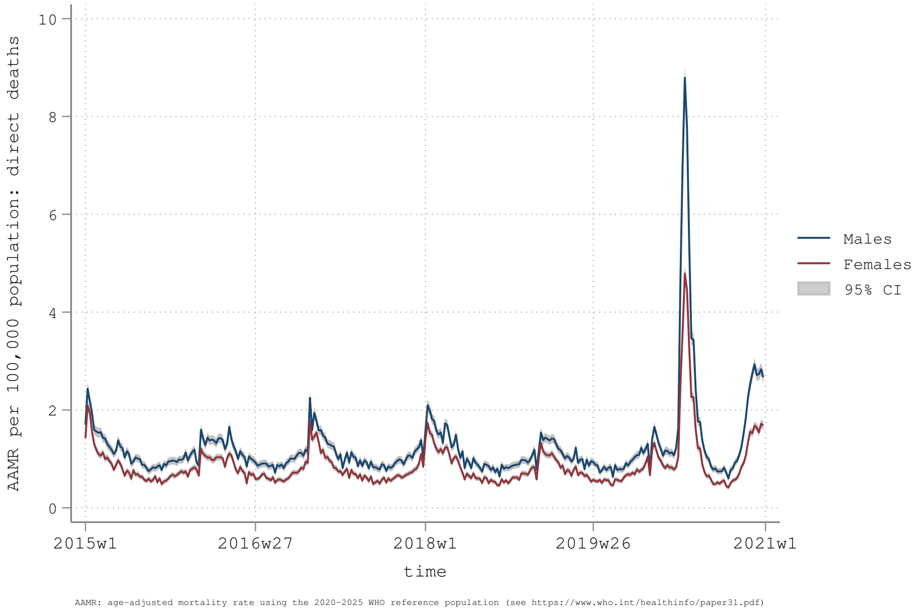


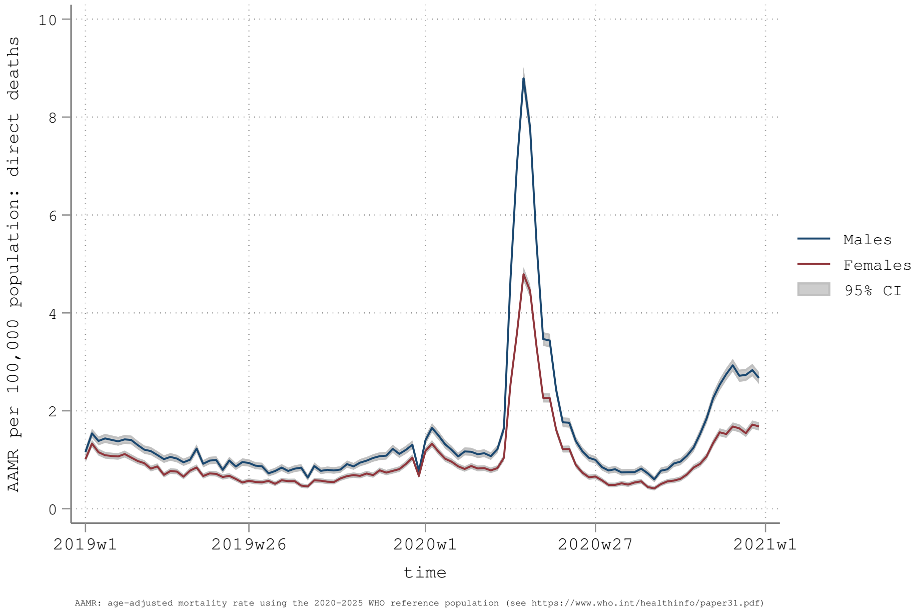

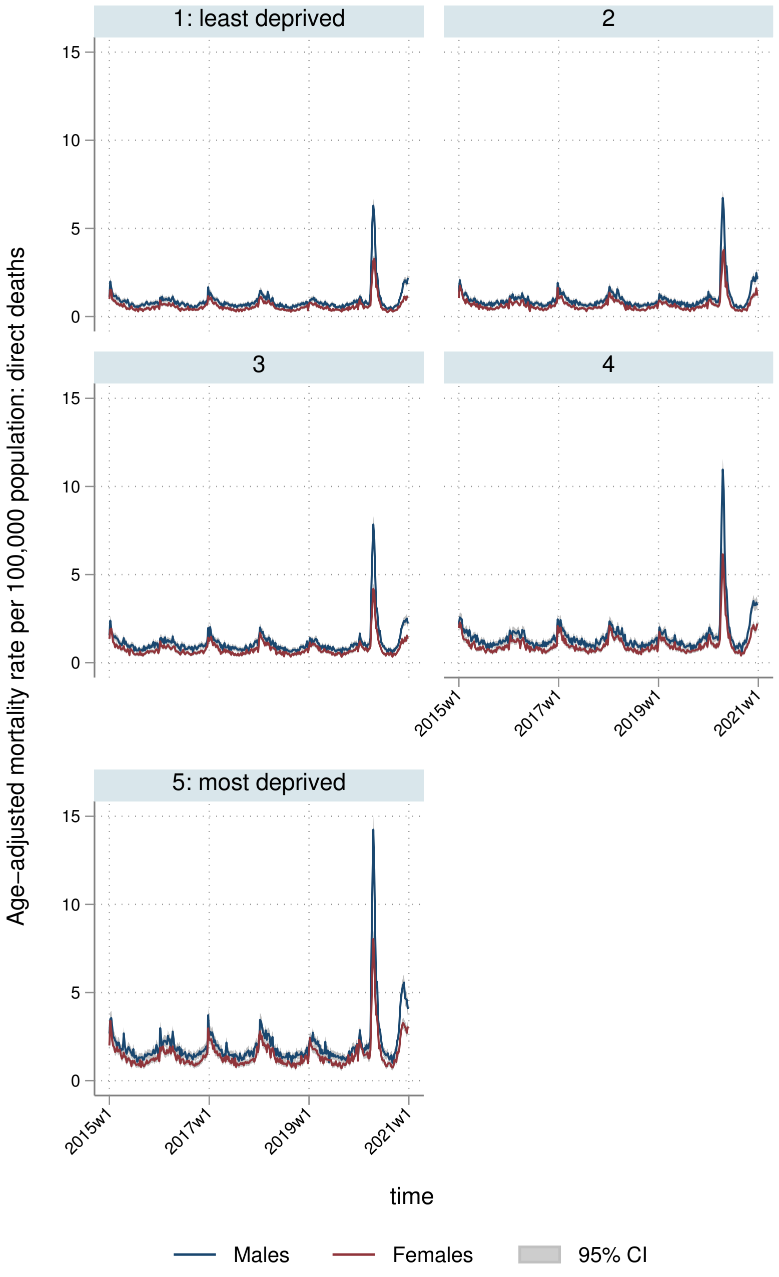
1: least deprived
2
3
4
5: most deprived
Age-adjusted mortality rate per 100,000 population: direct deaths
15
10
5
0
2015w1
2017w1
2019w1
2021w1
time
Males
Females
95% CI

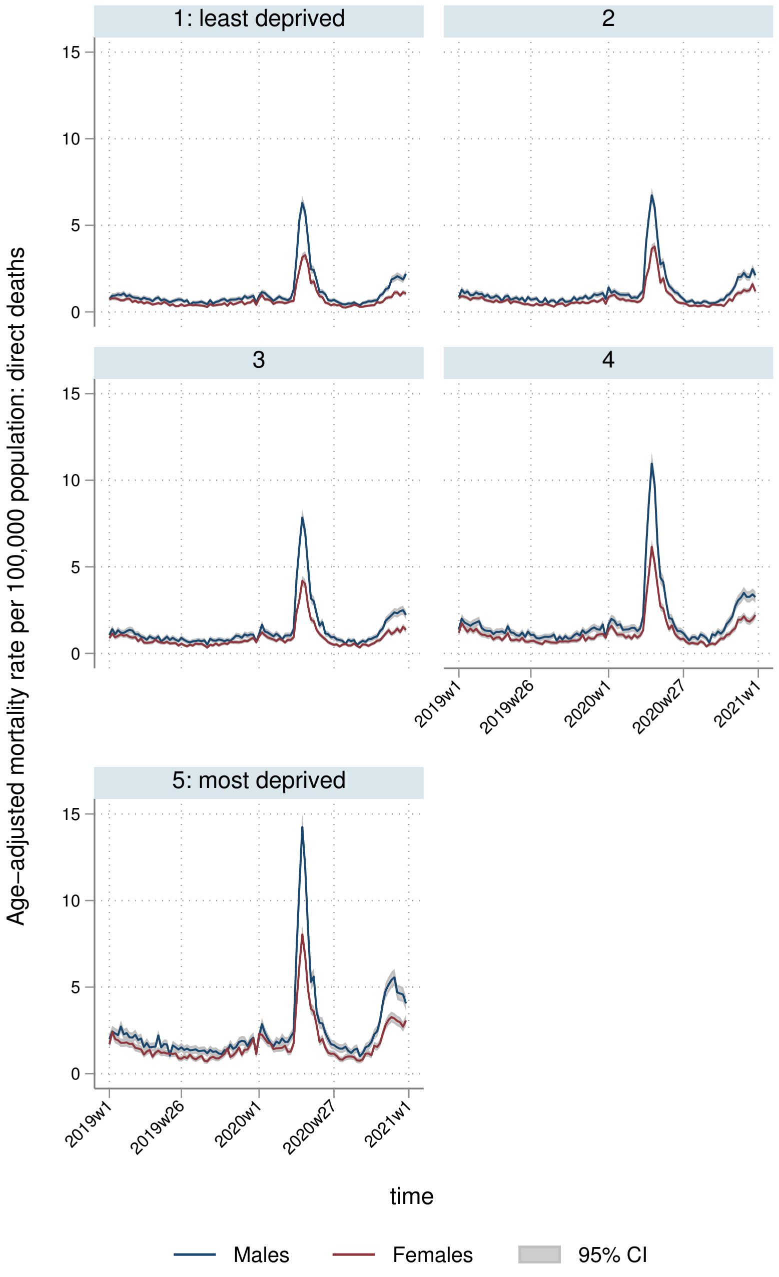
1: least deprived
2
3
4
5: most deprived
Age-adjusted mortality rate per 100,000 population: direct deaths
15
10
5
0
2019w1
2019w26
2020w1
2020w27
2021w1
time
Males
Females
95% CI

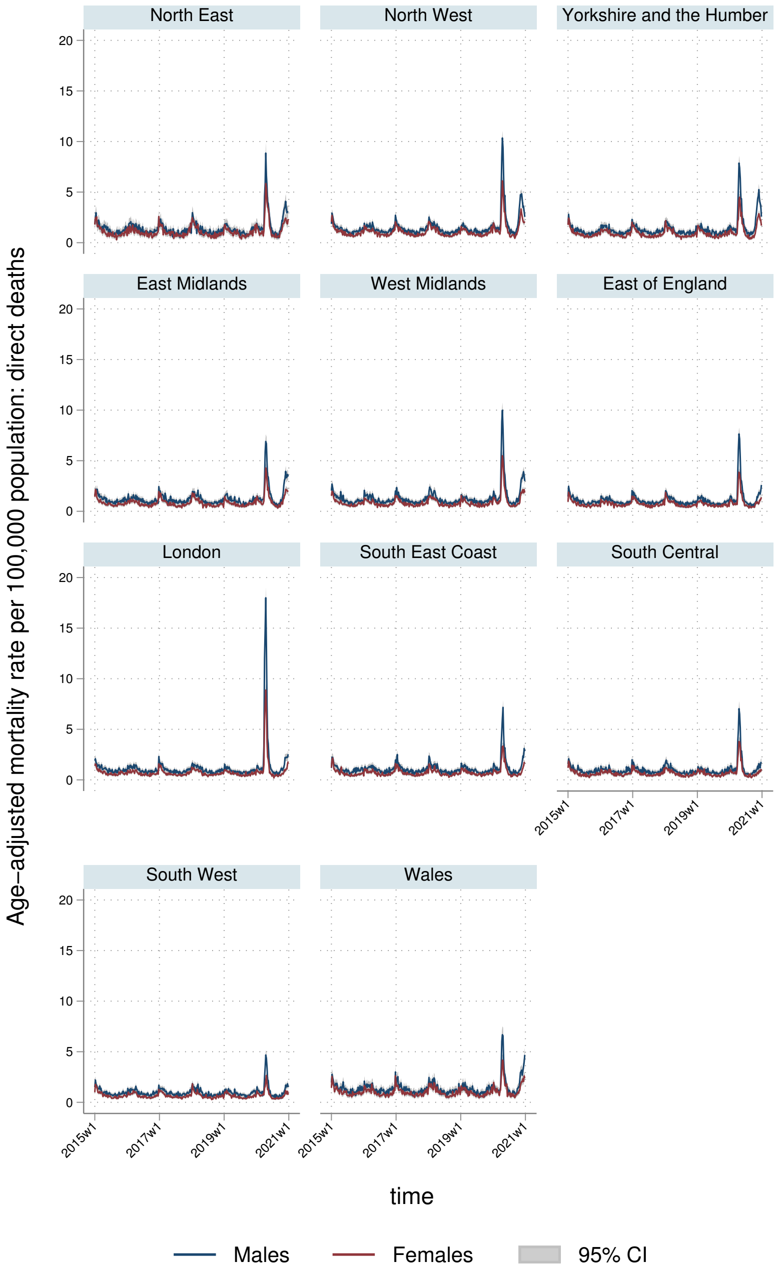
North East
North West
Yorkshire and the Humber
East Midlands
West Midlands
East of England
London
South East Coast
South Central
South West
Wales
0
5
10
15
20
2015w1
2017w1
2019w1
2021w1
Age–adjusted mortality rate per 100,000 population: direct deaths
time
Males
Females
95% CI

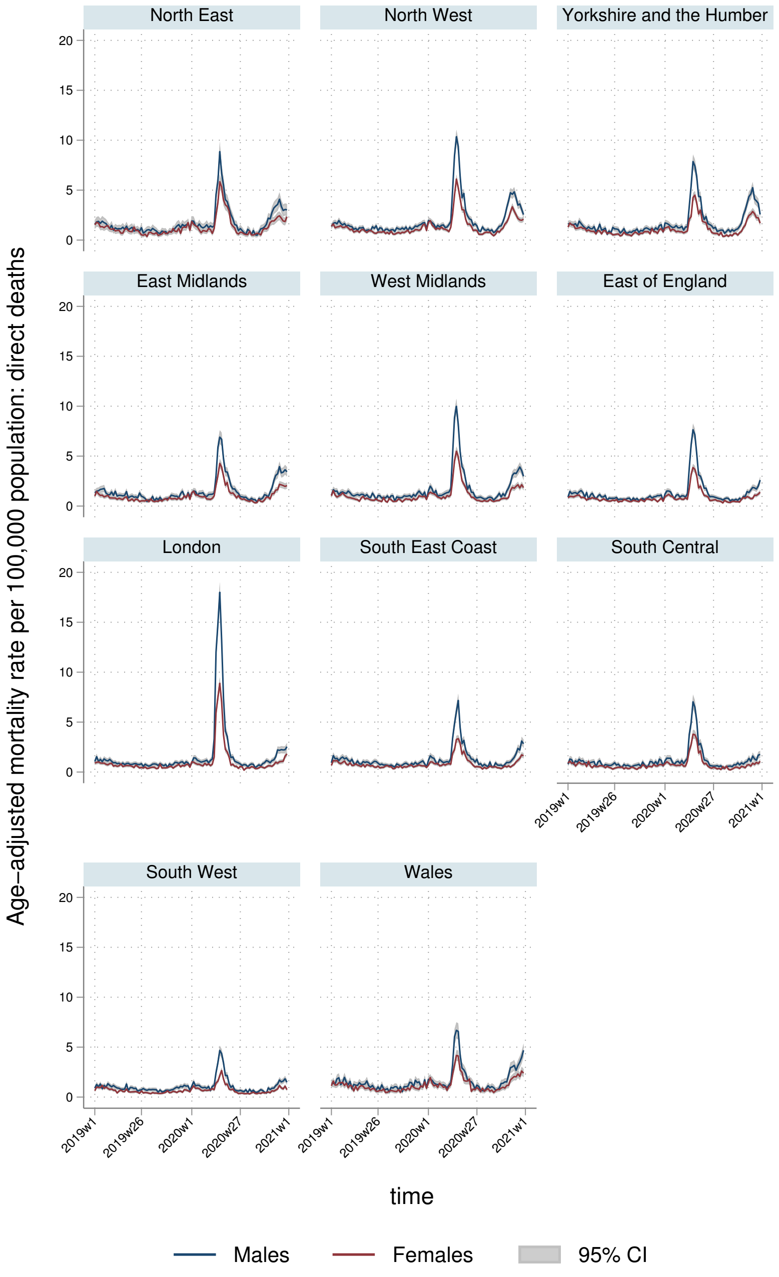

North East
North West
Yorkshire and the Humber
East Midlands
West Midlands
East of England
London
South East Coast
South Central
South West
Wales
Age-adjusted mortality rate per 100,000 population: direct deaths
0
5
10
15
20
2019w1
2019w26
2020w1
2020w27
2021w1
time
Males
Females
95% CI

#### 6.2 YLLs

##### 6.2.1 England-Wales aggregate

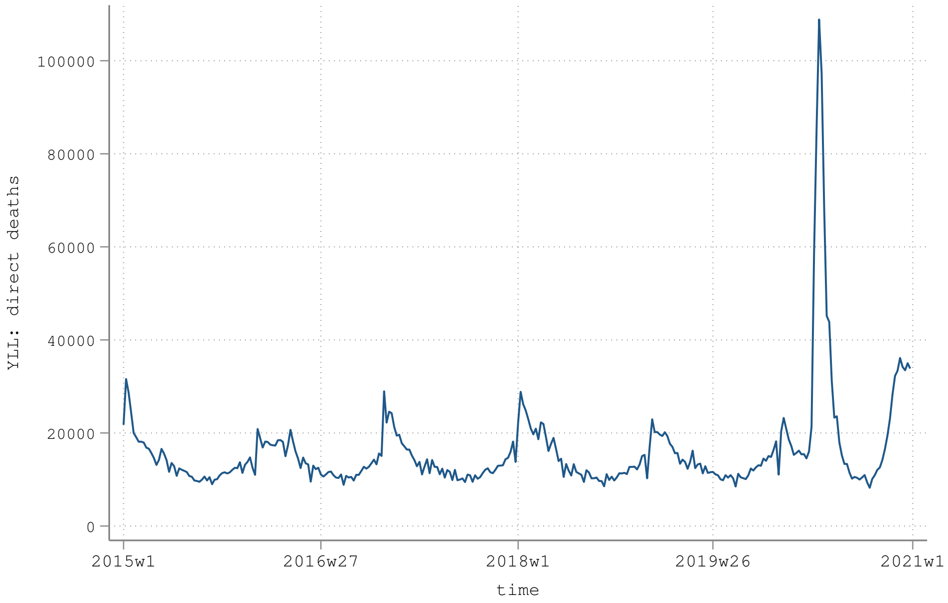


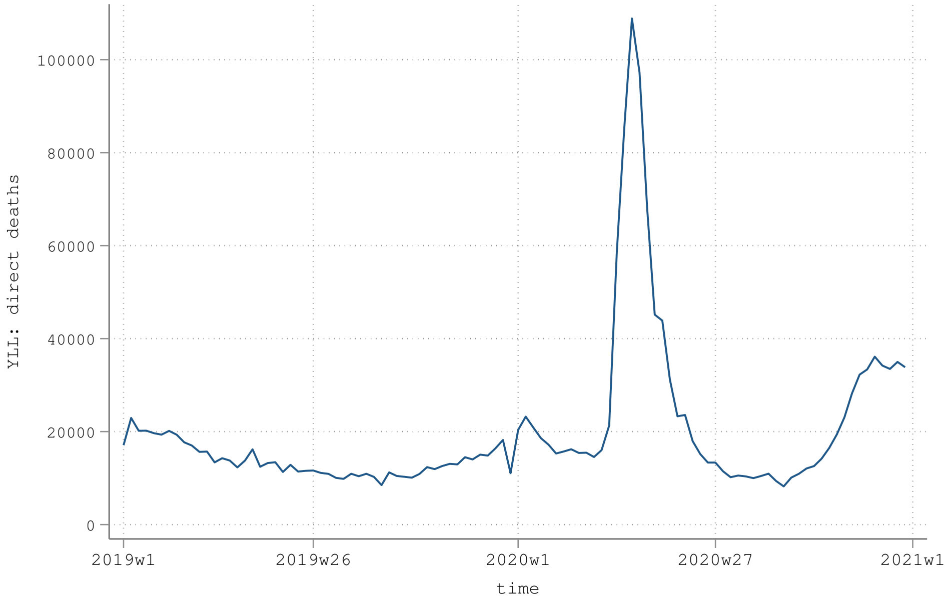

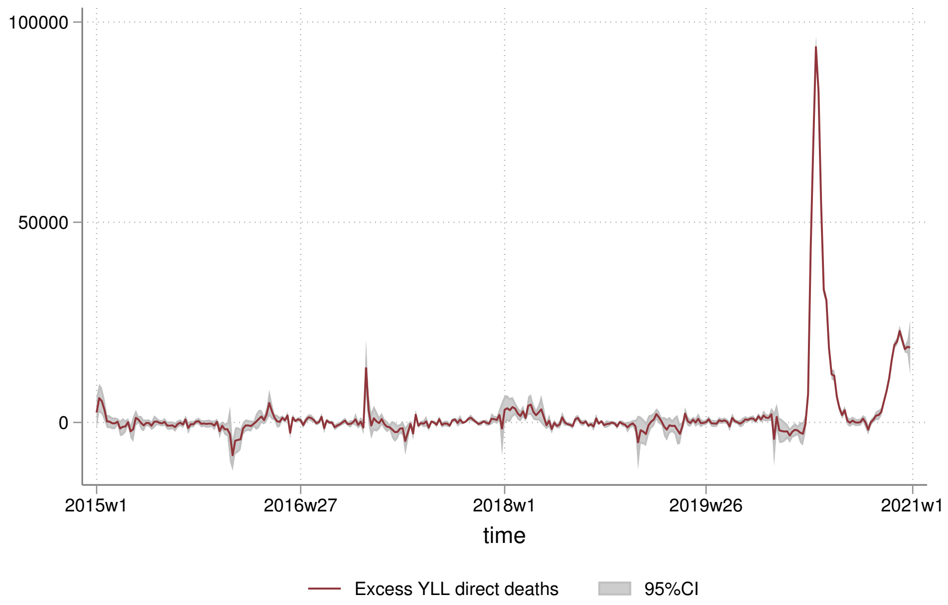
100000
50000
0
2015w1
2016w27
2018w1
2019w26
2021w1
time
Excess YLL direct deaths
95%CI

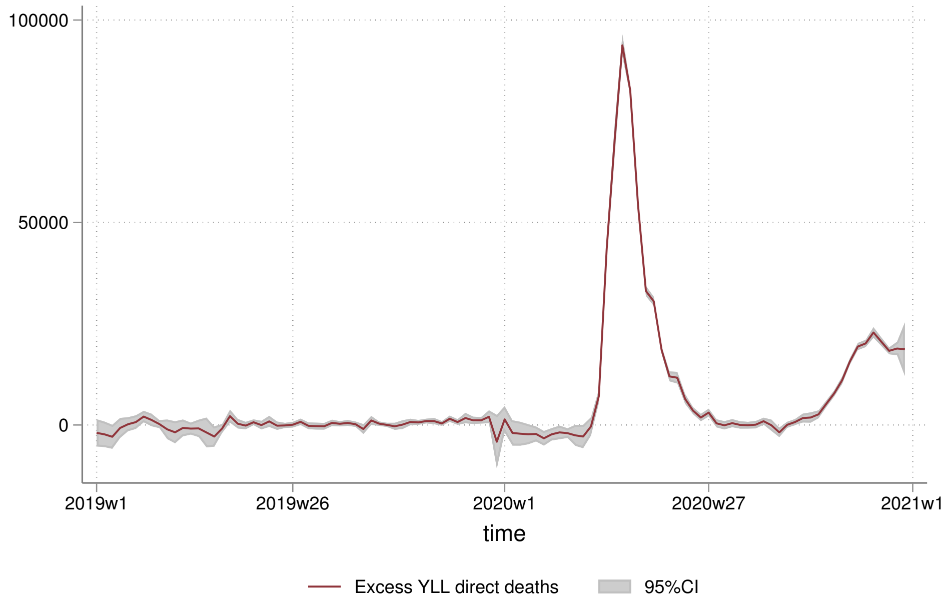
100000
50000
0
2019w1
2019w26
2020w1
2020w27
2021w1
time
Excess YLL direct deaths
95%CI

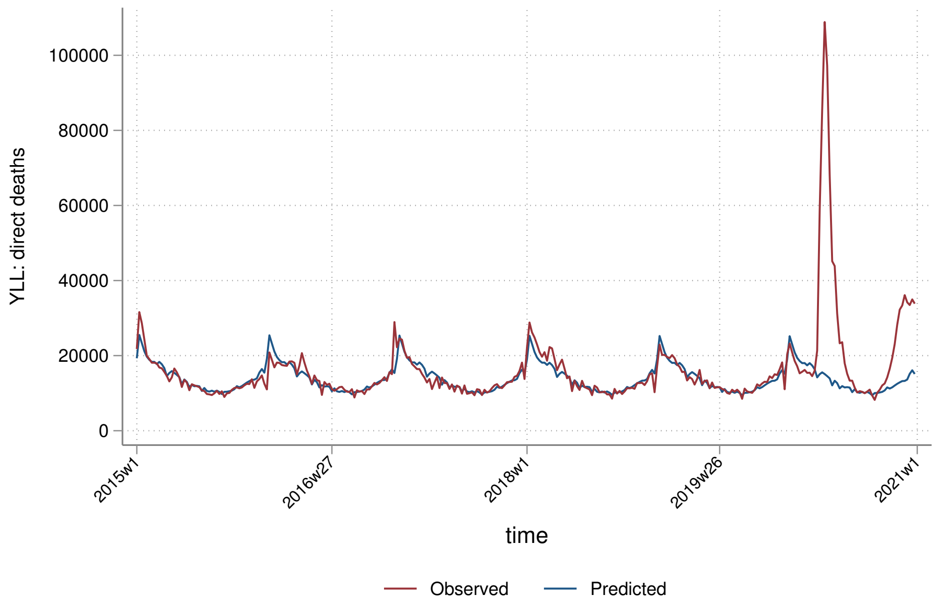
YLL: direct deaths
100000
80000
60000
40000
20000
0
2015w1
2016w27
2018w1
2019w26
2021w1
time
Observed
Predicted

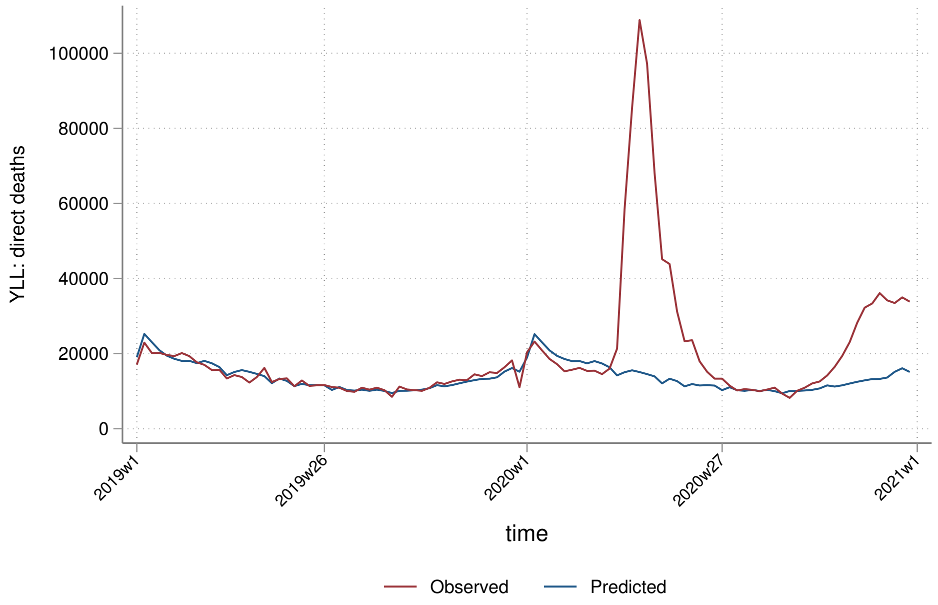
YLL: direct deaths
100000
80000
60000
40000
20000
0
2019w1
2019w26
2020w1
2020w27
2021w1
time
Observed
Predicted

##### 6.2.2 By sex

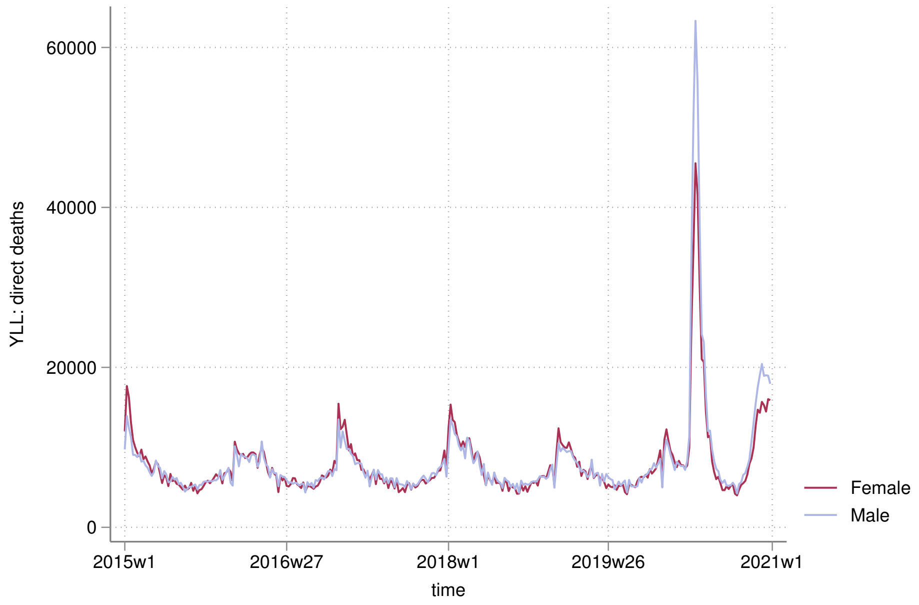

60000
40000
20000
0
YLL: direct deaths
2015w1
2016w27
2018w1
2019w26
2021w1
time
Female
Male


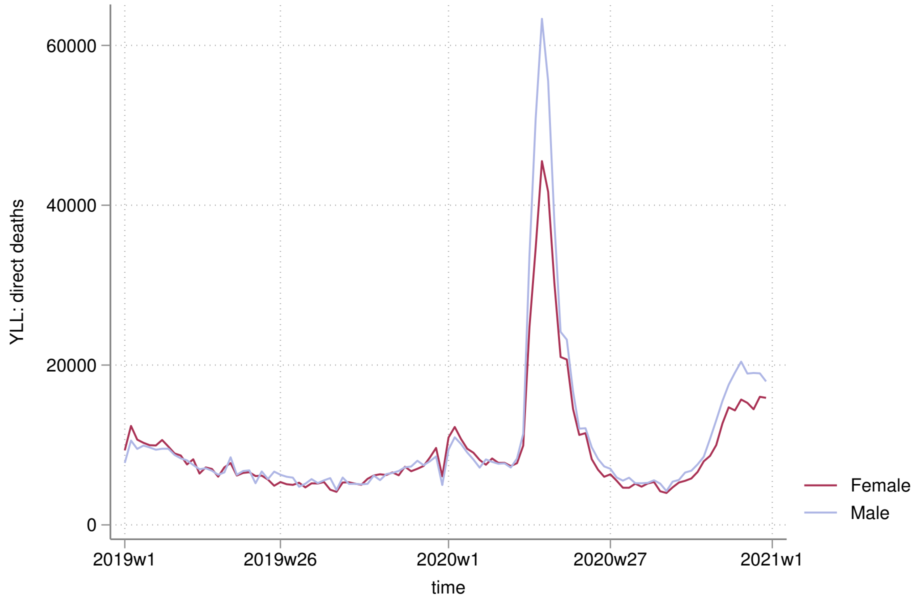

60000
40000
20000
0
YLL: direct deaths
2019w1
2019w26
2020w1
2020w27
2021w1
time
Female
Male

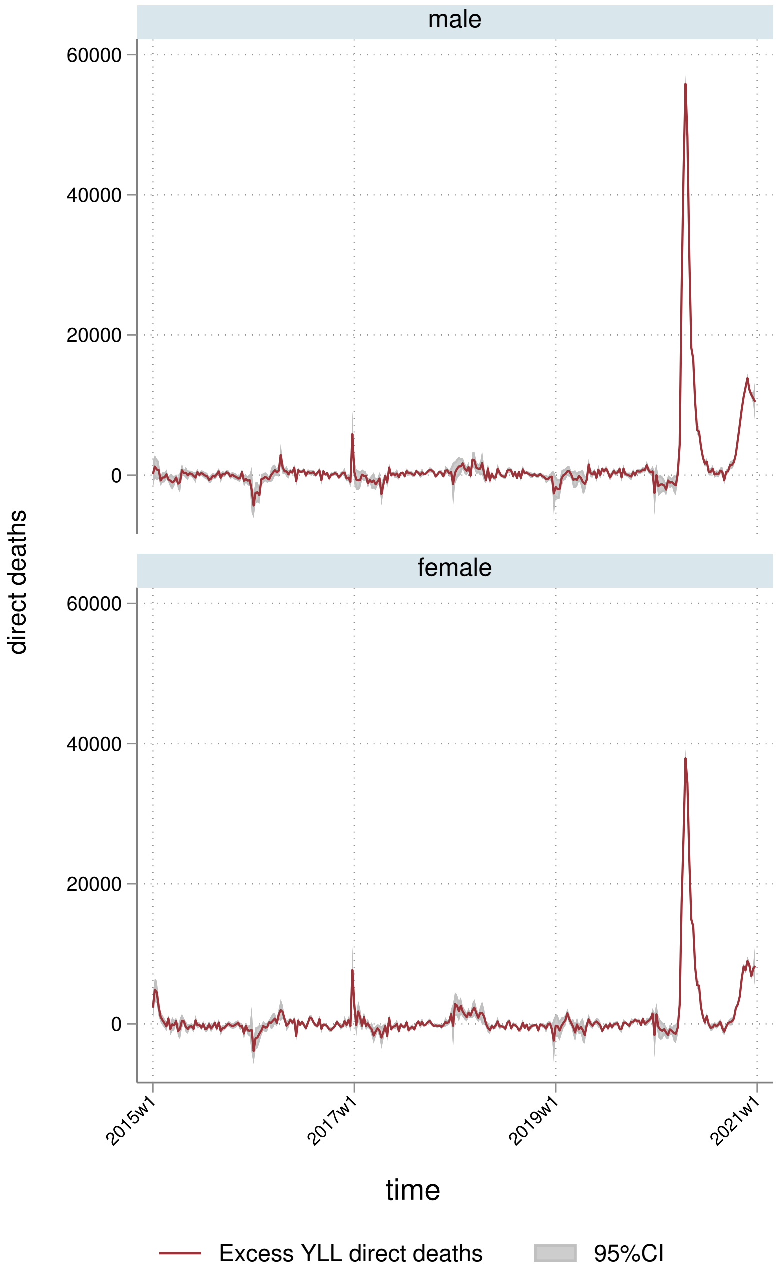
male
female
60000
40000
20000
0
direct deaths
2015w1
2017w1
2019w1
2021w1
time
Excess YLL direct deaths
95%CI

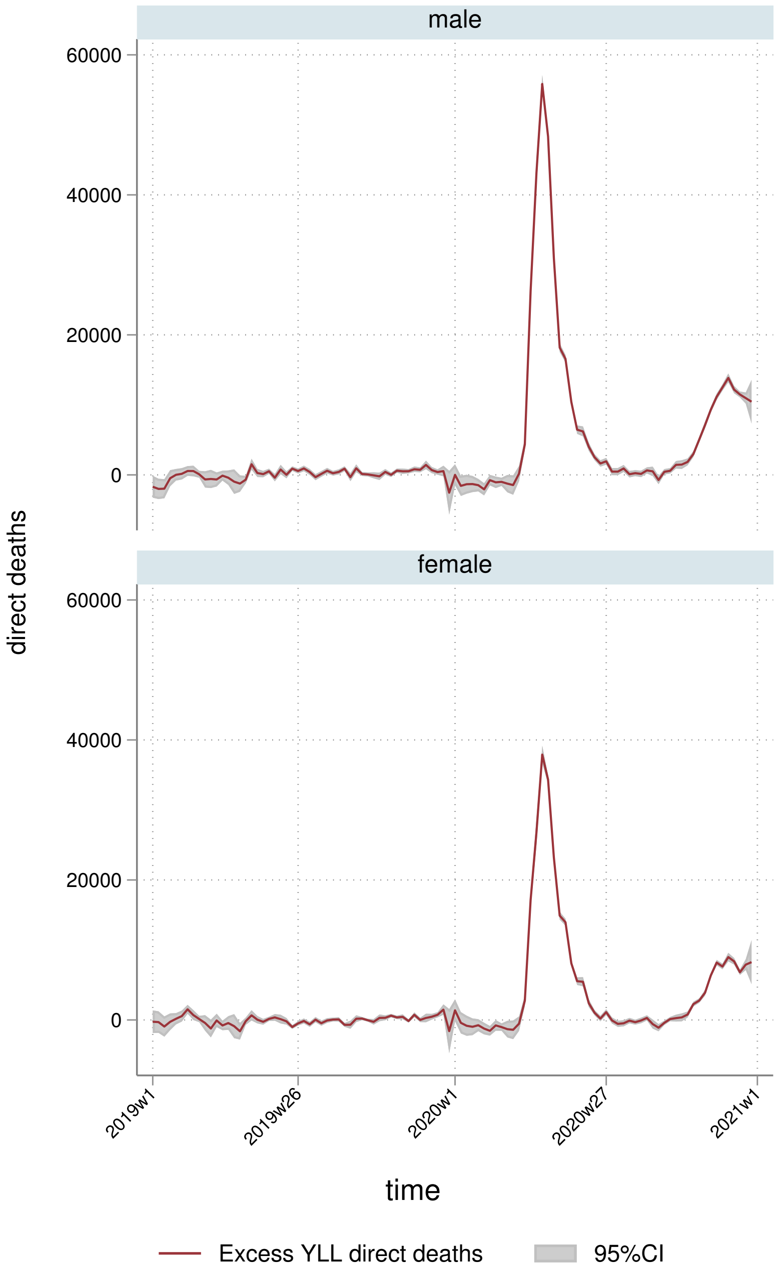
male
female
60000
40000
20000
0
direct deaths
2019w1
2019w26
2020w1
2020w27
2021w1
time
Excess YLL direct deaths
95%CI

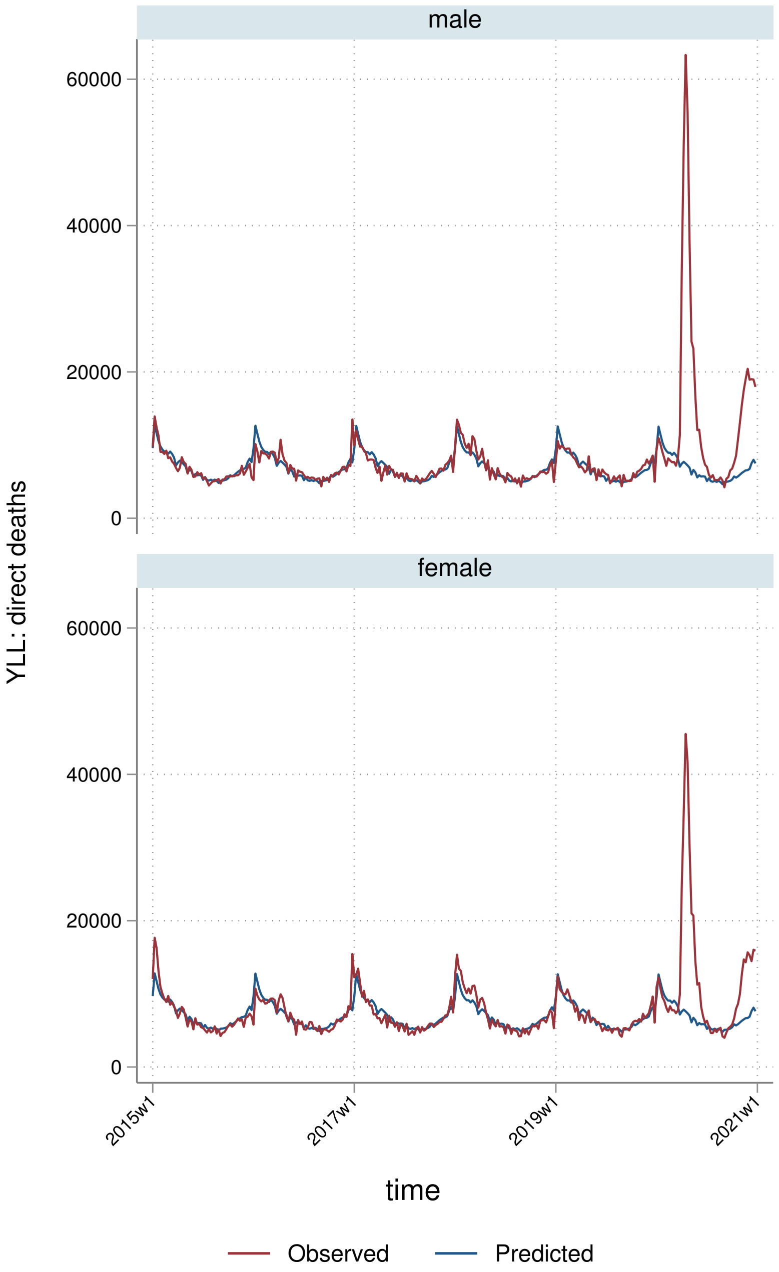
male
female
60000
40000
20000
0
YLL: direct deaths
2015w1
2017w1
2019w1
2021w1
time
Observed
Predicted

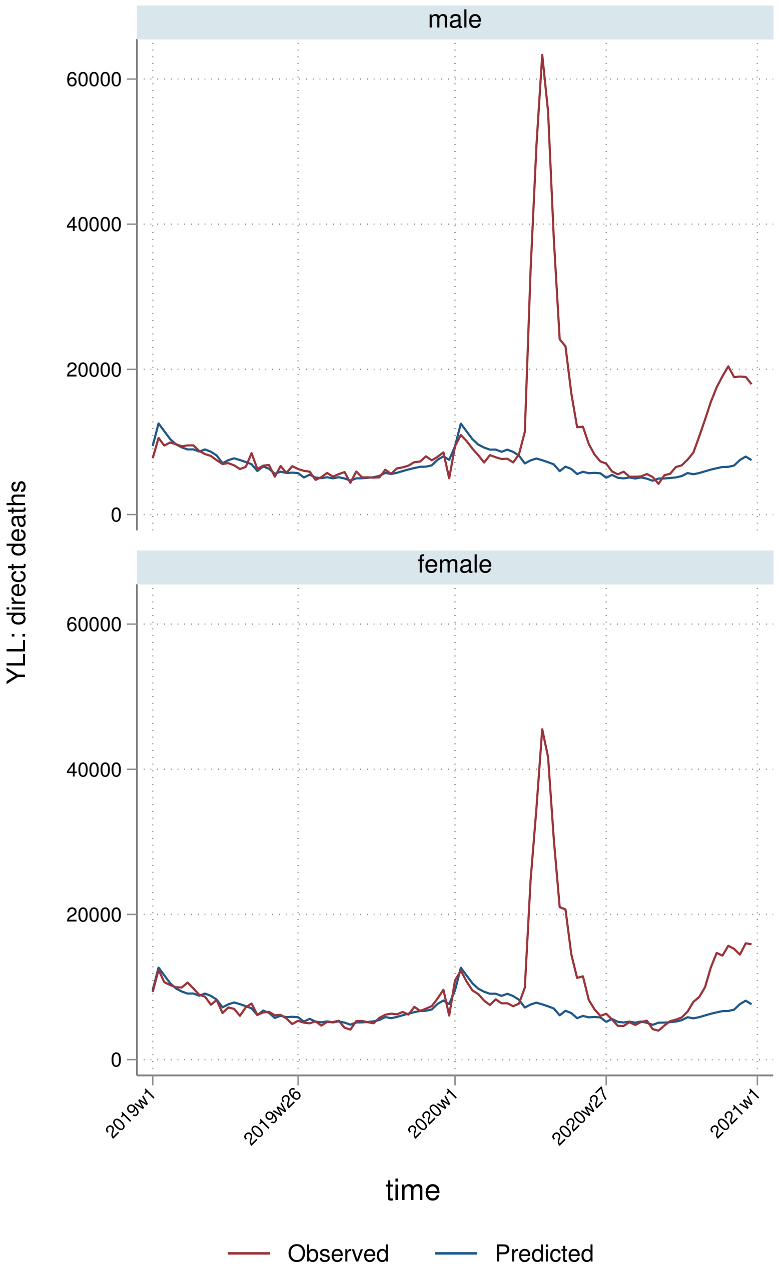
male
female
60000
40000
20000
0
YLL: direct deaths
2019w1
2019w26
2020w1
2020w27
2021w1
time
Observed
Predicted

##### 6.2.3 By deprivation quintile

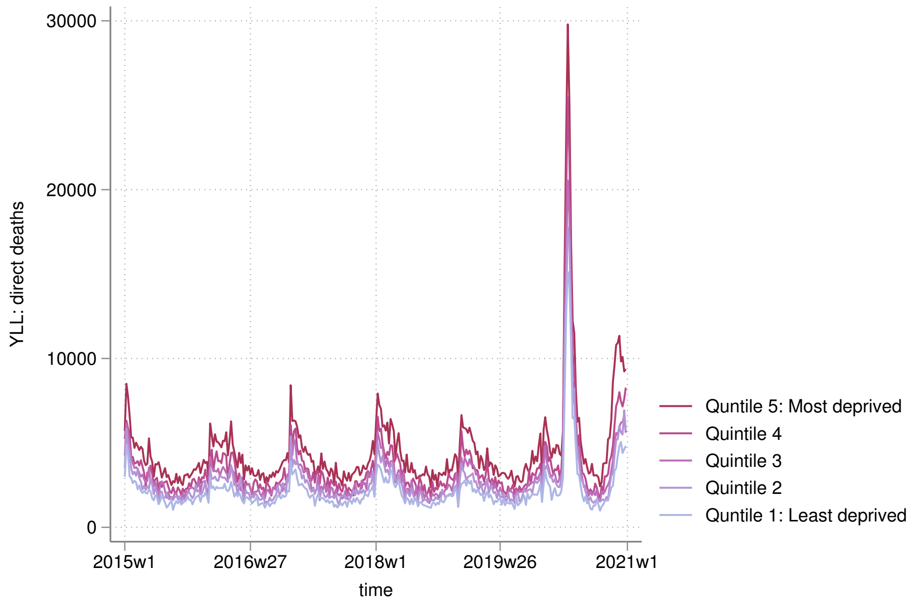


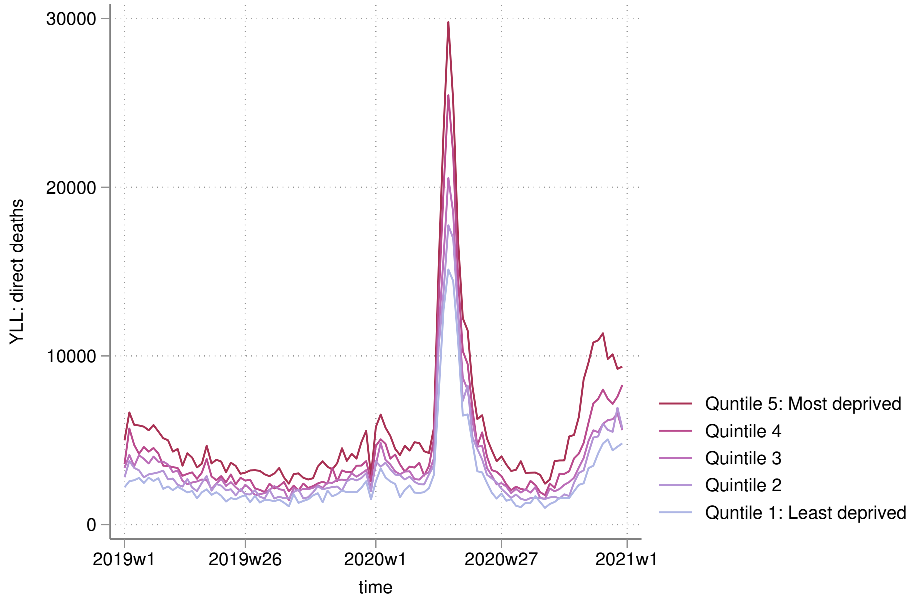

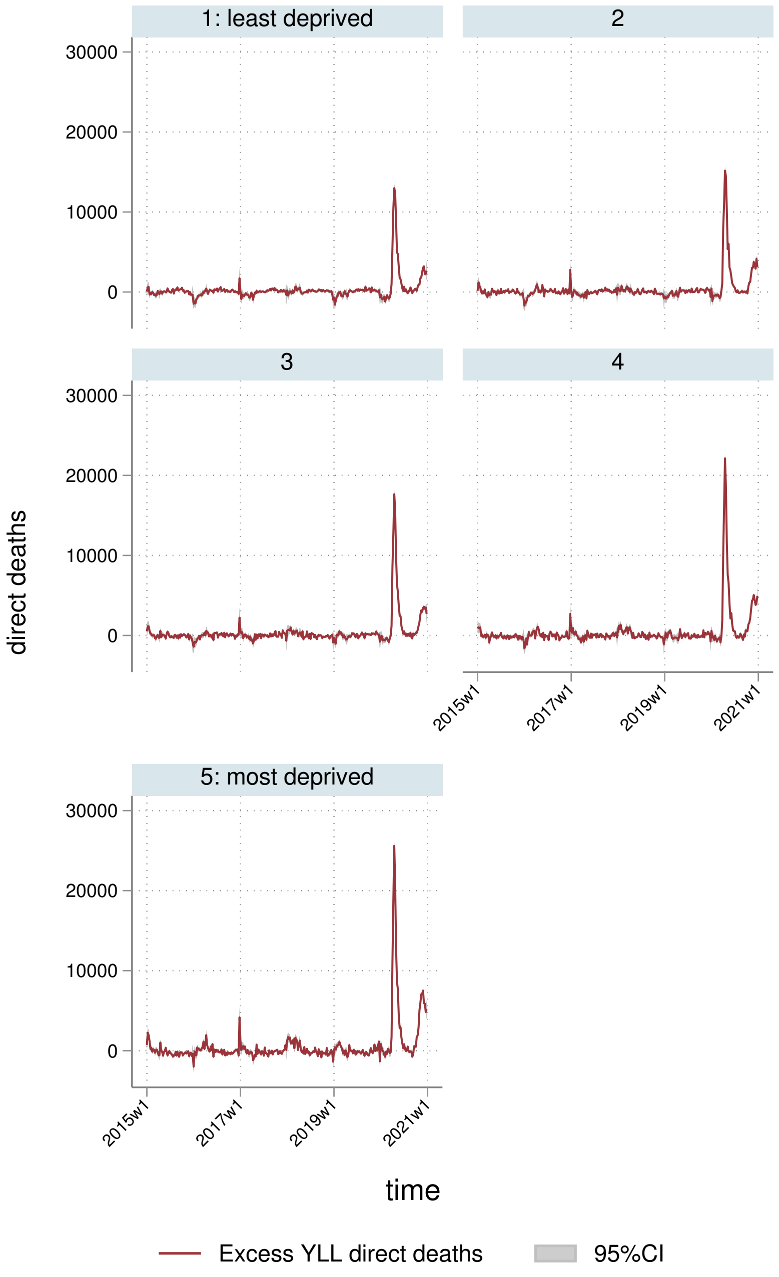
1: least deprived
2
3
4
5: most deprived
30000
20000
10000
0
direct deaths
2015w1
2017w1
2019w1
2021w1
time
Excess YLL direct deaths
95%CI

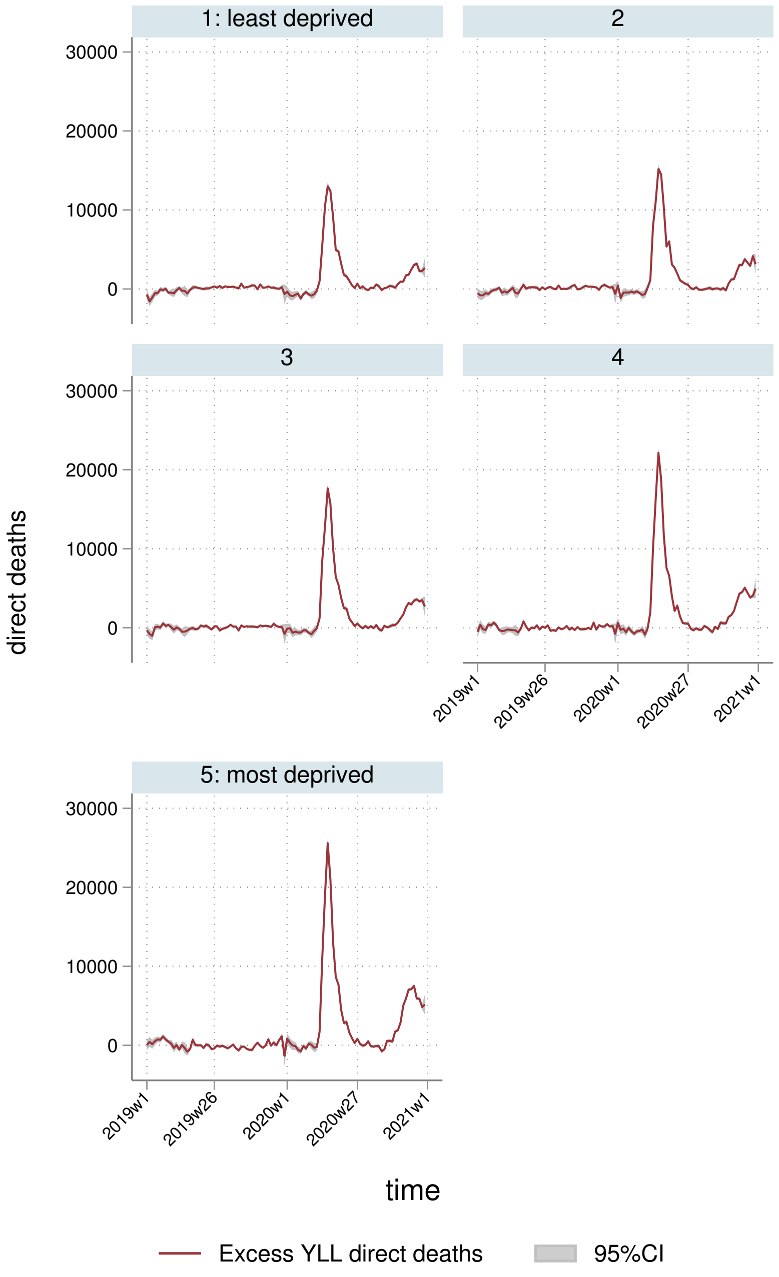
1: least deprived
2
3
4
5: most deprived
30000
20000
10000
0
direct deaths
2019w1
2019w26
2020w1
2020w27
2021w1
time
Excess YLL direct deaths
95%CI

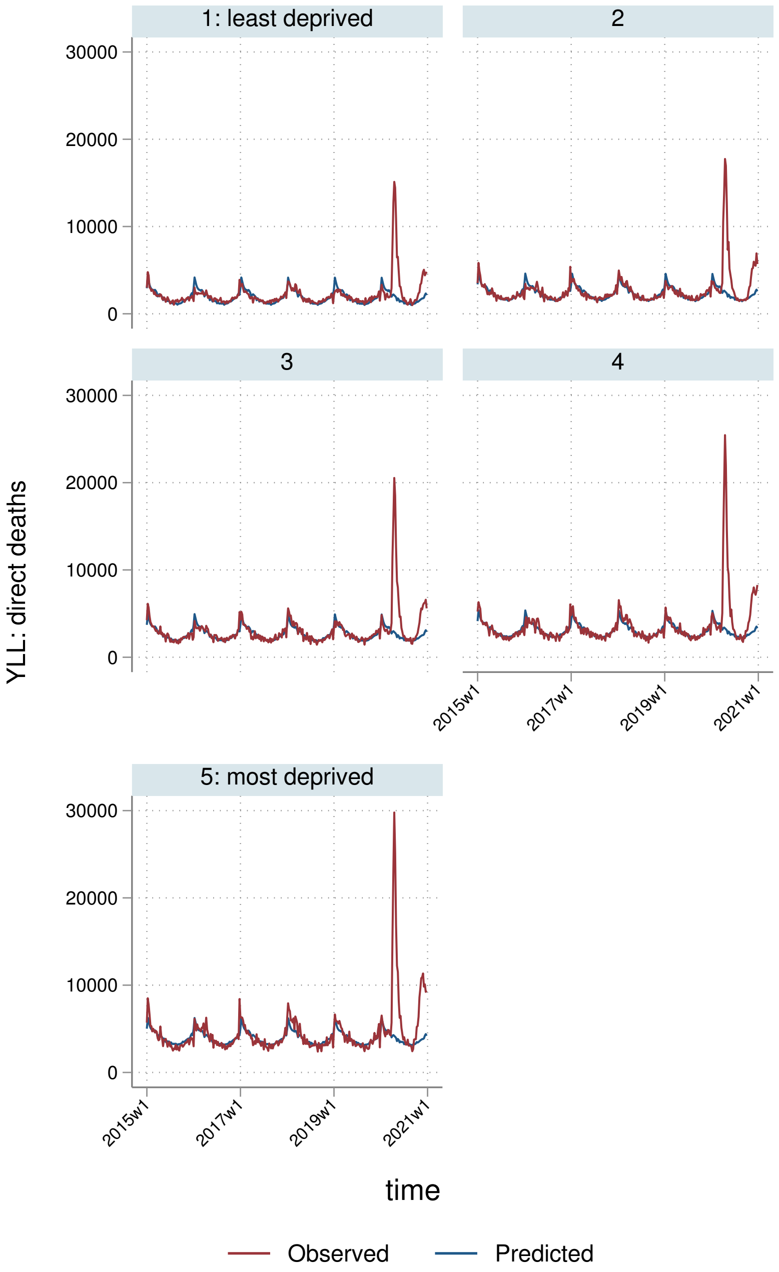

1: least deprived
2
3
4
5: most deprived
30000
20000
10000
0
YLL: direct deaths
2015w1
2017w1
2019w1
2021w1
time
Observed
Predicted

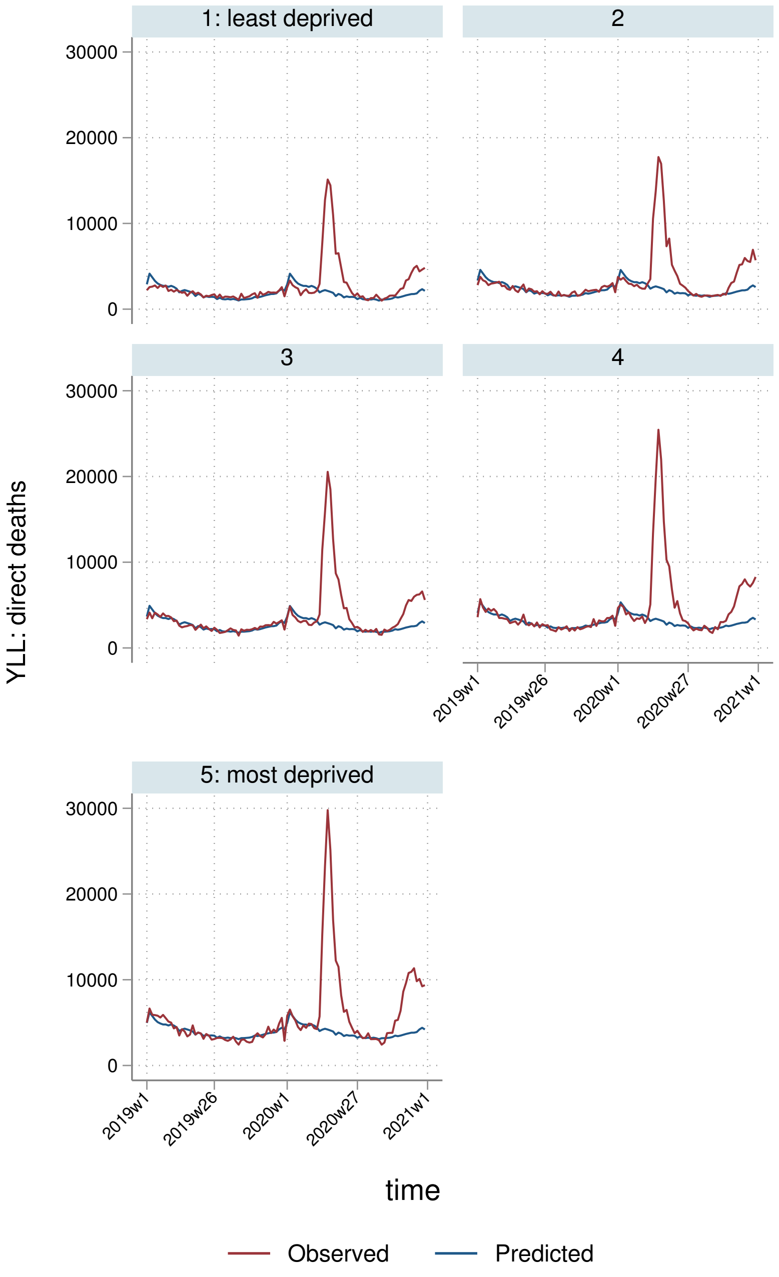

1: least deprived
2
3
4
5: most deprived
YLL: direct deaths
30000
20000
10000
0
2019w1
2019w26
2020w1
2020w27
2021w1
time
Observed
Predicted

##### 6.2.4 By Strategic Health Authority

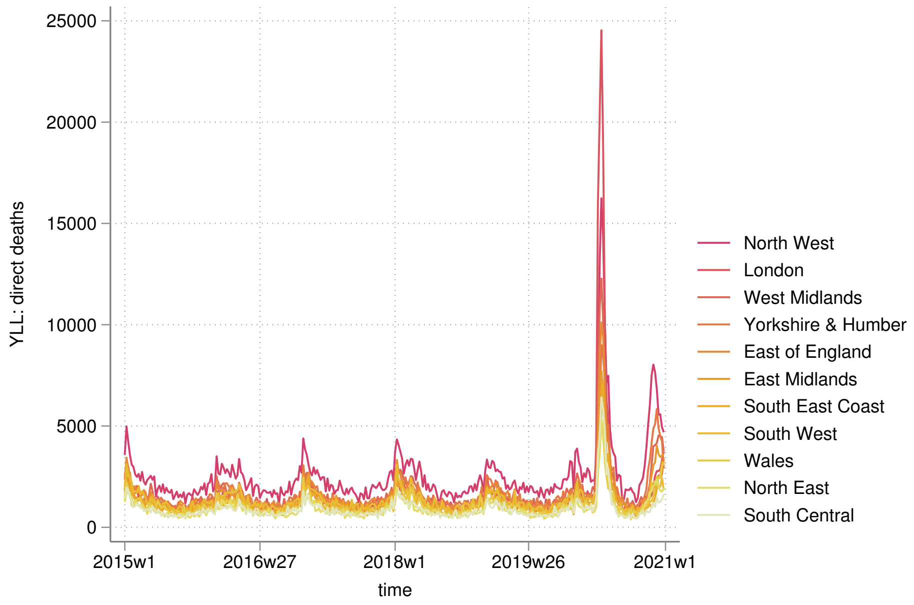


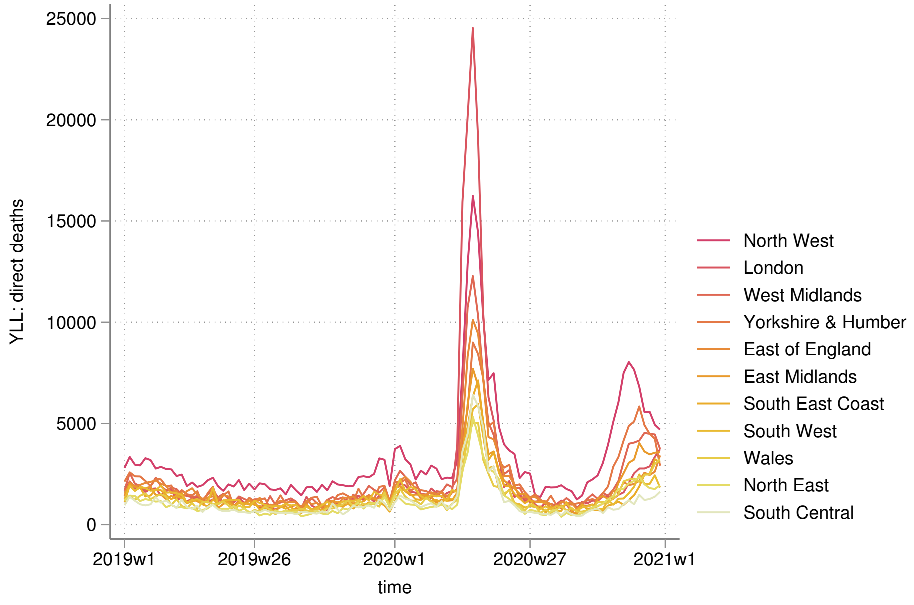

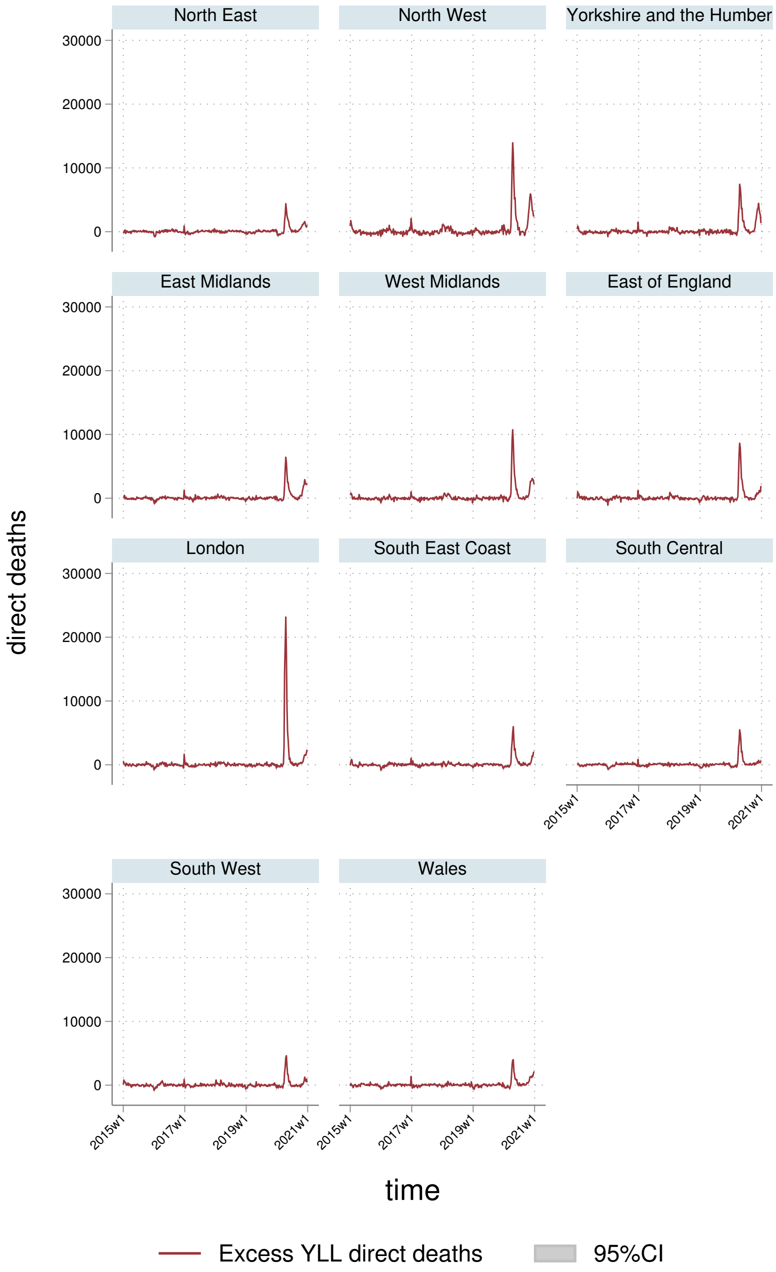
North East
North West
Yorkshire and the Humber
East Midlands
West Midlands
East of England
London
South East Coast
South Central
South West
Wales
30000
20000
10000
0
2015w1
2017w1
2019w1
2021w1
direct deaths
time
Excess YLL direct deaths
95%CI

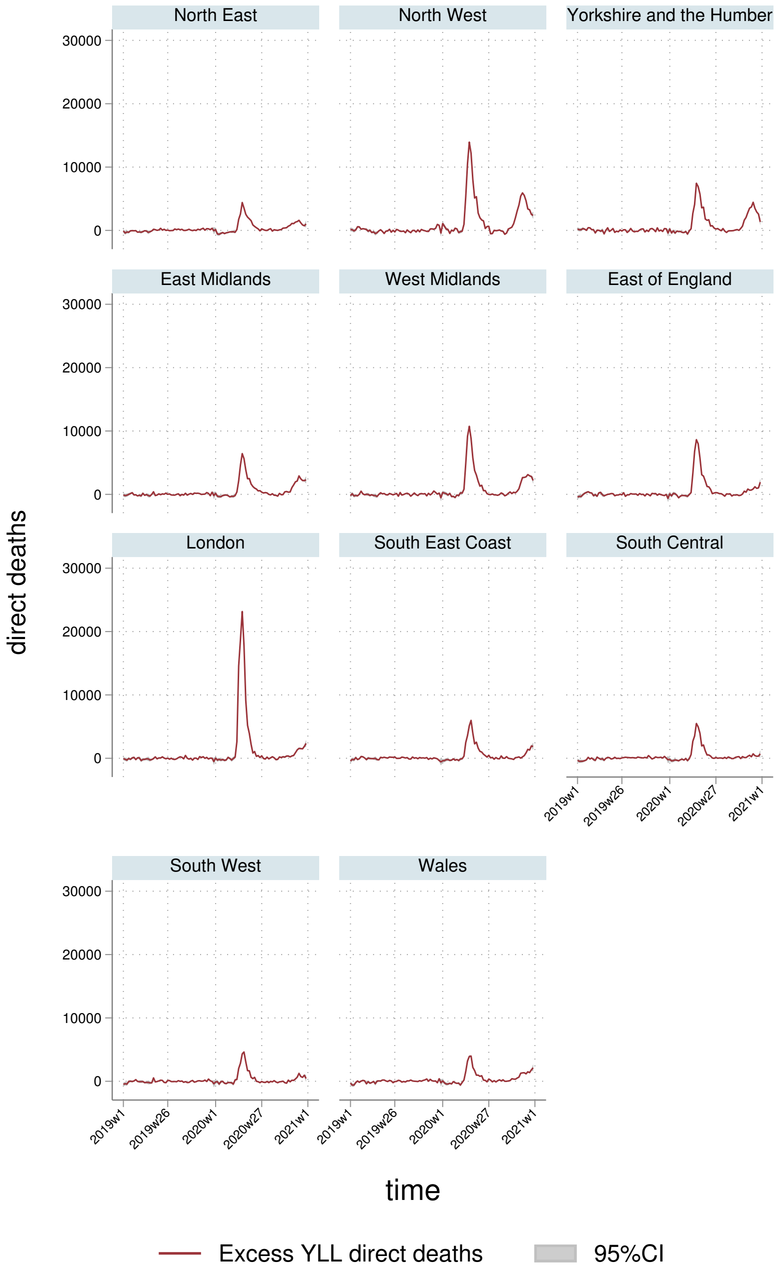
North East
North West
Yorkshire and the Humber
East Midlands
West Midlands
East of England
London
South East Coast
South Central
South West
Wales
30000
20000
10000
0
direct deaths
2019w1
2019w26
2020w1
2020w27
2021w1
time
Excess YLL direct deaths
95%CI

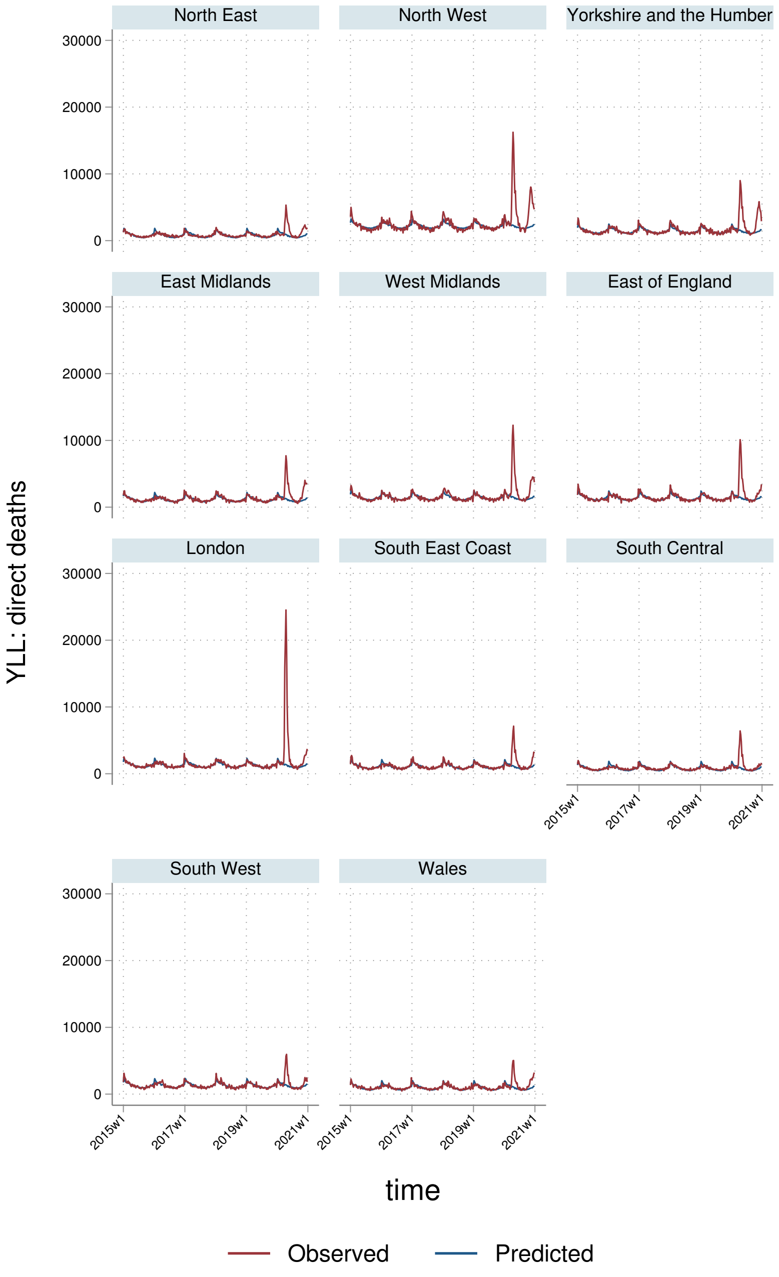
North East
North West
Yorkshire and the Humber
East Midlands
West Midlands
East of England
London
South East Coast
South Central
South West
Wales
YLL: direct deaths
30000
20000
10000
0
2015w1
2017w1
2019w1
2021w1
time
Observed
Predicted

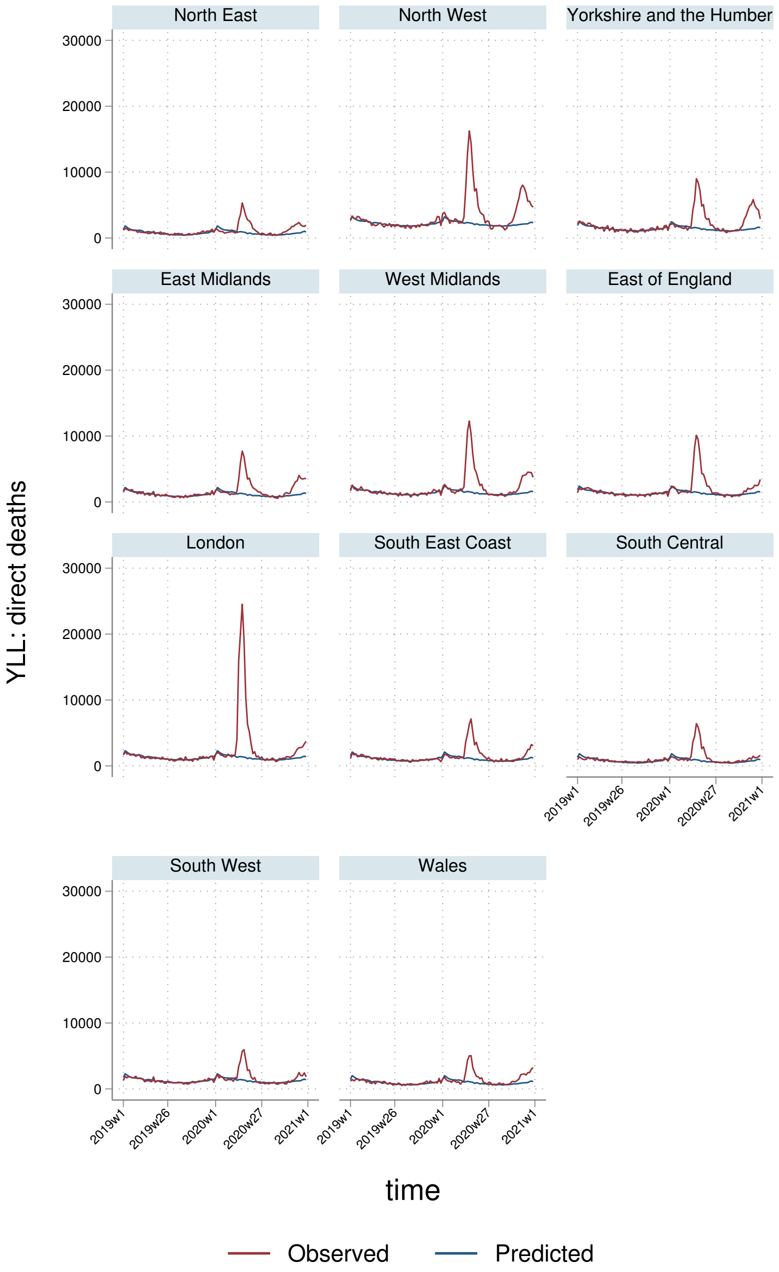
North East
North West
Yorkshire and the Humber
East Midlands
West Midlands
East of England
London
South East Coast
South Central
South West
Wales
YLL: direct deaths
30000
20000
10000
0
2019w1
2019w26
2020w1
2020w27
2021w1
time
Observed
Predicted

#### 6.3 YLLs per $100,000$ population

##### 6.3.1 England-Wales aggregate

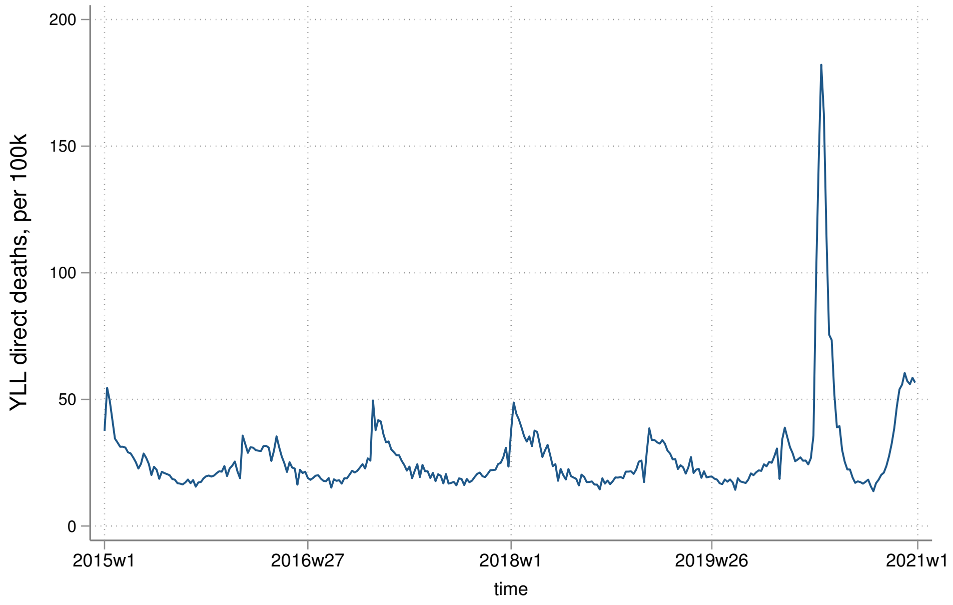


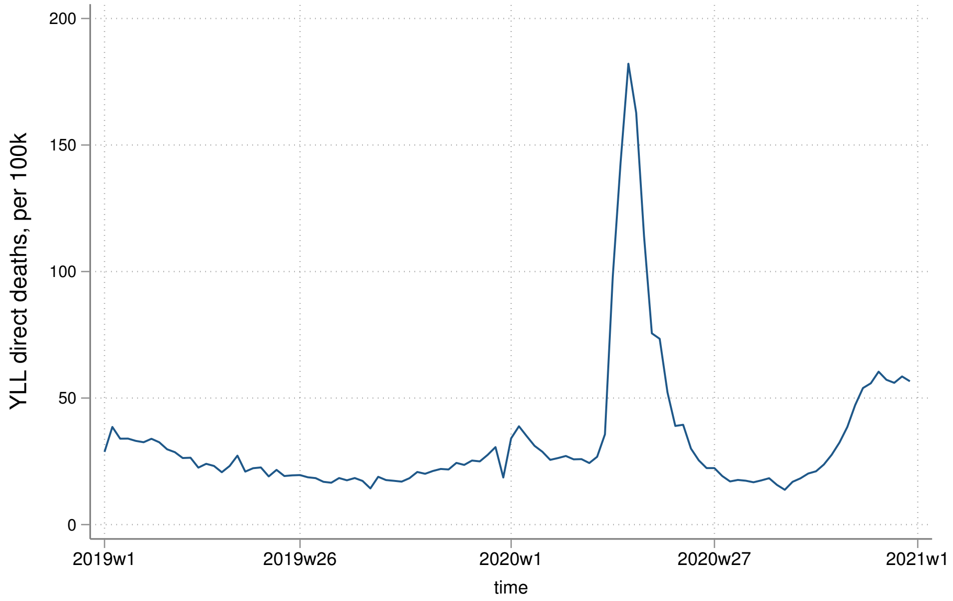

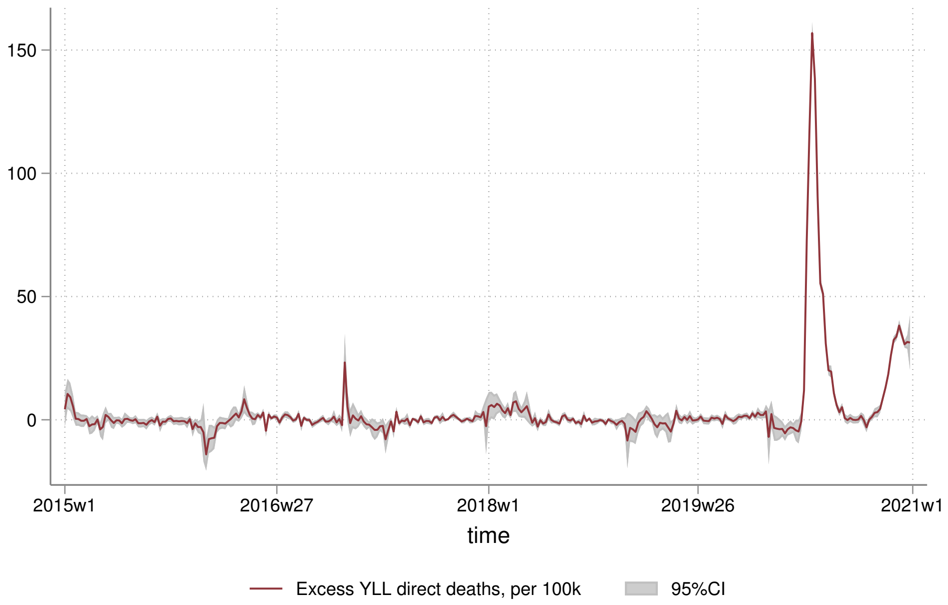
150
100
50
0
2015w1
2016w27
2018w1
2019w26
2021w1
time
Excess YLL direct deaths, per 100k
95%CI

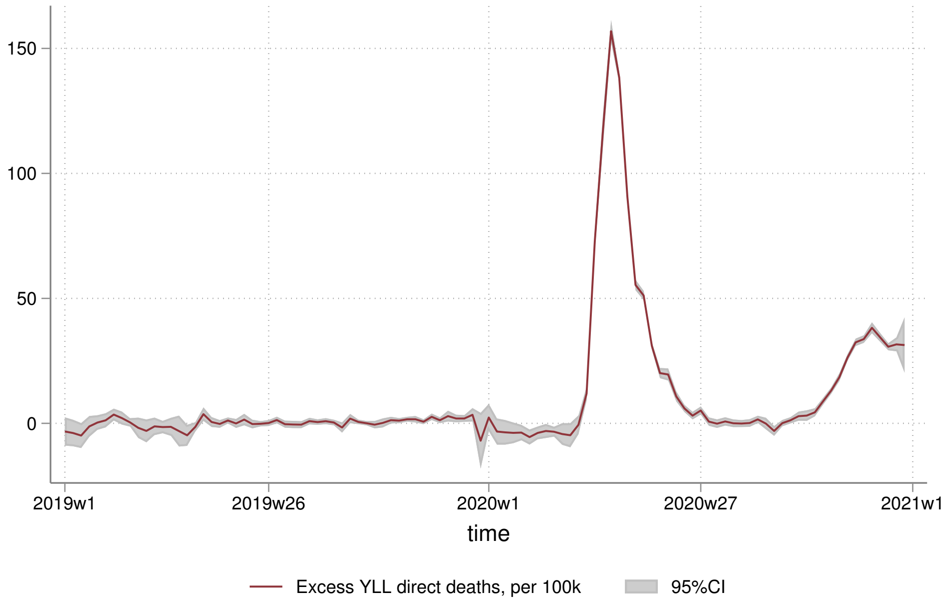
150
100
50
0
2019w1
2019w26
2020w1
2020w27
2021w1
time
Excess YLL direct deaths, per 100k
95%CI

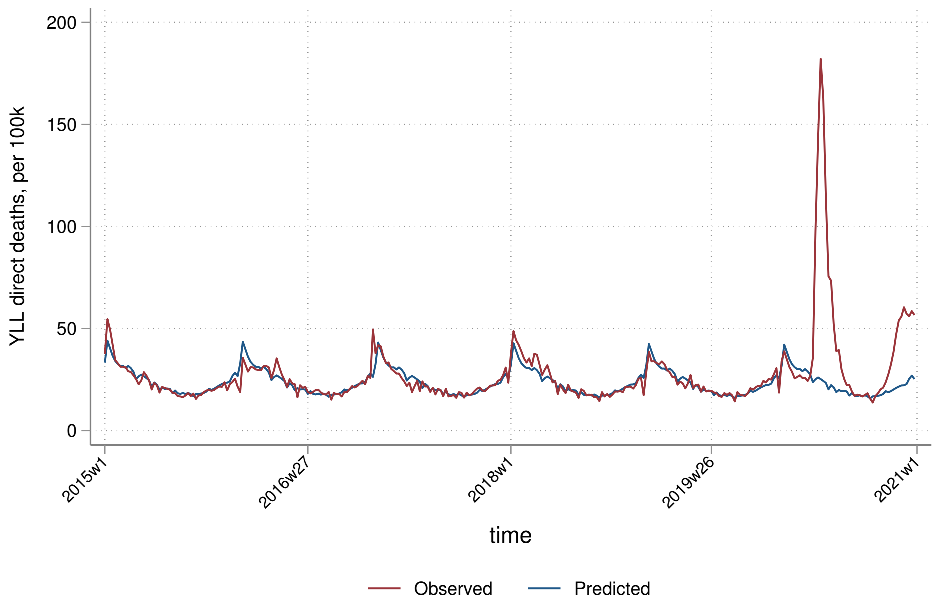
YLL direct deaths, per 100k
200
150
100
50
0
2015w1
2016w27
2018w1
2019w26
2021w1
time
Observed
Predicted

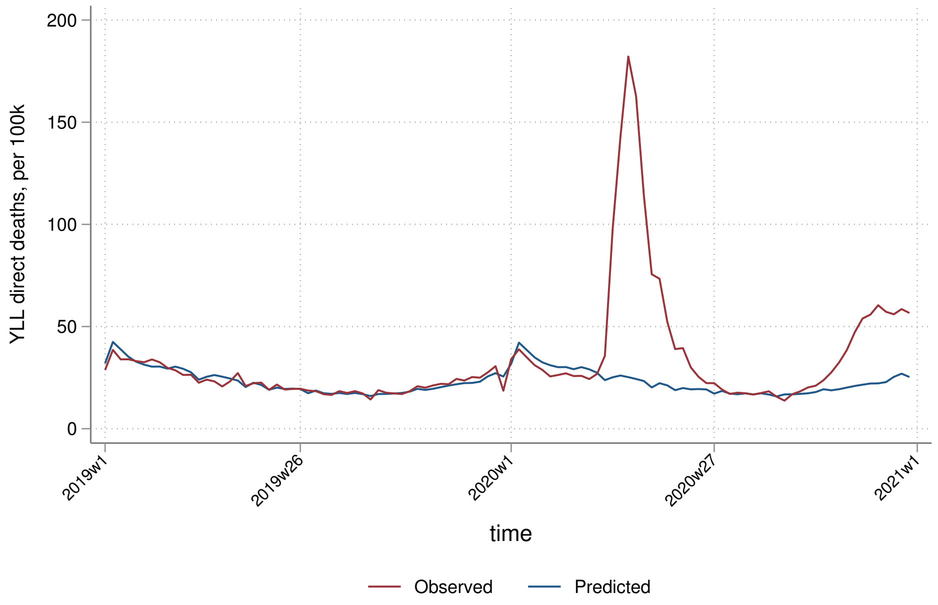
YLL direct deaths, per 100k
200
150
100
50
0
2019w1
2019w26
2020w1
2020w27
2021w1
time
Observed
Predicted

##### 6.3.2 By sex

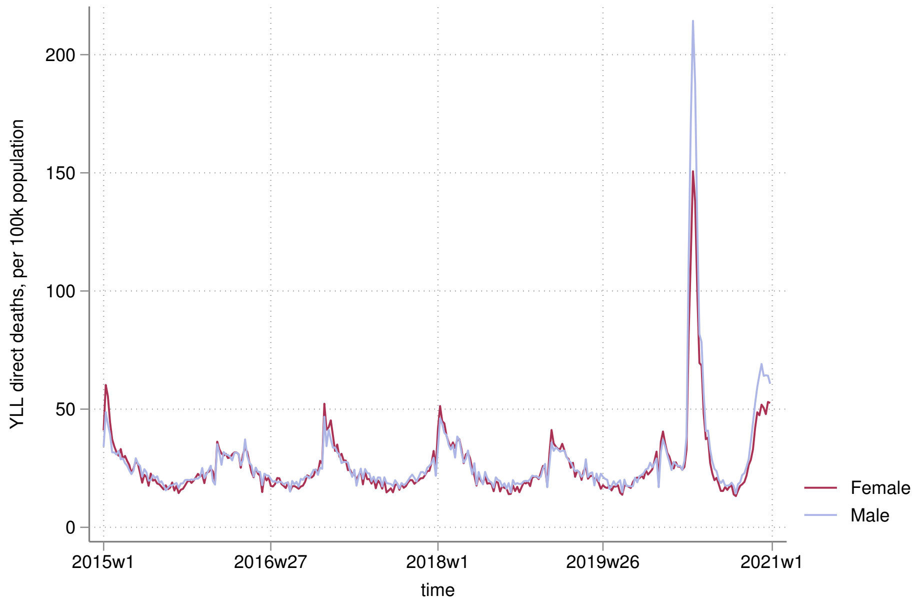

YLL direct deaths, per 100k population
0
50
100
150
200
2015w1
2016w27
2018w1
2019w26
2021w1
time
Female
Male


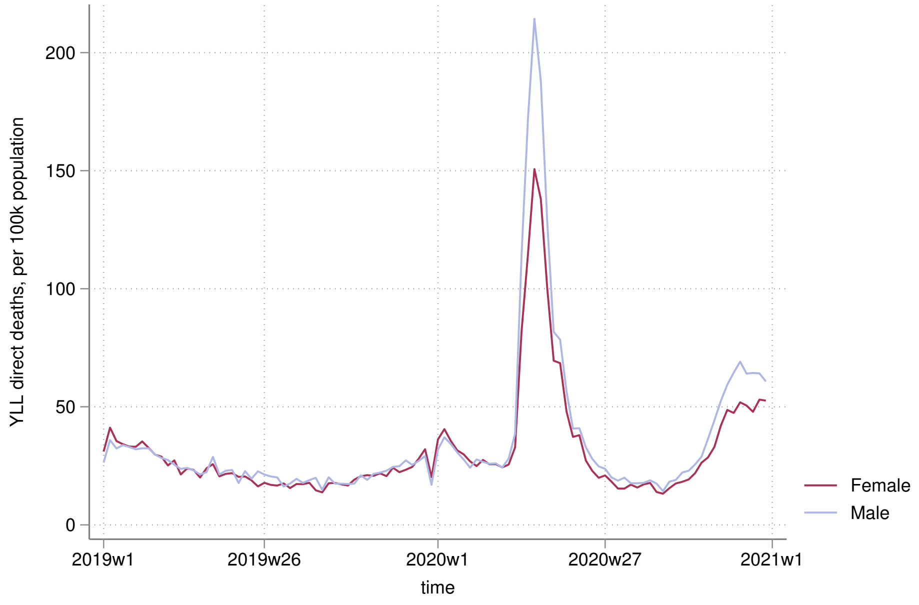

YLL direct deaths, per 100k population
0
50
100
150
200
2019w1
2019w26
2020w1
2020w27
2021w1
time
Female
Male

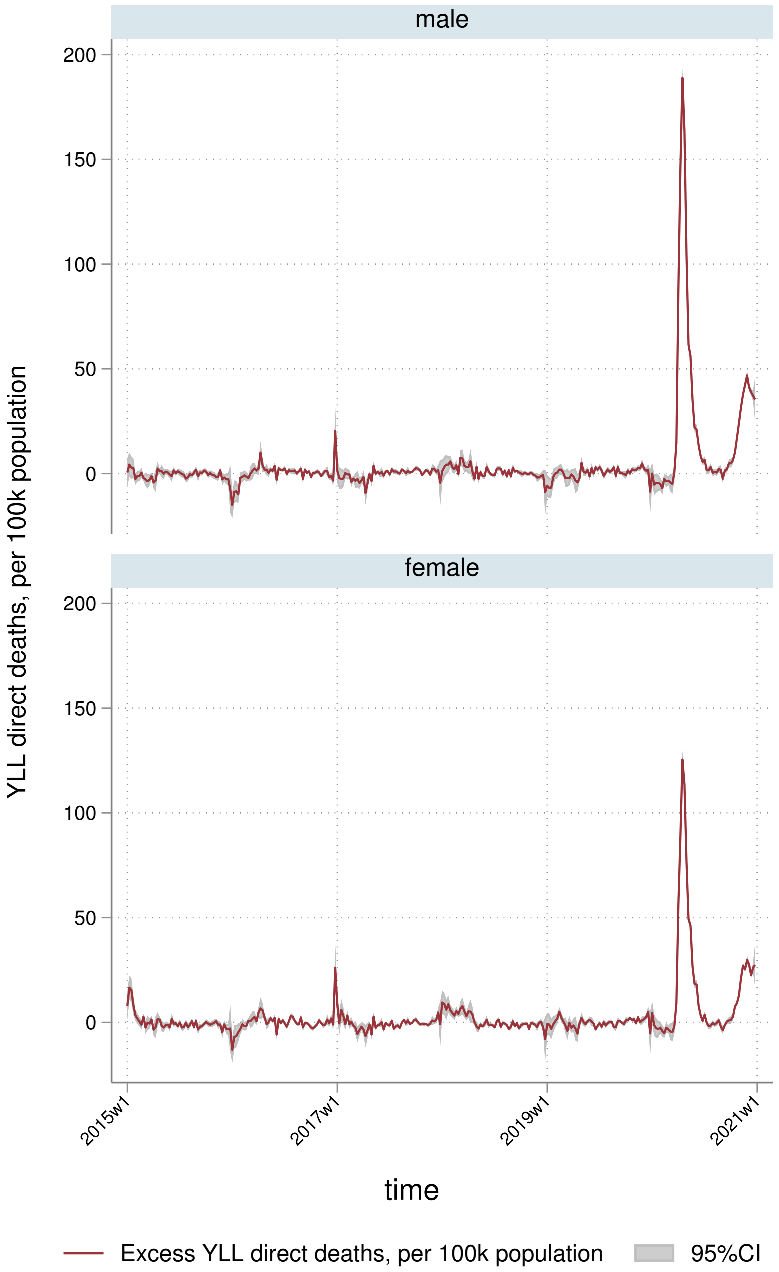
male
female
200
150
100
50
0
YLL direct deaths, per 100k population
2015w1
2017w1
2019w1
2021w1
time
Excess YLL direct deaths, per 100k population
95%CI

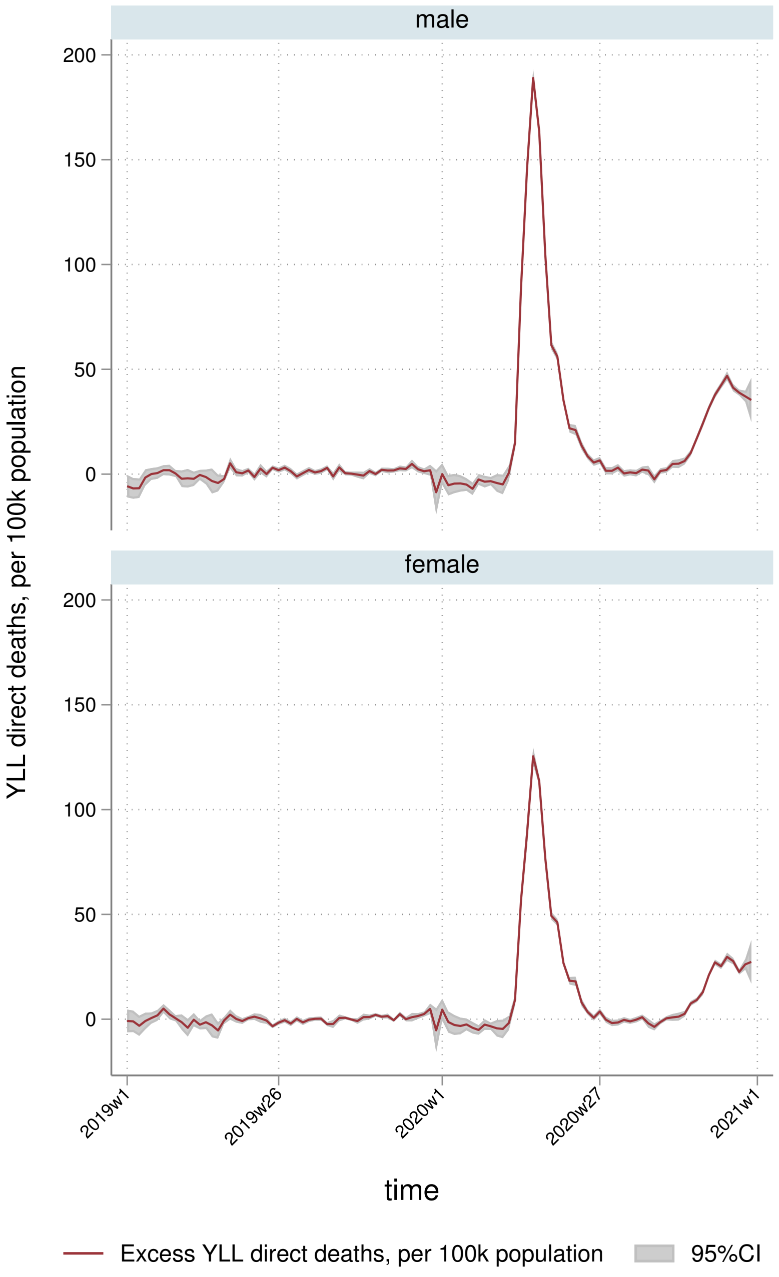
male
female
200
150
100
50
0
YLL direct deaths, per 100k population
2019w1
2019w26
2020w1
2020w27
2021w1
time
Excess YLL direct deaths, per 100k population
95%CI

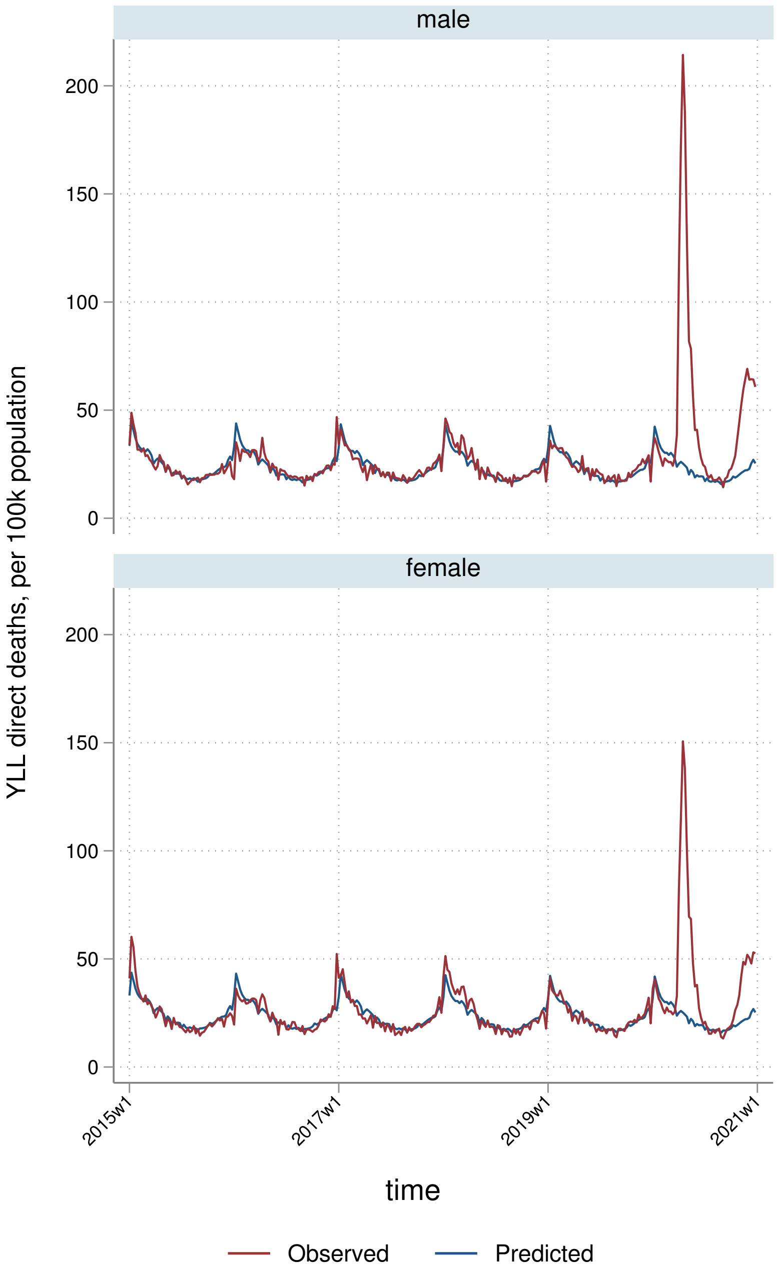

male
female
YLL direct deaths, per 100k population
200
150
100
50
0
2015w1
2017w1
2019w1
2021w1
time
Observed
Predicted

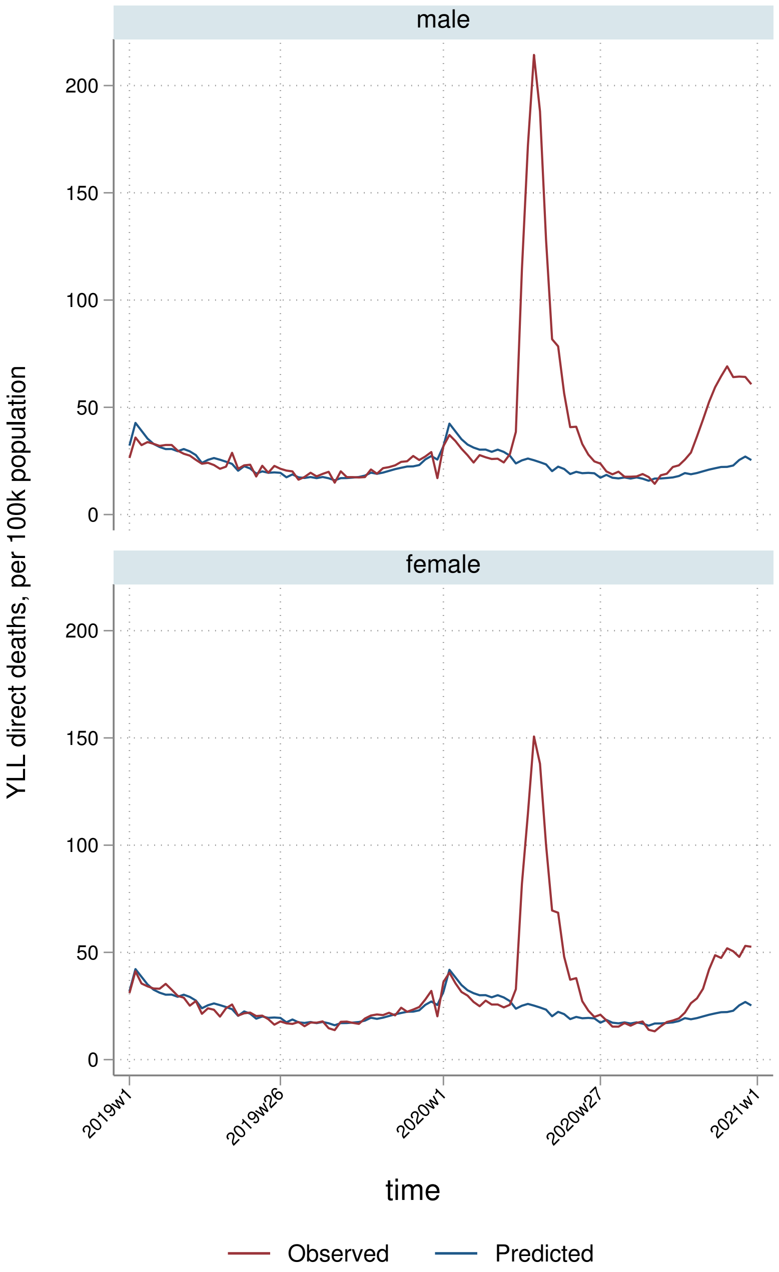

male
female
YLL direct deaths, per 100k population
200
150
100
50
0
2019w1
2019w26
2020w1
2020w27
2021w1
time
Observed
Predicted

##### 6.3.3 By deprivation quintile

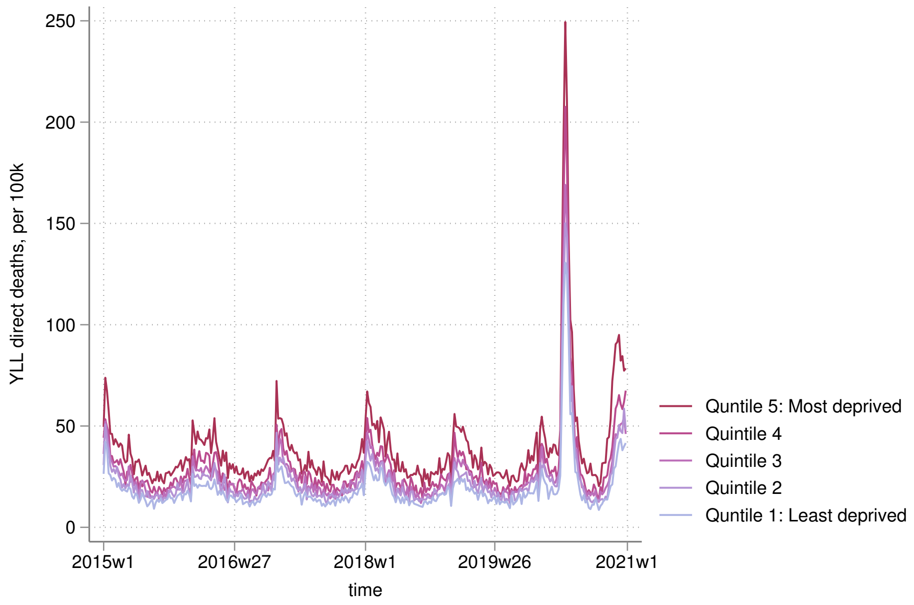


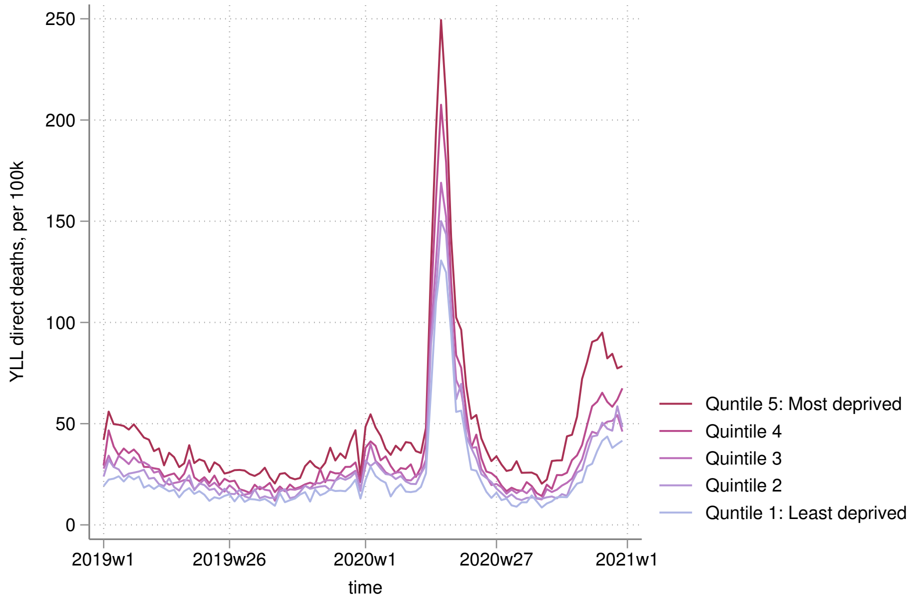

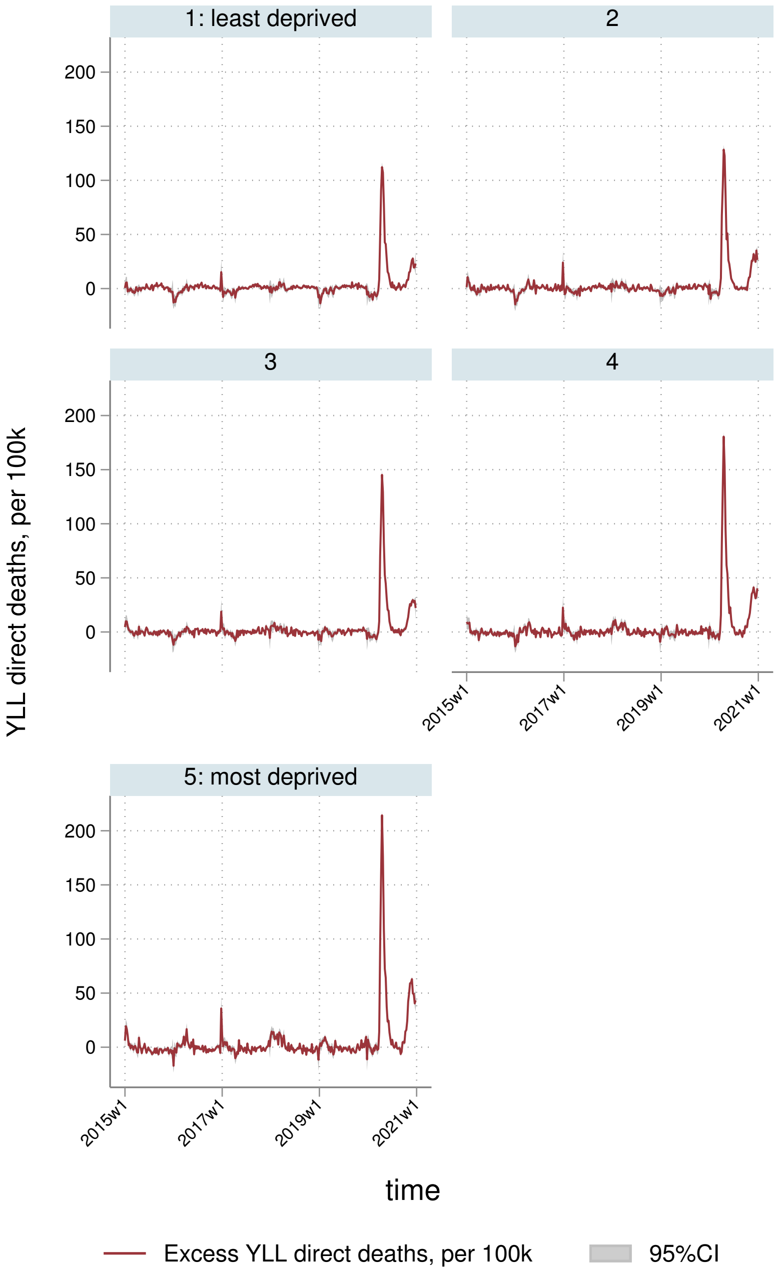
1: least deprived
2
3
4
5: most deprived
YLL direct deaths, per 100k
200
150
100
50
0
2015w1
2017w1
2019w1
2021w1
time
Excess YLL direct deaths, per 100k
95%CI

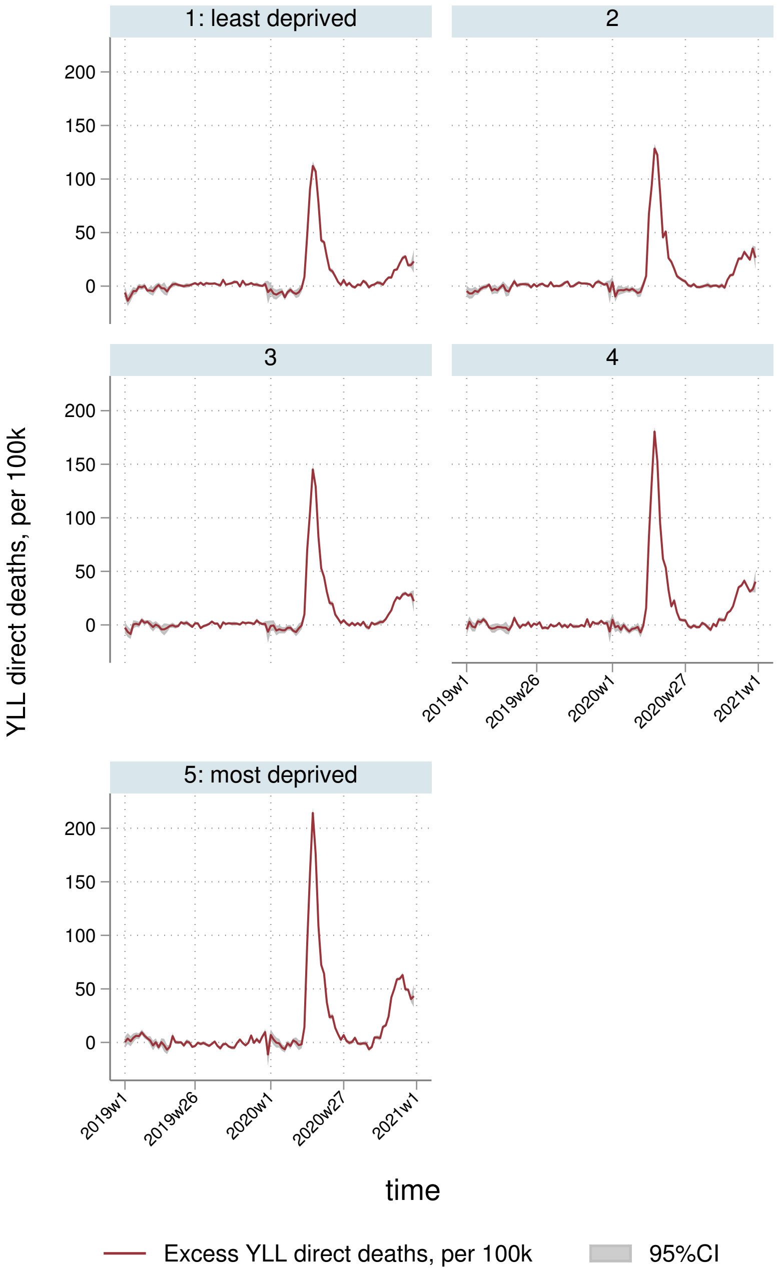
1: least deprived
2
3
4
5: most deprived
YLL direct deaths, per 100k
200
150
100
50
0
2019w1
2019w26
2020w1
2020w27
2021w1
time
Excess YLL direct deaths, per 100k
95%CI

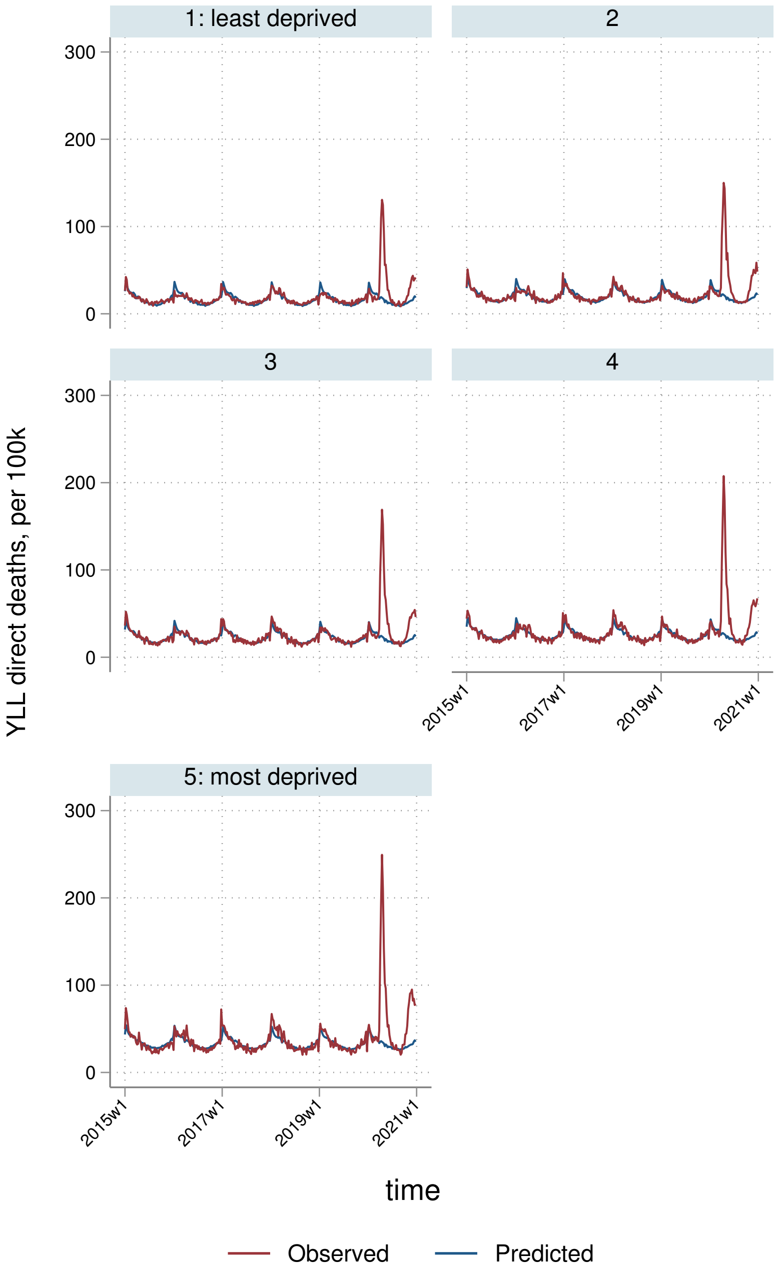
1: least deprived
2
3
4
5: most deprived
300
200
100
0
YLL direct deaths, per 100k
2015w1
2017w1
2019w1
2021w1
time
Observed
Predicted

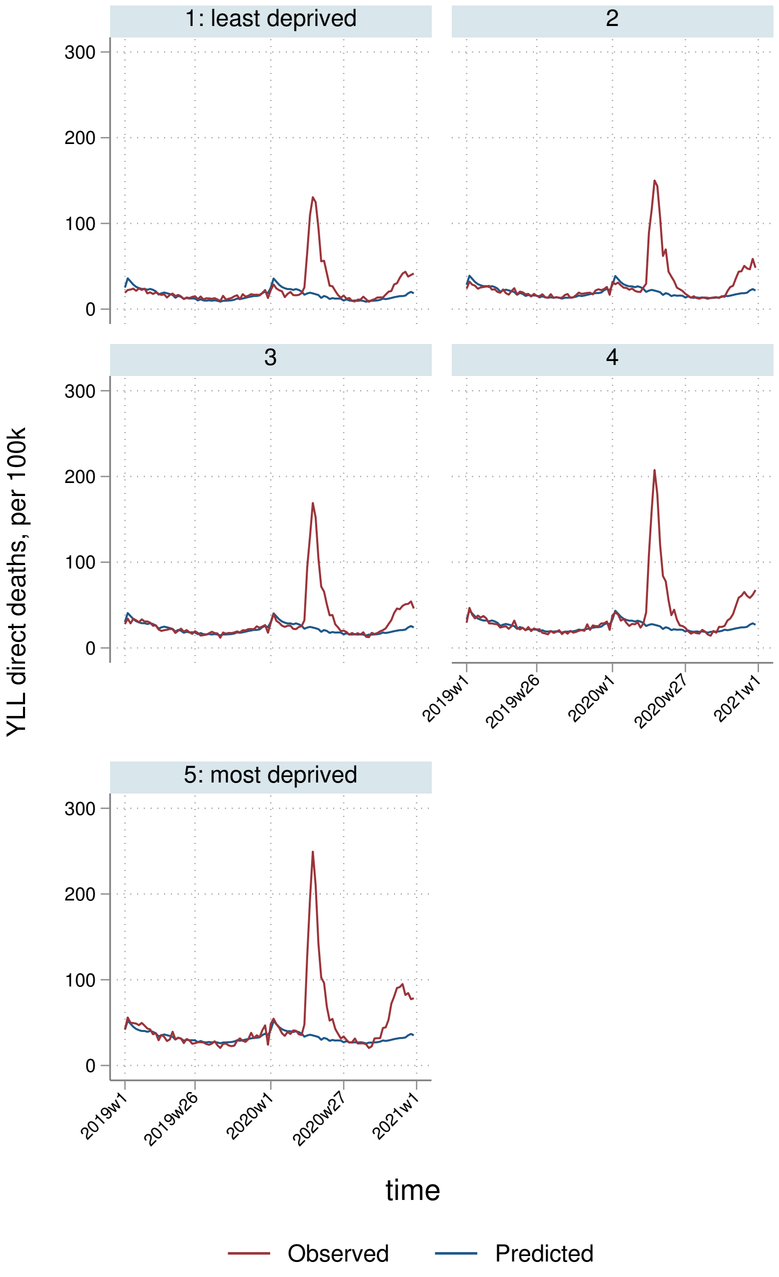
1: least deprived
2
3
4
5: most deprived
300
200
100
0
YLL direct deaths, per 100k
2019w1
2019w26
2020w1
2020w27
2021w1
time
Observed
Predicted

##### 6.3.4 By Strategic Health Authority

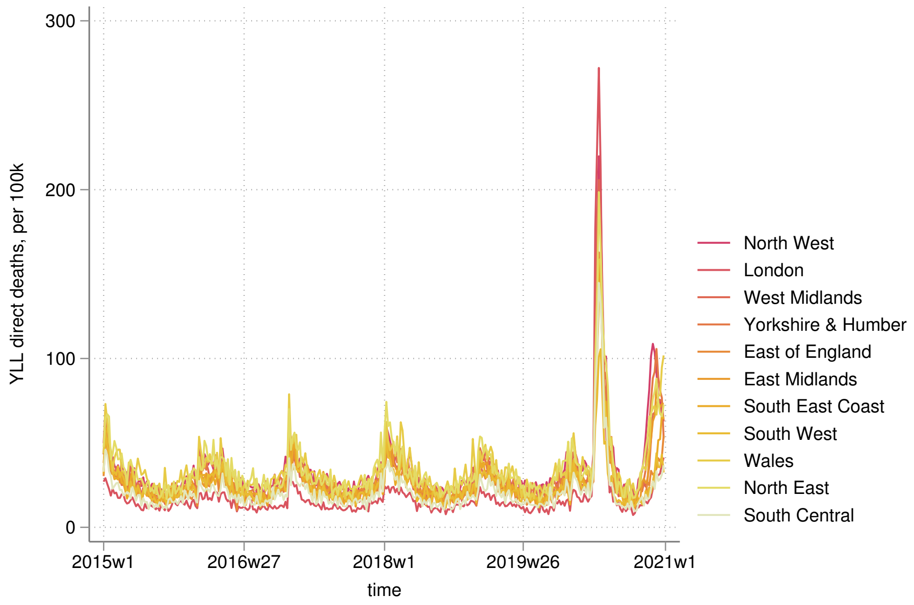


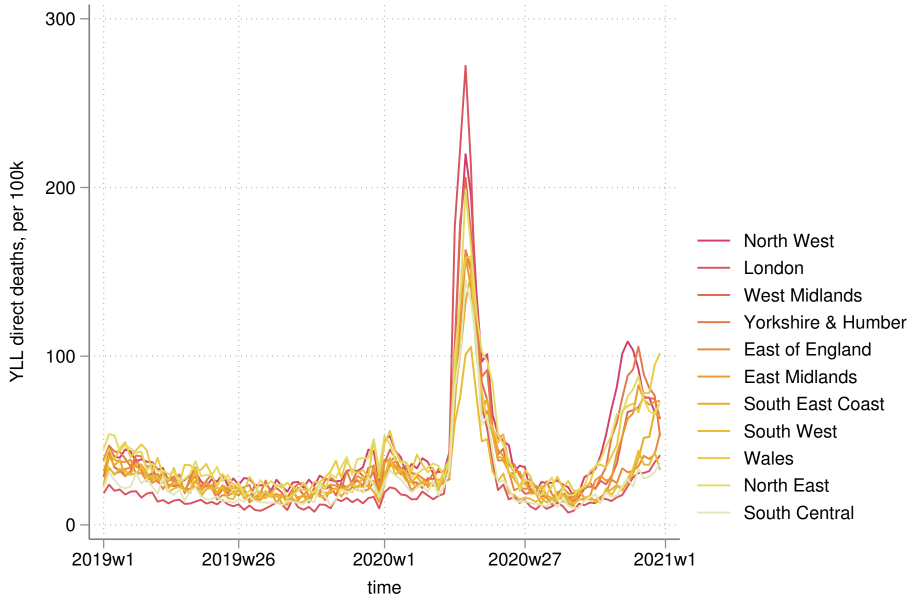

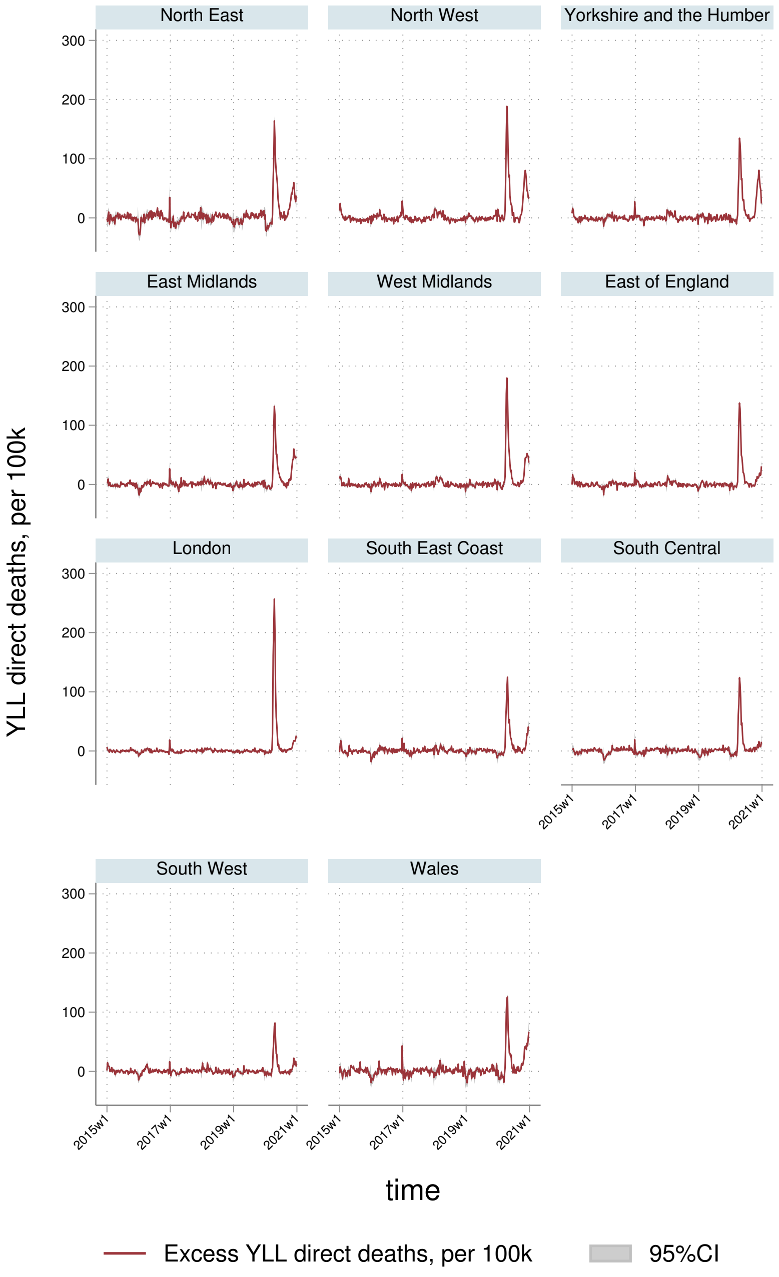
North East
North West
Yorkshire and the Humber
East Midlands
West Midlands
East of England
London
South East Coast
South Central
South West
Wales
YLL direct deaths, per 100k
300
200
100
0
2015w1
2017w1
2019w1
2021w1
time
Excess YLL direct deaths, per 100k
95%CI

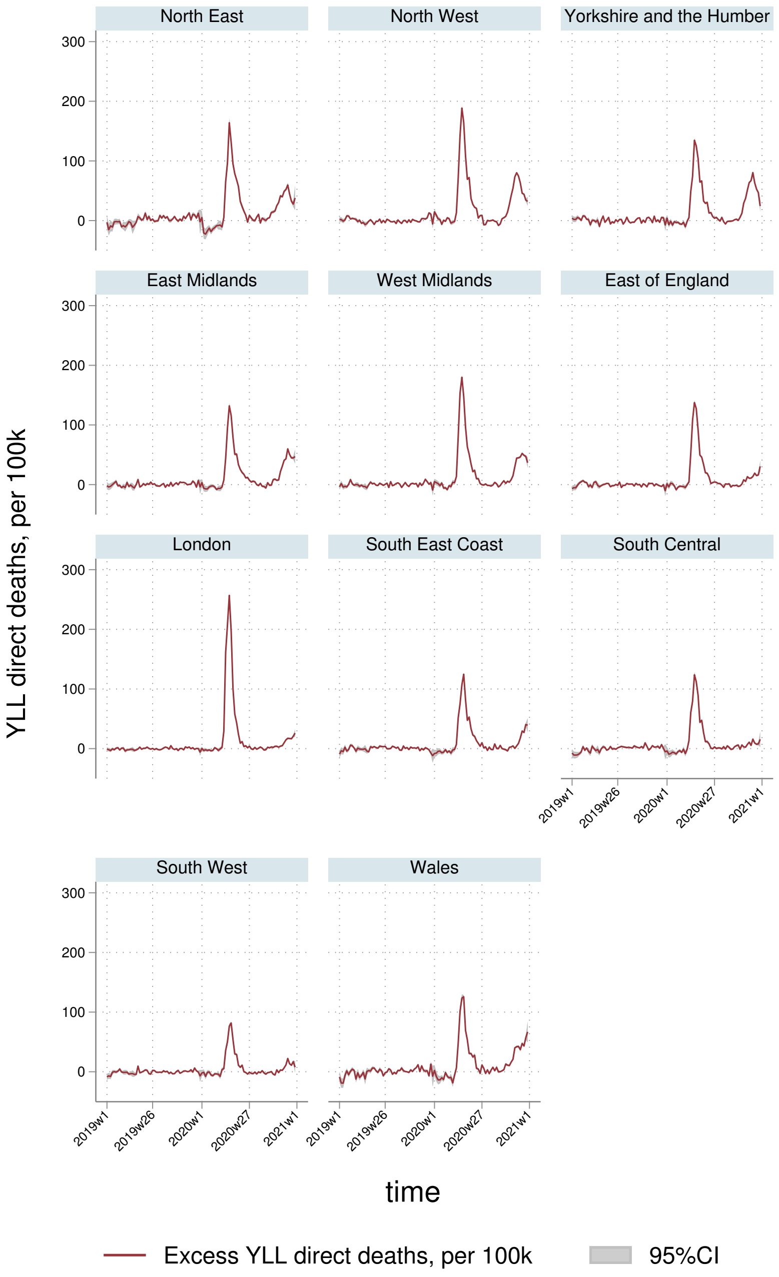
North East
North West
Yorkshire and the Humber
East Midlands
West Midlands
East of England
London
South East Coast
South Central
South West
Wales
300
200
100
0
YLL direct deaths, per 100k
2019w1
2019w26
2020w1
2020w27
2021w1
time
Excess YLL direct deaths, per 100k
95%CI

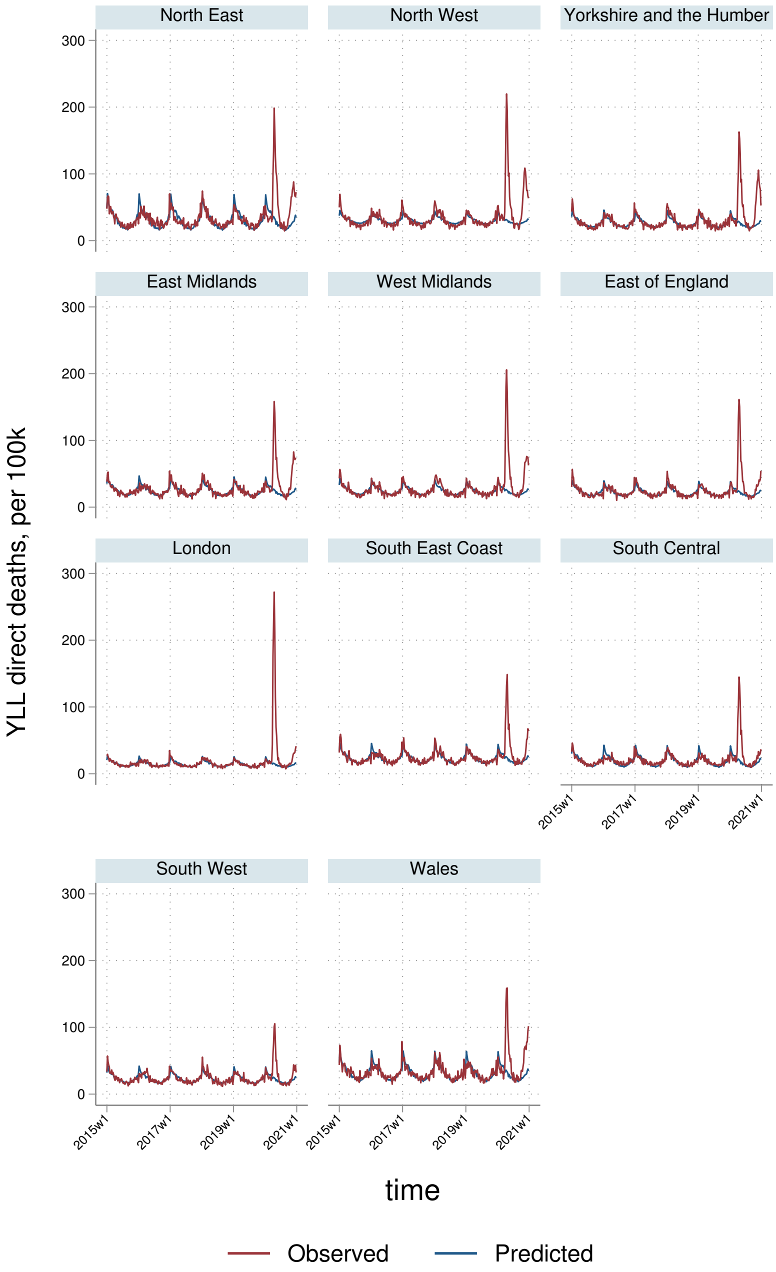
North East
North West
Yorkshire and the Humber
East Midlands
West Midlands
East of England
London
South East Coast
South Central
South West
Wales
YLL direct deaths, per 100k
300
200
100
0
2015w1
2017w1
2019w1
2021w1
time
Observed
Predicted

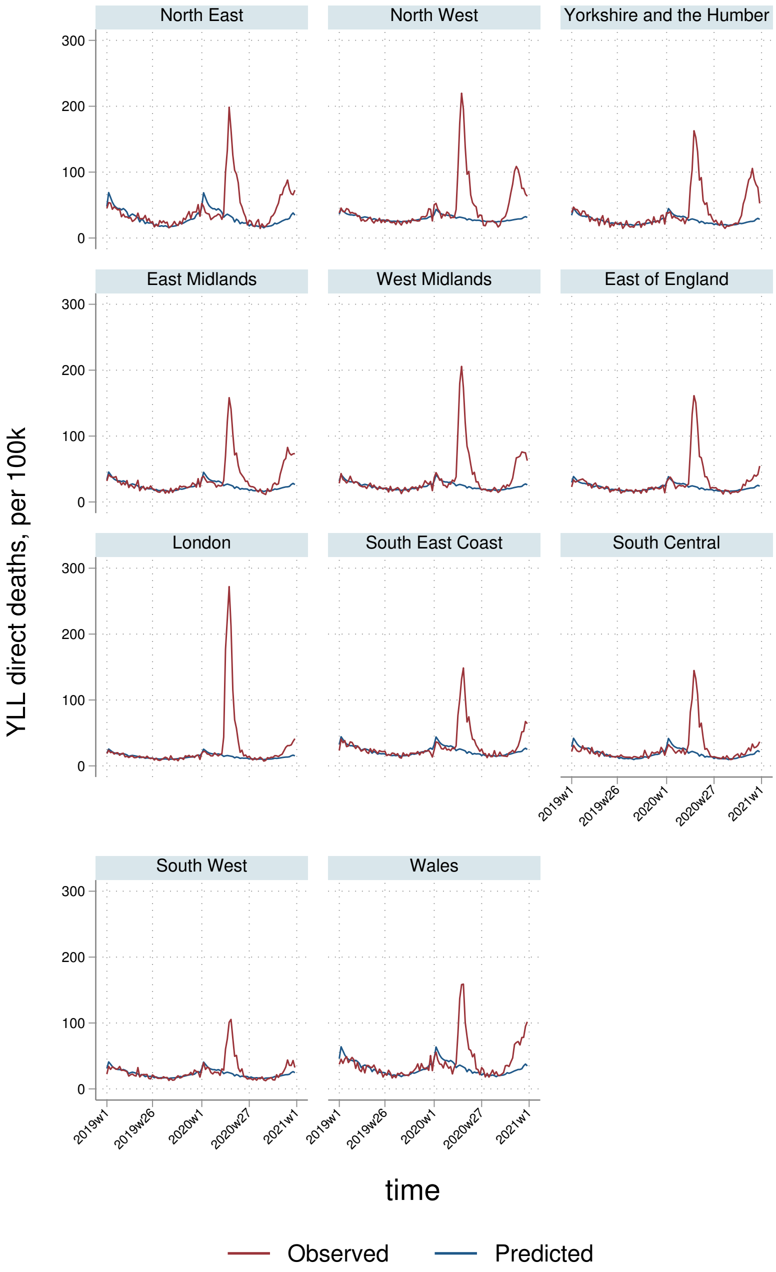

North East
North West
Yorkshire and the Humber
East Midlands
West Midlands
East of England
London
South East Coast
South Central
South West
Wales
YLL direct deaths, per 100k
300
200
100
0
2019w1
2019w26
2020w1
2020w27
2021w1
time
Observed
Predicted

### 7 Cardiovascular & diabetes

#### 7.1 AASMRs

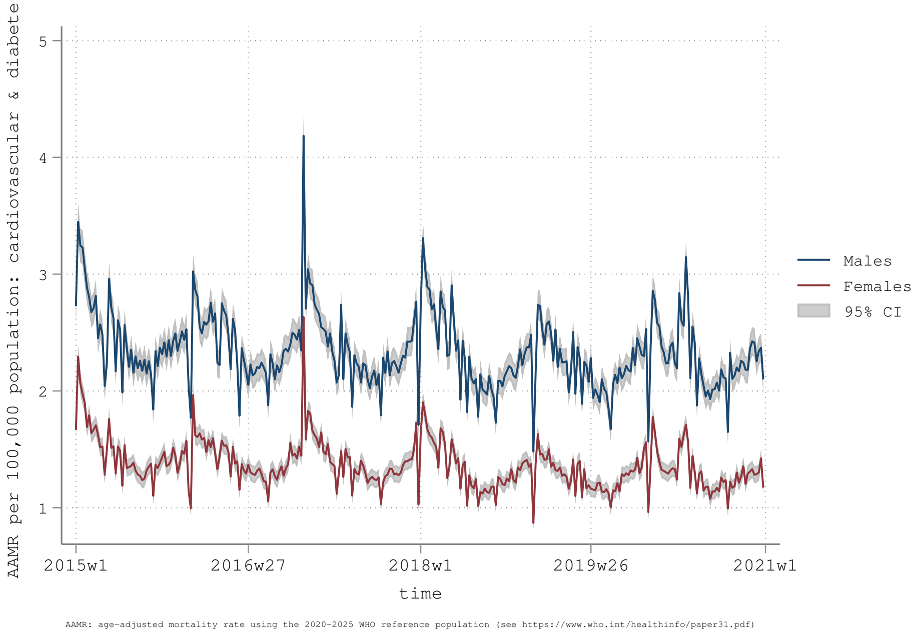


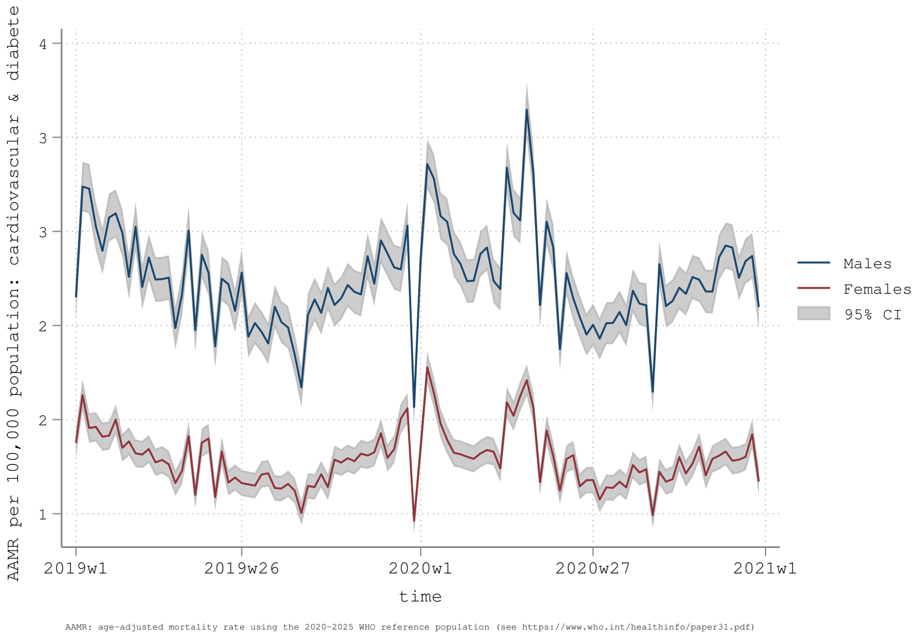

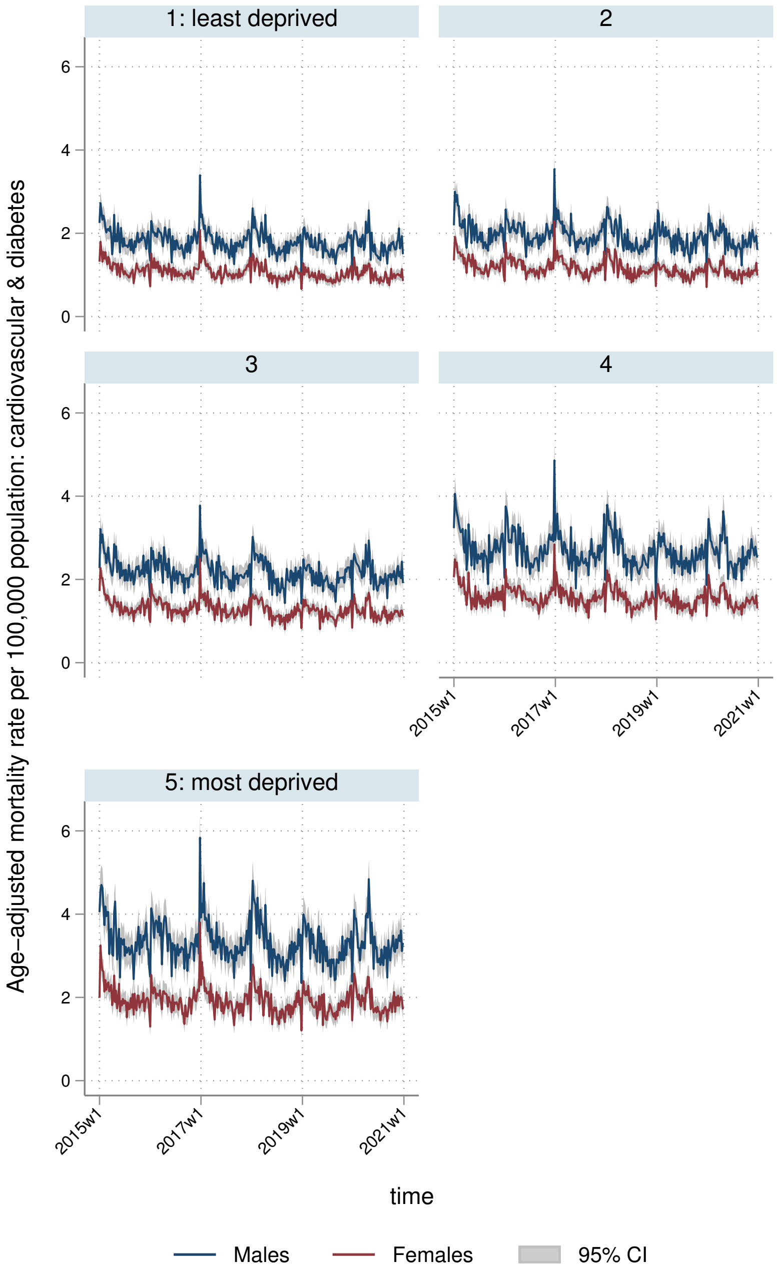
1: least deprived
2
3
4
5: most deprived
Age-adjusted mortality rate per 100,000 population: cardiovascular & diabetes
0
2
4
6
2015w1
2017w1
2019w1
2021w1
time
Males
Females
95% CI

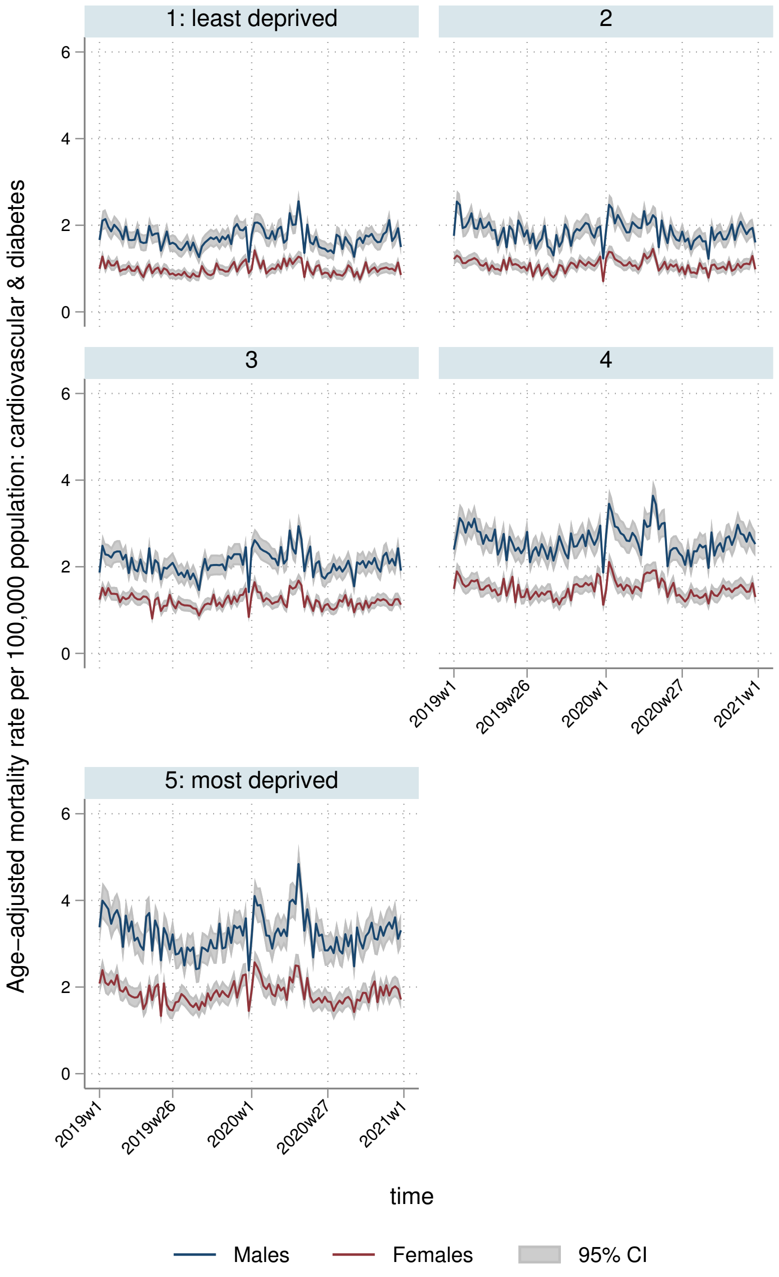
1: least deprived
2
3
4
5: most deprived
Age-adjusted mortality rate per 100,000 population: cardiovascular & diabetes
0
2
4
6
2019w1
2019w26
2020w1
2020w27
2021w1
time
Males
Females
95% CI

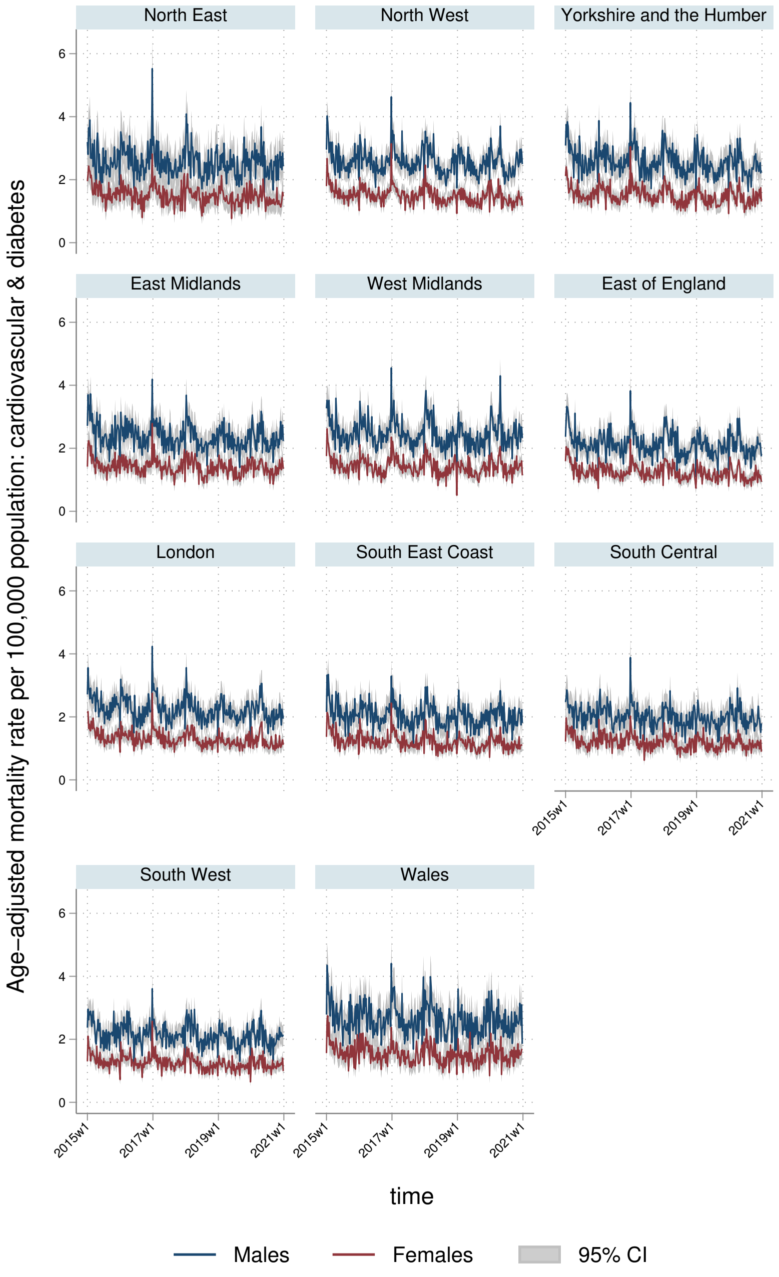
North East
North West
Yorkshire and the Humber
East Midlands
West Midlands
East of England
London
South East Coast
South Central
South West
Wales
0
2
4
6
2015w1
2017w1
2019w1
2021w1
Age-adjusted mortality rate per 100,000 population: cardiovascular & diabetes
time
Males
Females
95% CI

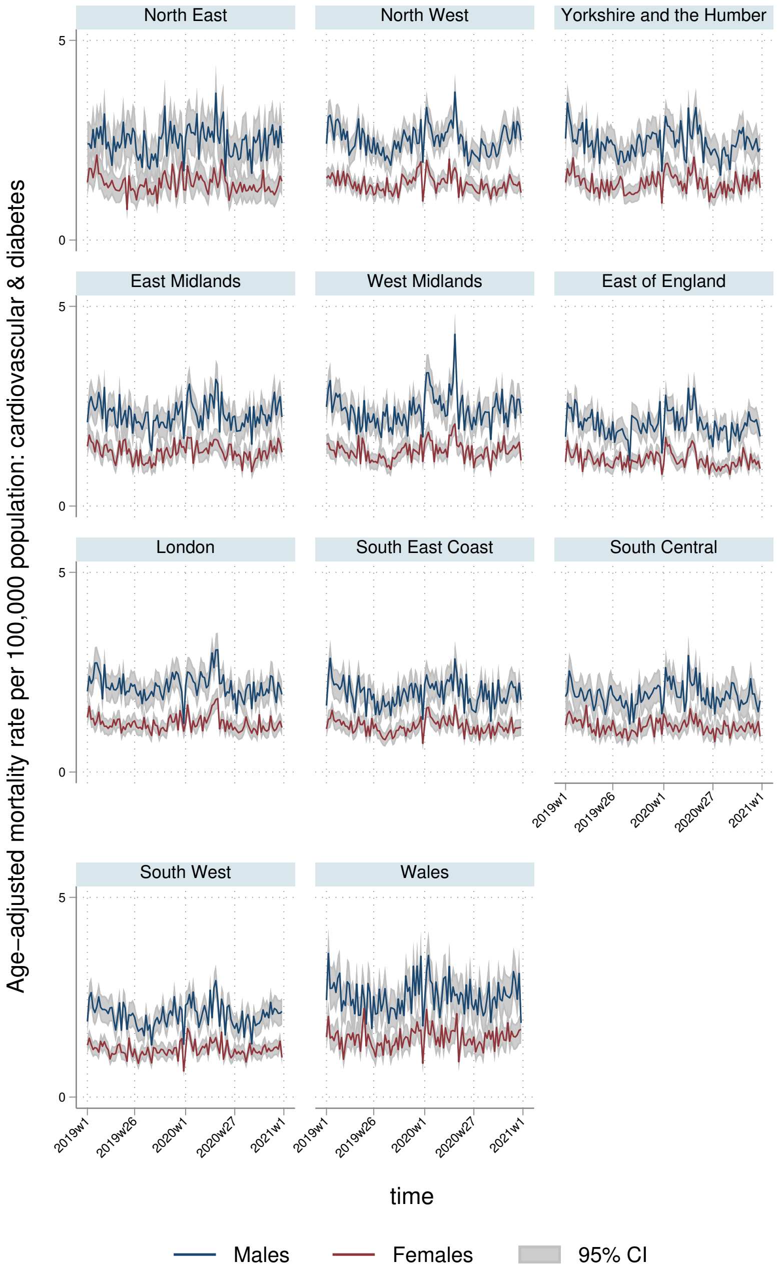
North East
North West
Yorkshire and the Humber
East Midlands
West Midlands
East of England
London
South East Coast
South Central
South West
Wales
Age-adjusted mortality rate per 100,000 population: cardiovascular & diabetes
0
5
2019w1
2019w26
2020w1
2020w27
2021w1
time
Males
Females
95% CI

#### 7.2 YLLs

##### 7.2.1 England-Wales aggregate

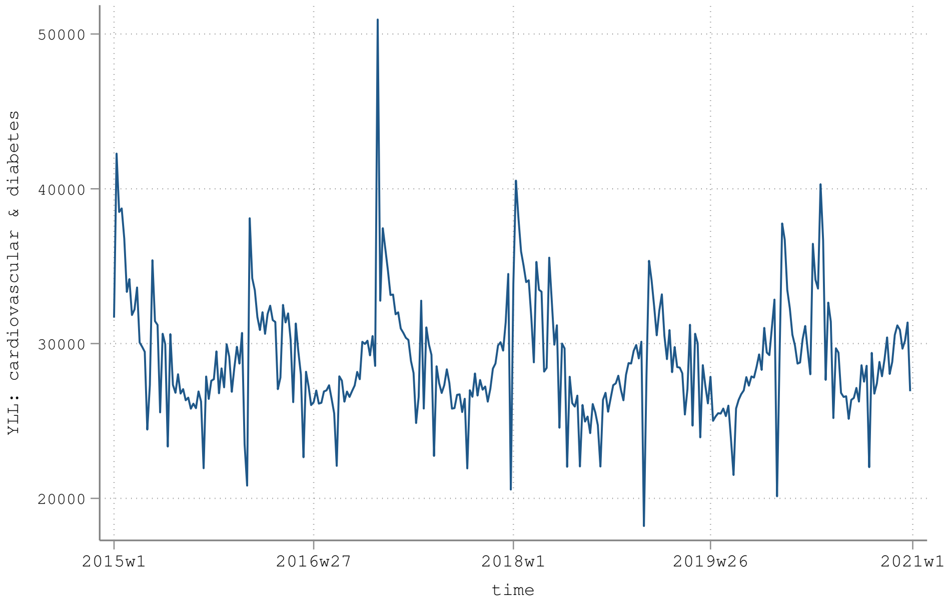


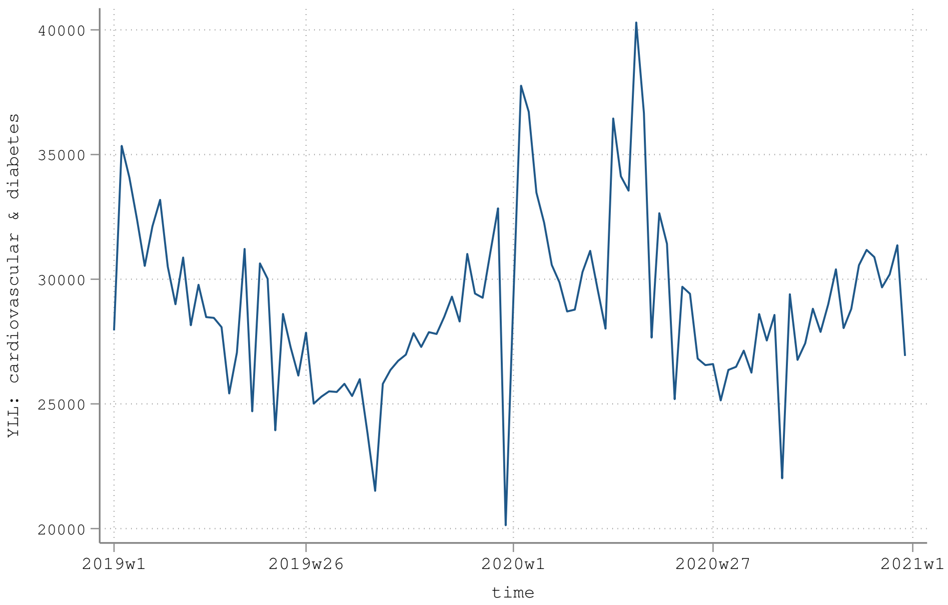

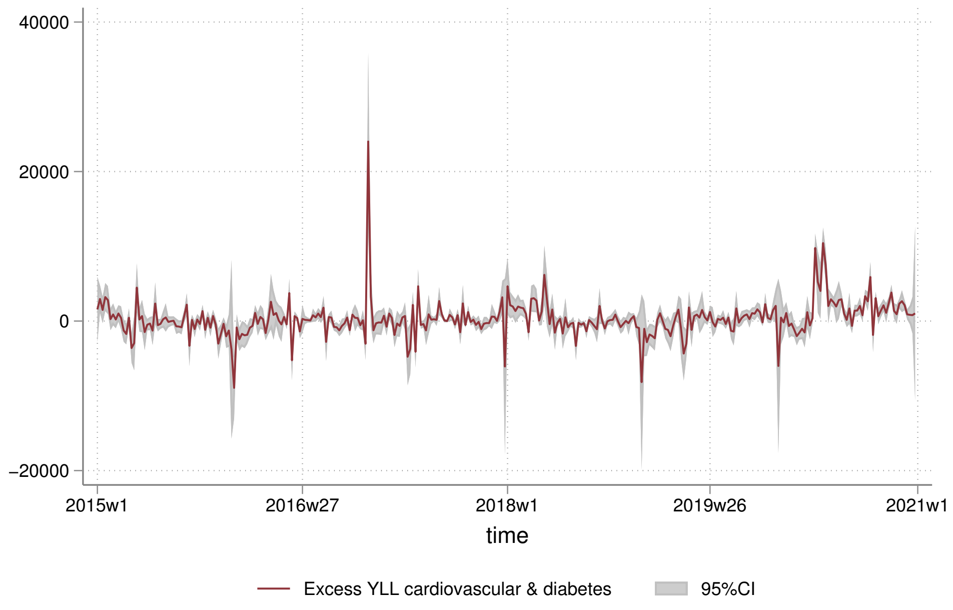
40000
20000
0
−20000
2015w1
2016w27
2018w1
2019w26
2021w1
time
Excess YLL cardiovascular & diabetes
95%CI

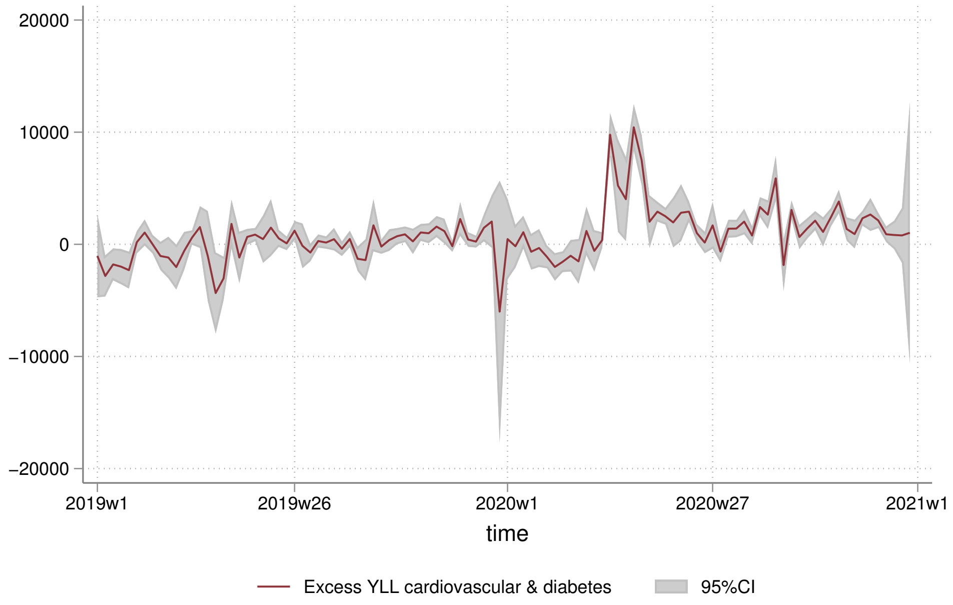
20000
10000
0
−10000
−20000
2019w1
2019w26
2020w1
2020w27
2021w1
time
Excess YLL cardiovascular & diabetes
95%CI

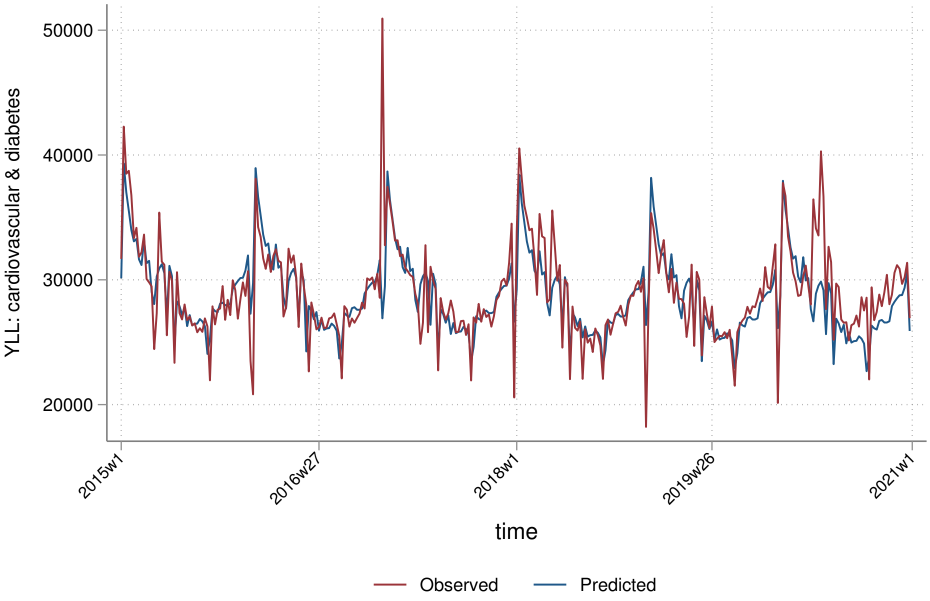

50000
40000
30000
20000
YLL: cardiovascular & diabetes
2015w1
2016w27
2018w1
2019w26
2021w1
time
Observed
Predicted


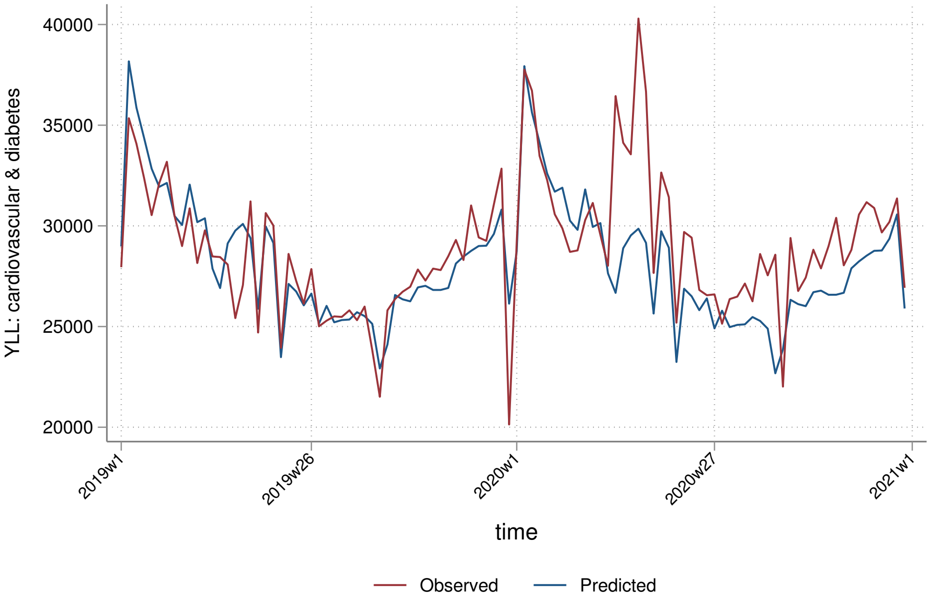

40000
35000
30000
25000
20000
YLL: cardiovascular & diabetes
2019w1
2019w26
2020w1
2020w27
2021w1
time
Observed
Predicted

##### 7.2.2 By sex

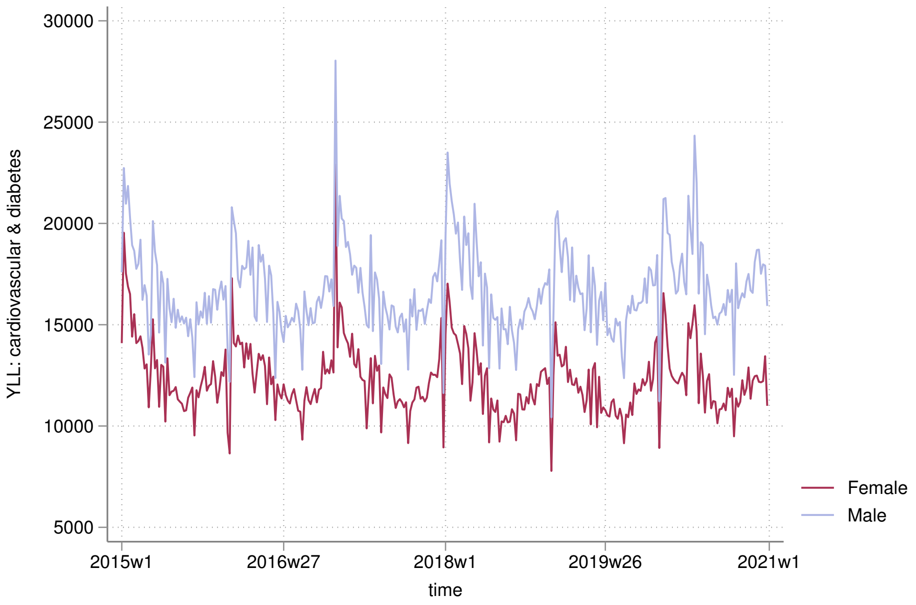

YLL: cardiovascular & diabetes
30000
25000
20000
15000
10000
5000
2015w1
2016w27
2018w1
2019w26
2021w1
time
Female
Male


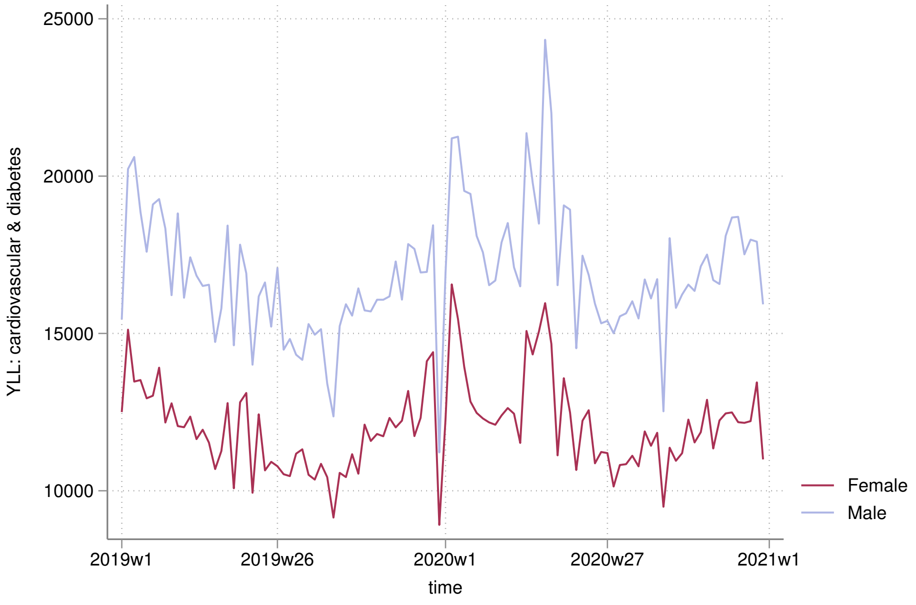

YLL: cardiovascular & diabetes
25000
20000
15000
10000
2019w1
2019w26
2020w1
2020w27
2021w1
time
Female
Male

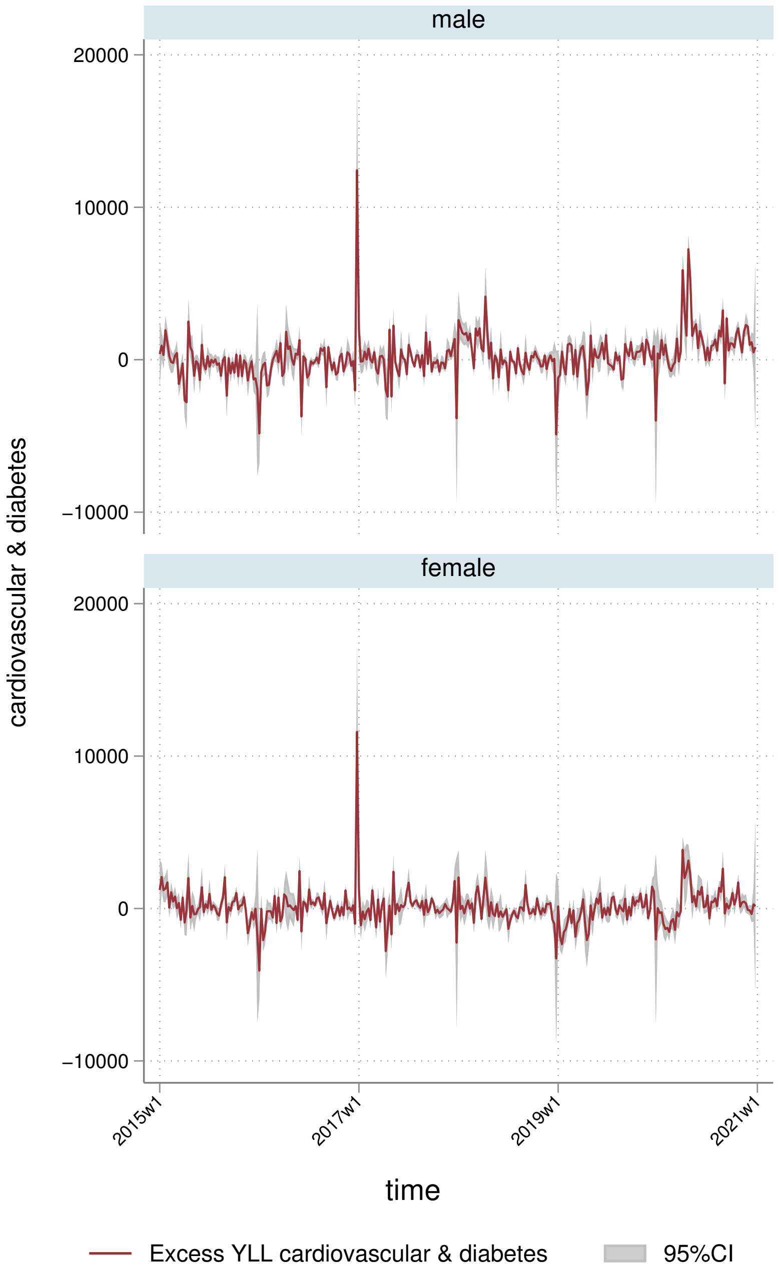
male
female
20000
10000
0
−10000
cardiovascular & diabetes
2015w1
2017w1
2019w1
2021w1
time
Excess YLL cardiovascular & diabetes
95%CI

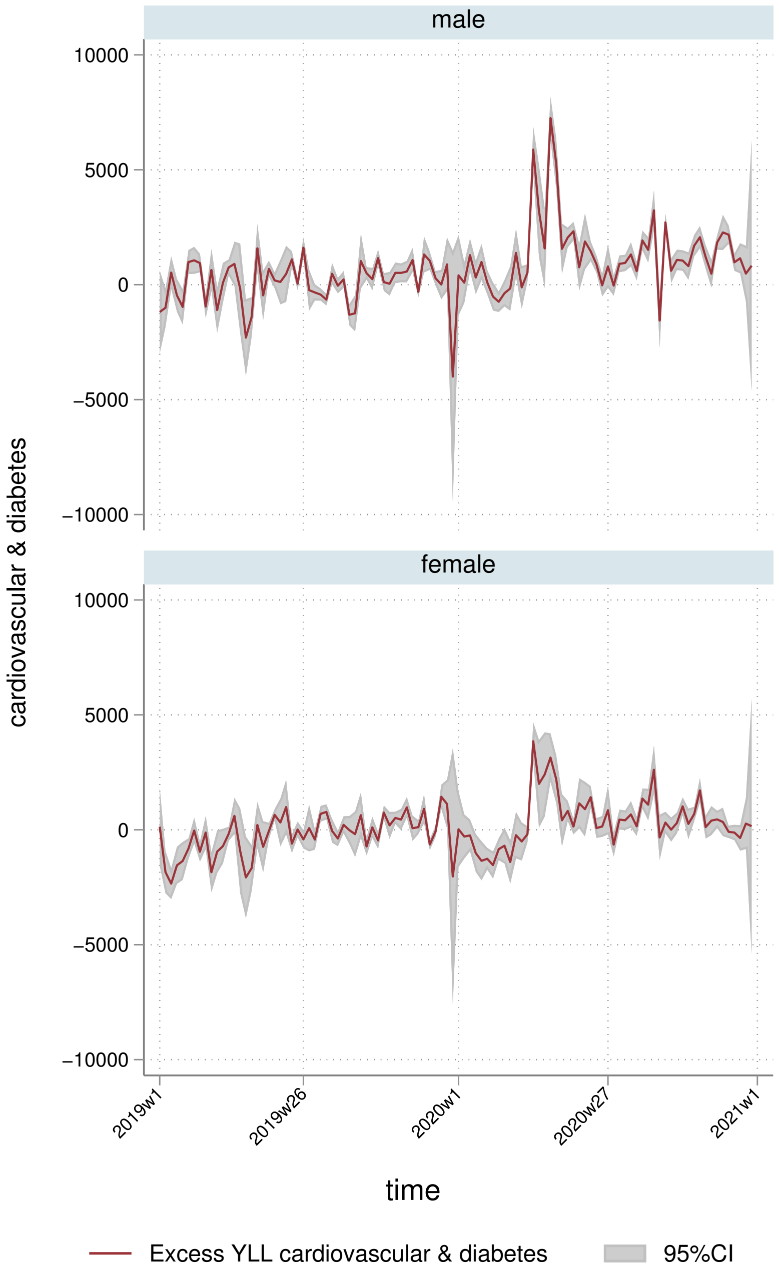
male
female
10000
5000
0
−5000
−10000
cardiovascular & diabetes
2019w1
2019w26
2020w1
2020w27
2021w1
time
Excess YLL cardiovascular & diabetes
95%CI

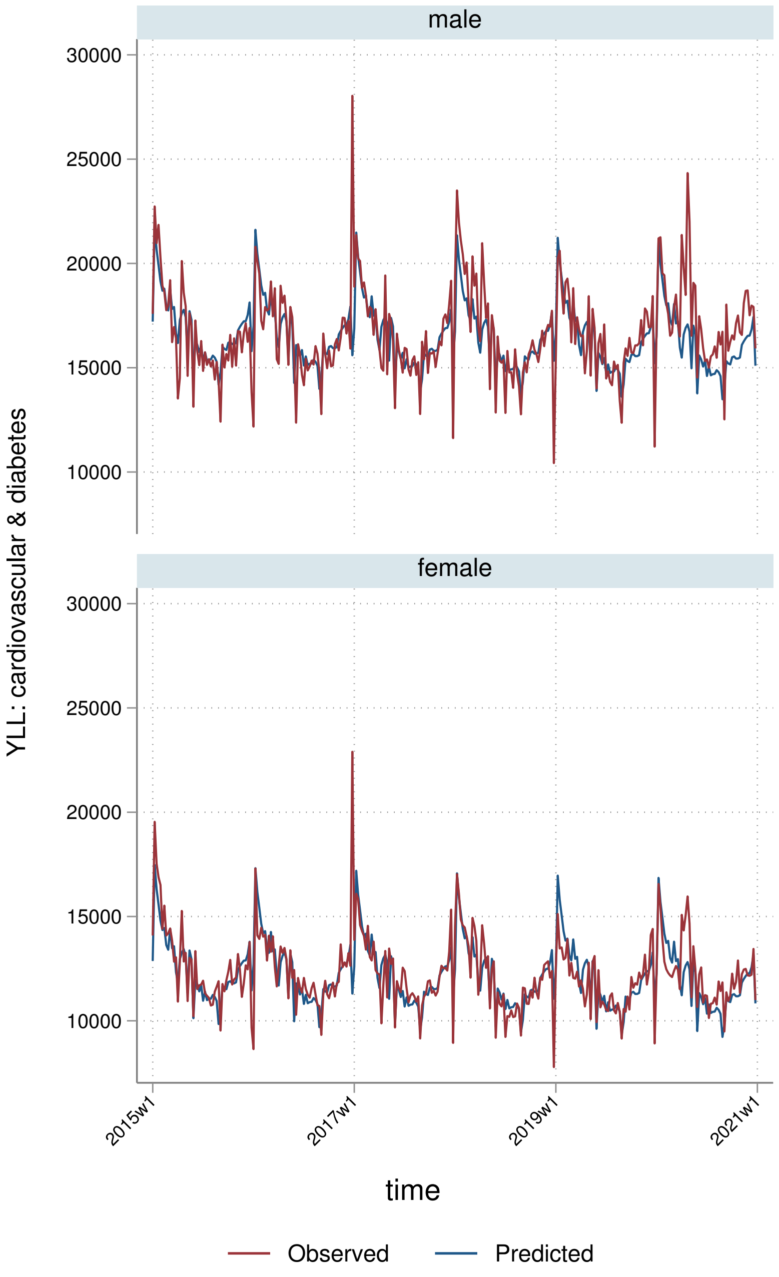
male
female
30000
25000
20000
15000
10000
YLL: cardiovascular & diabetes
2015w1
2017w1
2019w1
2021w1
time
Observed
Predicted

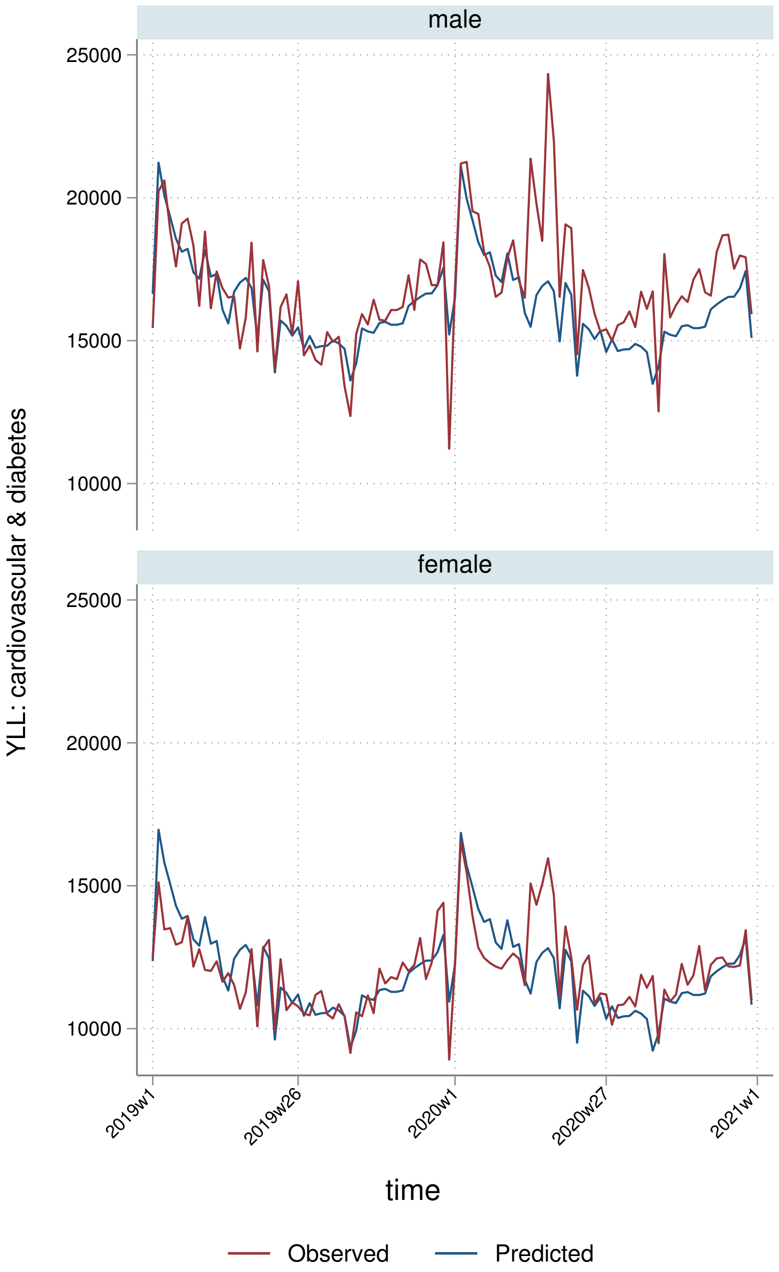
male
female
25000
20000
15000
10000
YLL: cardiovascular & diabetes
2019w1
2019w26
2020w1
2020w27
2021w1
time
Observed
Predicted

##### 7.2.3 By deprivation quintile

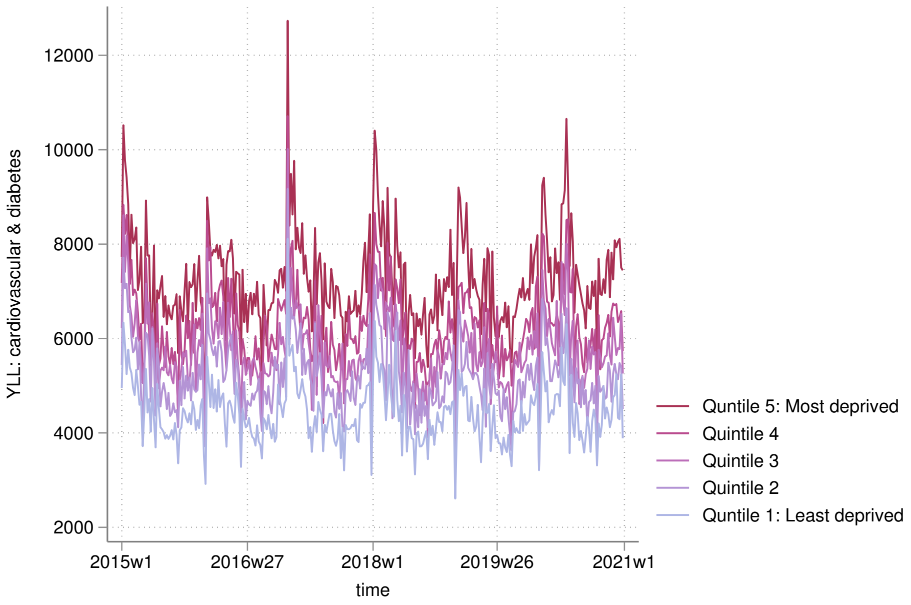


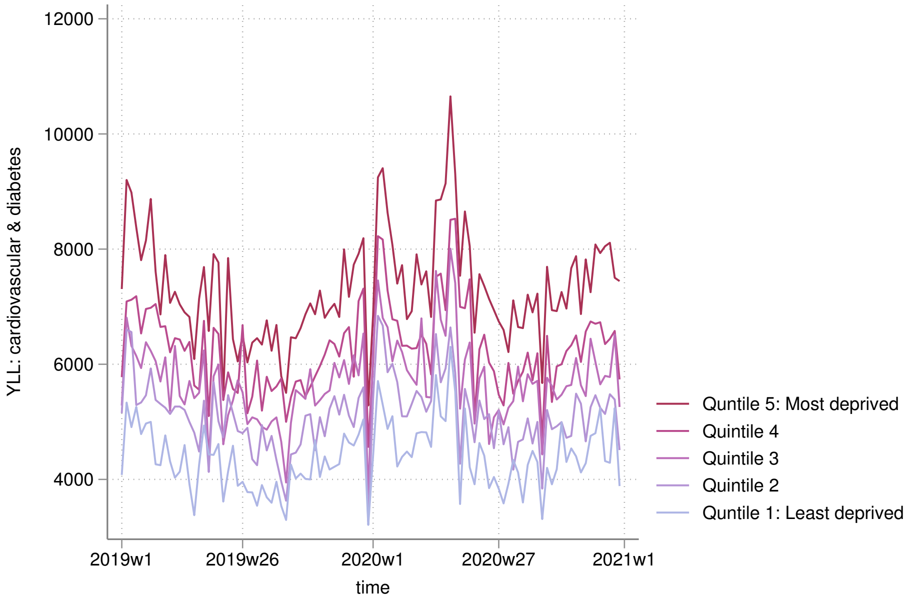

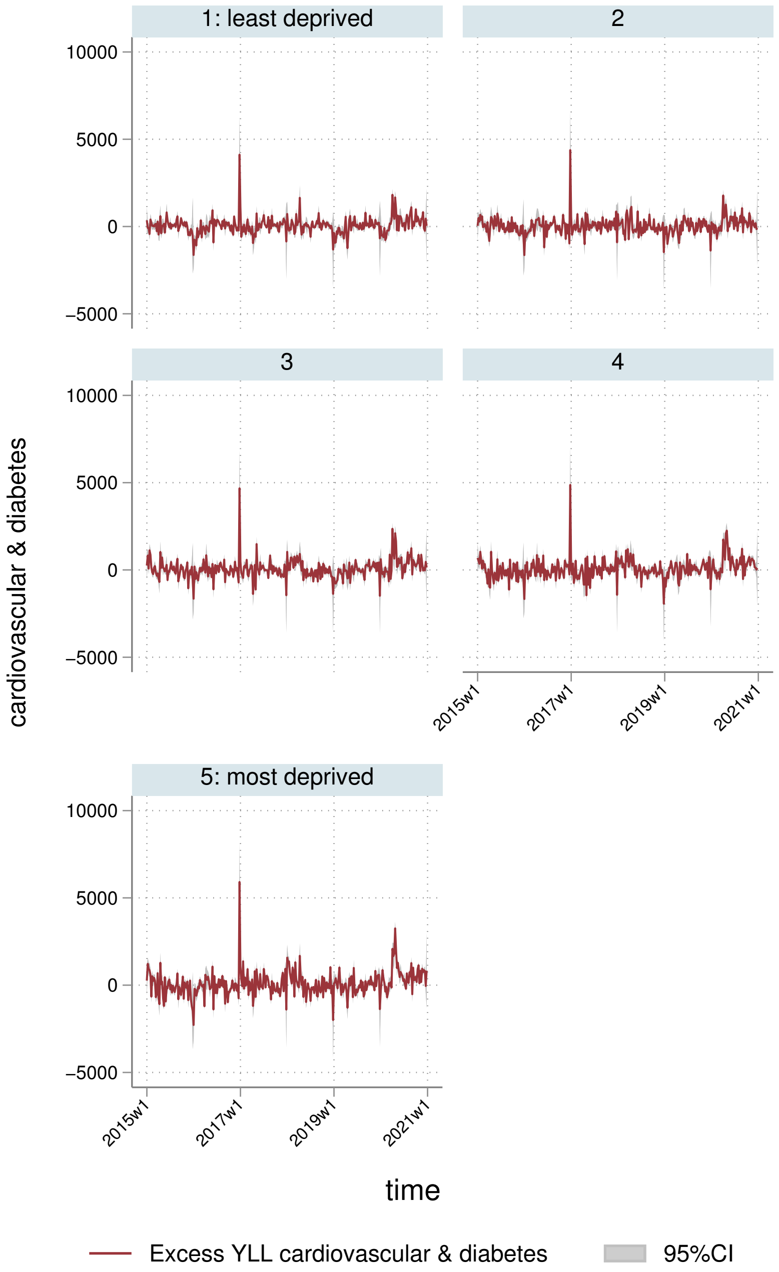
1: least deprived
2
3
4
5: most deprived
10000
5000
0
−5000
cardiovascular & diabetes
2015w1
2017w1
2019w1
2021w1
time
Excess YLL cardiovascular & diabetes
95%CI

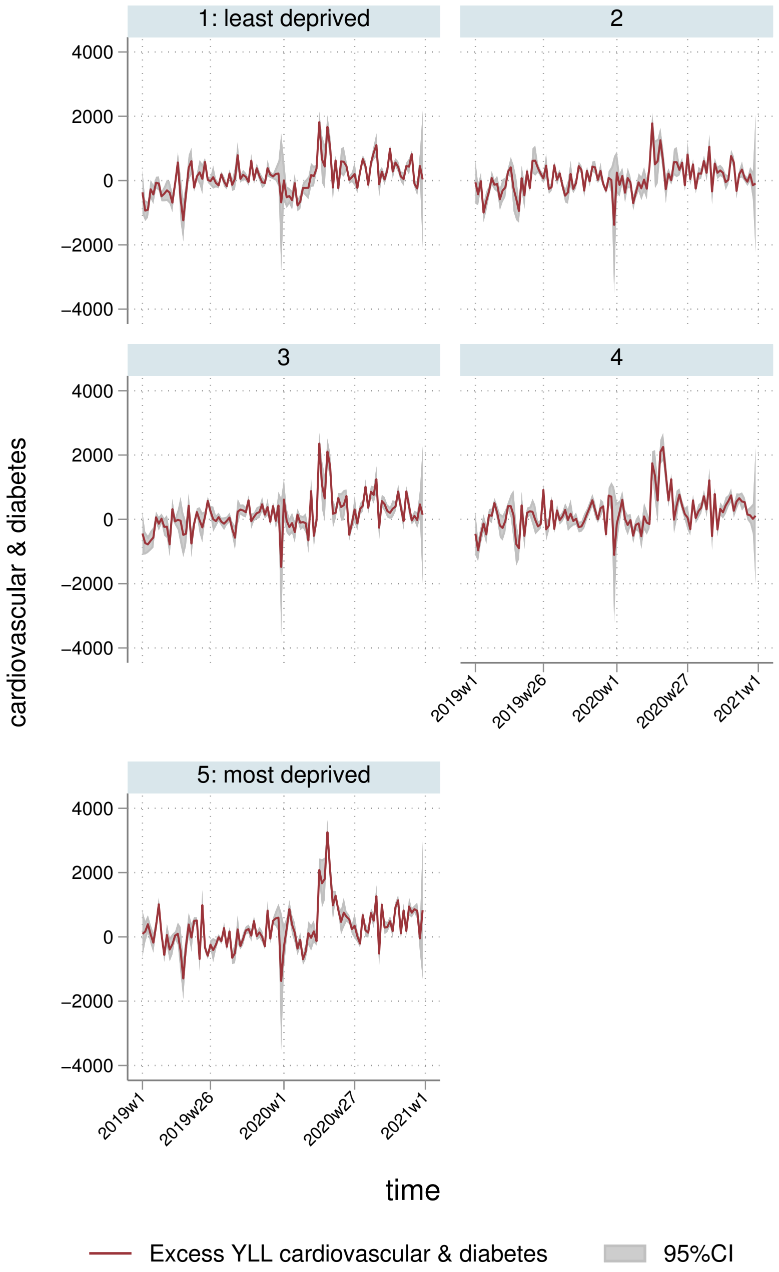
1: least deprived
2
3
4
5: most deprived
cardiovascular & diabetes
4000
2000
0
−2000
−4000
2019w1
2019w26
2020w1
2020w27
2021w1
time
Excess YLL cardiovascular & diabetes
95%CI

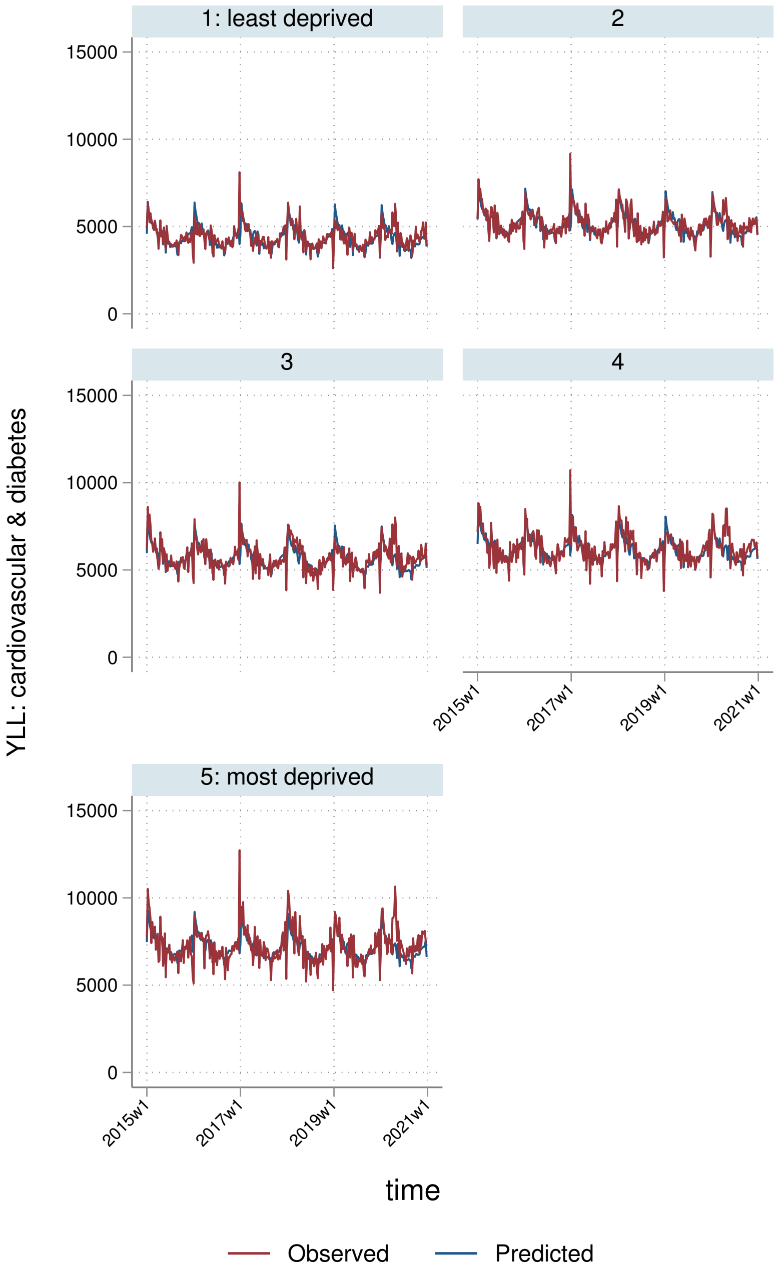

1: least deprived
2
3
4
5: most deprived
15000
10000
5000
0
2015w1
2017w1
2019w1
2021w1
YLL: cardiovascular & diabetes
time
Observed
Predicted

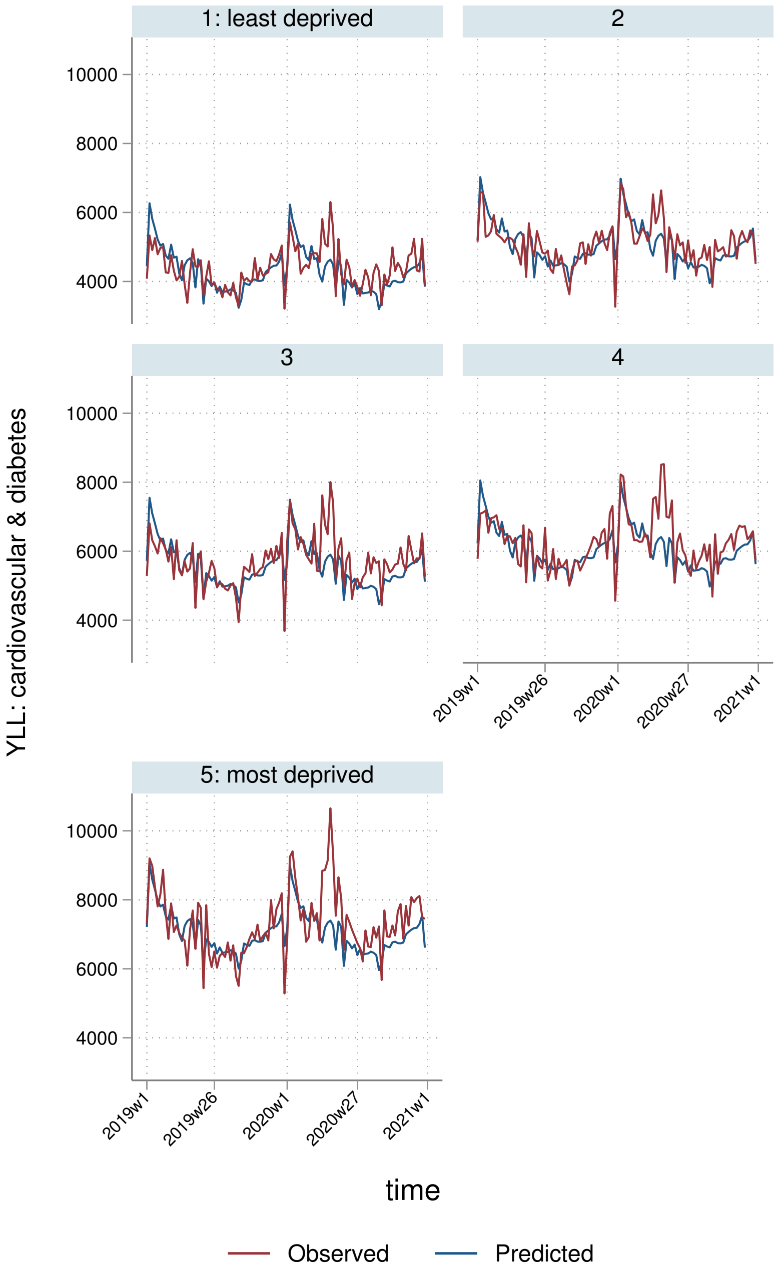
1: least deprived
2
3
4
5: most deprived
YLL: cardiovascular & diabetes
10000
8000
6000
4000
2019w1
2019w26
2020w1
2020w27
2021w1
time
Observed
Predicted

##### 7.2.4 By Strategic Health Authority

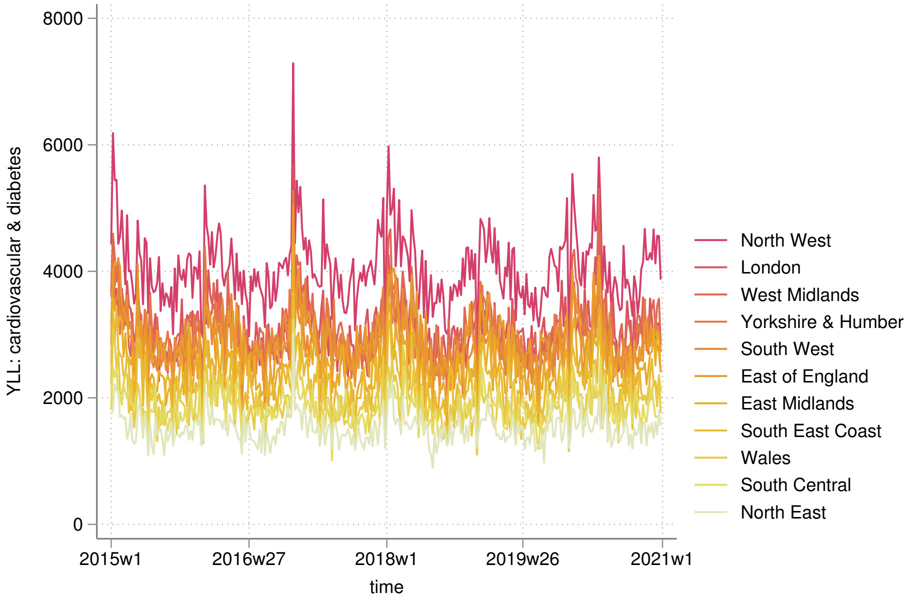


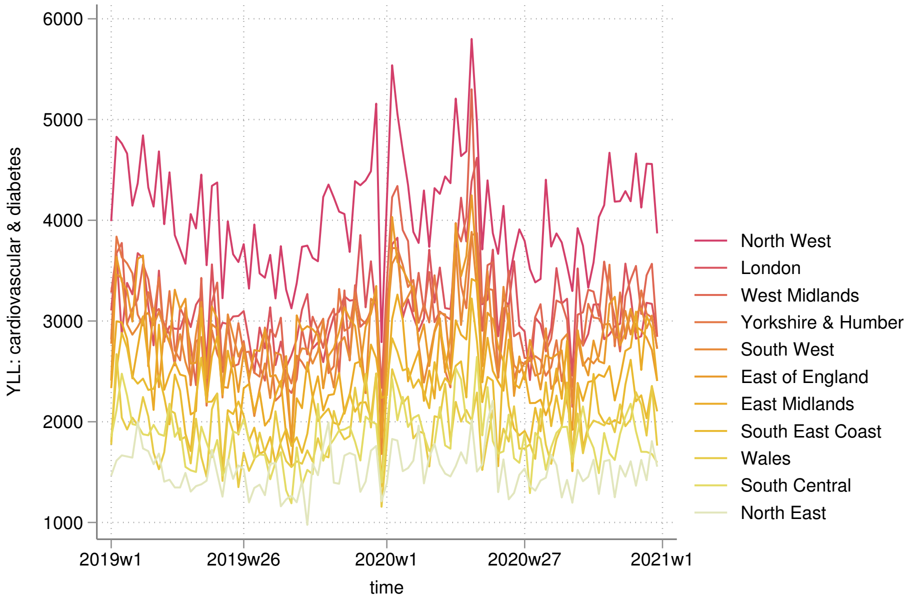

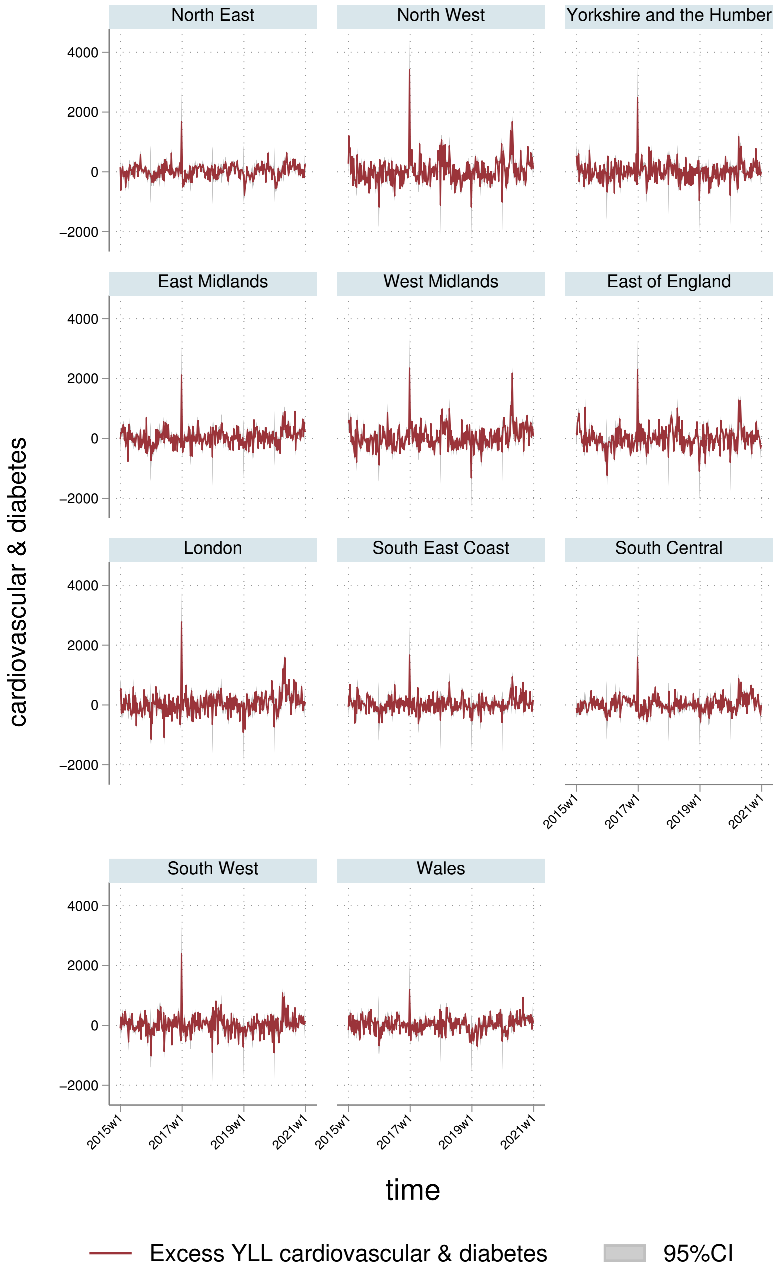
North East
North West
Yorkshire and the Humber
East Midlands
West Midlands
East of England
London
South East Coast
South Central
South West
Wales
4000
2000
0
−2000
cardiovascular & diabetes
2015w1
2017w1
2019w1
2021w1
time
Excess YLL cardiovascular & diabetes
95%CI

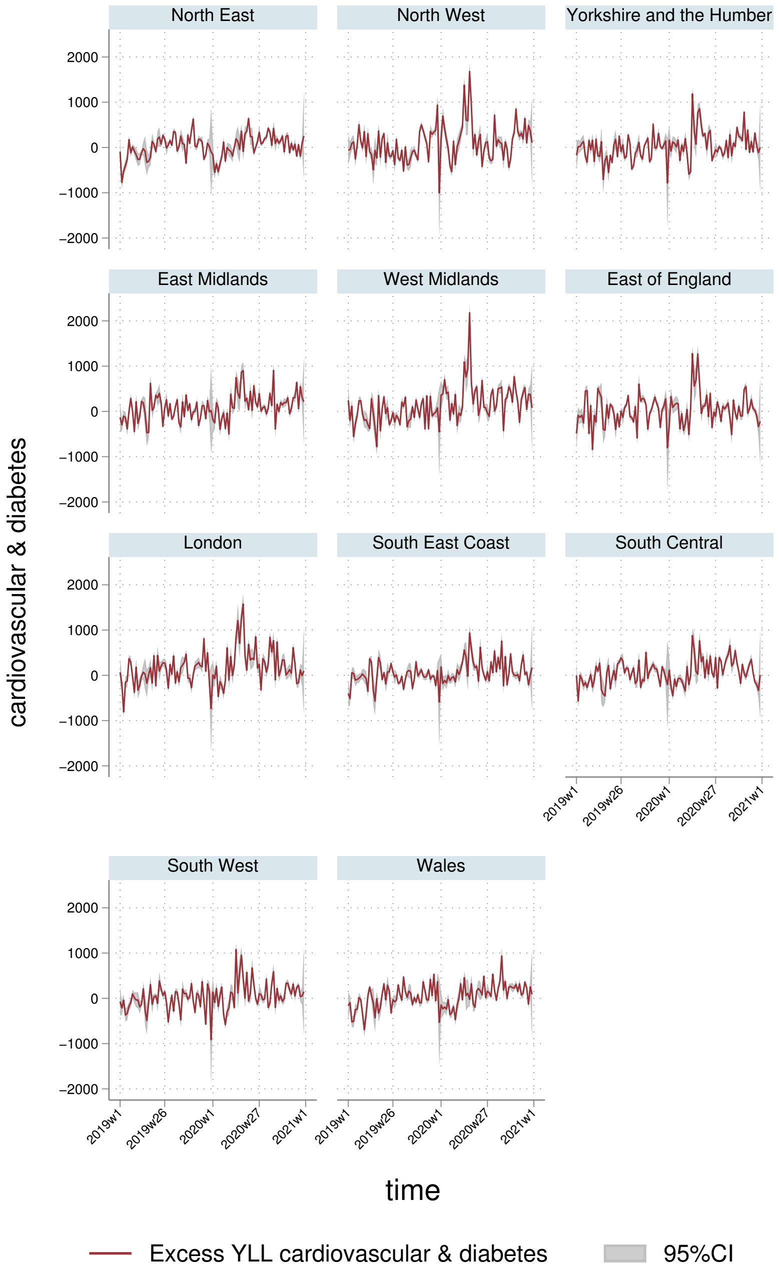
North East
North West
Yorkshire and the Humber
East Midlands
West Midlands
East of England
London
South East Coast
South Central
South West
Wales
2000
1000
0
−1000
−2000
2019w1
2019w26
2020w1
2020w27
2021w1
cardiovascular & diabetes
time
Excess YLL cardiovascular & diabetes
95%CI

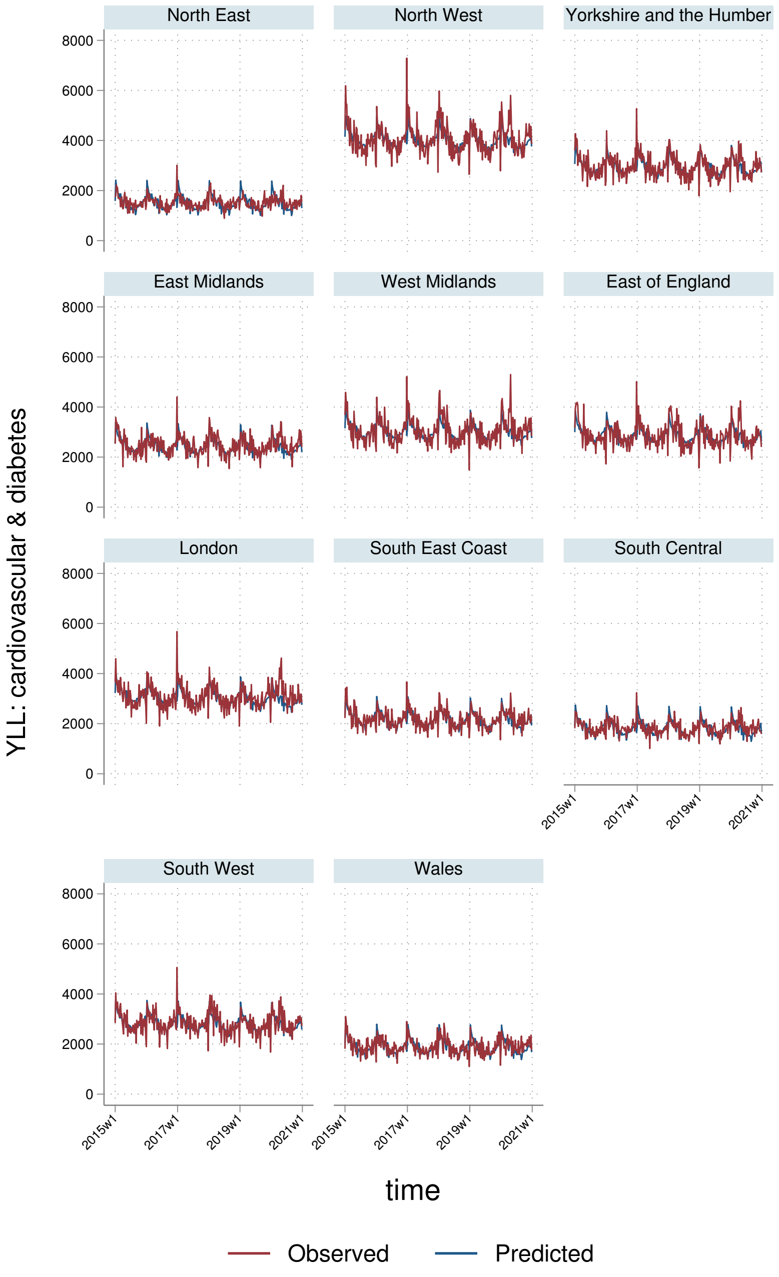
North East
North West
Yorkshire and the Humber
East Midlands
West Midlands
East of England
London
South East Coast
South Central
South West
Wales
YLL: cardiovascular & diabetes
0
2000
4000
6000
8000
2015w1
2017w1
2019w1
2021w1
time
Observed
Predicted

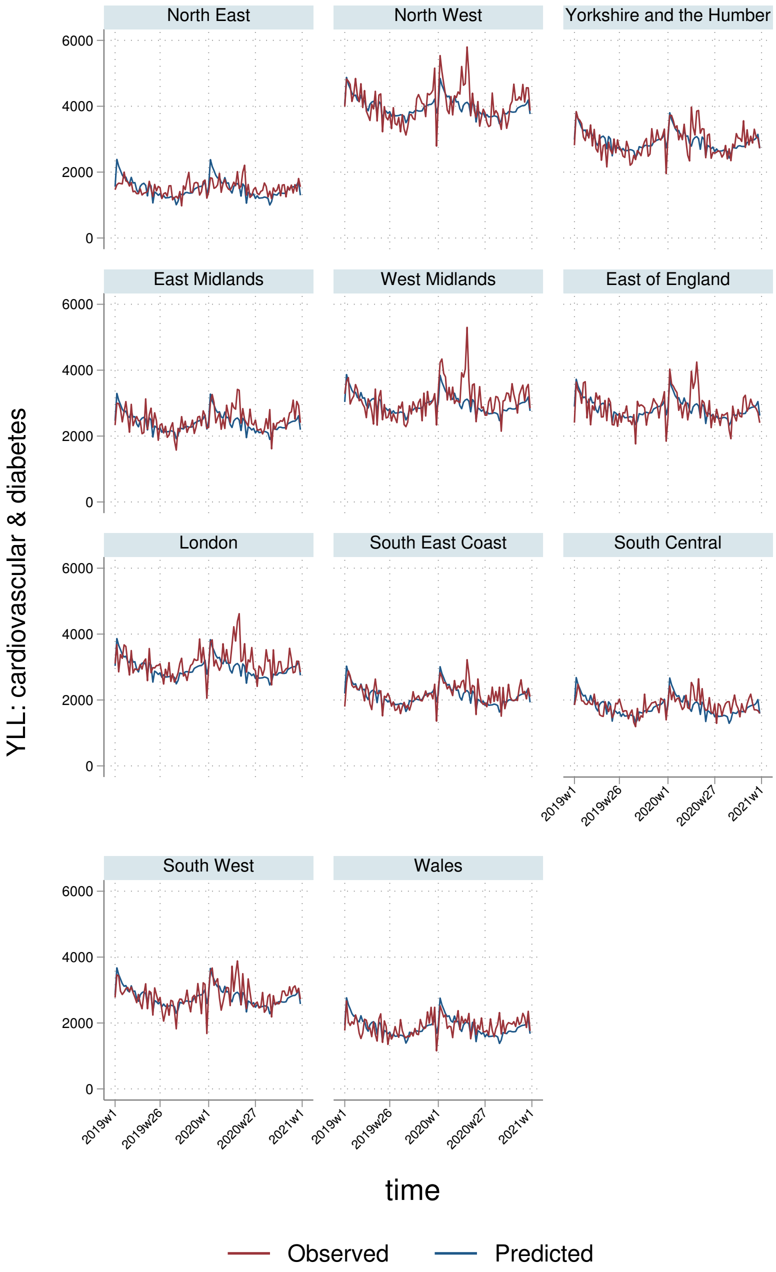
North East
North West
Yorkshire and the Humber
East Midlands
West Midlands
East of England
London
South East Coast
South Central
South West
Wales
YLL: cardiovascular & diabetes
0
2000
4000
6000
2019w1
2019w26
2020w1
2020w27
2021w1
time
Observed
Predicted

#### 7.3 YLLs per $100,000$ population

##### 7.3.1 England-Wales aggregate

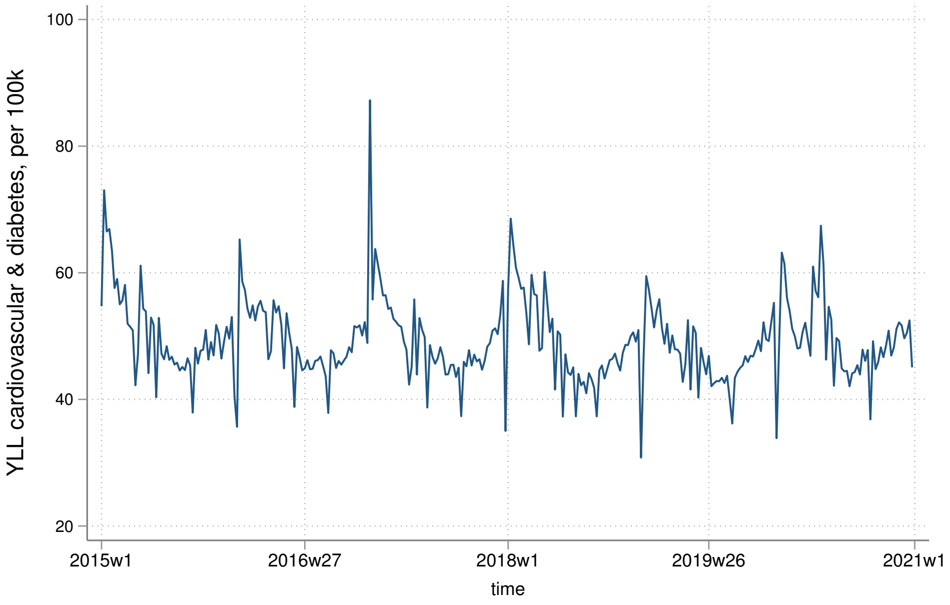


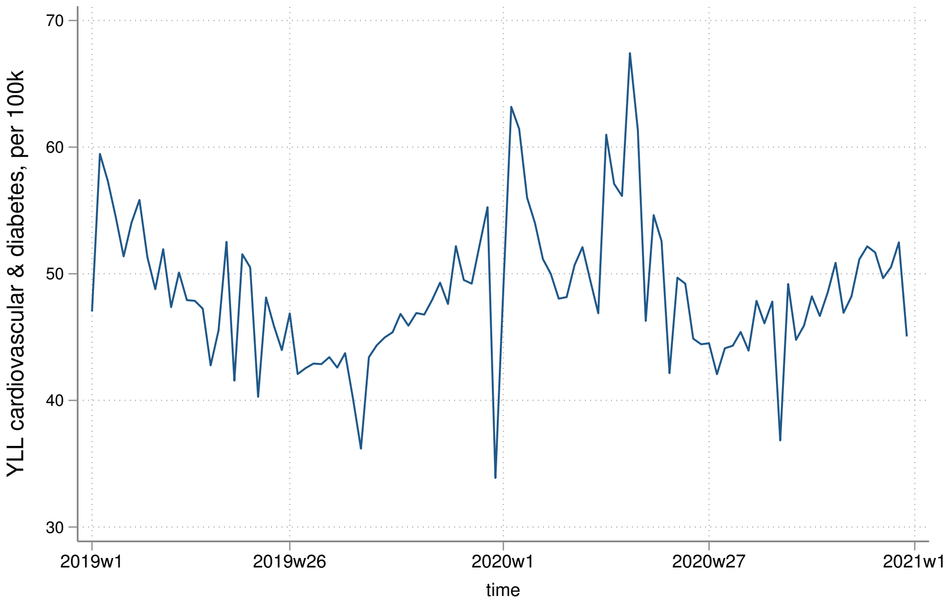

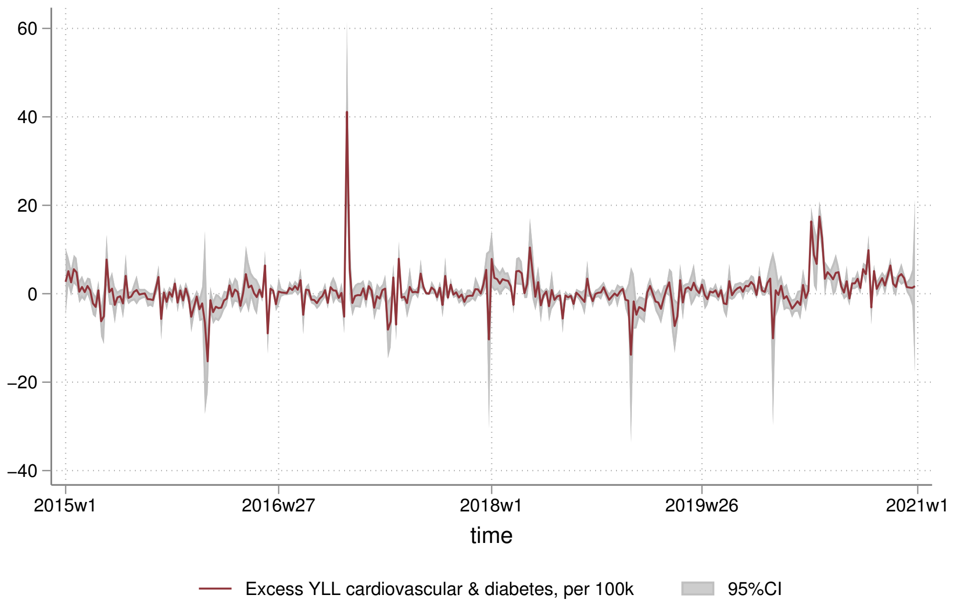
60
40
20
0
−20
−40
2015w1
2016w27
2018w1
2019w26
2021w1
time
Excess YLL cardiovascular & diabetes, per 100k
95%CI

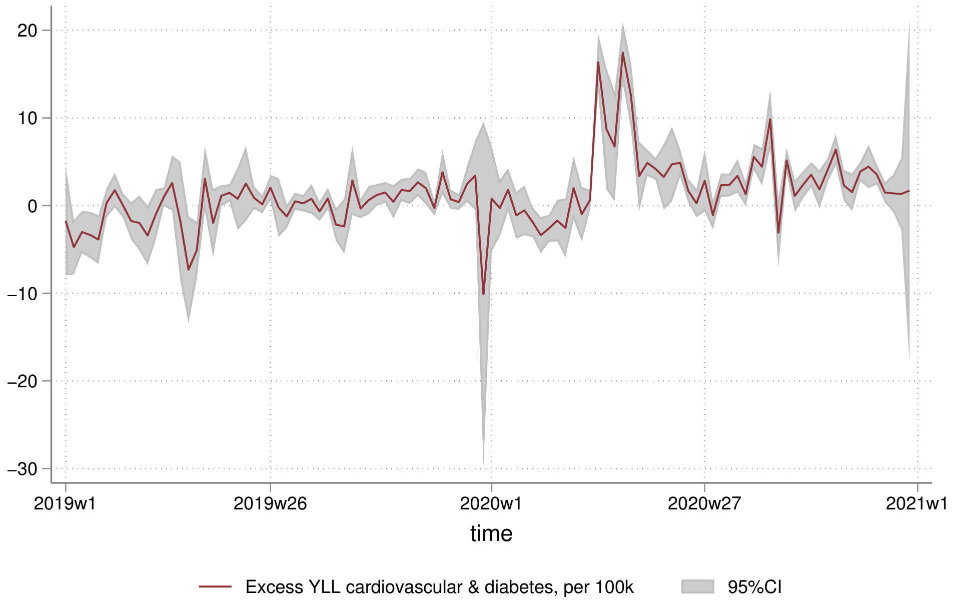
20
10
0
−10
−20
−30
2019w1
2019w26
2020w1
2020w27
2021w1
time
Excess YLL cardiovascular & diabetes, per 100k
95%CI

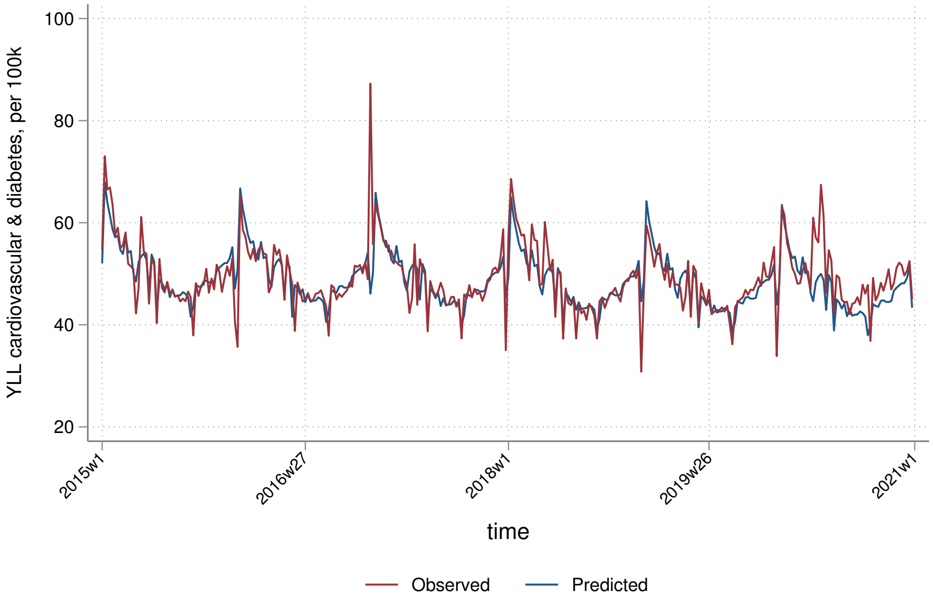

100
80
60
40
20
YLL cardiovascular & diabetes, per 100k
2015w1
2016w27
2018w1
2019w26
2021w1
time
Observed
Predicted


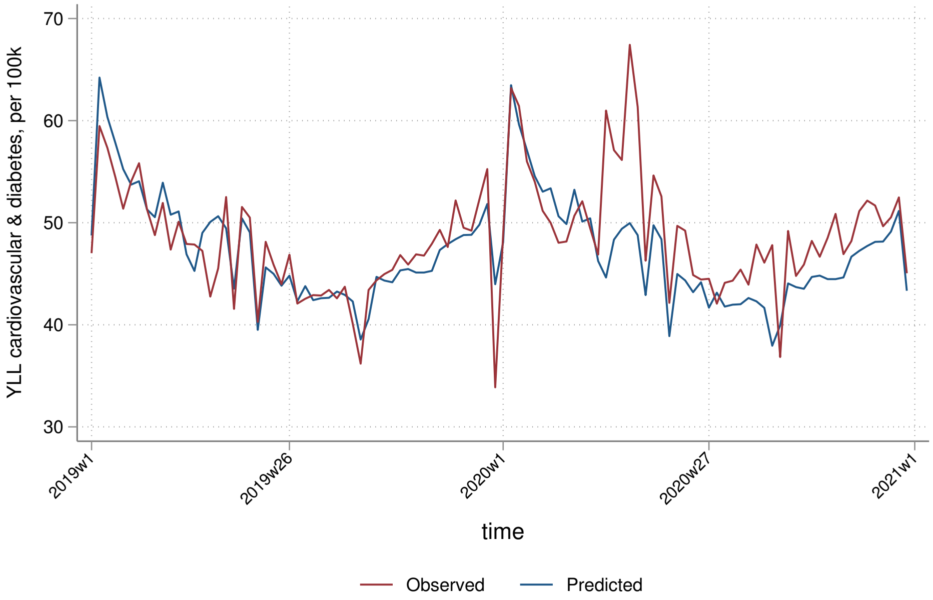

70
60
50
40
30
YLL cardiovascular & diabetes, per 100k
2019w1
2019w26
2020w1
2020w27
2021w1
time
Observed
Predicted

##### 7.3.2 By sex

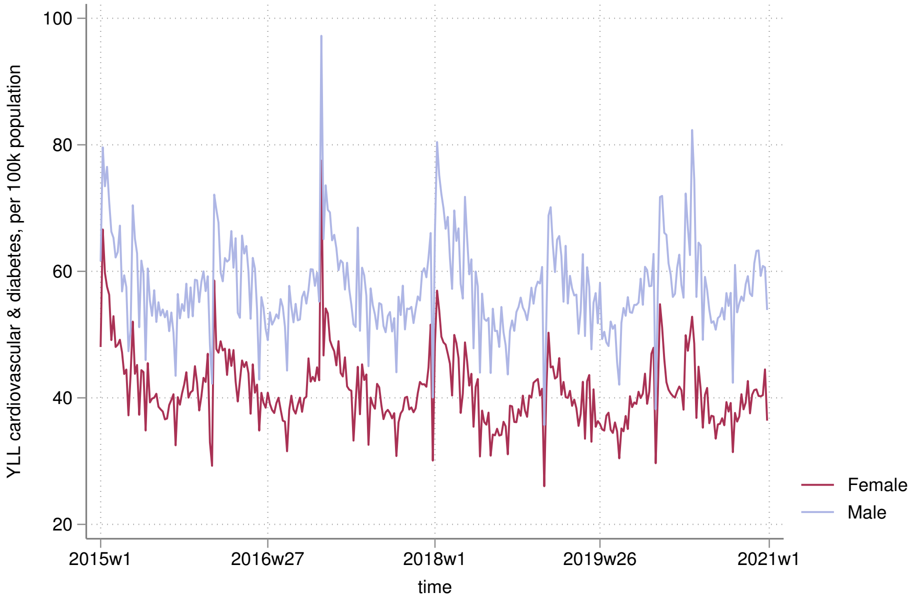

YLL cardiovascular & diabetes, per 100k population
100
80
60
40
20
2015w1
2016w27
2018w1
2019w26
2021w1
time
Female
Male


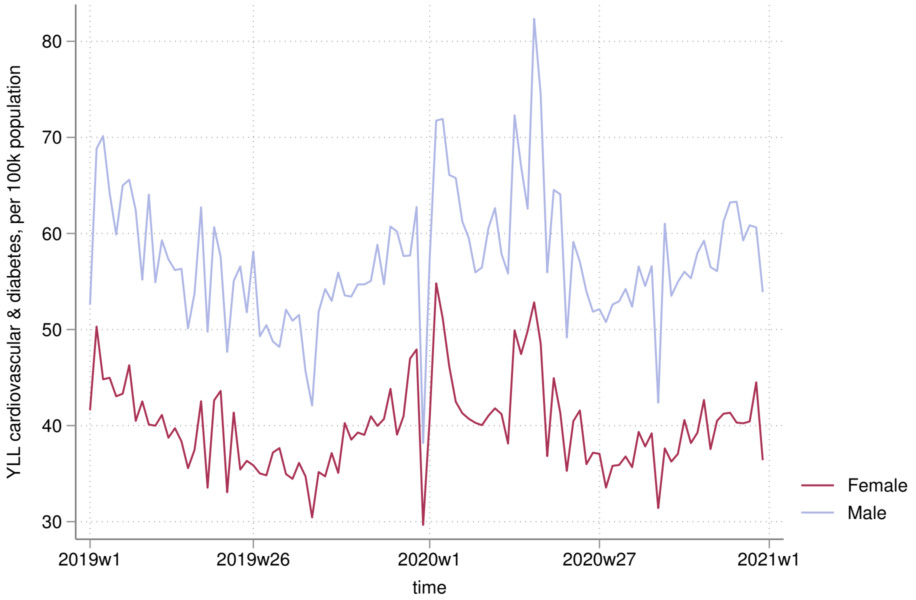

YLL cardiovascular & diabetes, per 100k population
80
70
60
50
40
30
2019w1
2019w26
2020w1
2020w27
2021w1
time
Female
Male

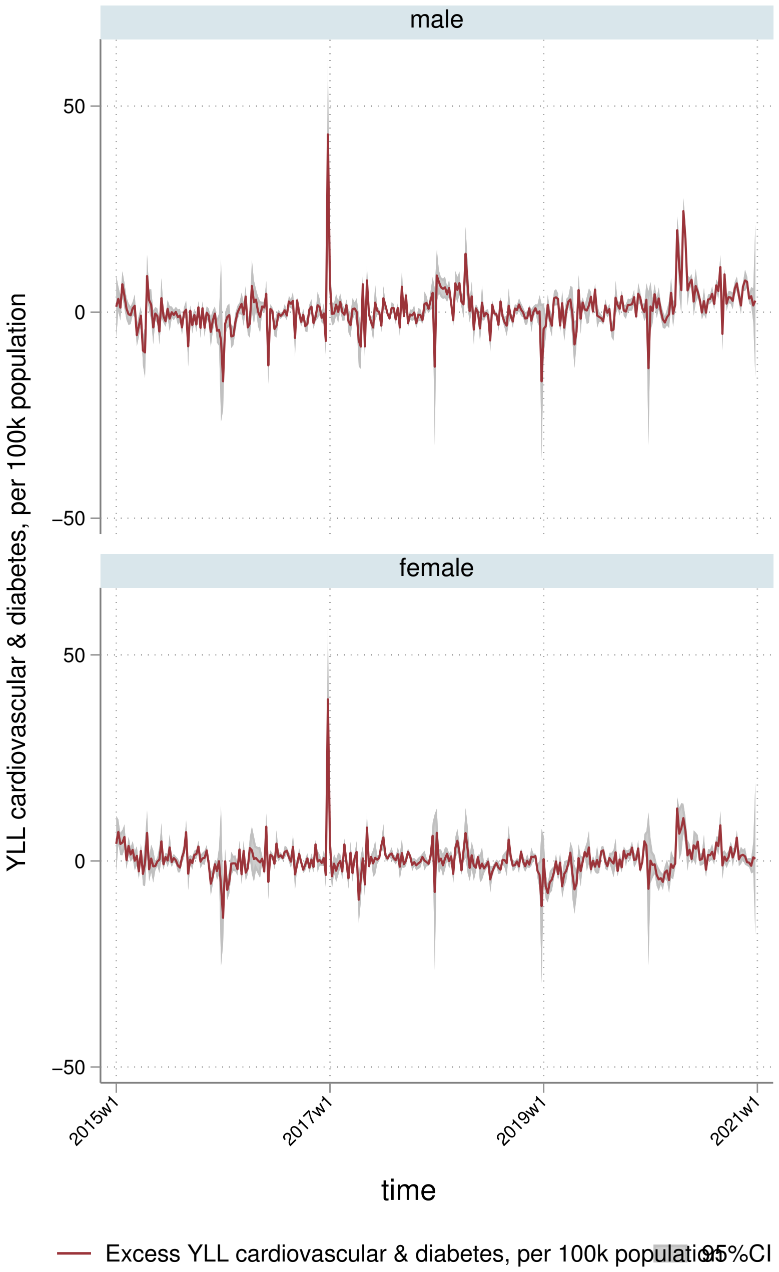
male
female
50
0
−50
YLL cardiovascular & diabetes, per 100k population
2015w1
2017w1
2019w1
2021w1
time
Excess YLL cardiovascular & diabetes, per 100k population
95%CI

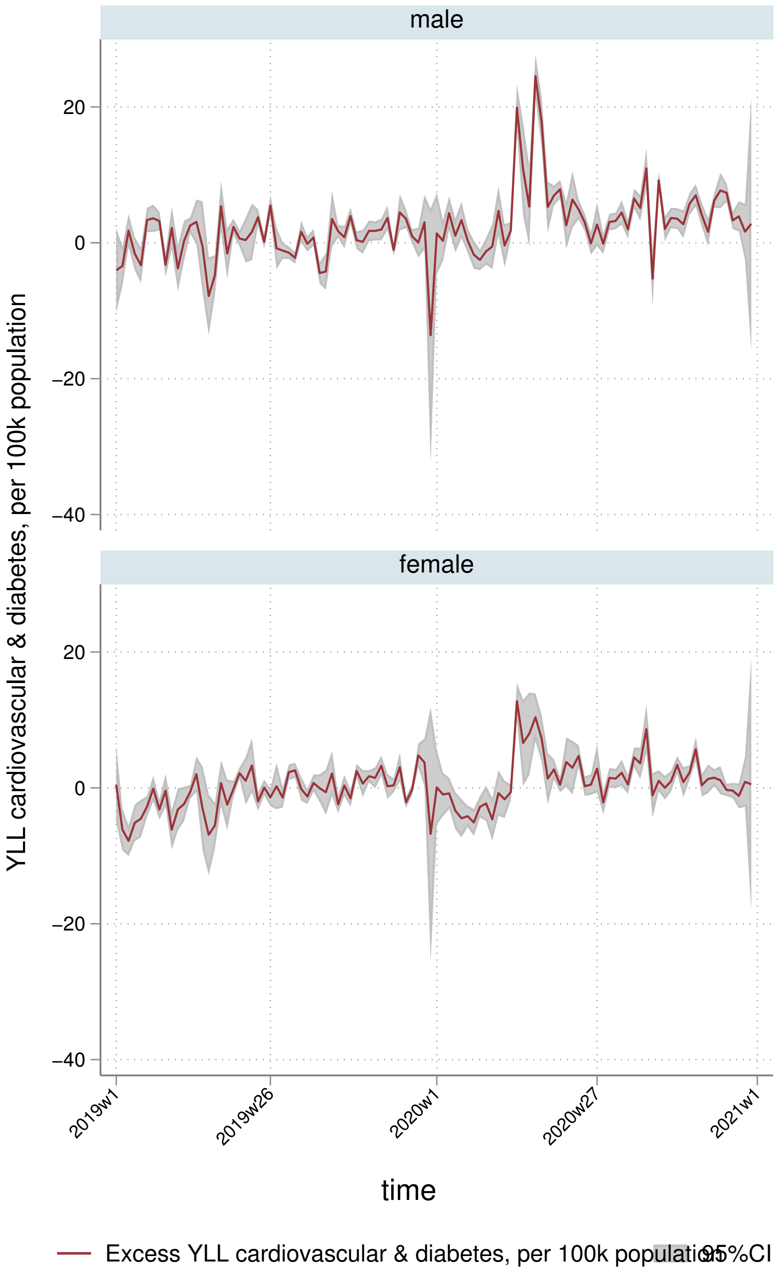
male
20
0
−20
−40
female
20
0
−20
−40
YLL cardiovascular & diabetes, per 100k population
2019w1
2019w26
2020w1
2020w27
2021w1
time
Excess YLL cardiovascular & diabetes, per 100k population
95%CI

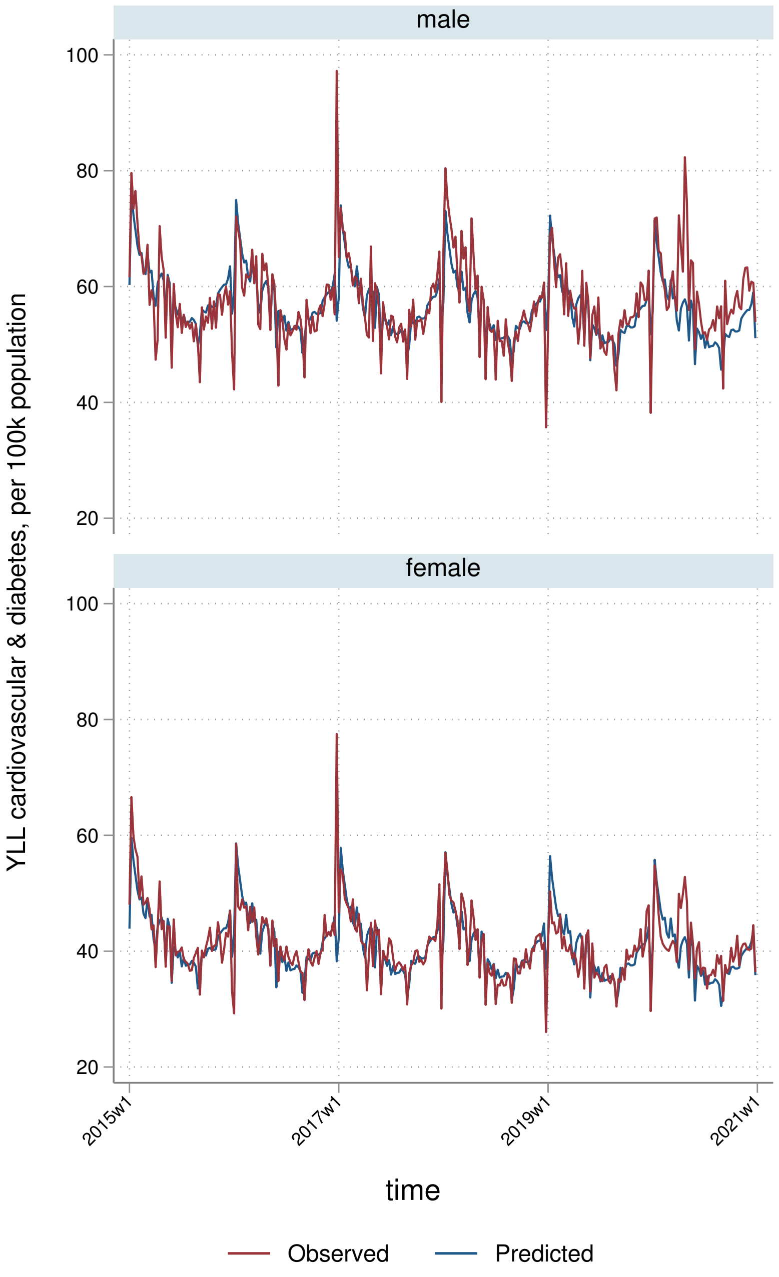
male
female
YLL cardiovascular & diabetes, per 100k population
100
80
60
40
20
2015w1
2017w1
2019w1
2021w1
time
Observed
Predicted

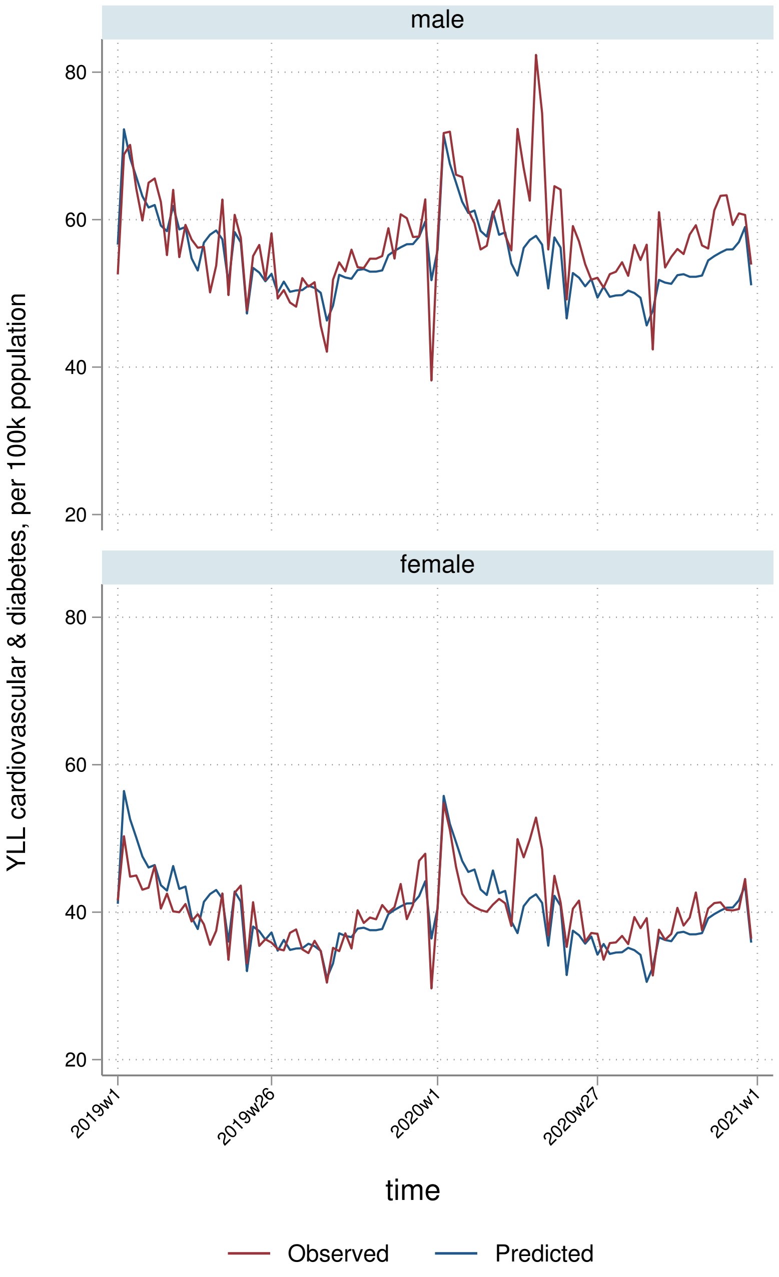
male
female
80
60
40
20
YLL cardiovascular & diabetes, per 100k population
2019w1
2019w26
2020w1
2020w27
2021w1
time
Observed
Predicted

##### 7.3.3 By deprivation quintile

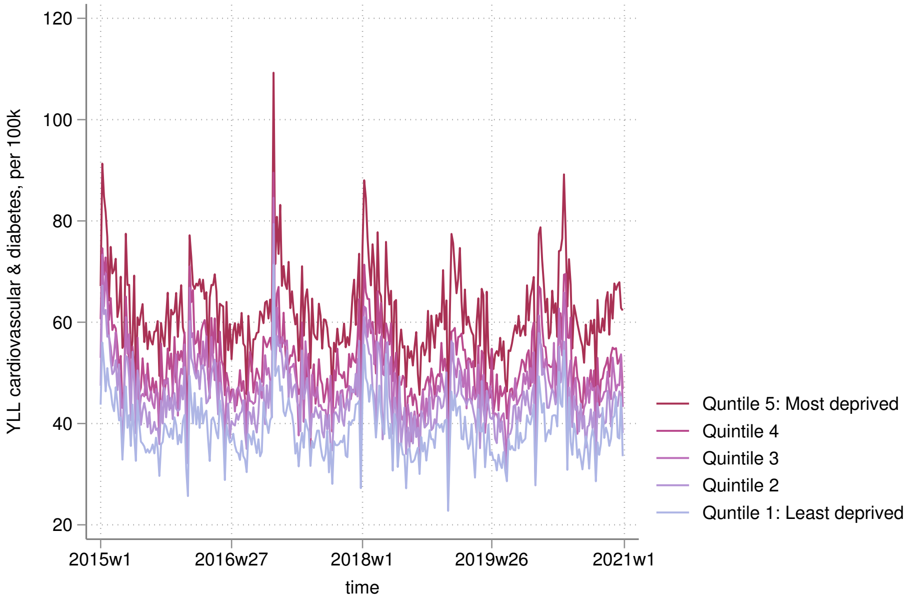


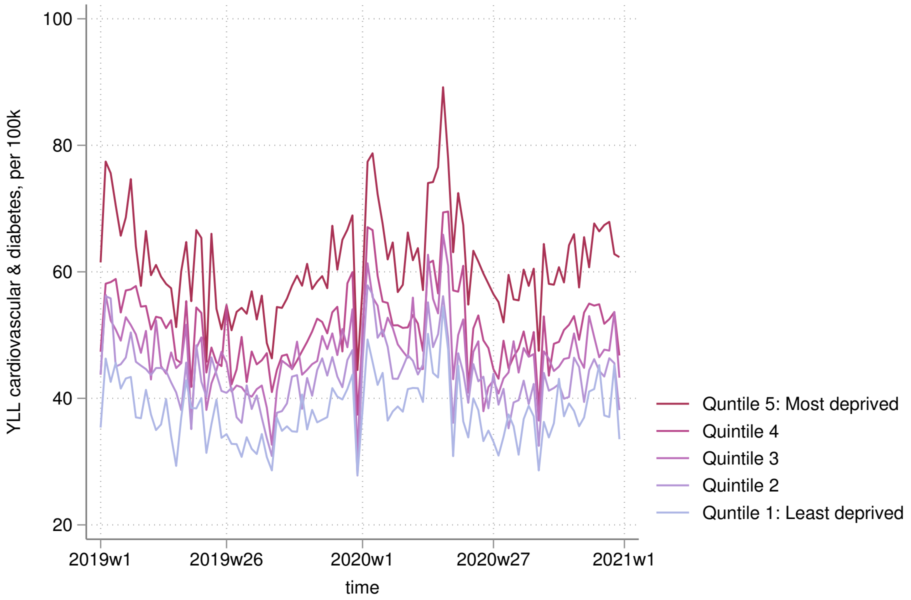

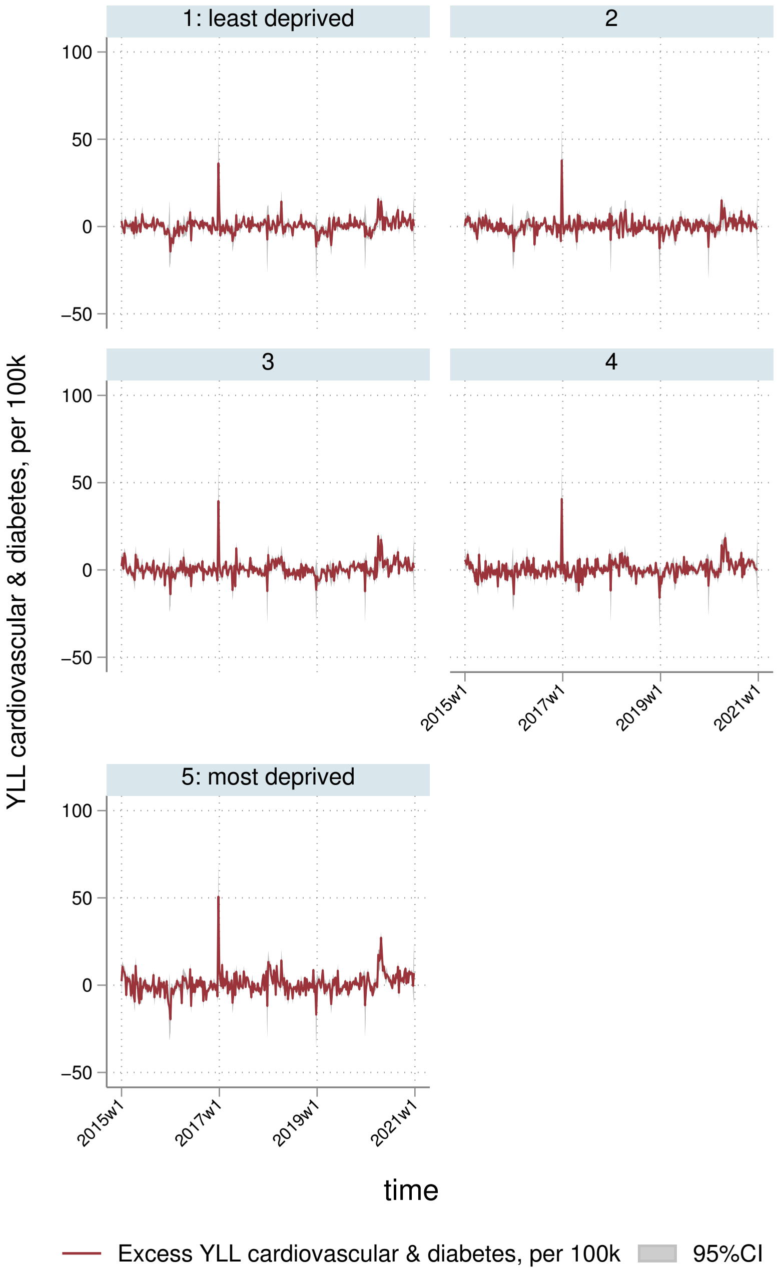

1: least deprived
2
3
4
5: most deprived
100
50
0
−50
2015w1
2017w1
2019w1
2021w1
YLL cardiovascular & diabetes, per 100k
time
Excess YLL cardiovascular & diabetes, per 100k
95%CI

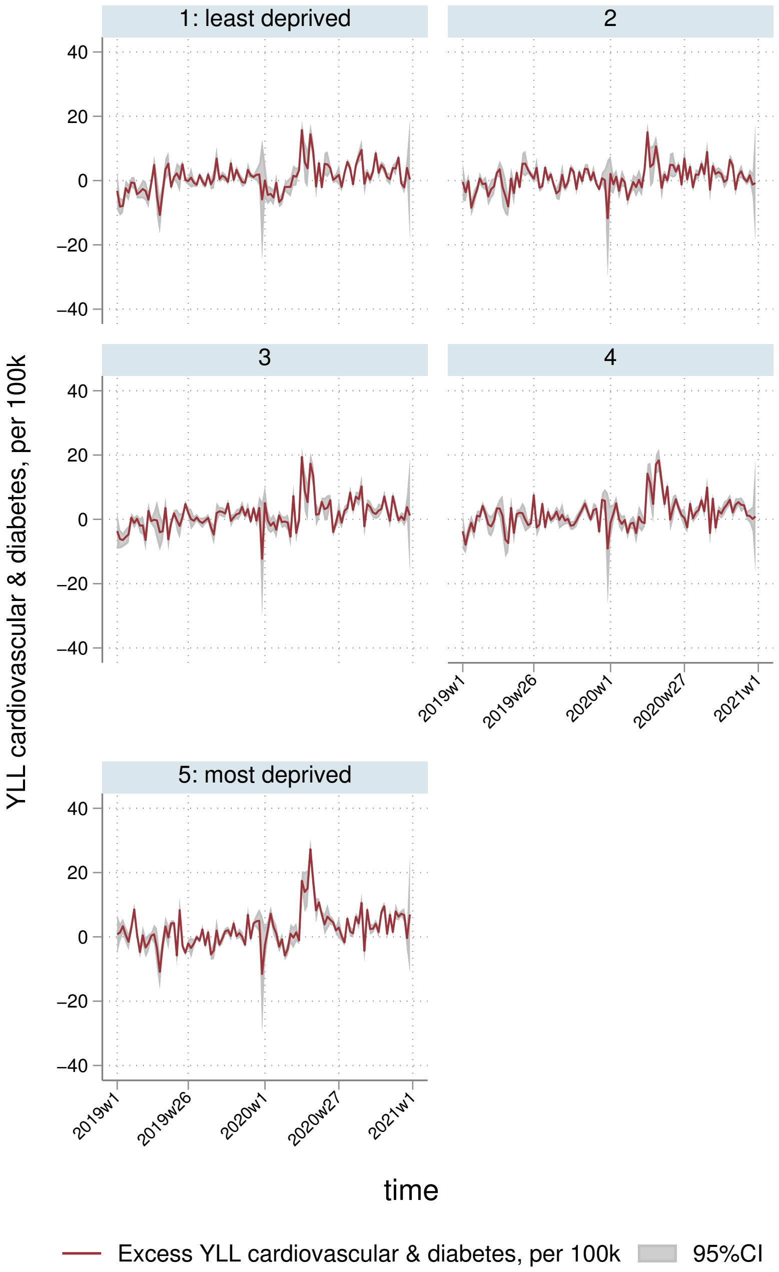
1: least deprived
2
3
4
5: most deprived
YLL cardiovascular & diabetes, per 100k
40
20
0
−20
−40
2019w1
2019w26
2020w1
2020w27
2021w1
time
Excess YLL cardiovascular & diabetes, per 100k
95%CI

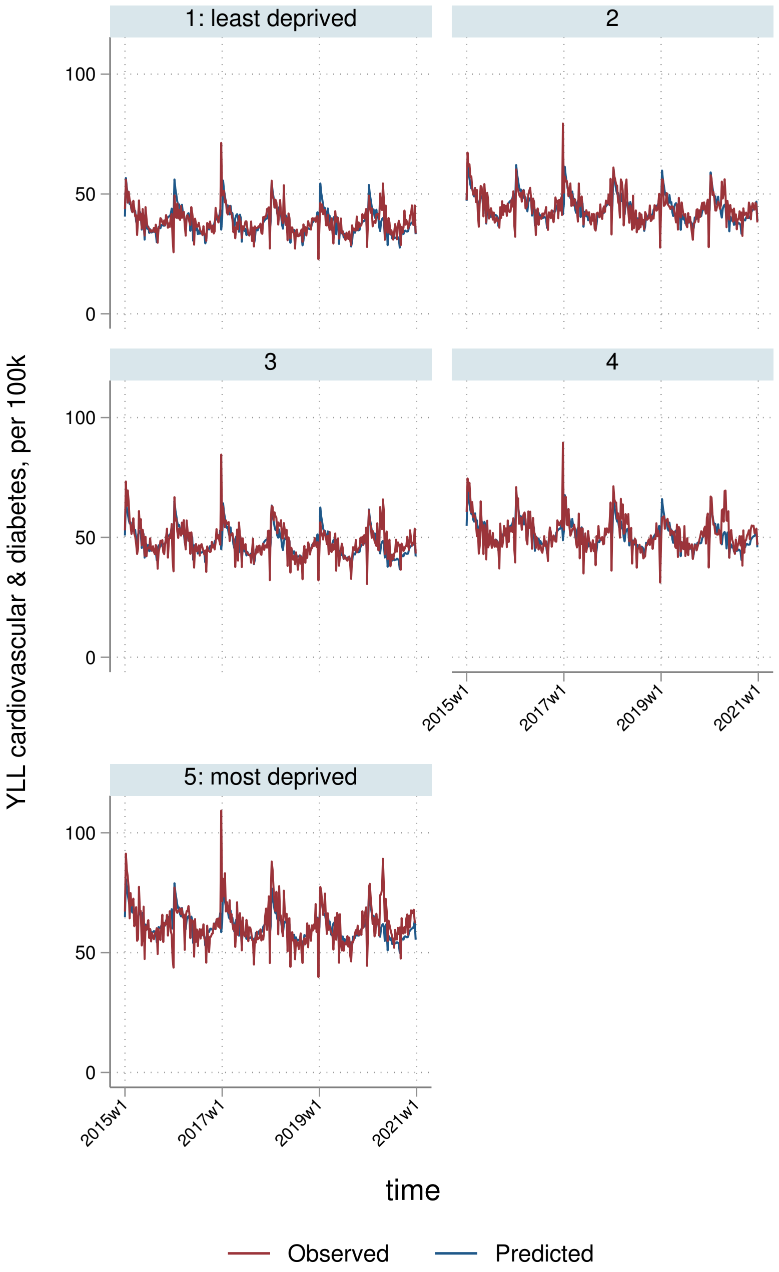
1: least deprived
2
3
4
5: most deprived
100
50
0
2015w1
2017w1
2019w1
2021w1
YLL cardiovascular & diabetes, per 100k
time
Observed
Predicted

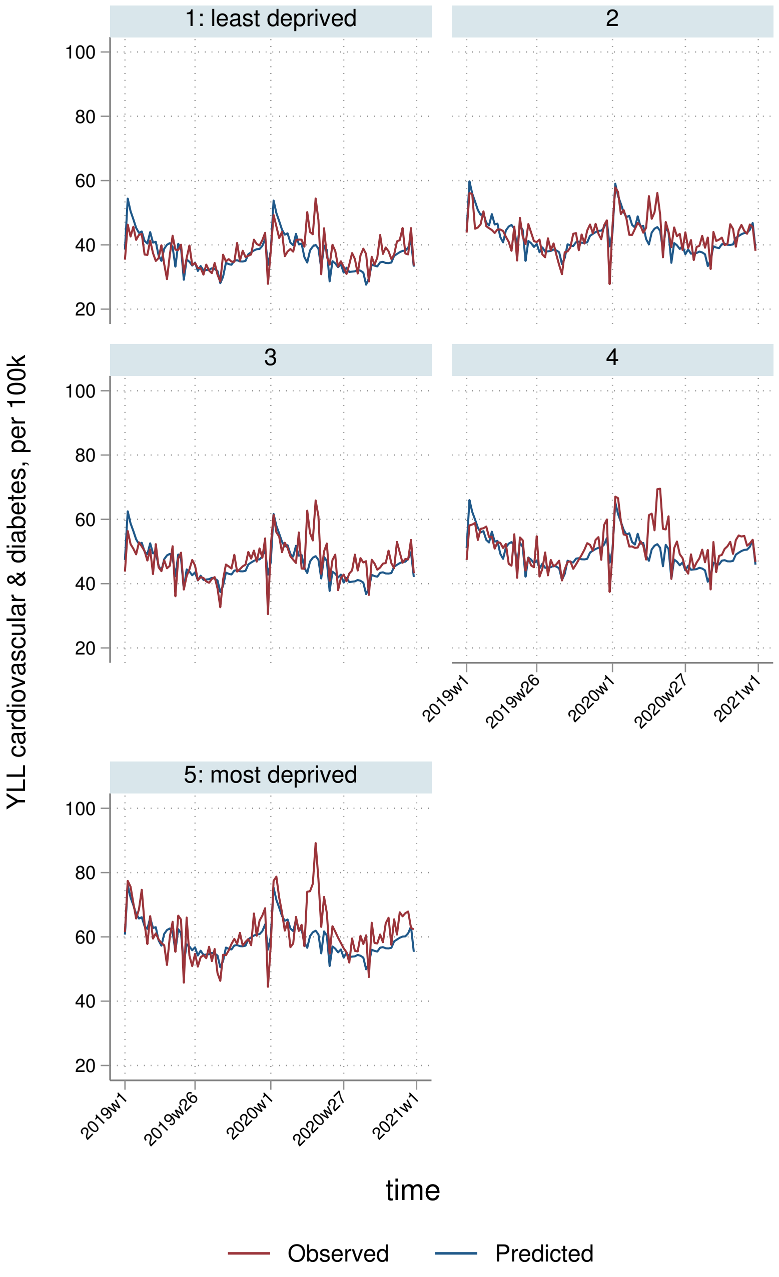
1: least deprived
2
3
4
5: most deprived
YLL cardiovascular & diabetes, per 100k
100
80
60
40
20
2019w1
2019w26
2020w1
2020w27
2021w1
time
Observed
Predicted

##### 7.3.4 By Strategic Health Authority

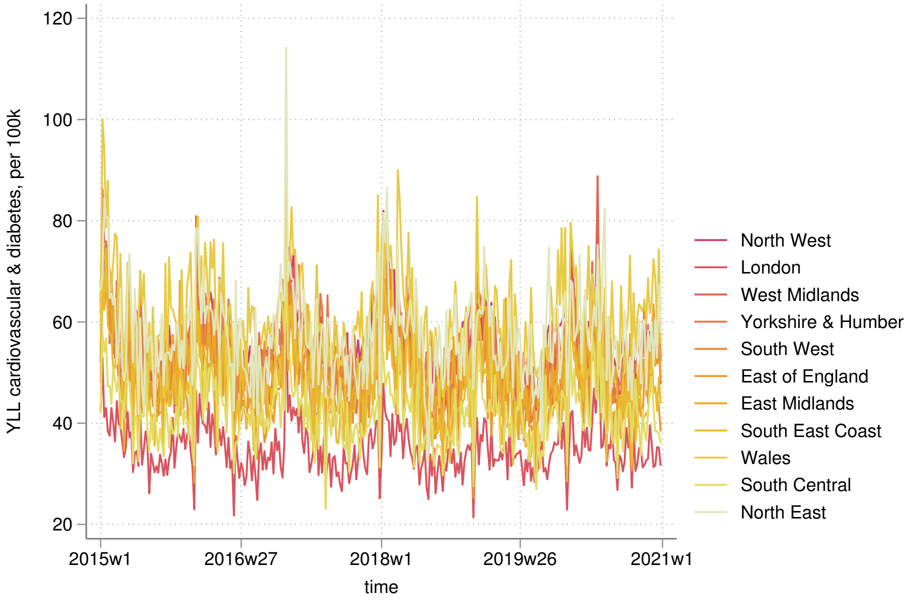


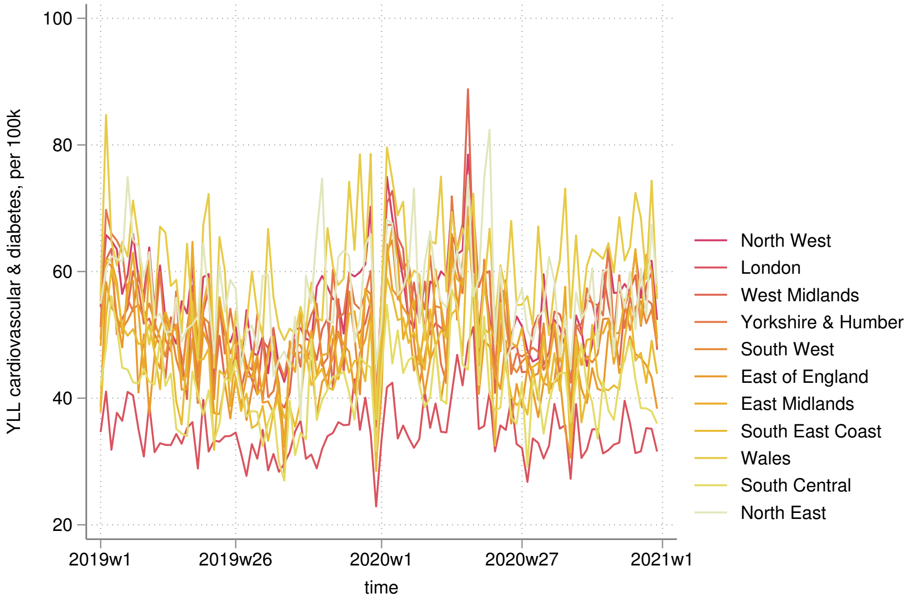

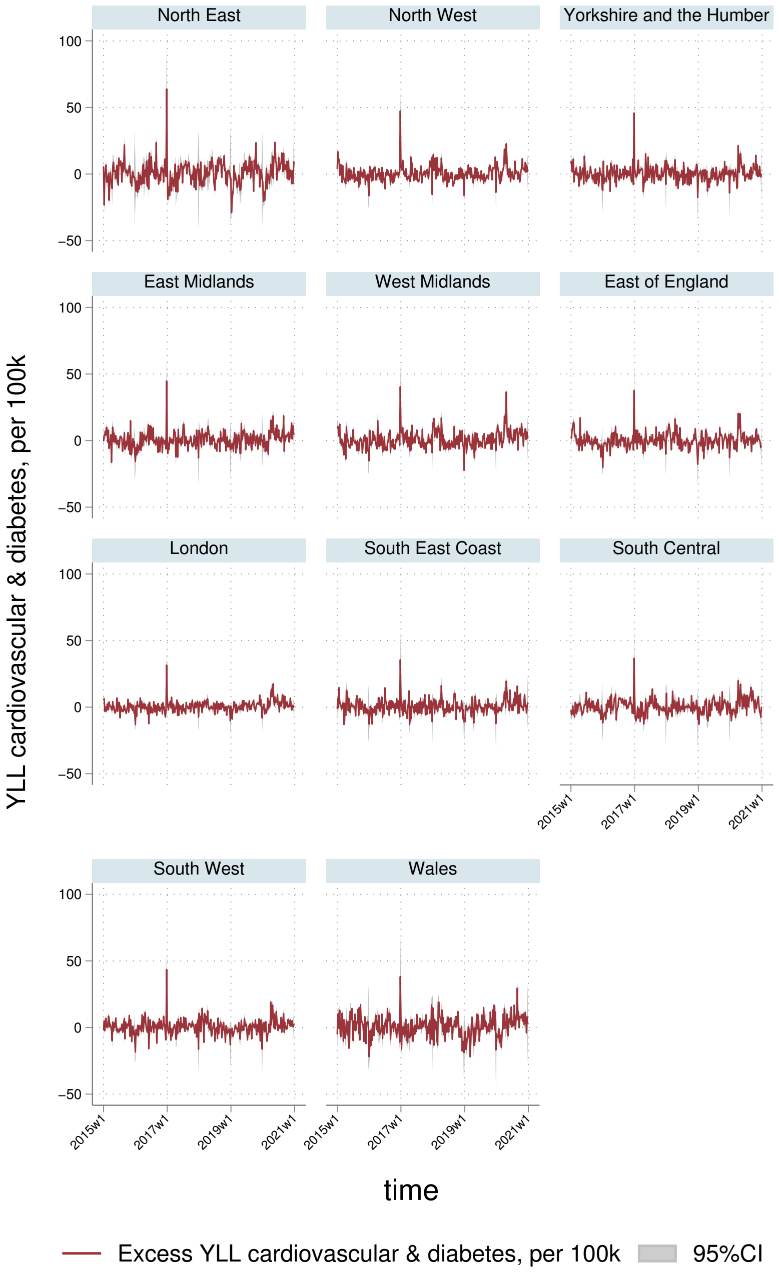
North East
North West
Yorkshire and the Humber
East Midlands
West Midlands
East of England
London
South East Coast
South Central
South West
Wales
100
50
0
−50
2015w1
2017w1
2019w1
2021w1
YLL cardiovascular & diabetes, per 100k
time
Excess YLL cardiovascular & diabetes, per 100k
95%CI

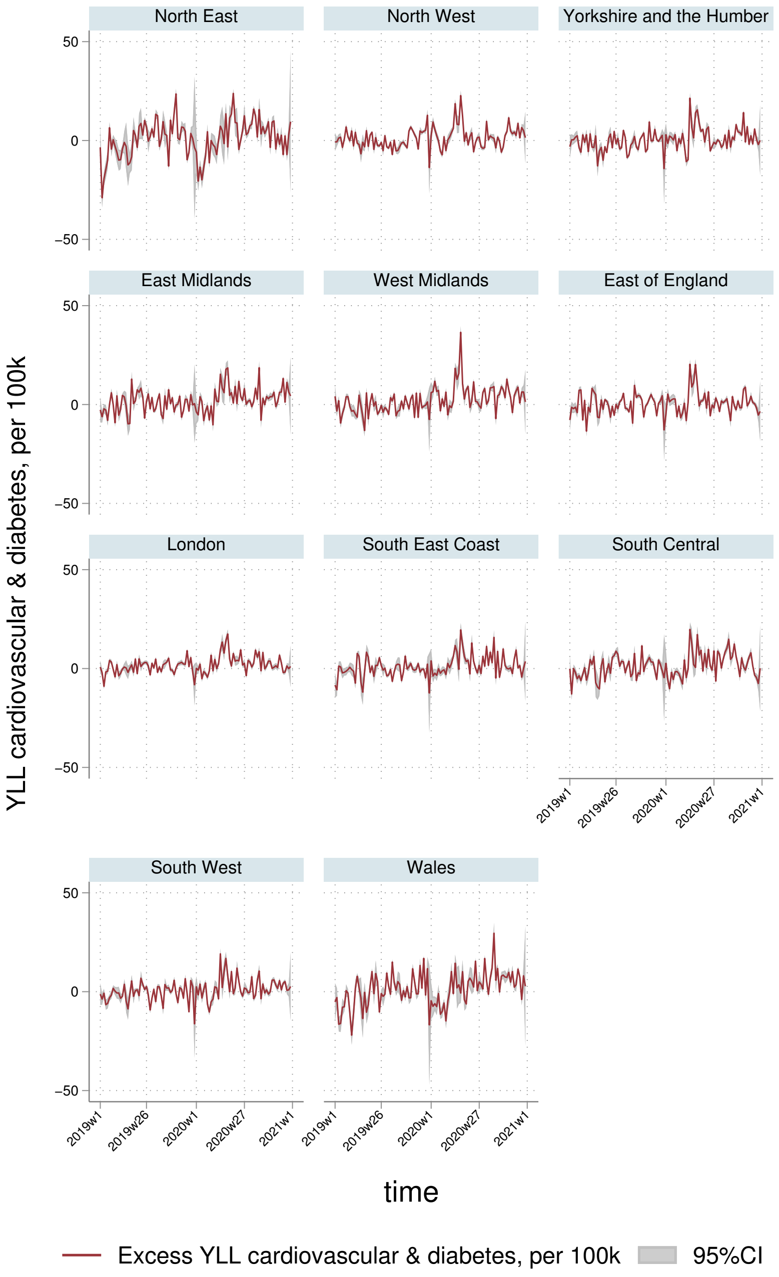
North East
North West
Yorkshire and the Humber
East Midlands
West Midlands
East of England
London
South East Coast
South Central
South West
Wales
50
0
−50
2019w1
2019w26
2020w1
2020w27
2021w1
YLL cardiovascular & diabetes, per 100k
time
Excess YLL cardiovascular & diabetes, per 100k
95%CI

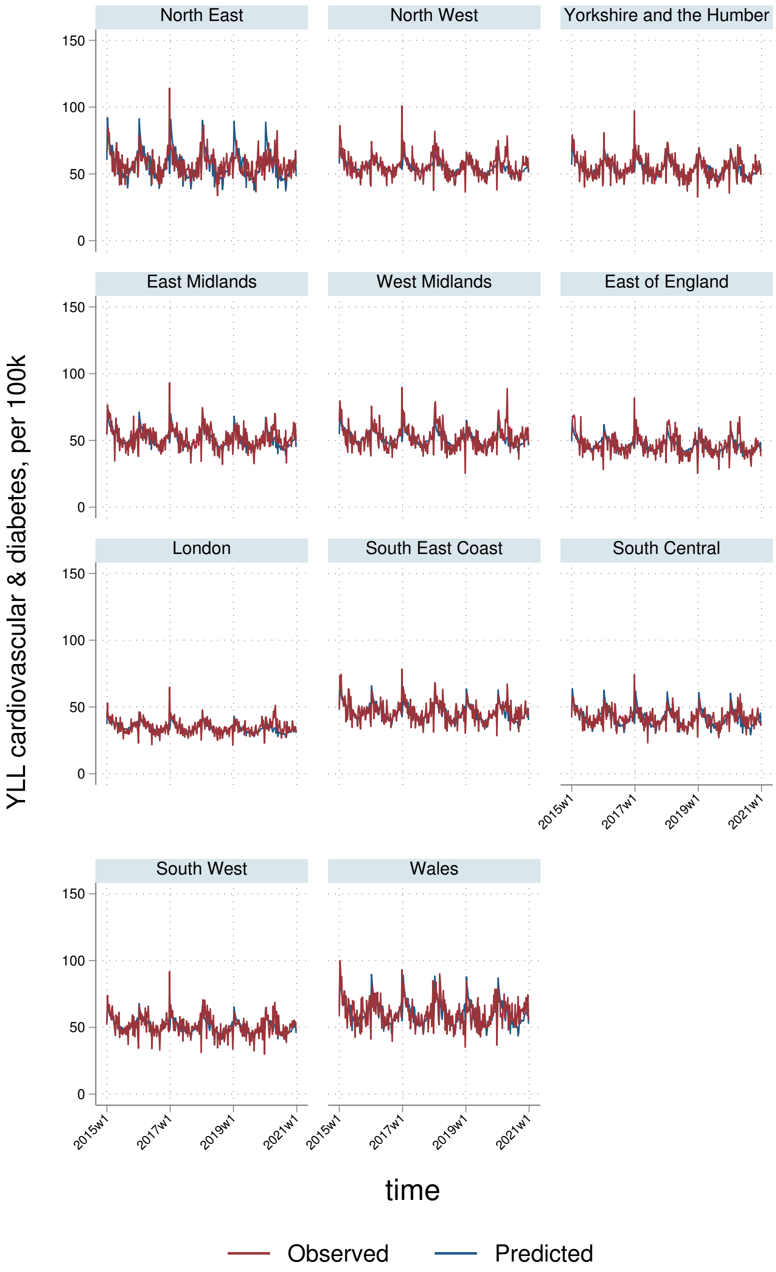

North East
North West
Yorkshire and the Humber
East Midlands
West Midlands
East of England
London
South East Coast
South Central
South West
Wales
150
100
50
0
2015w1
2017w1
2019w1
2021w1
YLL cardiovascular & diabetes, per 100k
time
Observed
Predicted

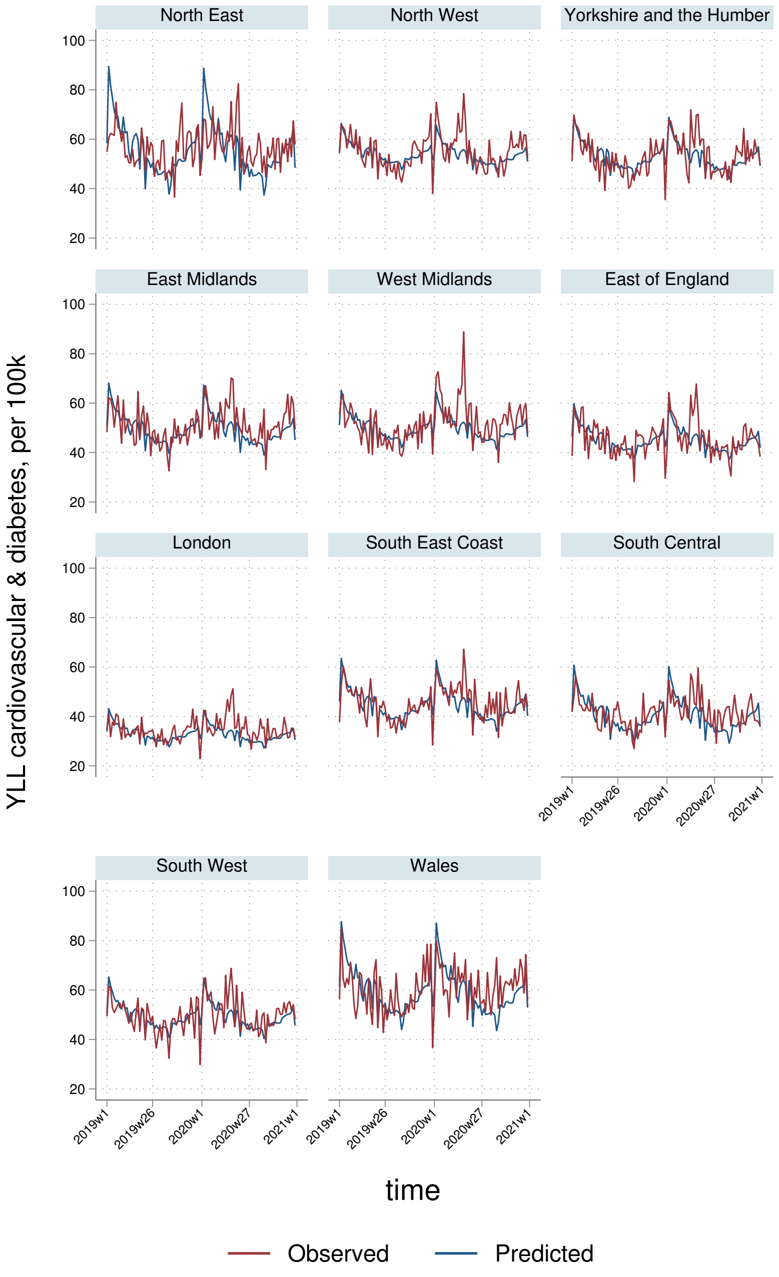
North East
North West
Yorkshire and the Humber
East Midlands
West Midlands
East of England
London
South East Coast
South Central
South West
Wales
YLL cardiovascular & diabetes, per 100k
100
80
60
40
20
2019w1
2019w26
2020w1
2020w27
2021w1
time
Observed
Predicted

### 8 Cancer

#### 8.1 AASMRs

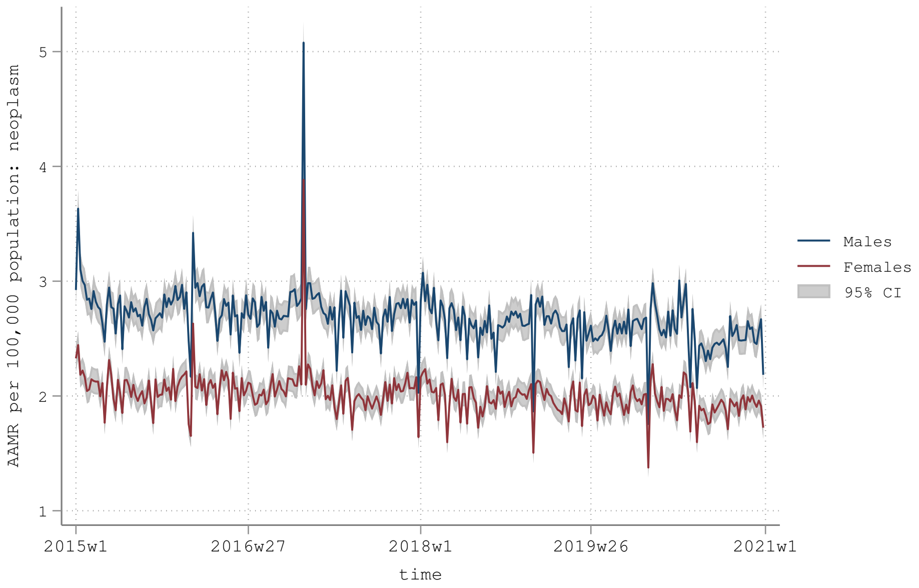


AAMR: age-adjusted mortality rate using the 2020-2025 WHO reference population (see https://www.who.int/healthinfo/paper31.pdf)

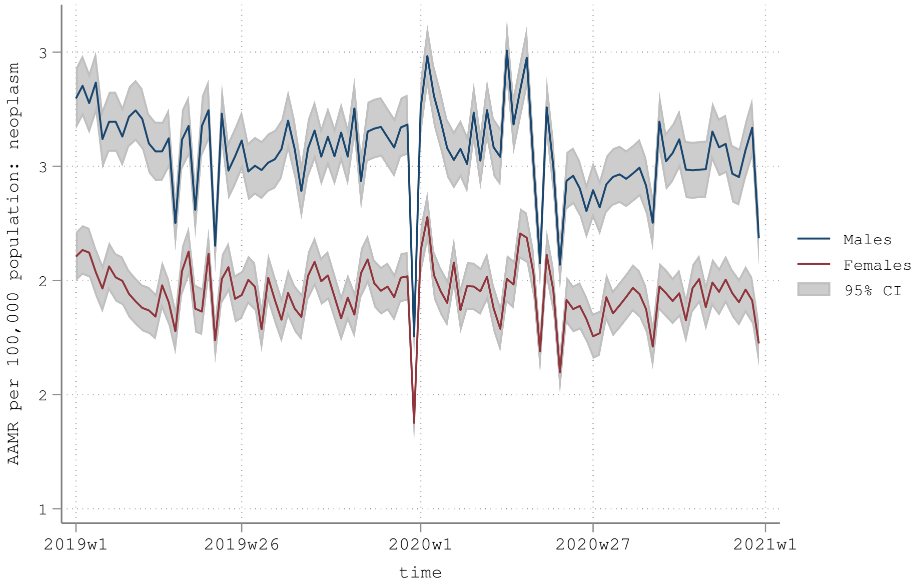


AAMR: age-adjusted mortality rate using the 2020-2025 WHO reference population (see https://www.who.int/healthinfo/paper31.pdf)

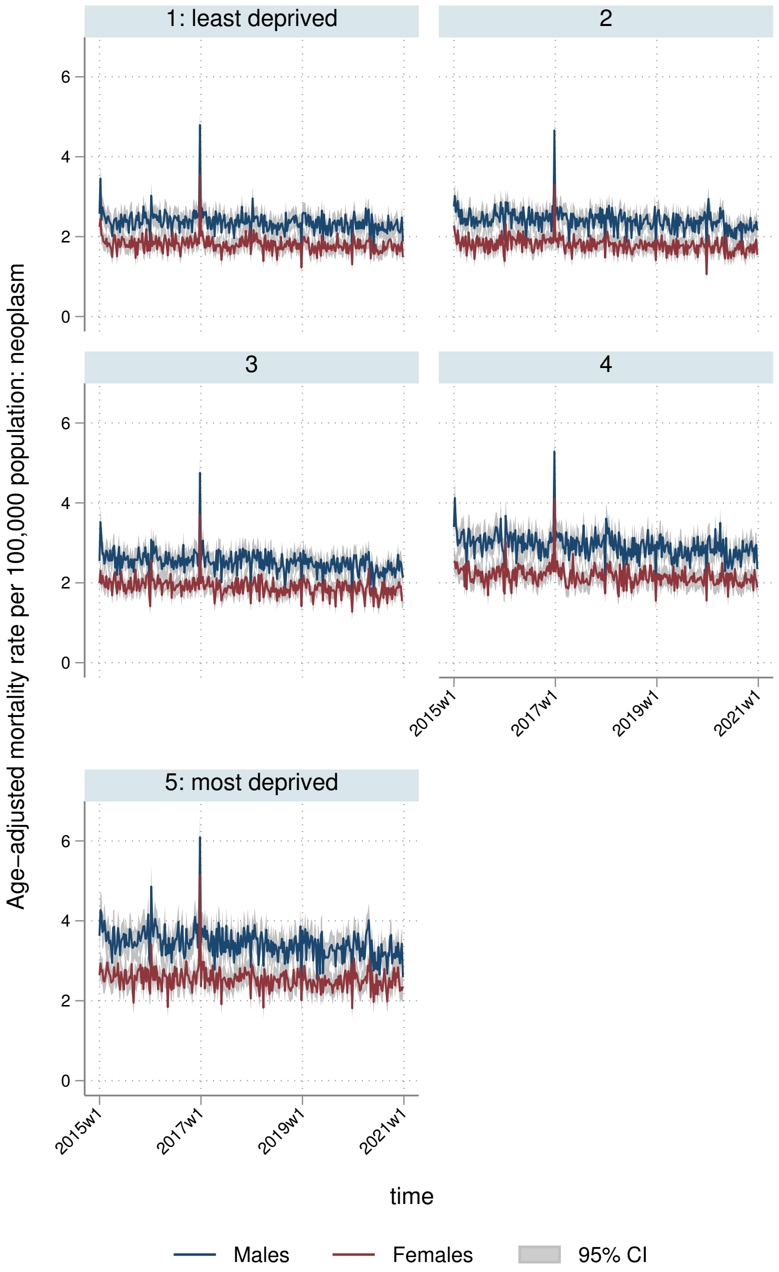
1: least deprived
2
3
4
5: most deprived
Age-adjusted mortality rate per 100,000 population: neoplasm
0
2
4
6
2015w1
2017w1
2019w1
2021w1
time
Males
Females
95% CI

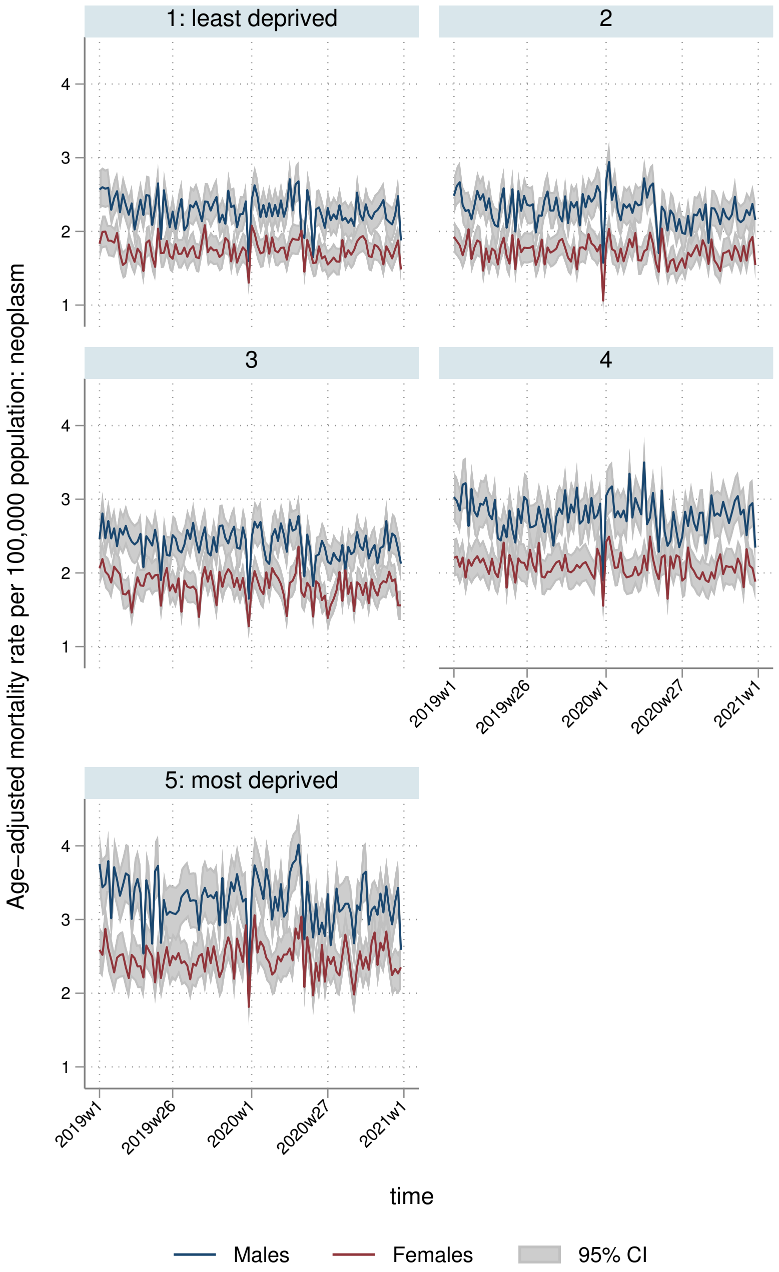

1: least deprived
2
3
4
5: most deprived
Age−adjusted mortality rate per 100,000 population: neoplasm
1
2
3
4
2019w1
2019w26
2020w1
2020w27
2021w1
time
Males
Females
95% CI

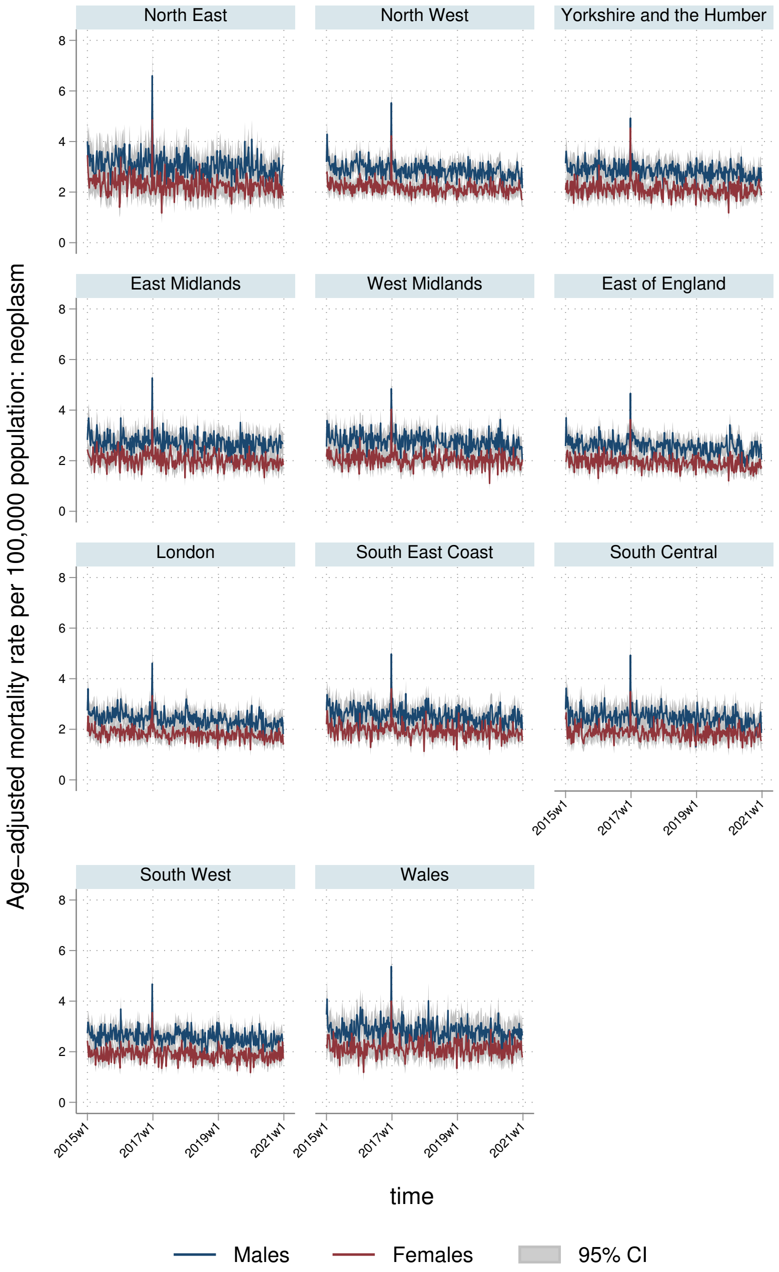
North East
North West
Yorkshire and the Humber
East Midlands
West Midlands
East of England
London
South East Coast
South Central
South West
Wales
Age-adjusted mortality rate per 100,000 population: neoplasm
0
2
4
6
8
2015w1
2017w1
2019w1
2021w1
time
Males
Females
95% CI

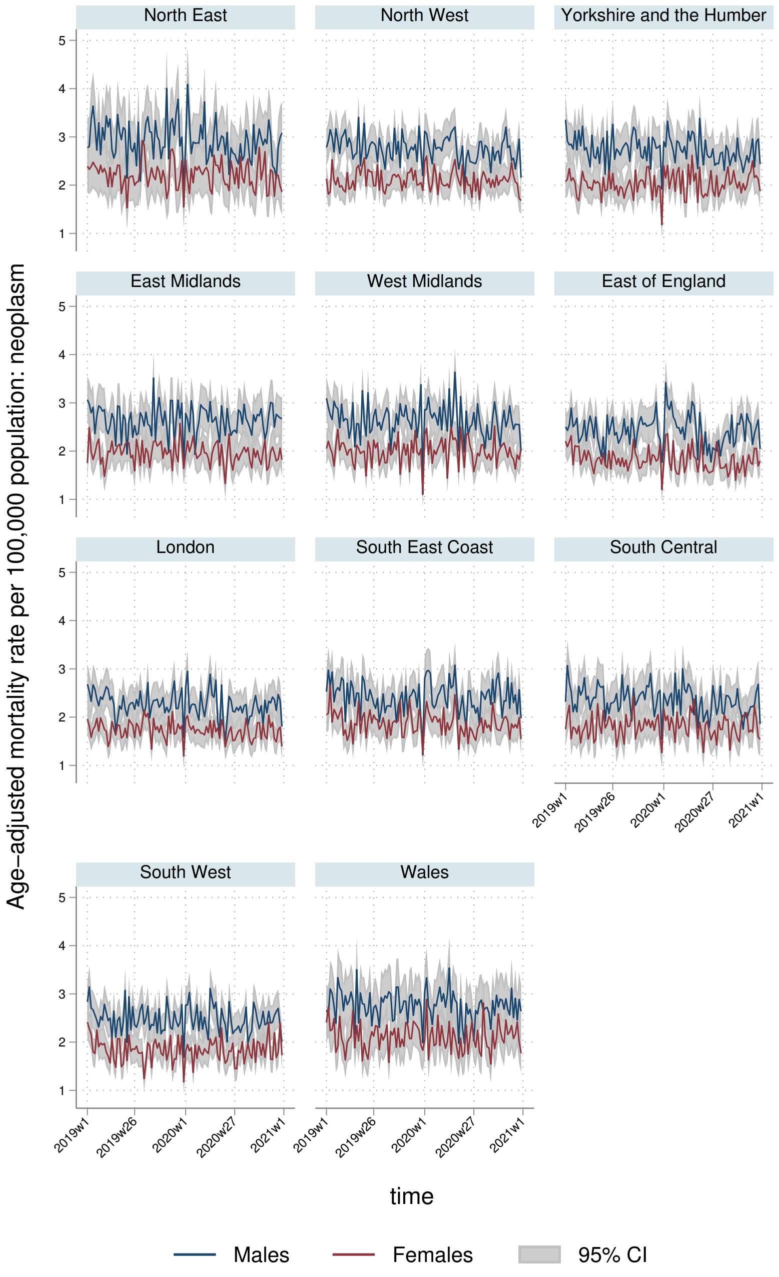
North East
North West
Yorkshire and the Humber
East Midlands
West Midlands
East of England
London
South East Coast
South Central
South West
Wales
Age-adjusted mortality rate per 100,000 population: neoplasm
1
2
3
4
5
2019w1
2019w26
2020w1
2020w27
2021w1
time
Males
Females
95% CI

#### 8.2 YLLs

##### 8.2.1 England-Wales aggregate

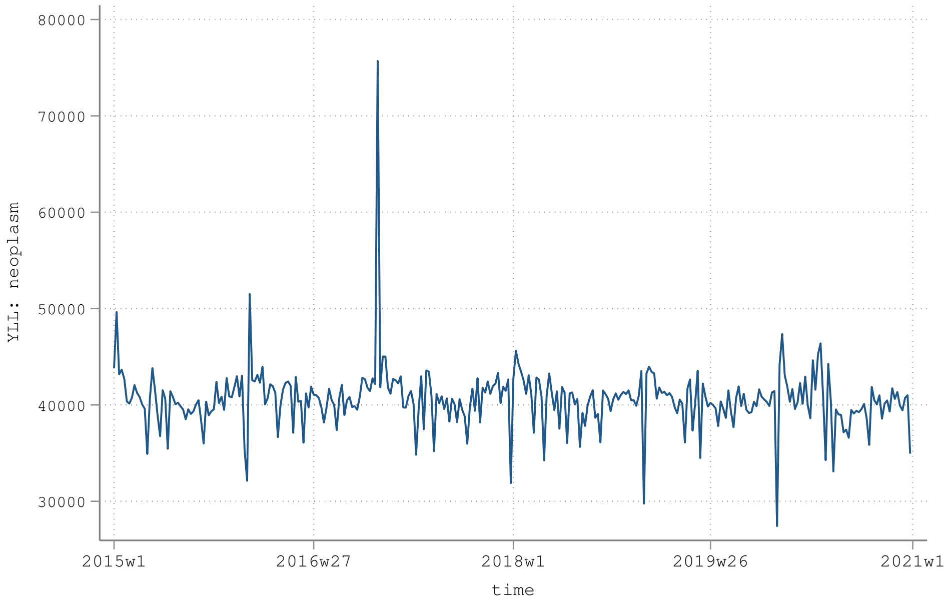


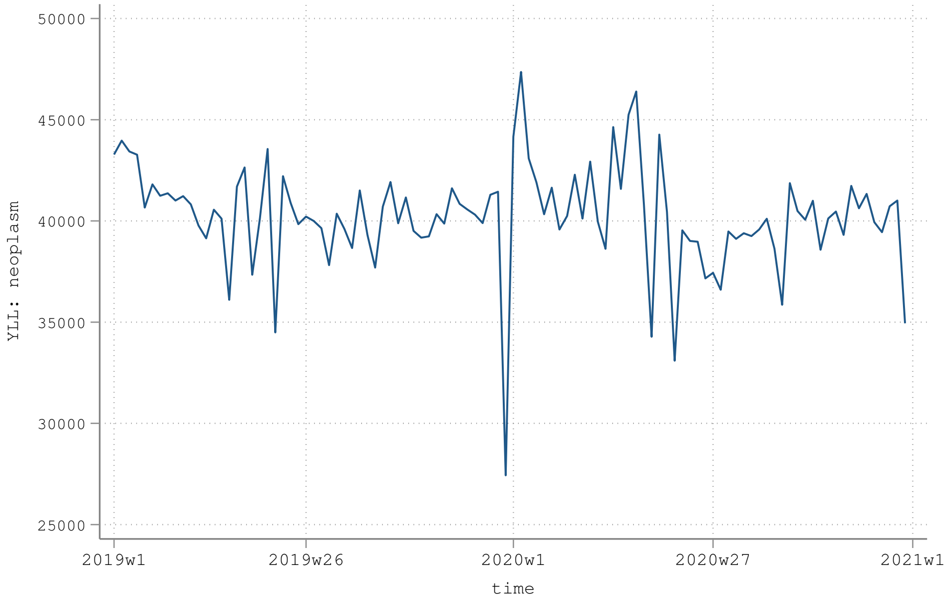

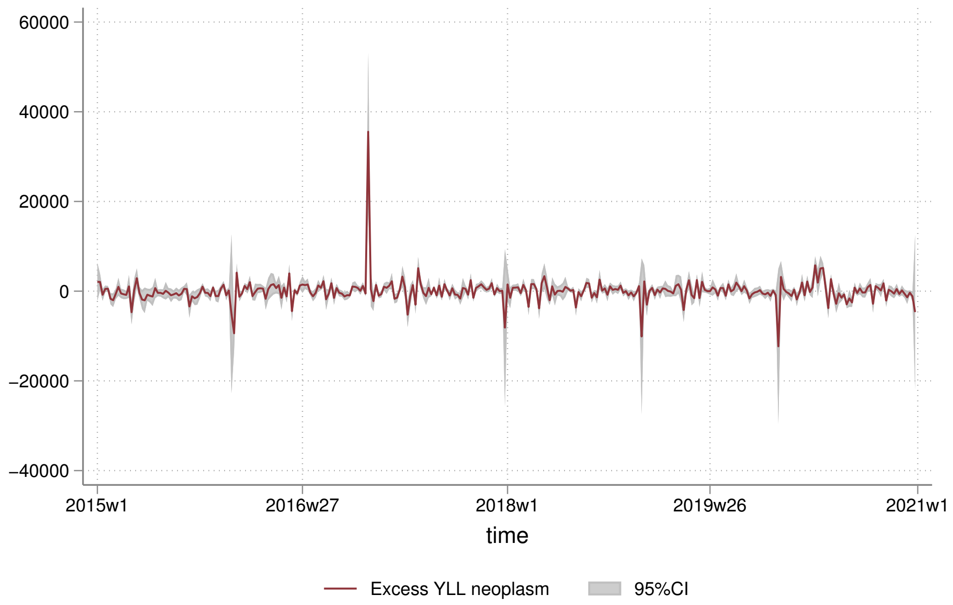
60000
40000
20000
0
−20000
−40000
2015w1
2016w27
2018w1
2019w26
2021w1
time
Excess YLL neoplasm
95%CI

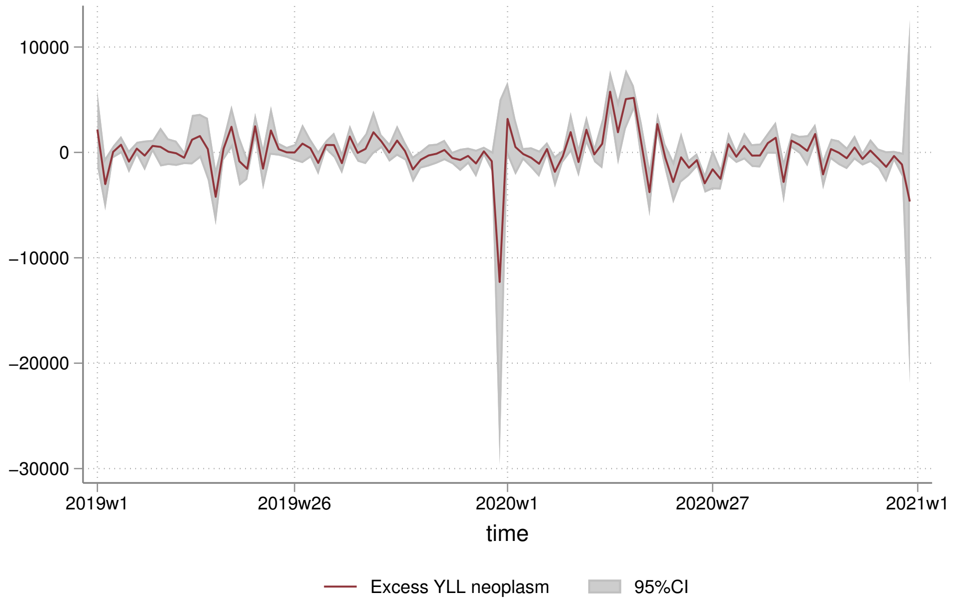
10000
0
−10000
−20000
−30000
2019w1
2019w26
2020w1
2020w27
2021w1
time
Excess YLL neoplasm
95%CI

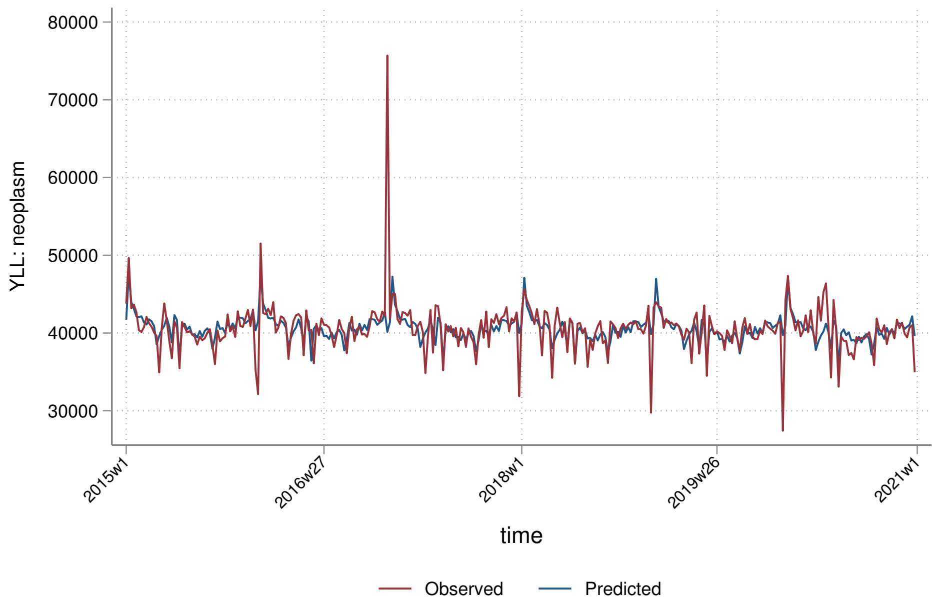
80000
70000
60000
50000
40000
30000
YLL: neoplasm
2015w1
2016w27
2018w1
2019w26
2021w1
time
Observed
Predicted

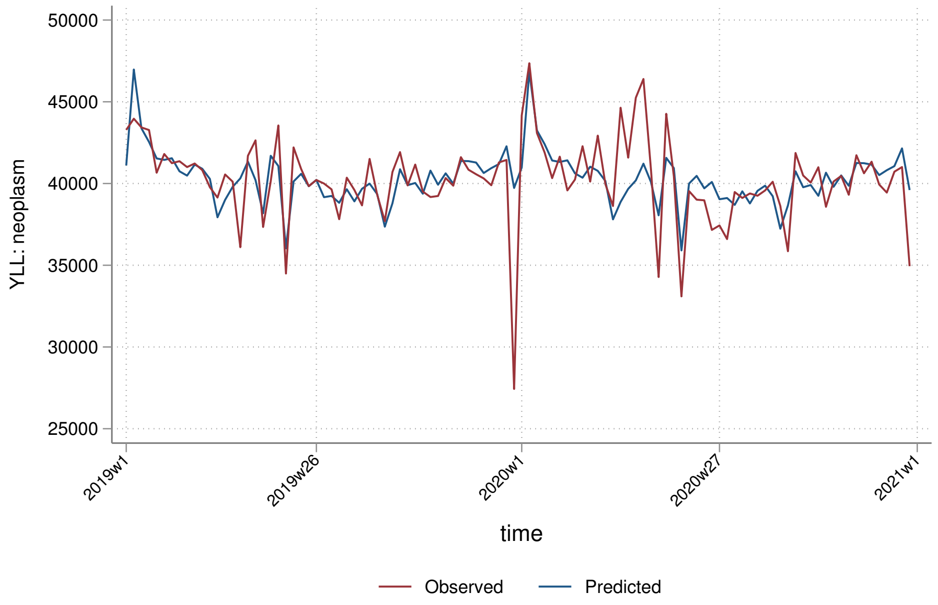
50000
45000
40000
35000
30000
25000
YLL: neoplasm
2019w1
2019w26
2020w1
2020w27
2021w1
time
Observed
Predicted

##### 8.2.2 By sex

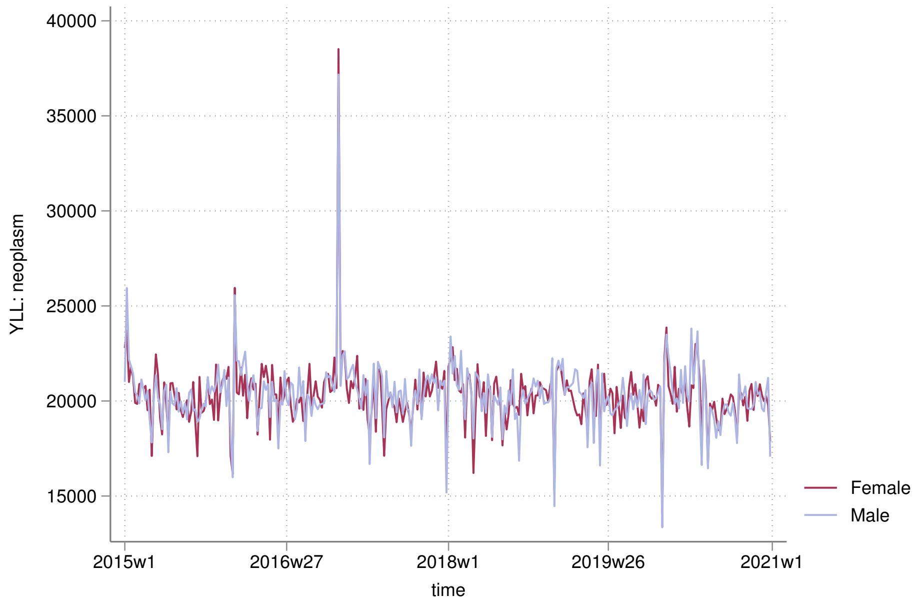

40000
35000
30000
25000
20000
15000
YLL: neoplasm
2015w1
2016w27
2018w1
2019w26
2021w1
time
Female
Male


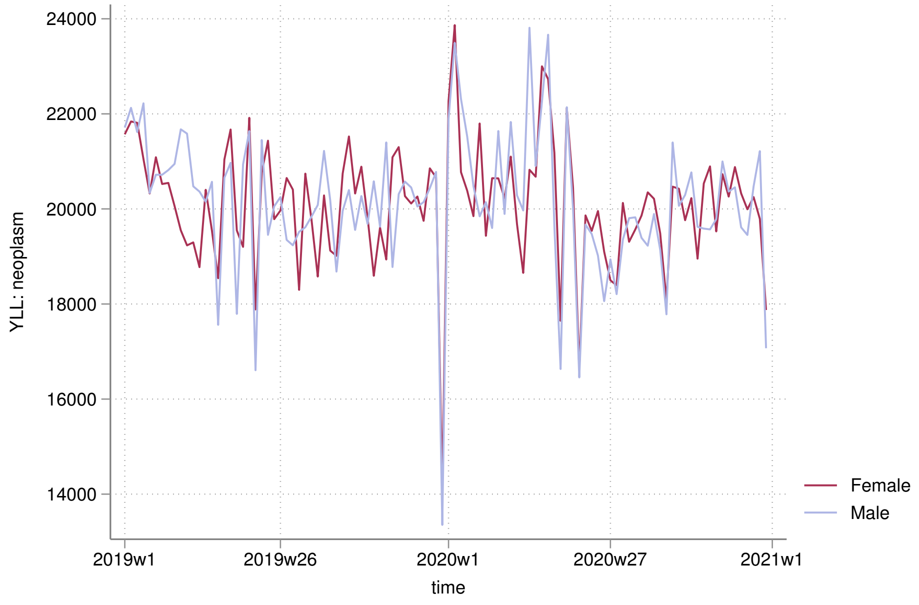

24000
22000
20000
18000
16000
14000
YLL: neoplasm
2019w1
2019w26
2020w1
2020w27
2021w1
time
Female
Male

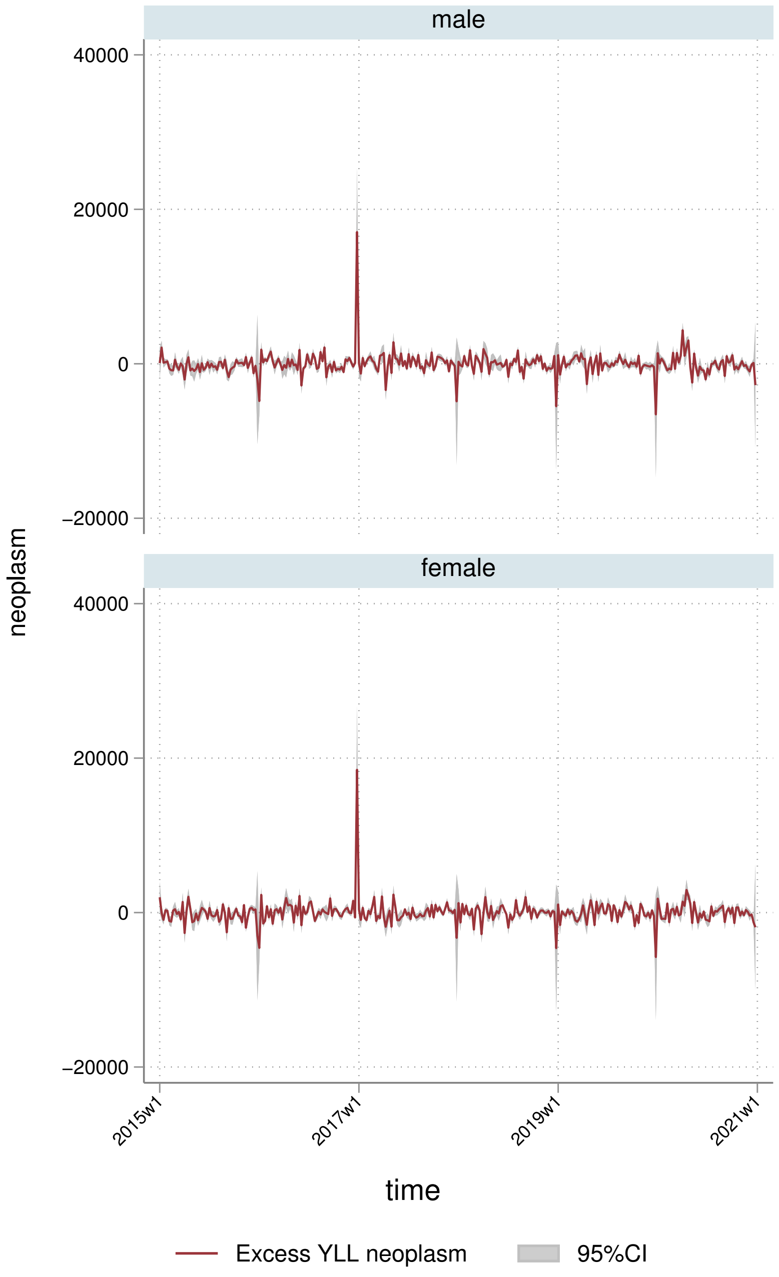
male
female
40000
20000
0
−20000
neoplasm
2015w1
2017w1
2019w1
2021w1
time
Excess YLL neoplasm
95%CI

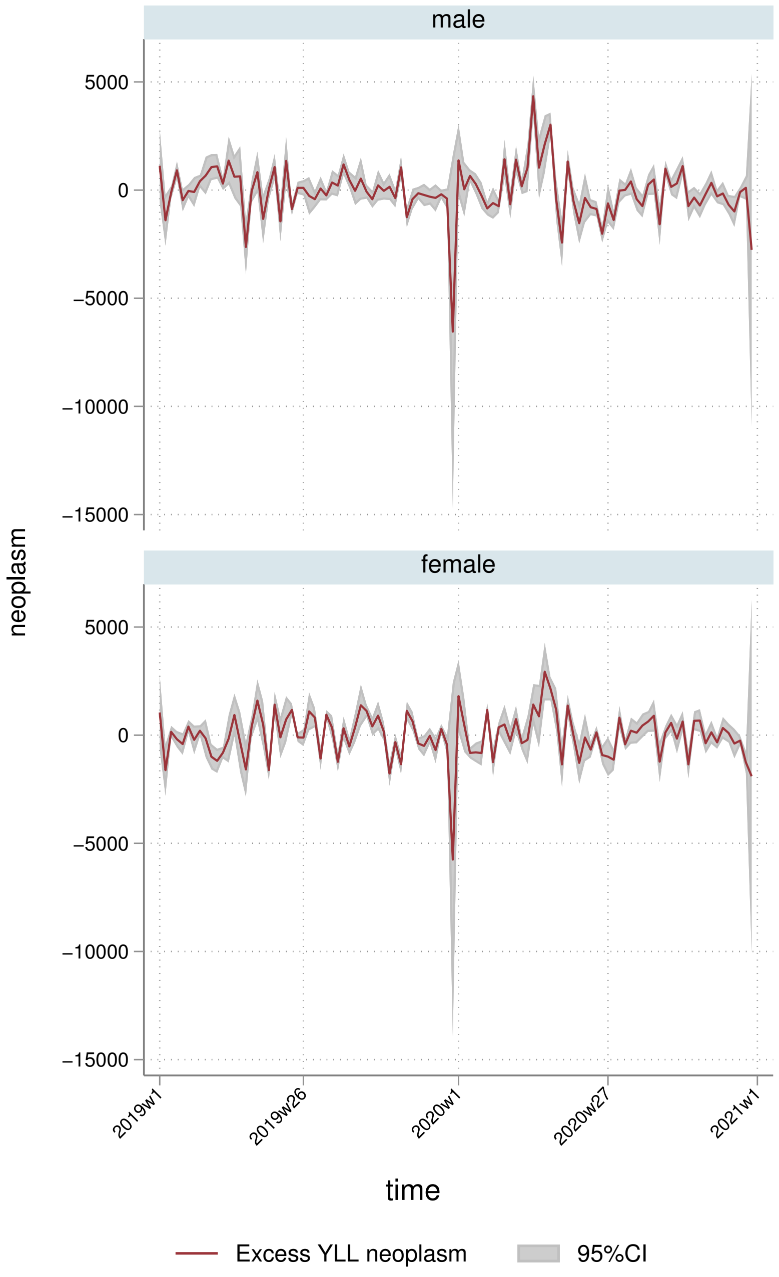
male
female
5000
0
−5000
−10000
−15000
neoplasm
2019w1
2019w26
2020w1
2020w27
2021w1
time
Excess YLL neoplasm
95%CI

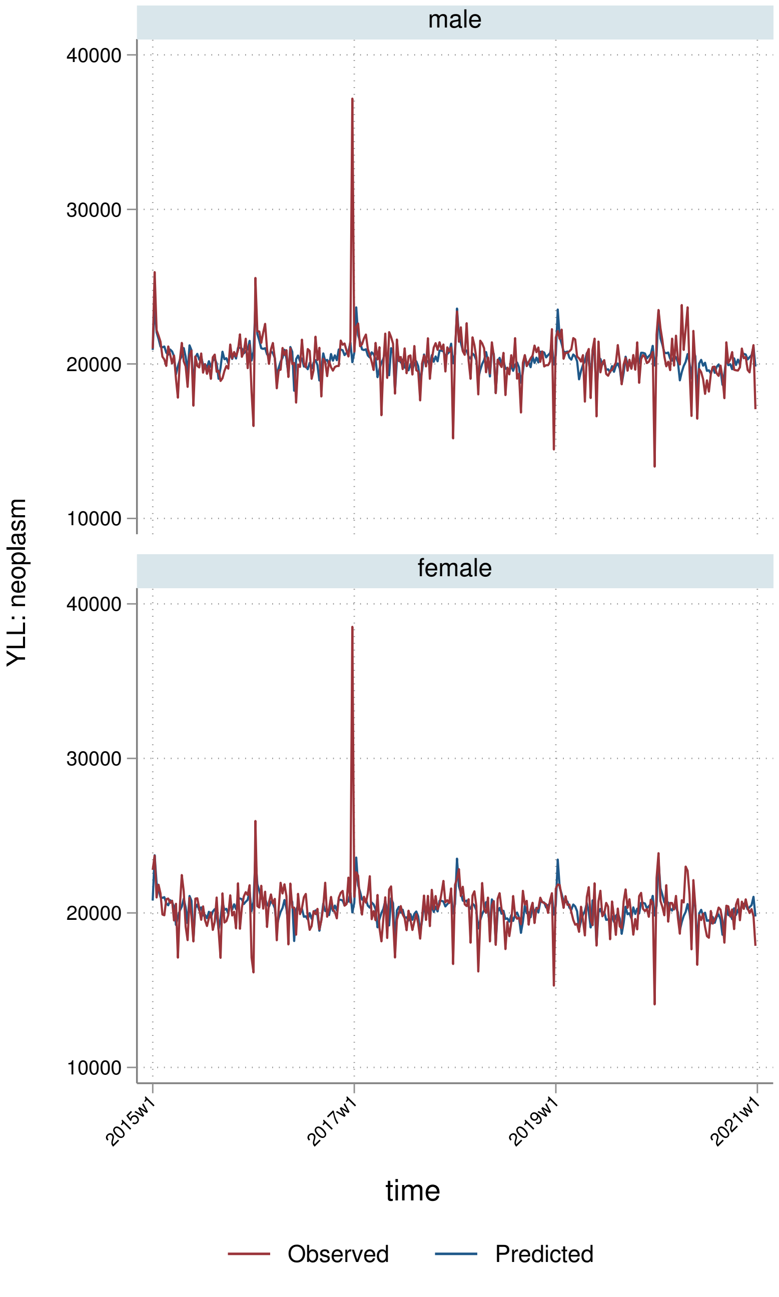
male
female
YLL: neoplasm
40000
30000
20000
10000
2015w1
2017w1
2019w1
2021w1
time
Observed
Predicted

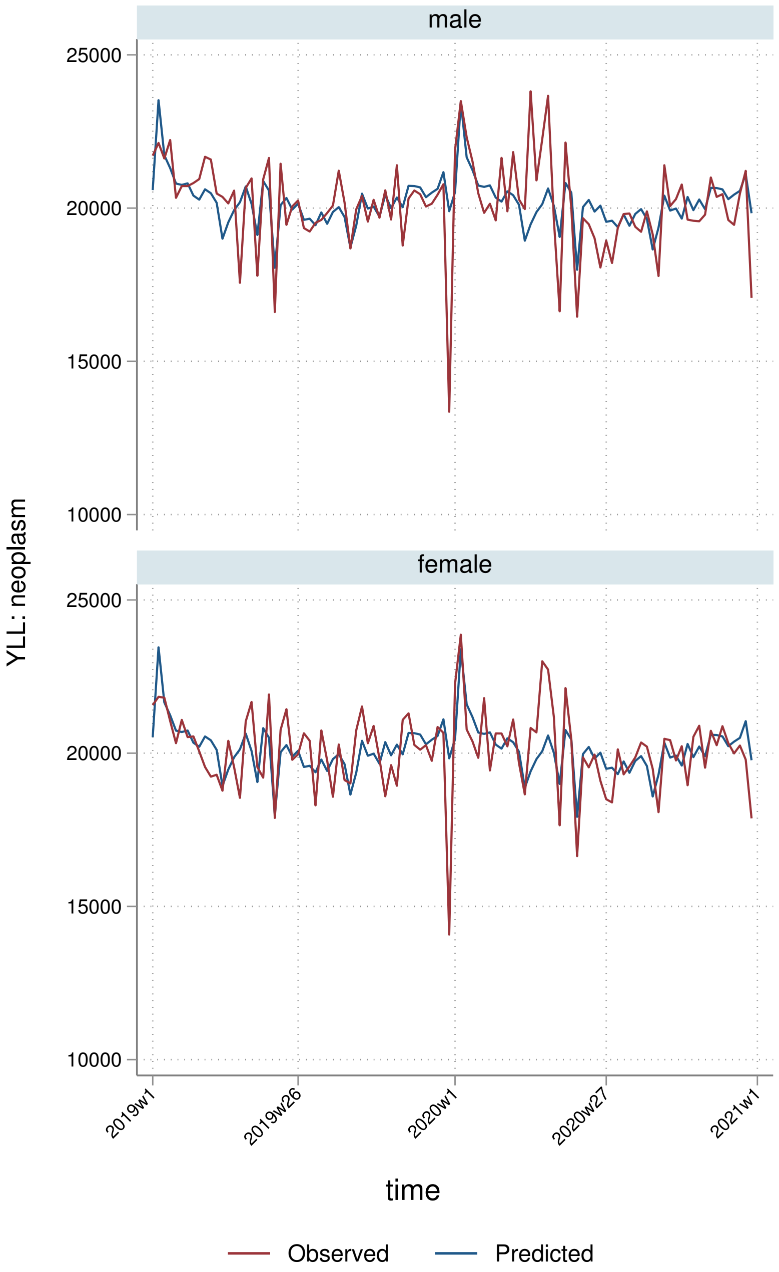
male
female
YLL: neoplasm
25000
20000
15000
10000
2019w1
2019w26
2020w1
2020w27
2021w1
time
Observed
Predicted

##### 8.2.3 By deprivation quintile

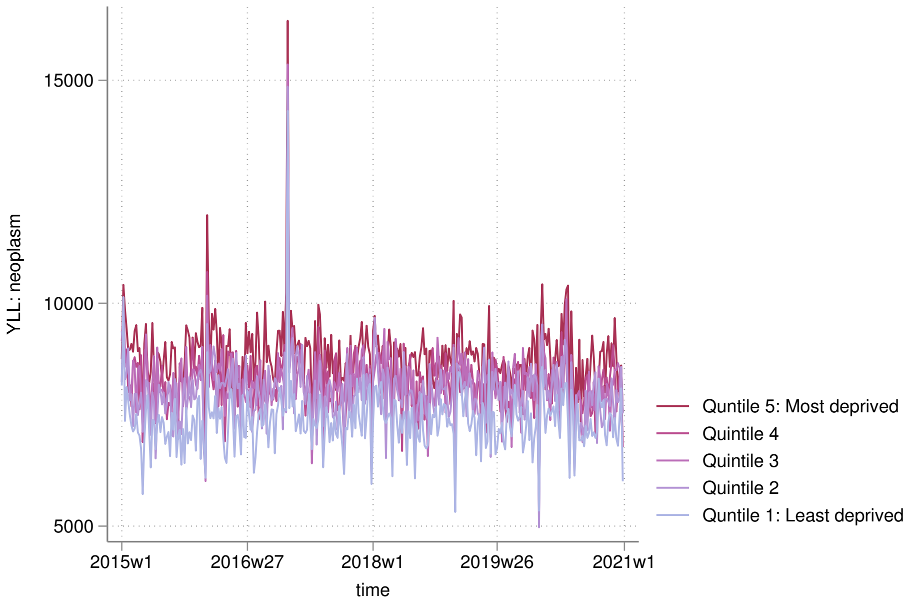


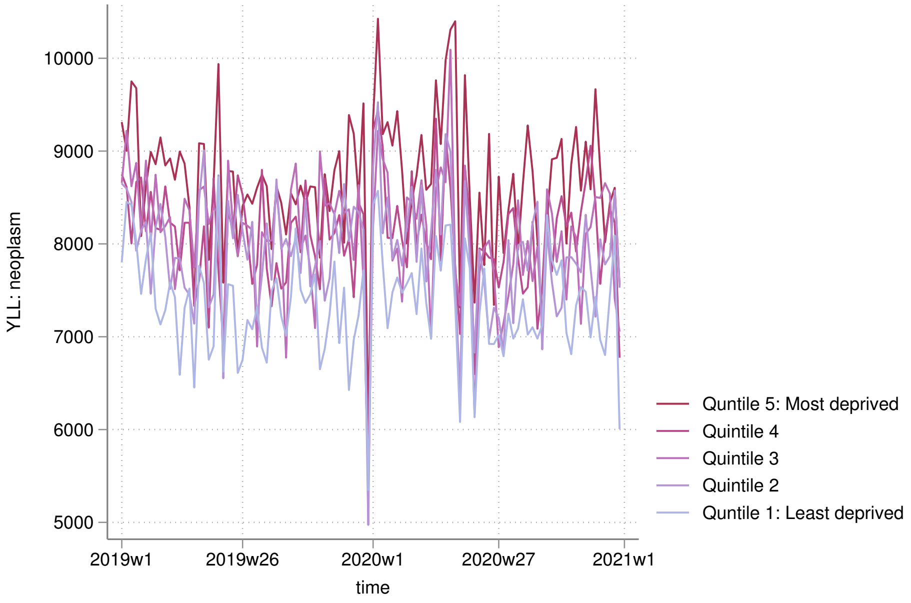

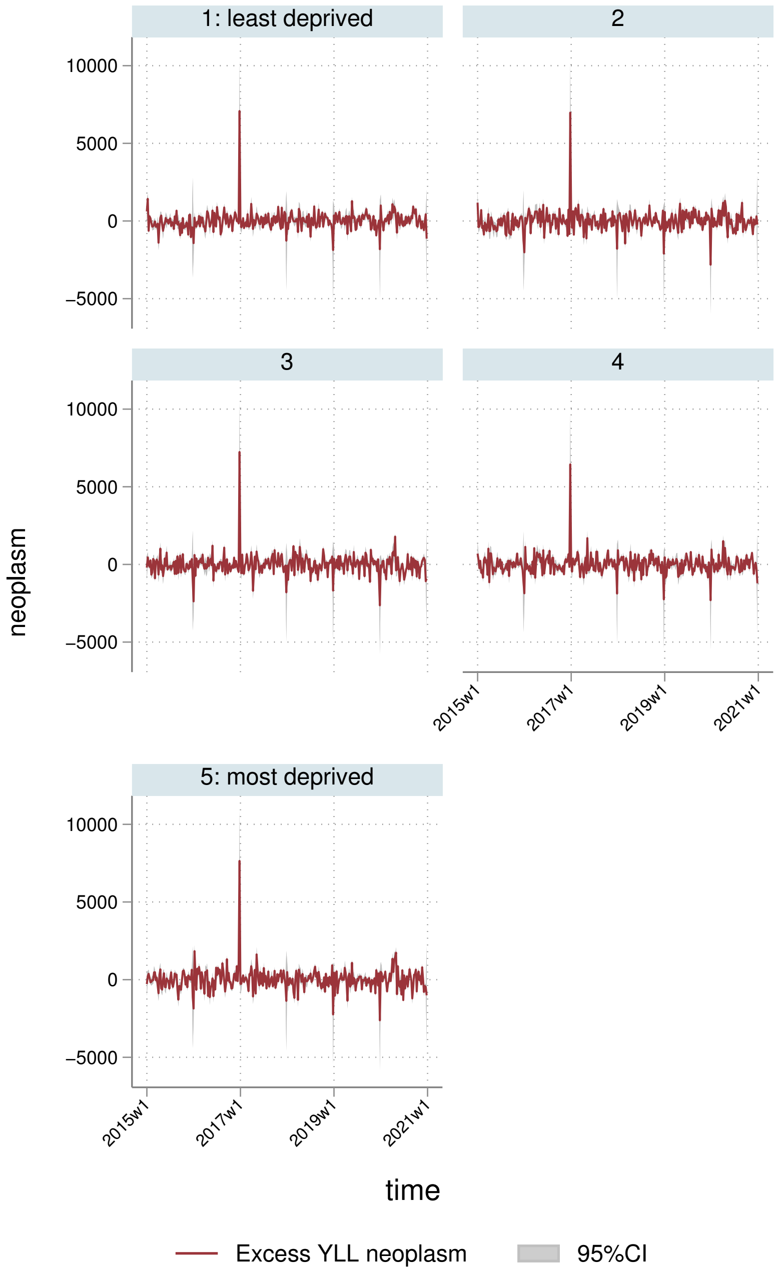

1: least deprived
2
3
4
5: most deprived
10000
5000
0
−5000
neoplasm
2015w1
2017w1
2019w1
2021w1
time
Excess YLL neoplasm
95%CI

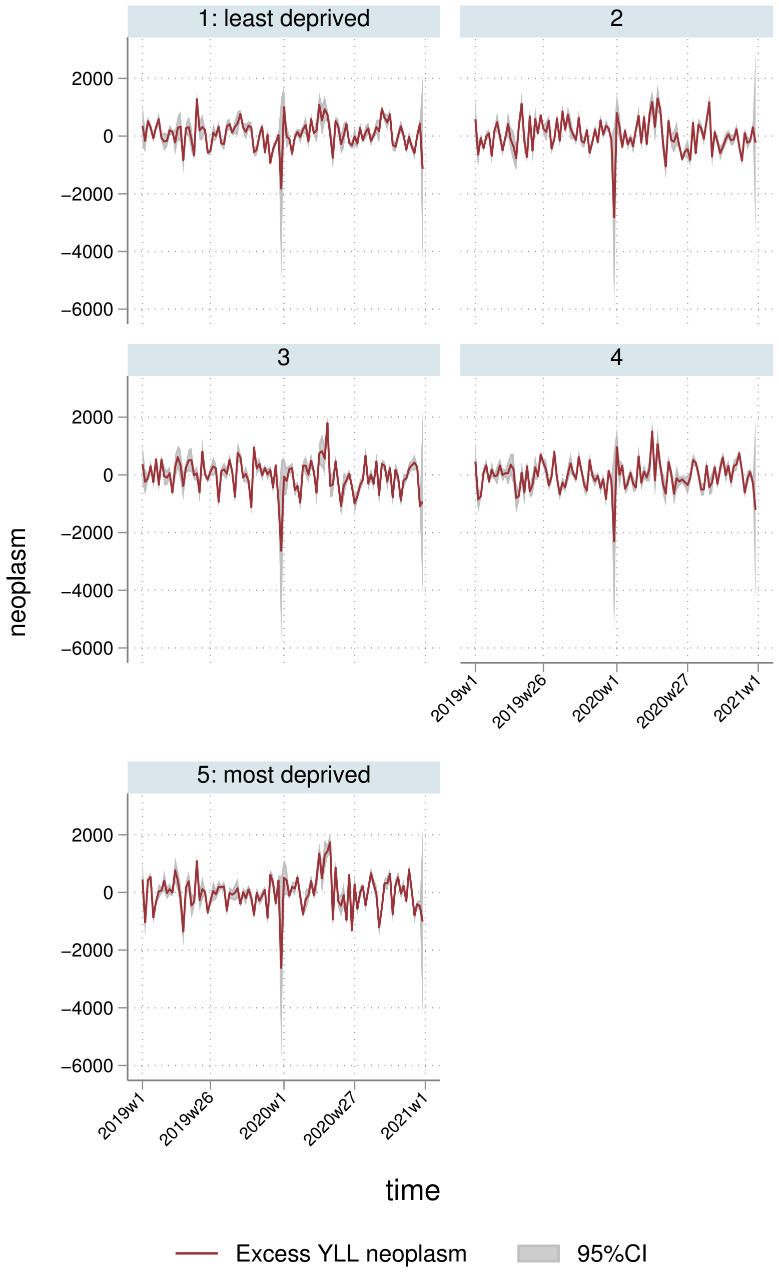

1: least deprived
2
3
4
5: most deprived
2000
0
−2000
−4000
−6000
neoplasm
2019w1
2019w26
2020w1
2020w27
2021w1
time
Excess YLL neoplasm
95%CI

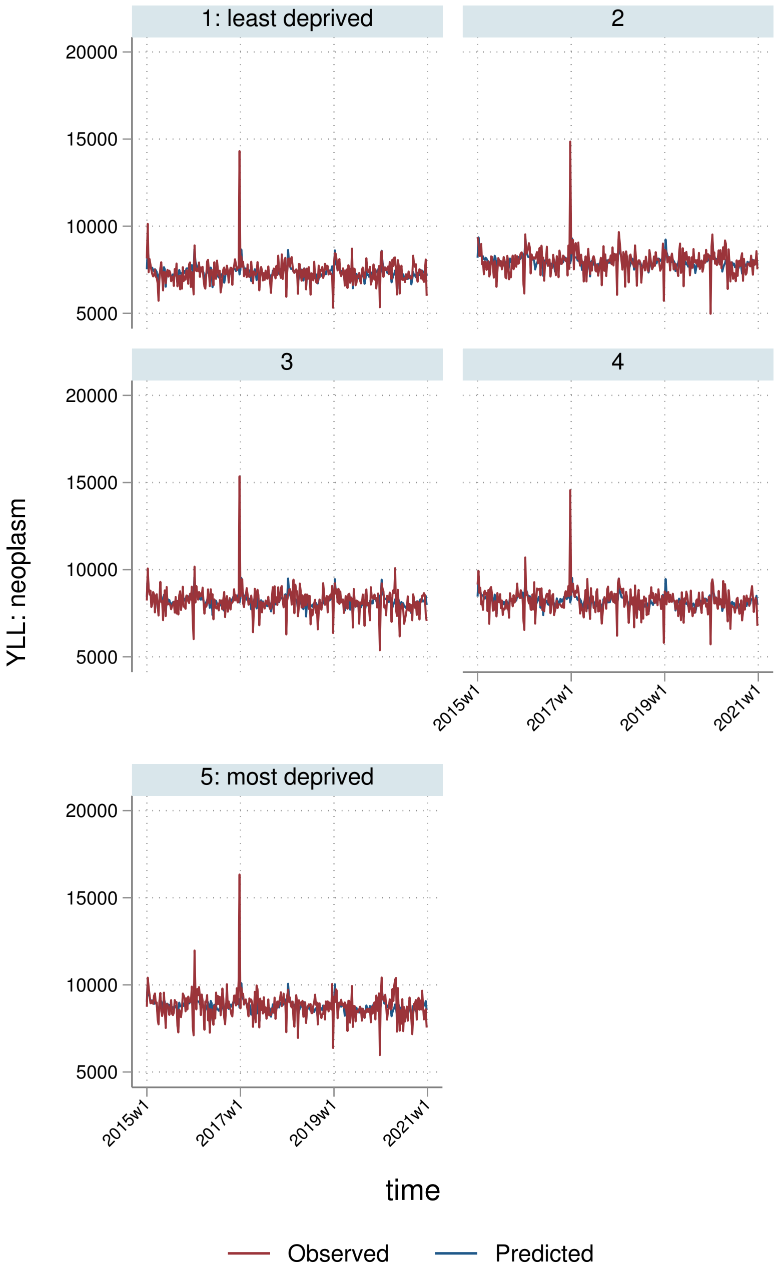

1: least deprived
2
3
4
5: most deprived
20000
15000
10000
5000
YLL: neoplasm
2015w1
2017w1
2019w1
2021w1
time
Observed
Predicted

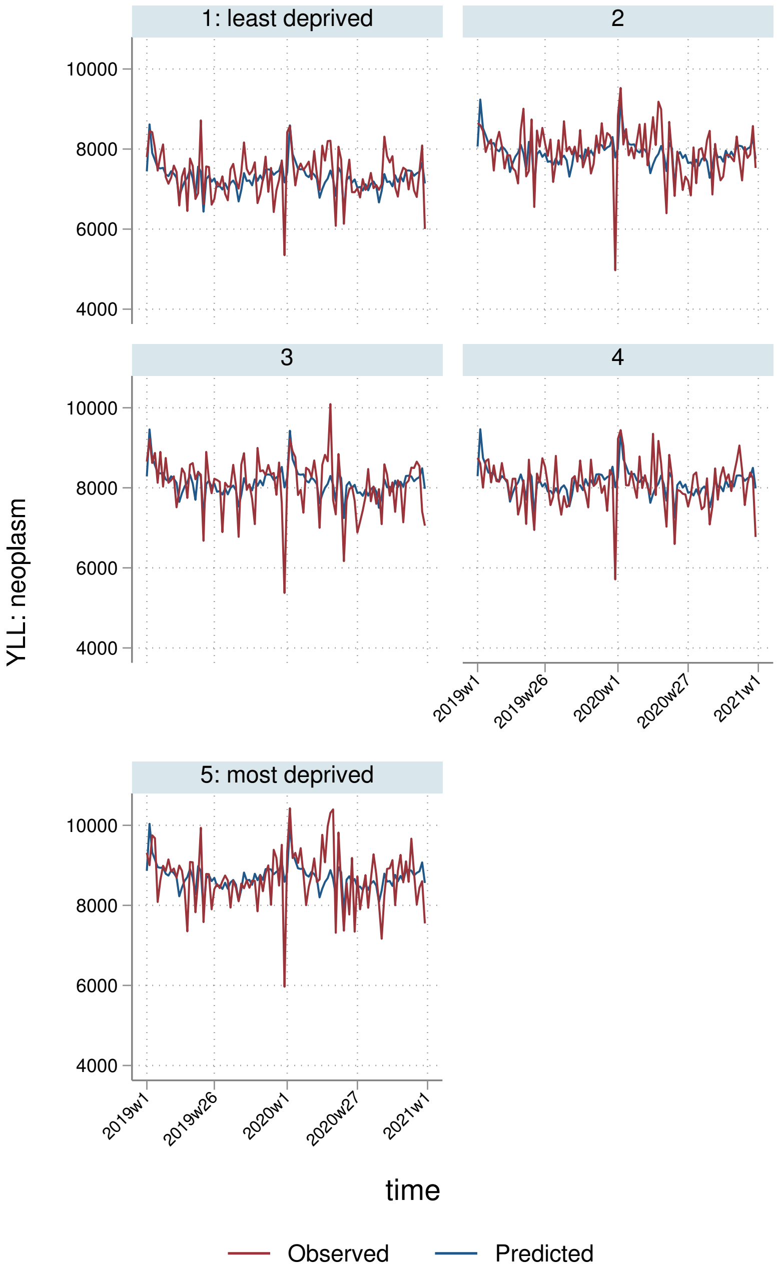
1: least deprived
2
3
4
5: most deprived
10000
8000
6000
4000
2019w1
2019w26
2020w1
2020w27
2021w1
YLL: neoplasm
time
Observed
Predicted

##### 8.2.4 By Strategic Health Authority

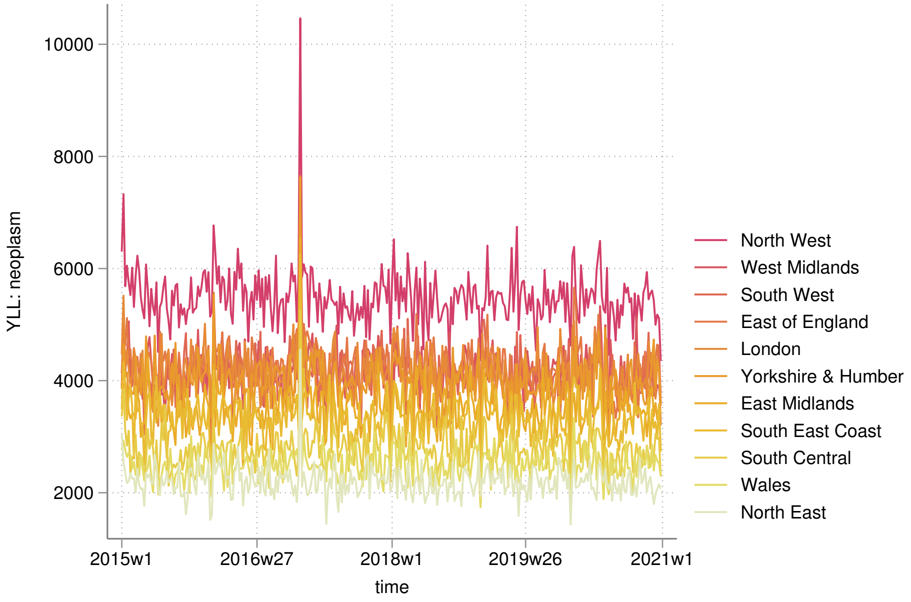


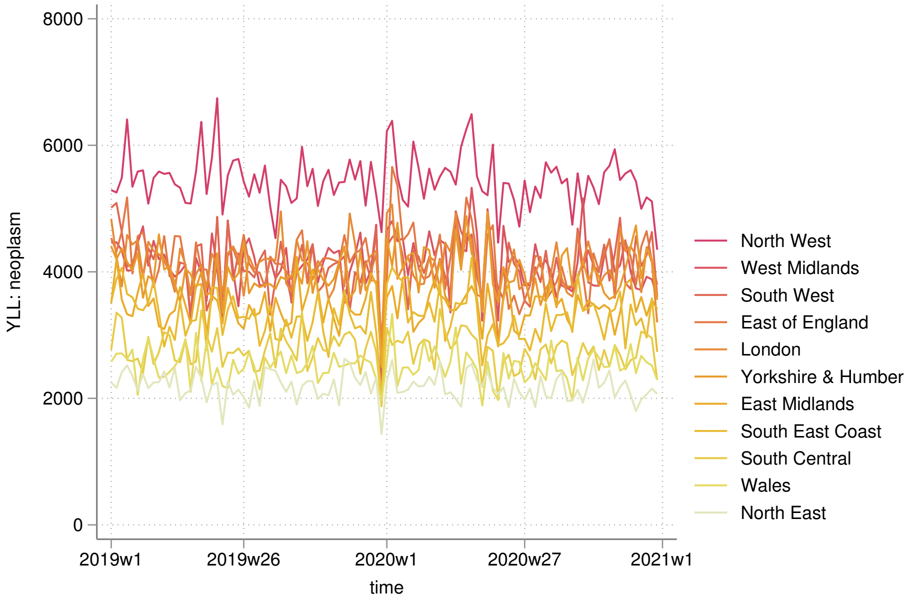

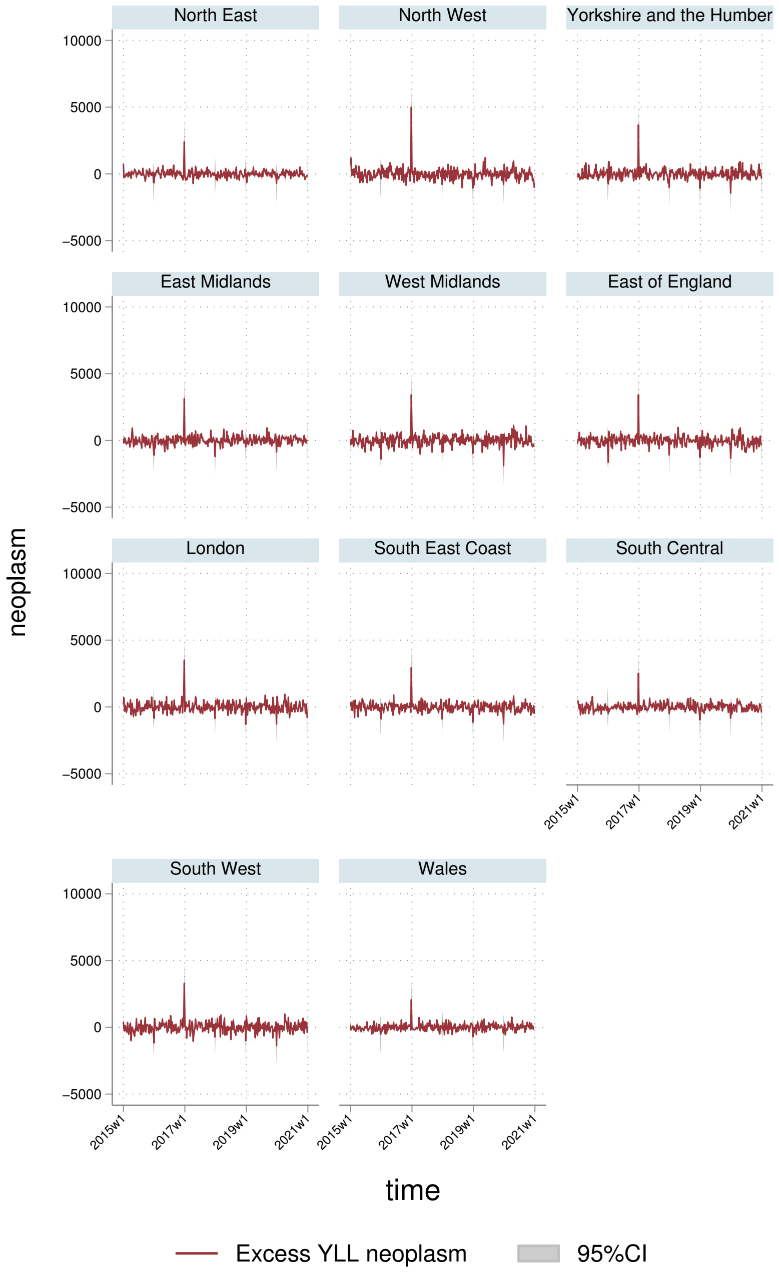
North East
North West
Yorkshire and the Humber
East Midlands
West Midlands
East of England
London
South East Coast
South Central
South West
Wales
10000
5000
0
−5000
2015w1
2017w1
2019w1
2021w1
neoplasm
time
Excess YLL neoplasm
95%CI

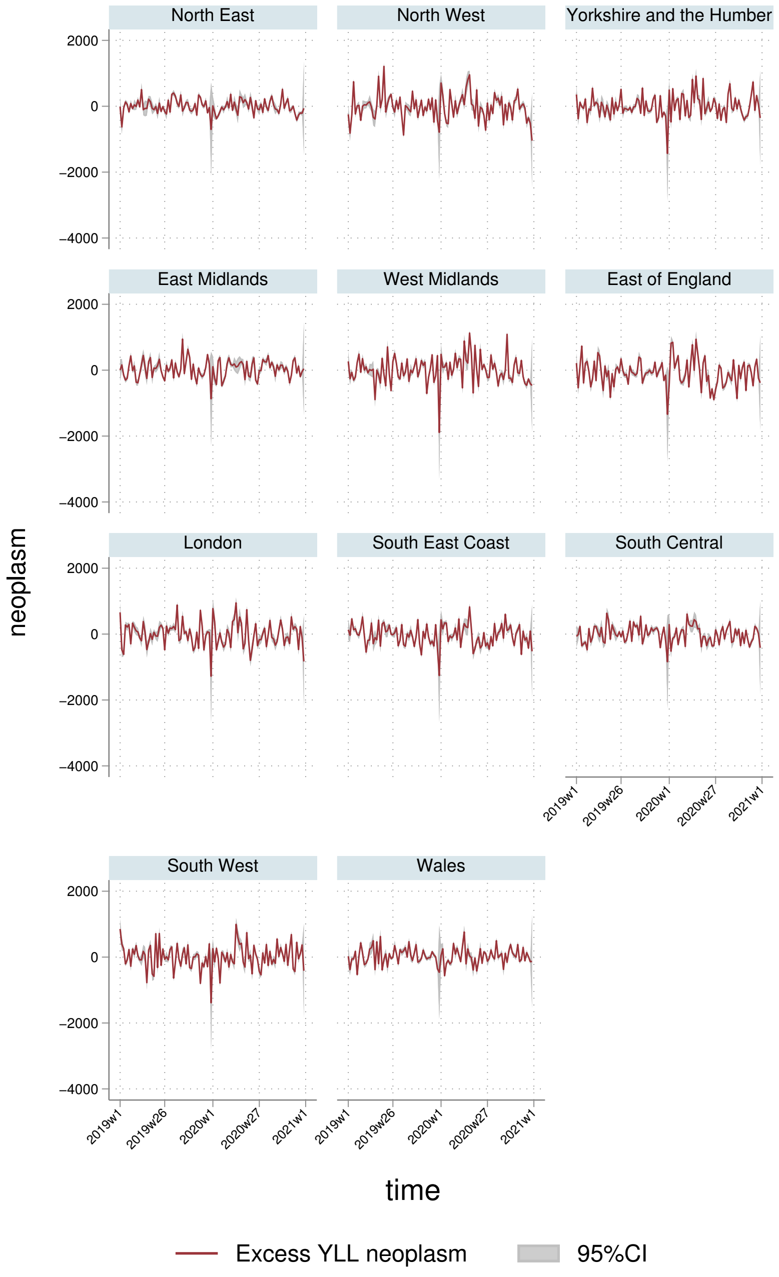
North East
North West
Yorkshire and the Humber
East Midlands
West Midlands
East of England
London
South East Coast
South Central
South West
Wales
2000
0
−2000
−4000
neoplasm
2019w1
2019w26
2020w1
2020w27
2021w1
time
Excess YLL neoplasm
95%CI

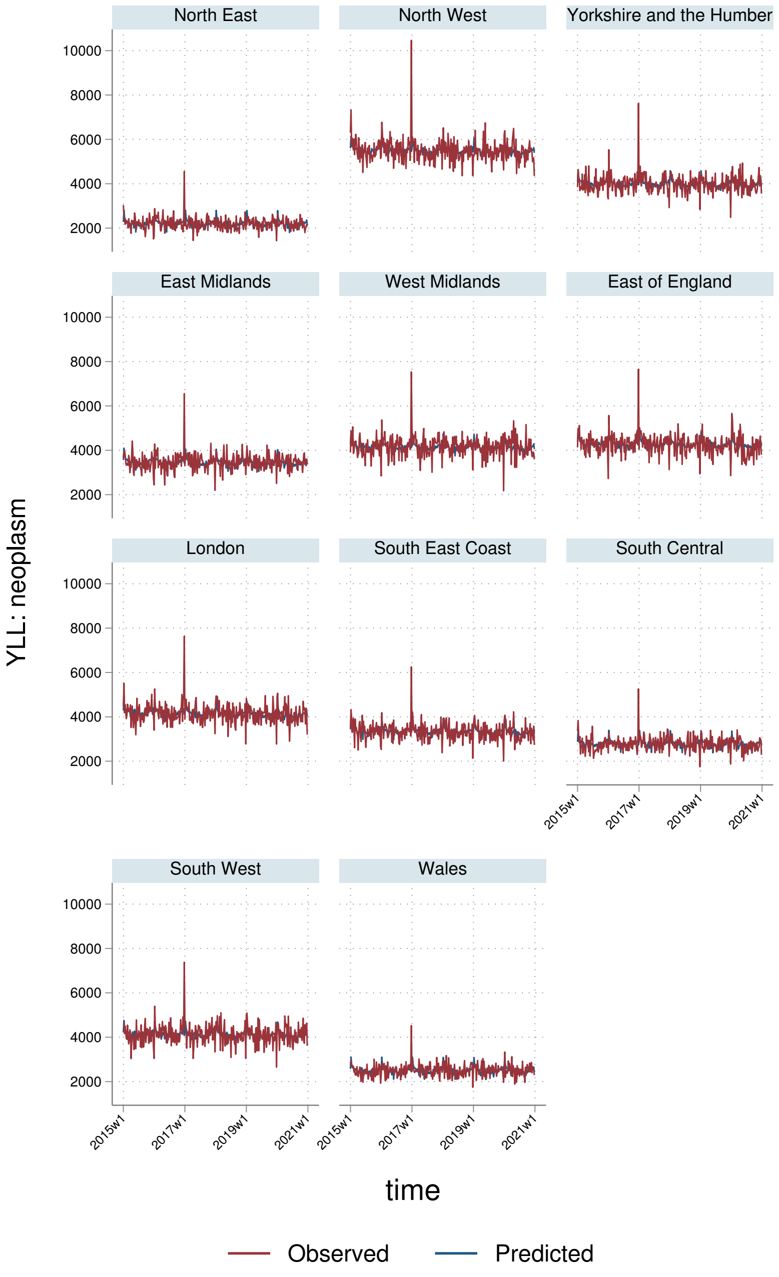
North East
North West
Yorkshire and the Humber
East Midlands
West Midlands
East of England
London
South East Coast
South Central
South West
Wales
YLL: neoplasm
10000
8000
6000
4000
2000
2015w1
2017w1
2019w1
2021w1
time
Observed
Predicted

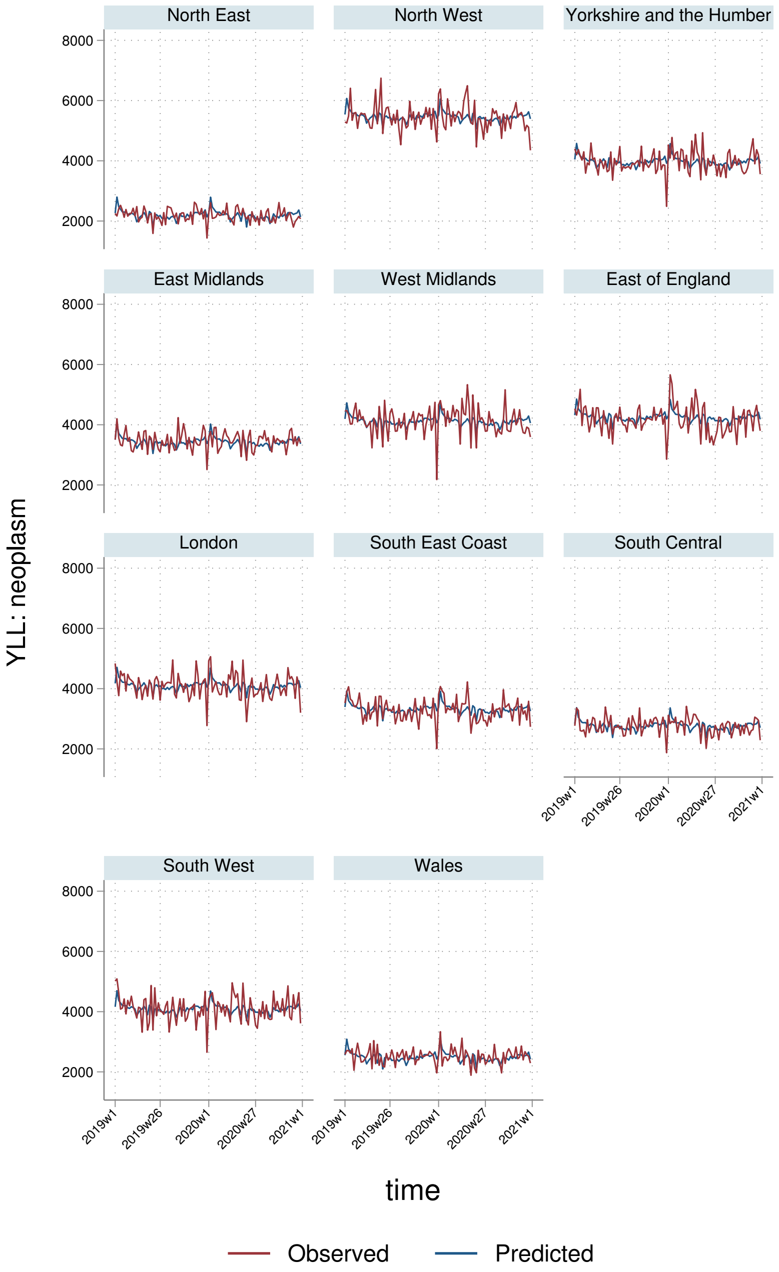
North East
North West
Yorkshire and the Humber
East Midlands
West Midlands
East of England
London
South East Coast
South Central
South West
Wales
YLL: neoplasm
8000
6000
4000
2000
2019w1
2019w26
2020w1
2020w27
2021w1
time
Observed
Predicted

#### 8.3 YLLs per $100,000$ population

##### 8.3.1 England-Wales aggregate

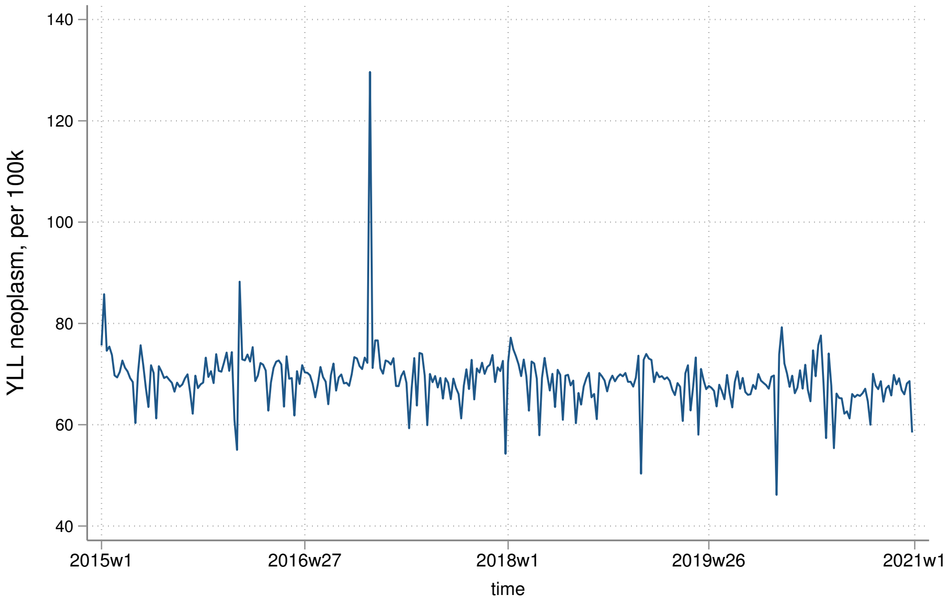


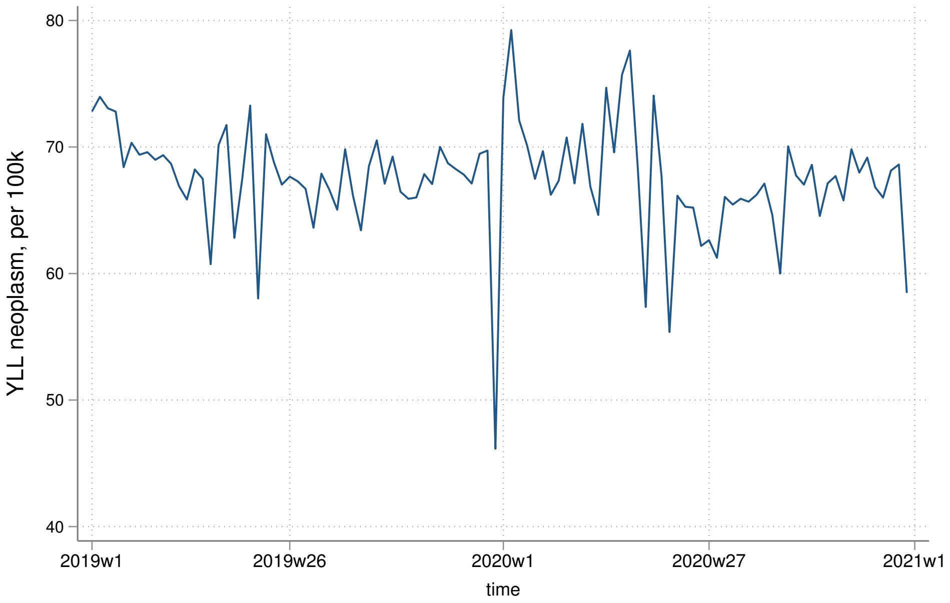

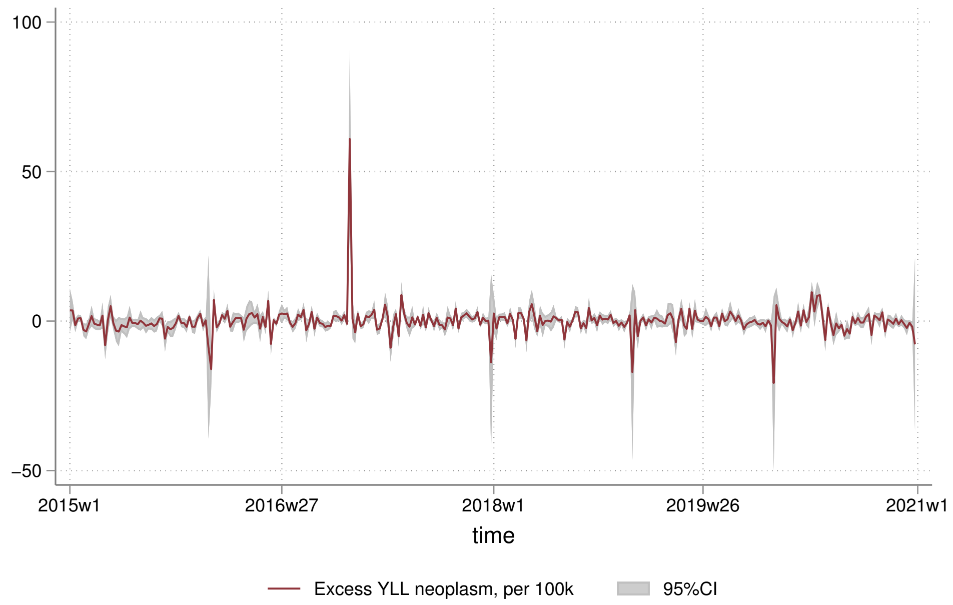
100
50
0
−50
2015w1
2016w27
2018w1
2019w26
2021w1
time
Excess YLL neoplasm, per 100k
95%CI

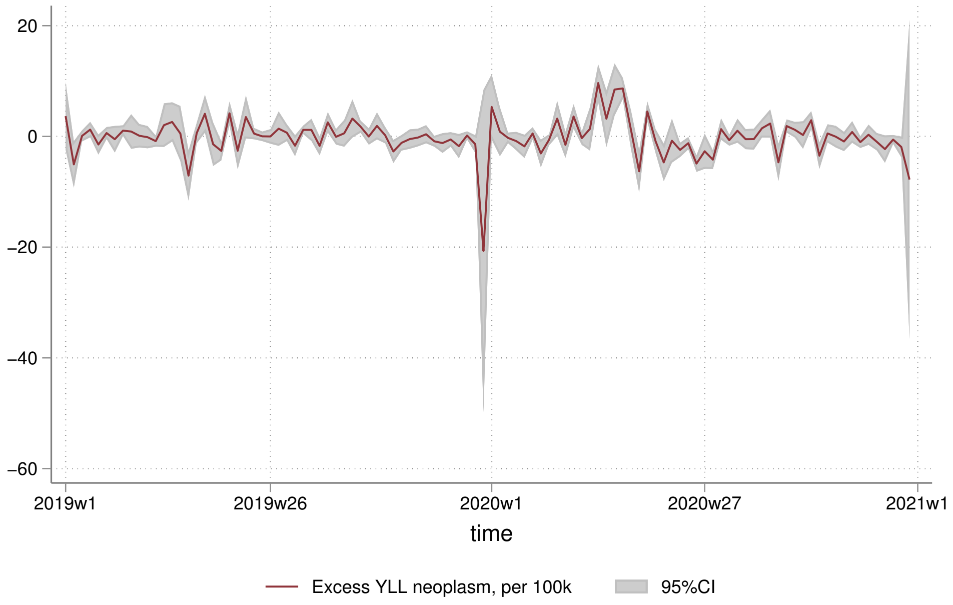
20
0
−20
−40
−60
2019w1
2019w26
2020w1
2020w27
2021w1
time
Excess YLL neoplasm, per 100k
95%CI

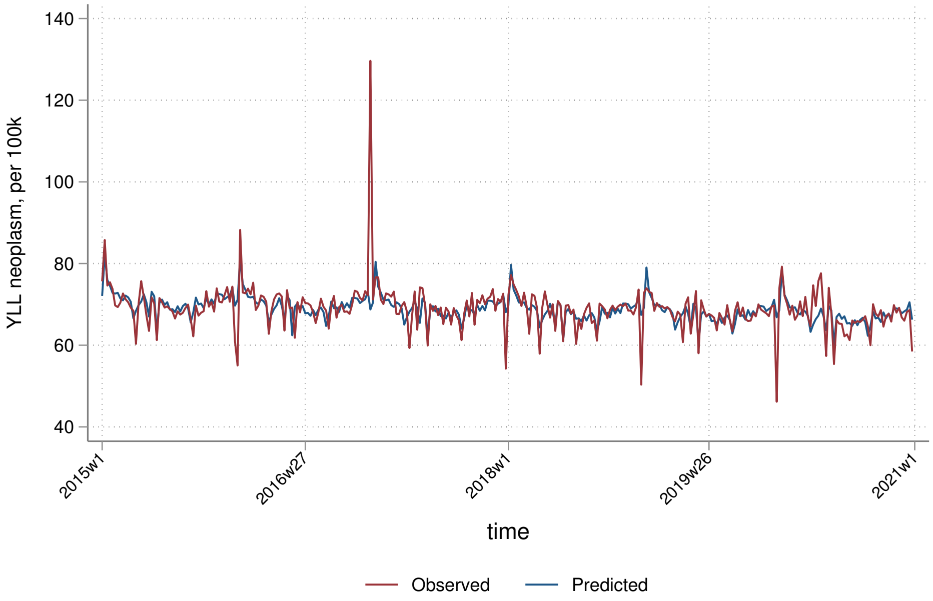
140
120
100
80
60
40
YLL neoplasm, per 100k
2015w1
2016w27
2018w1
2019w26
2021w1
time
Observed
Predicted

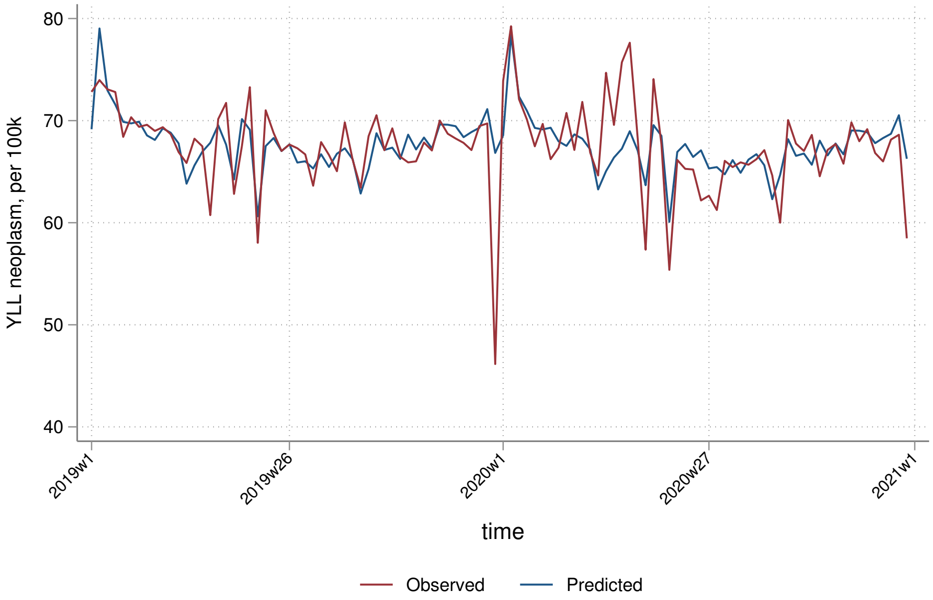
80
70
60
50
40
YLL neoplasm, per 100k
2019w1
2019w26
2020w1
2020w27
2021w1
time
Observed
Predicted

##### 8.3.2 By sex

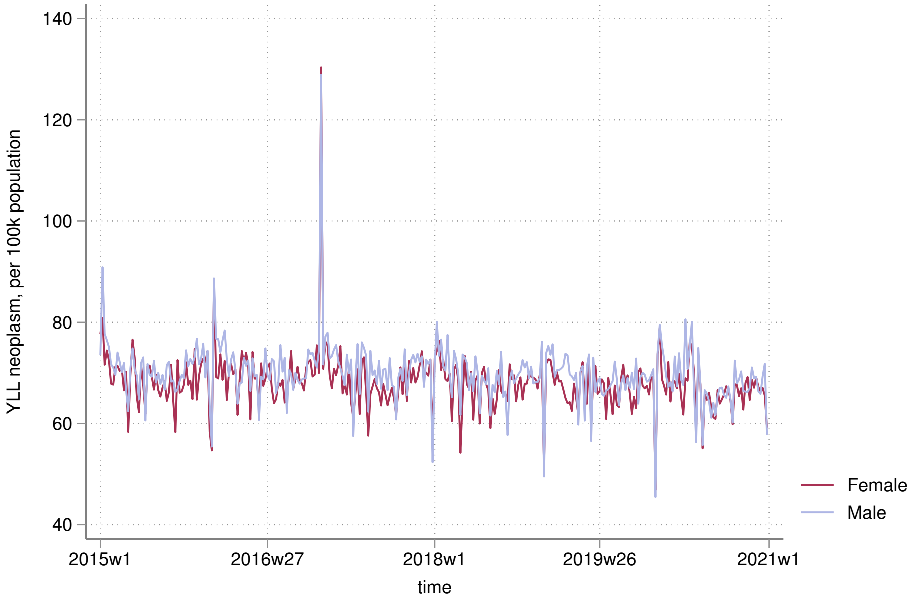

YLL neoplasm, per 100k population
140
120
100
80
60
40
2015w1
2016w27
2018w1
2019w26
2021w1
time
Female
Male


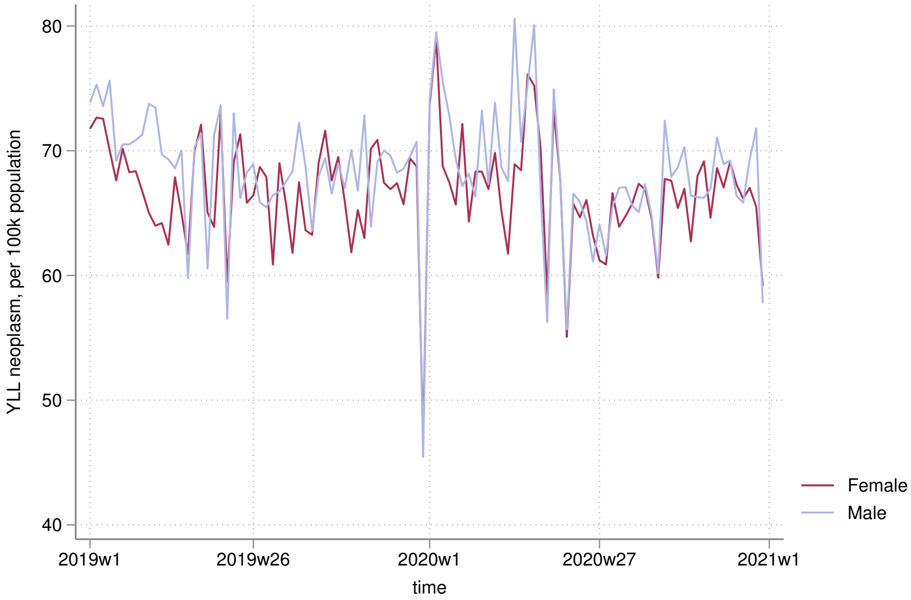

YLL neoplasm, per 100k population
80
70
60
50
40
2019w1
2019w26
2020w1
2020w27
2021w1
time
Female
Male

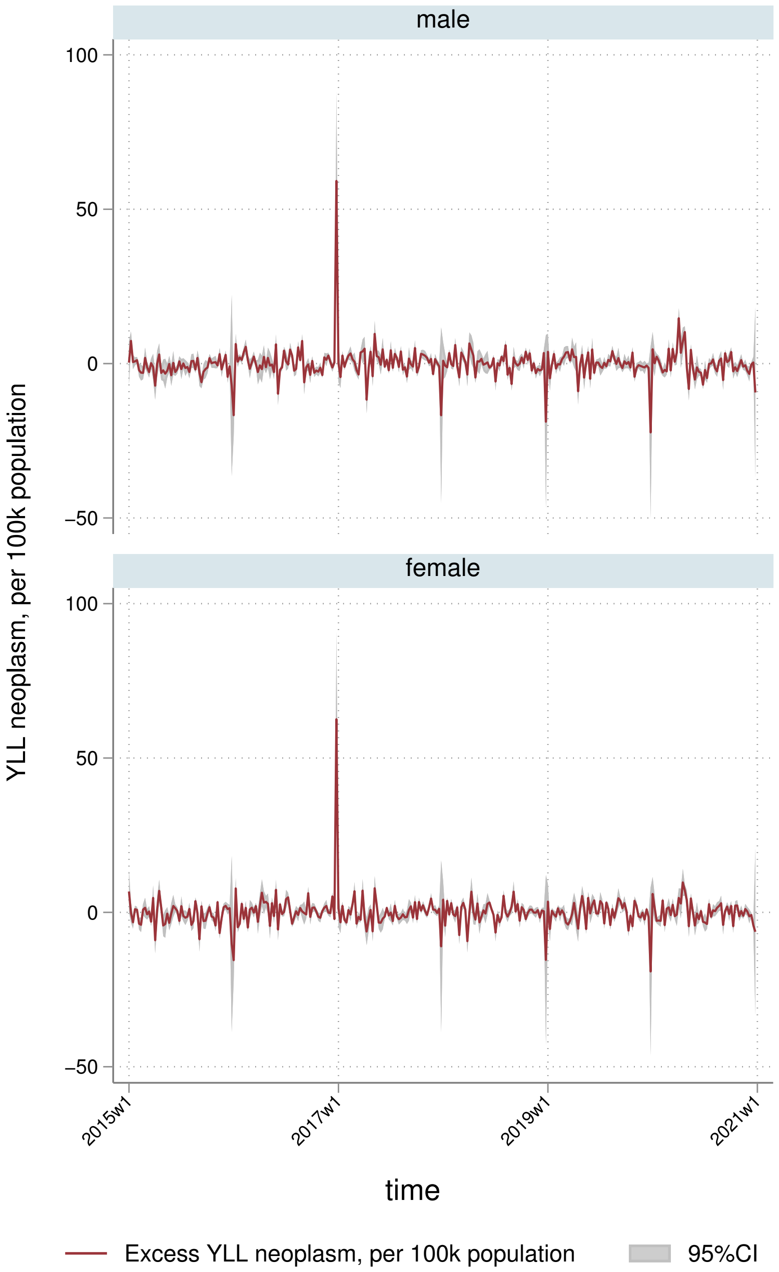

male
female
100
50
0
−50
YLL neoplasm, per 100k population
2015w1
2017w1
2019w1
2021w1
time
Excess YLL neoplasm, per 100k population
95%CI

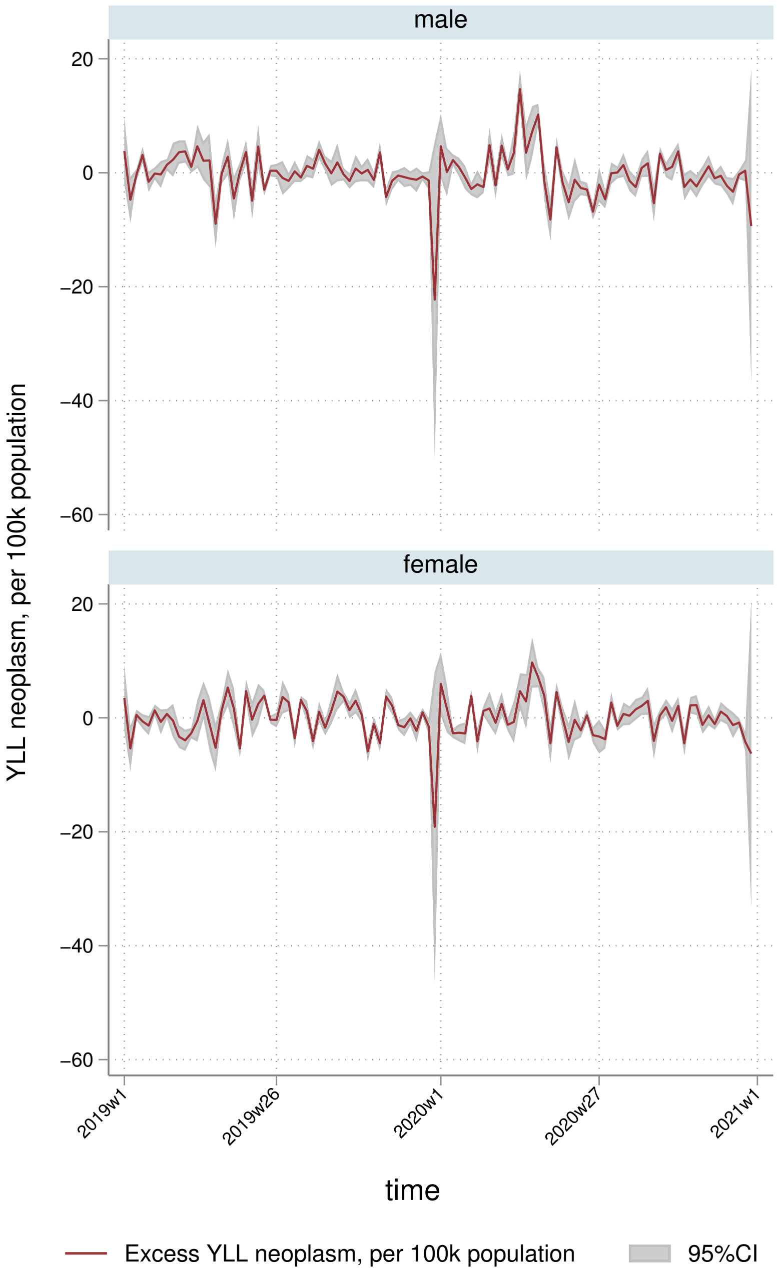
male
female
20
0
−20
−40
−60
YLL neoplasm, per 100k population
2019w1
2019w26
2020w1
2020w27
2021w1
time
Excess YLL neoplasm, per 100k population
95%CI

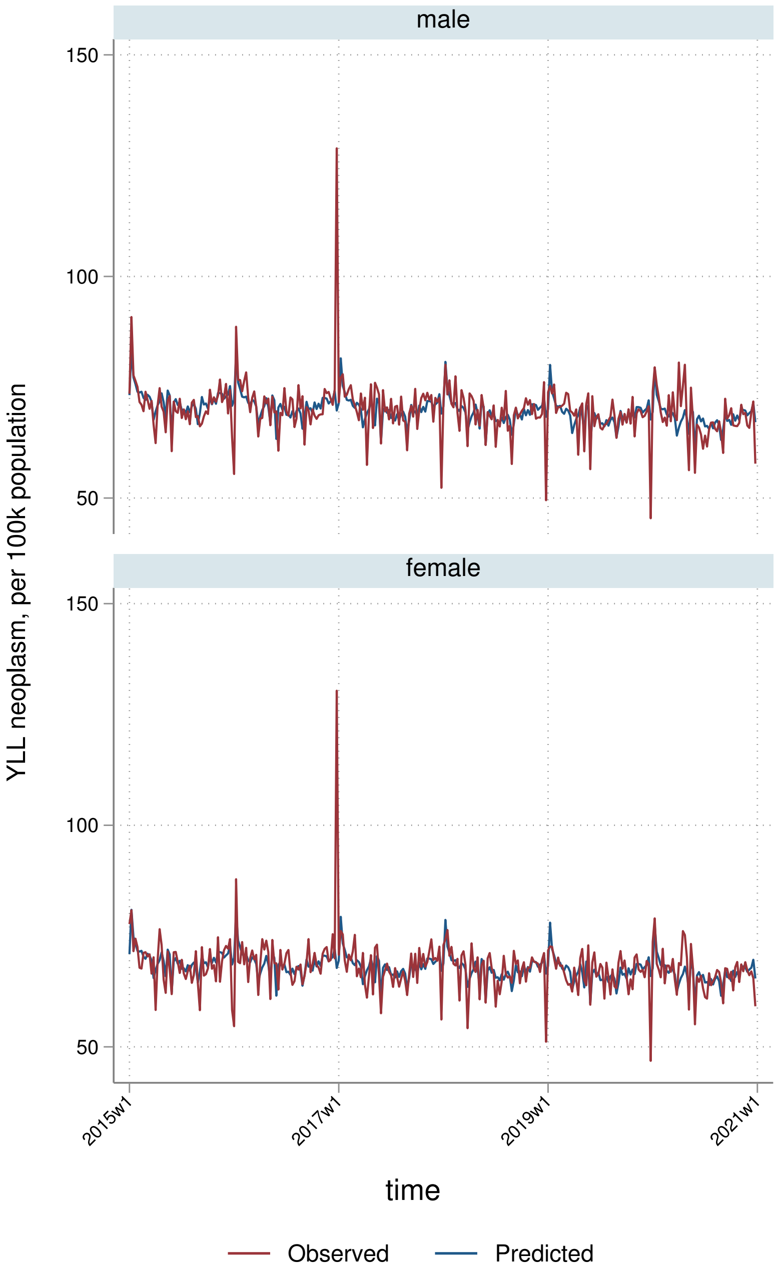
male
female
YLL neoplasm, per 100k population
150
100
50
2015w1
2017w1
2019w1
2021w1
time
Observed
Predicted

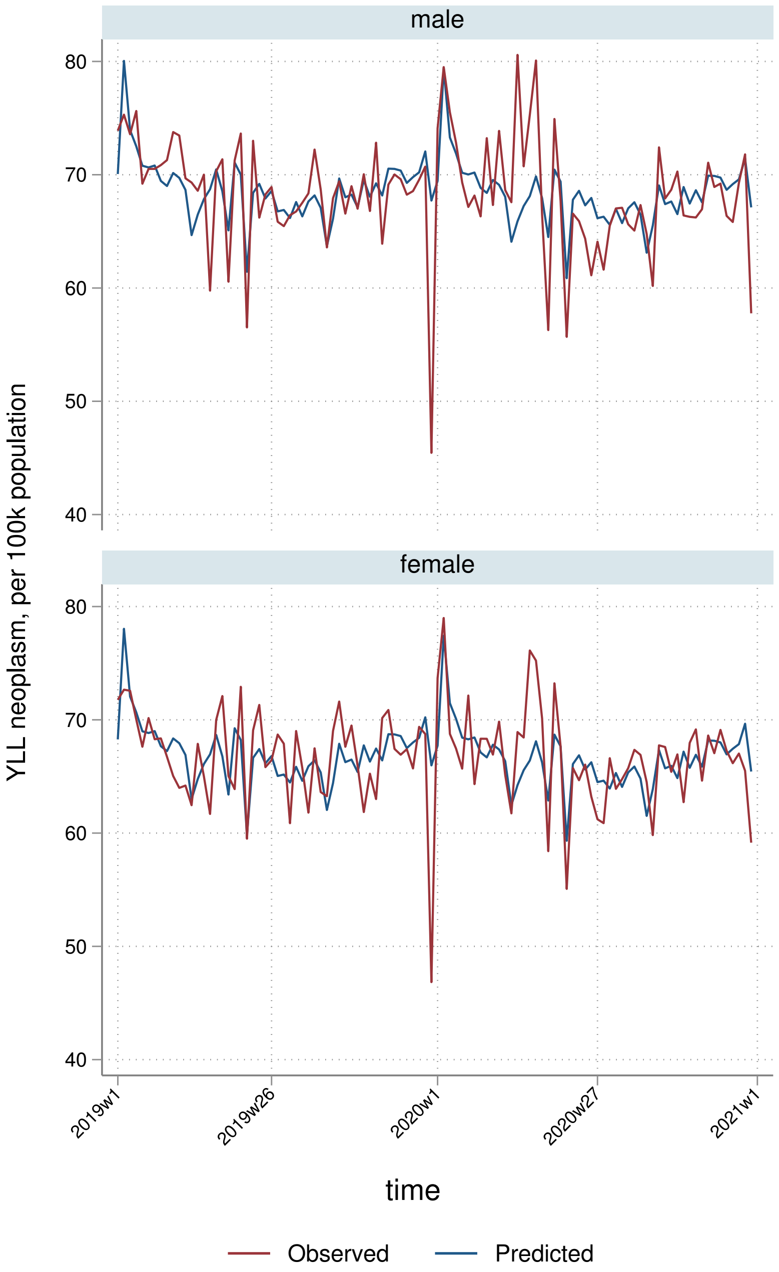

male
female
80
70
60
50
40
YLL neoplasm, per 100k population
2019w1
2019w26
2020w1
2020w27
2021w1
time
Observed
Predicted

##### 8.3.3 By deprivation quintile

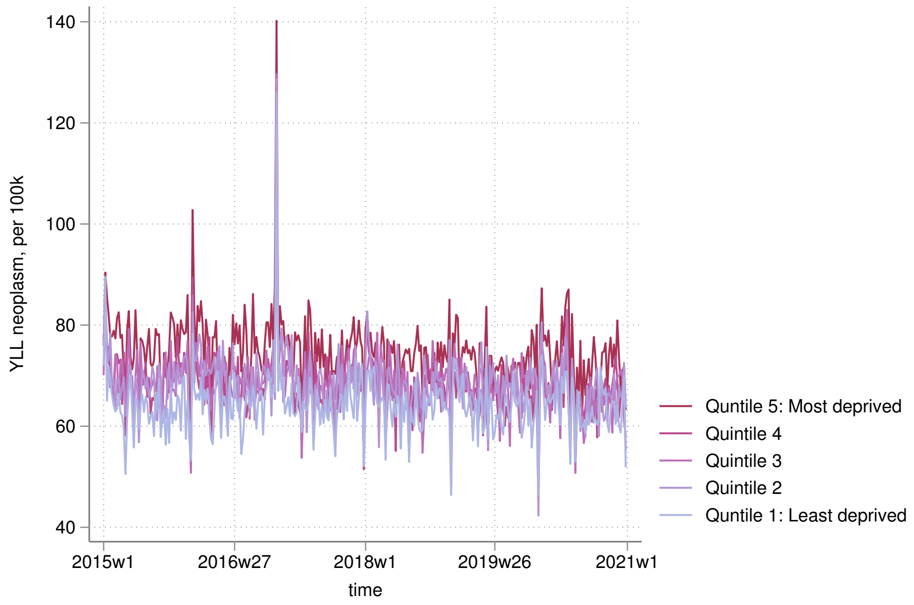


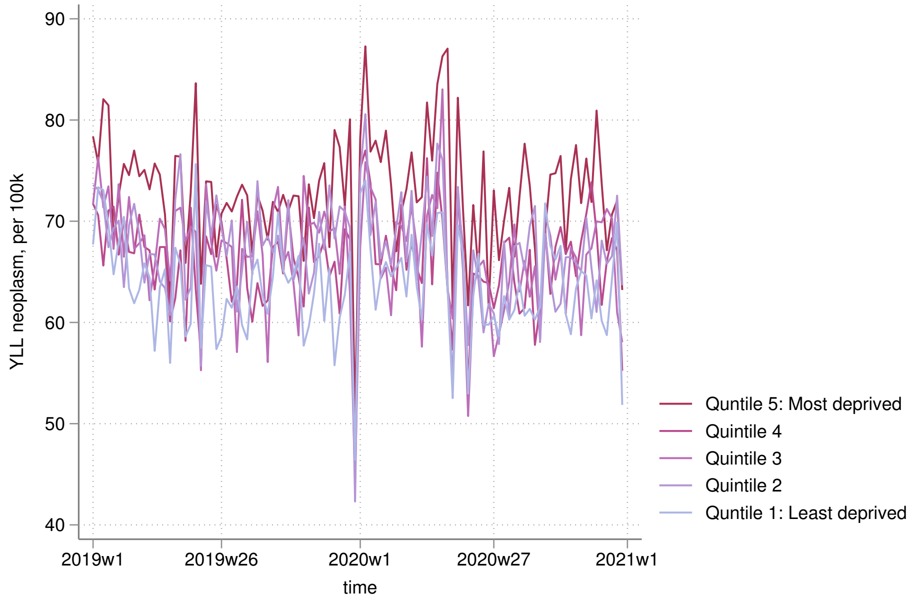

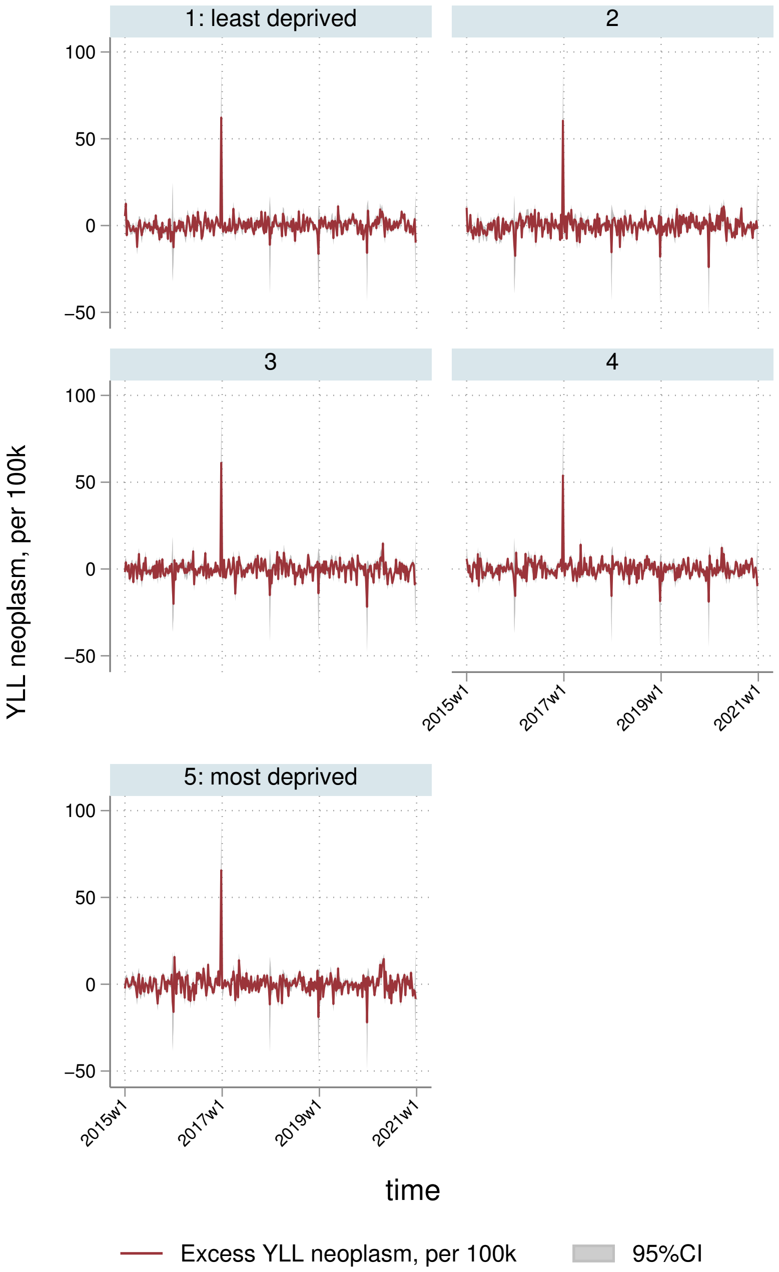
1: least deprived
2
3
4
5: most deprived
100
50
0
−50
2015w1
2017w1
2019w1
2021w1
YLL neoplasm, per 100k
time
Excess YLL neoplasm, per 100k
95%CI

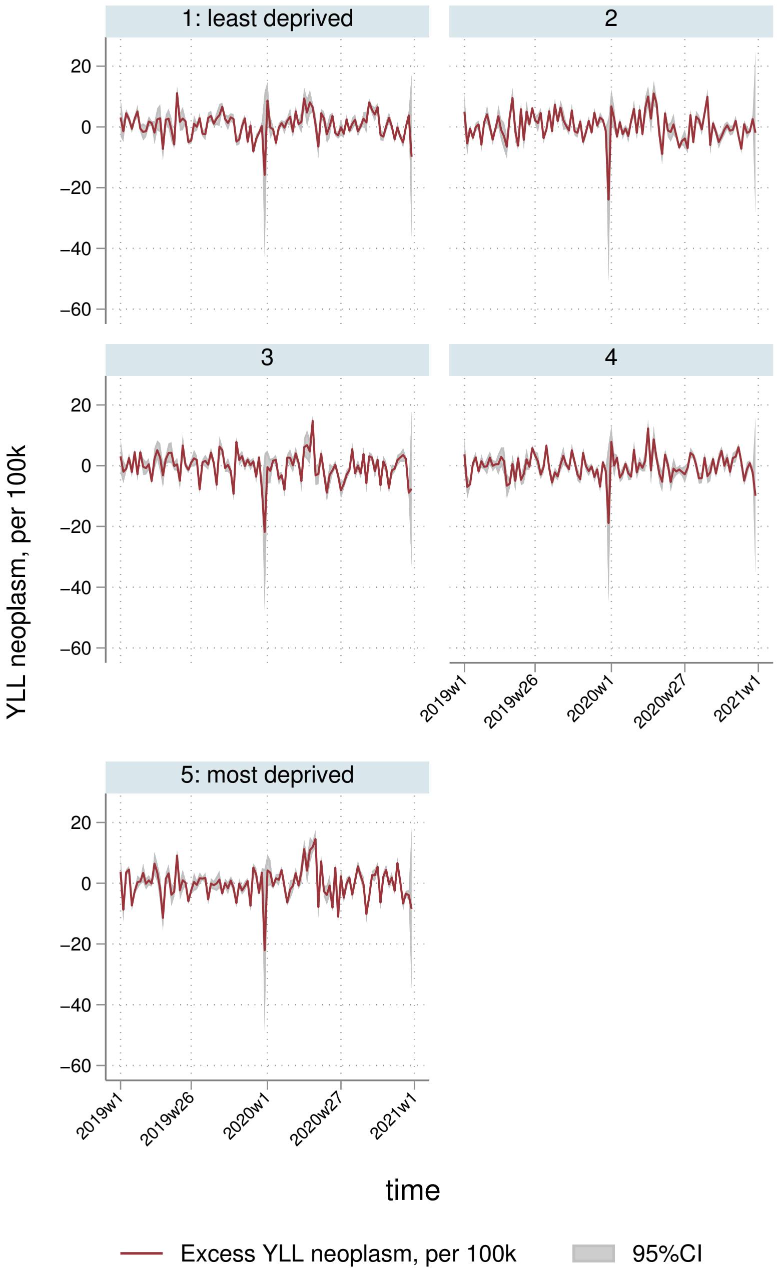

1: least deprived
2
3
4
5: most deprived
20
0
−20
−40
−60
YLL neoplasm, per 100k
2019w1
2019w26
2020w1
2020w27
2021w1
time
Excess YLL neoplasm, per 100k
95%CI

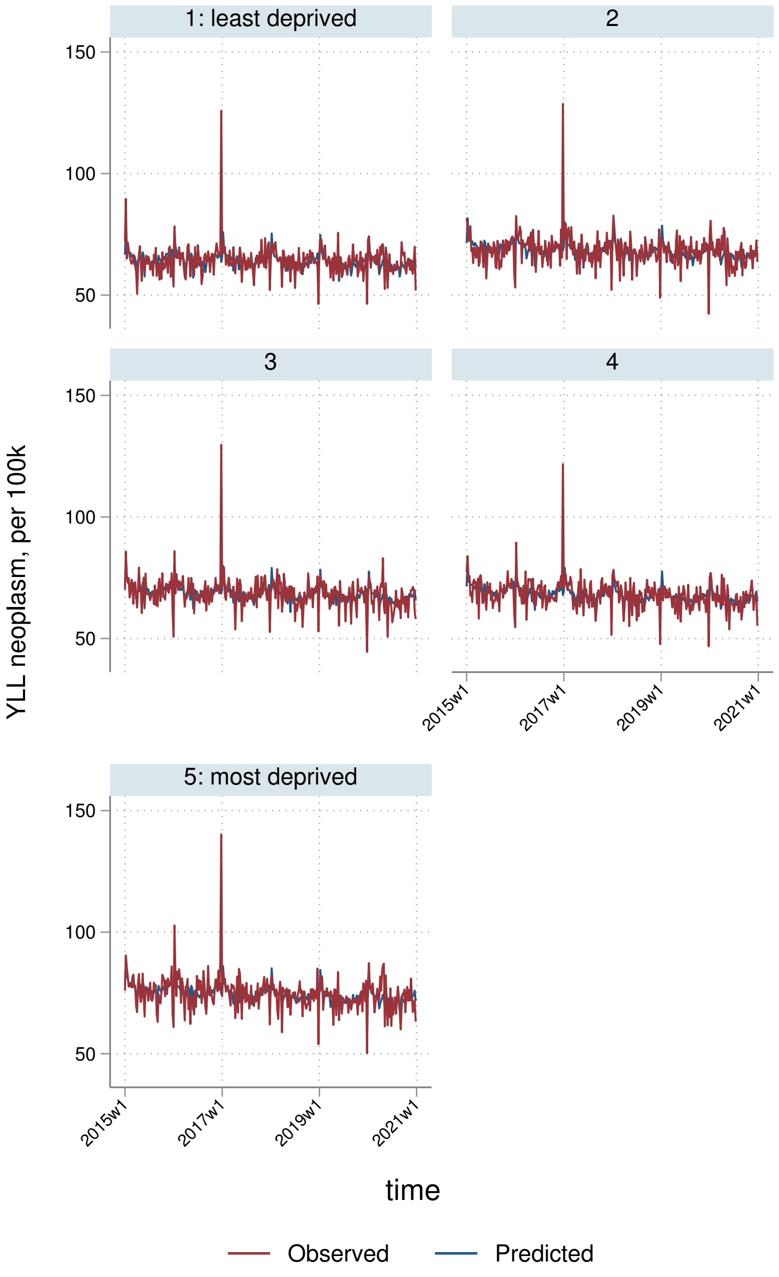
1: least deprived
2
3
4
5: most deprived
150
100
50
YLL neoplasm, per 100k
2015w1
2017w1
2019w1
2021w1
time
Observed
Predicted

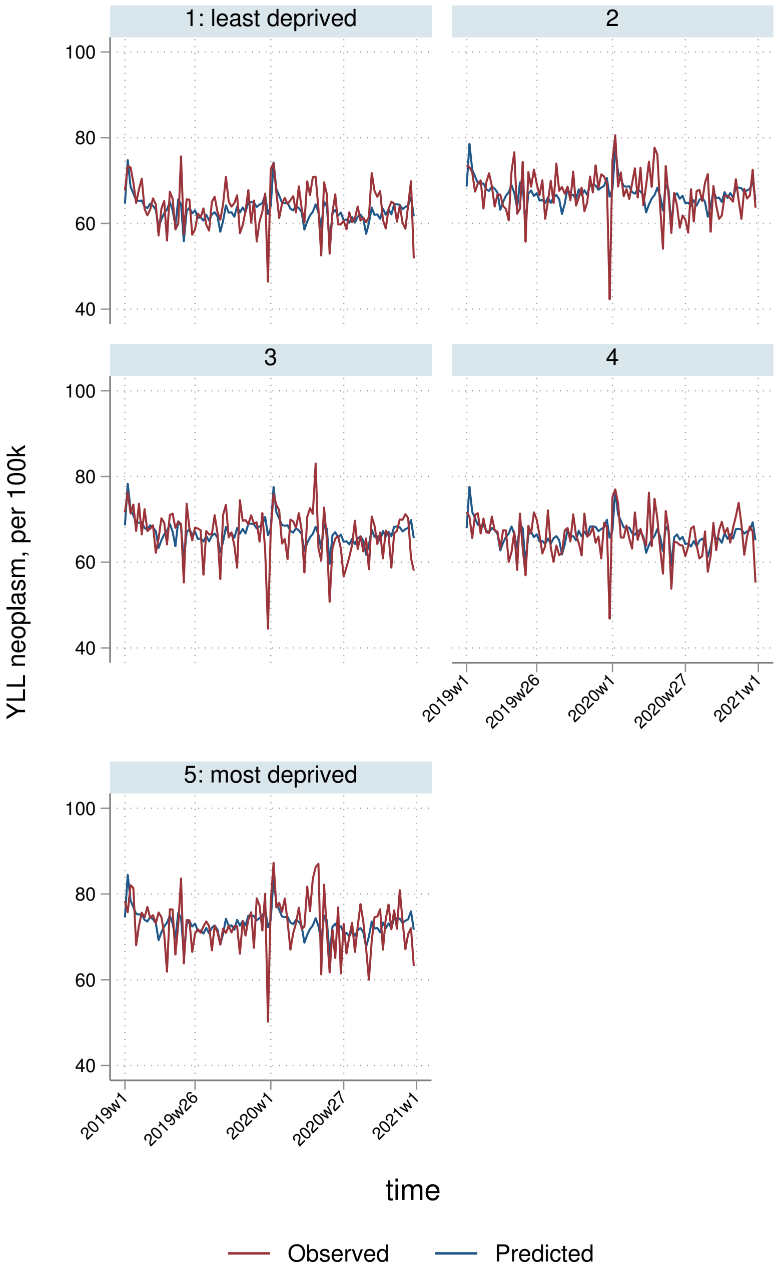

1: least deprived
2
3
4
5: most deprived
100
80
60
40
YLL neoplasm, per 100k
2019w1
2019w26
2020w1
2020w27
2021w1
time
Observed
Predicted

##### 8.3.4 By Strategic Health Authority

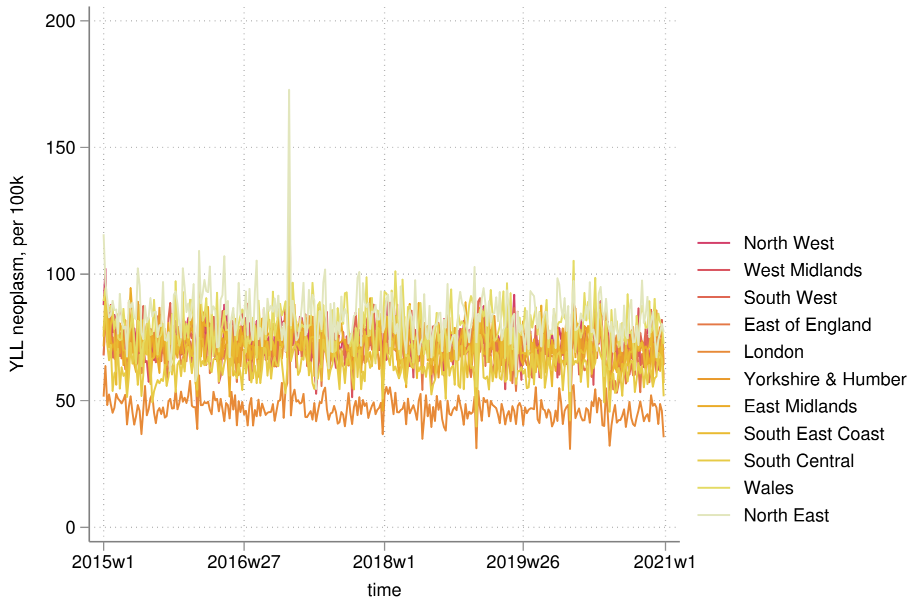


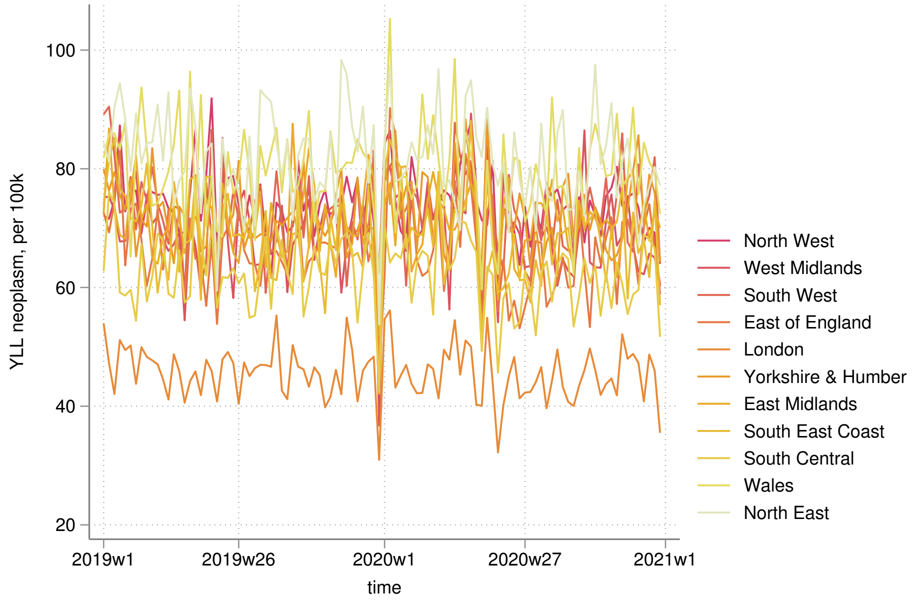

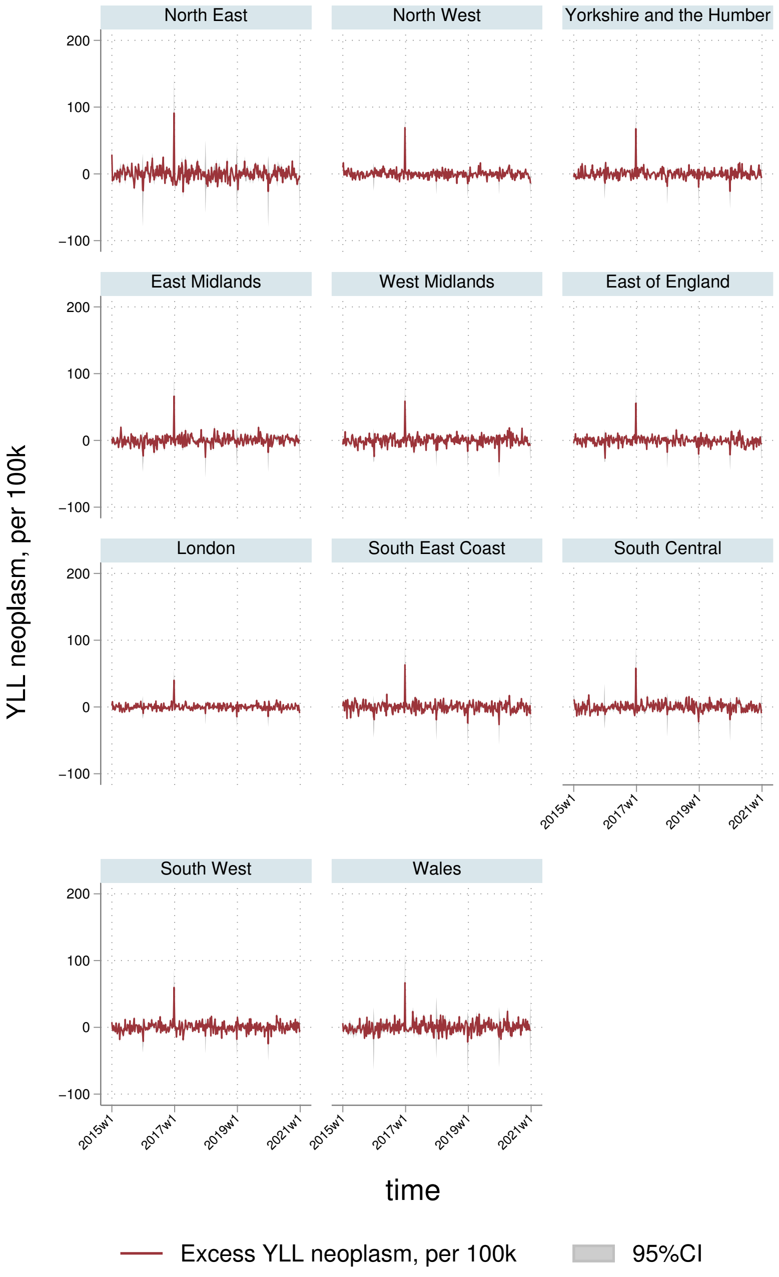
North East
North West
Yorkshire and the Humber
East Midlands
West Midlands
East of England
London
South East Coast
South Central
South West
Wales
YLL neoplasm, per 100k
200
100
0
−100
2015w1
2017w1
2019w1
2021w1
time
Excess YLL neoplasm, per 100k
95%CI

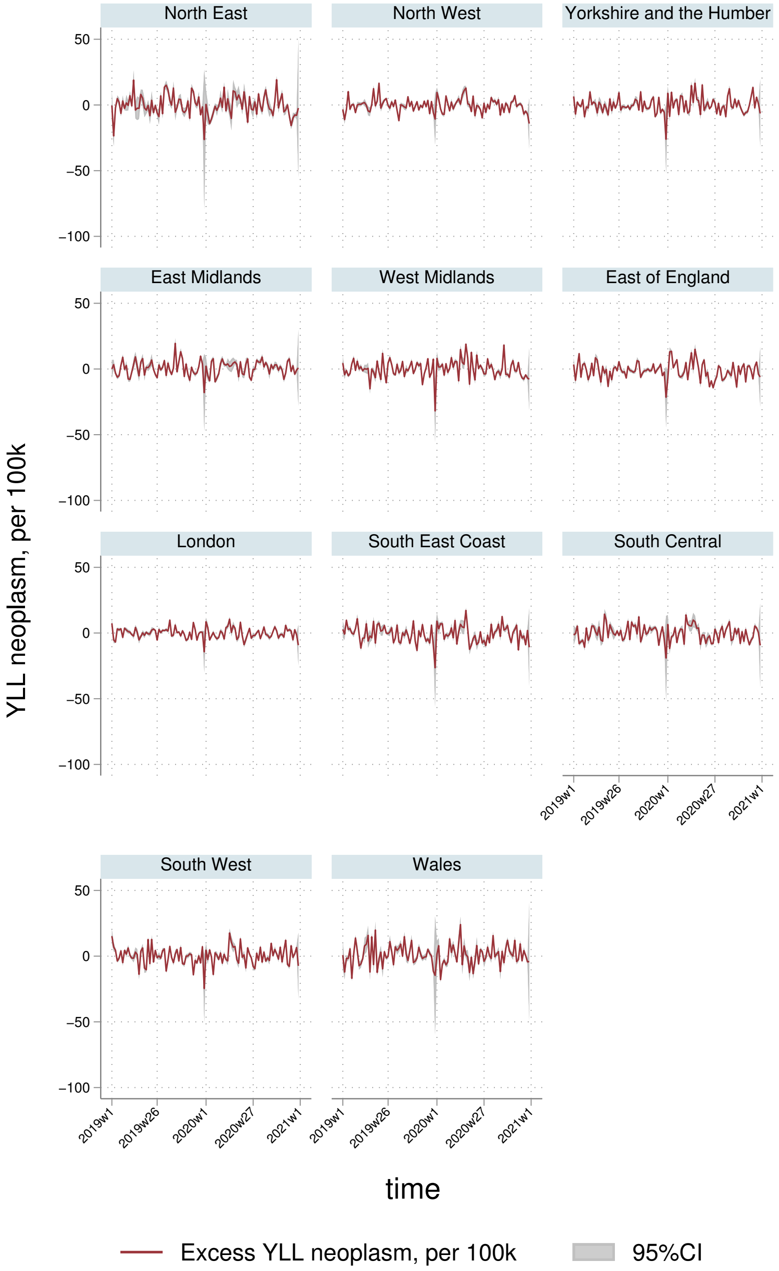

North East
North West
Yorkshire and the Humber
East Midlands
West Midlands
East of England
London
South East Coast
South Central
South West
Wales
YLL neoplasm, per 100k
50
0
−50
−100
2019w1
2019w26
2020w1
2020w27
2021w1
time
Excess YLL neoplasm, per 100k
95%CI

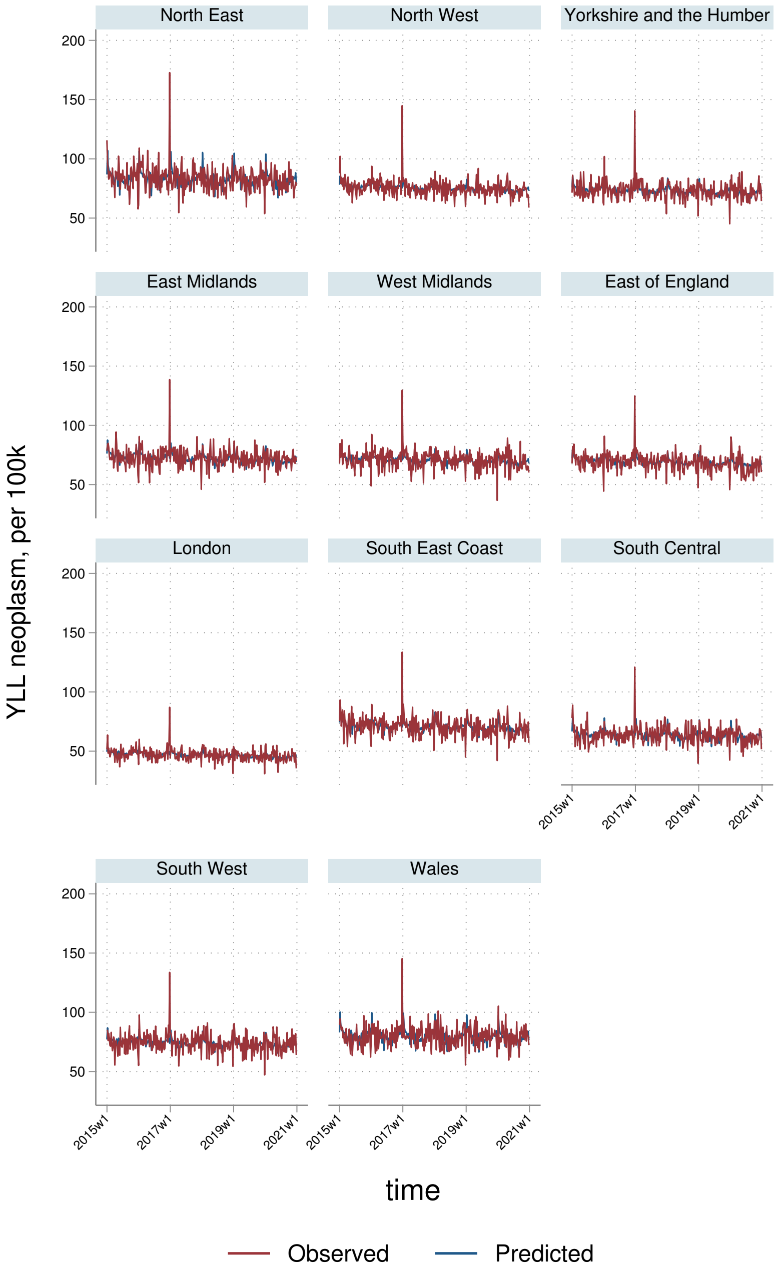
North East
North West
Yorkshire and the Humber
East Midlands
West Midlands
East of England
London
South East Coast
South Central
South West
Wales
YLL neoplasm, per 100k
200
150
100
50
2015w1
2017w1
2019w1
2021w1
time
Observed
Predicted

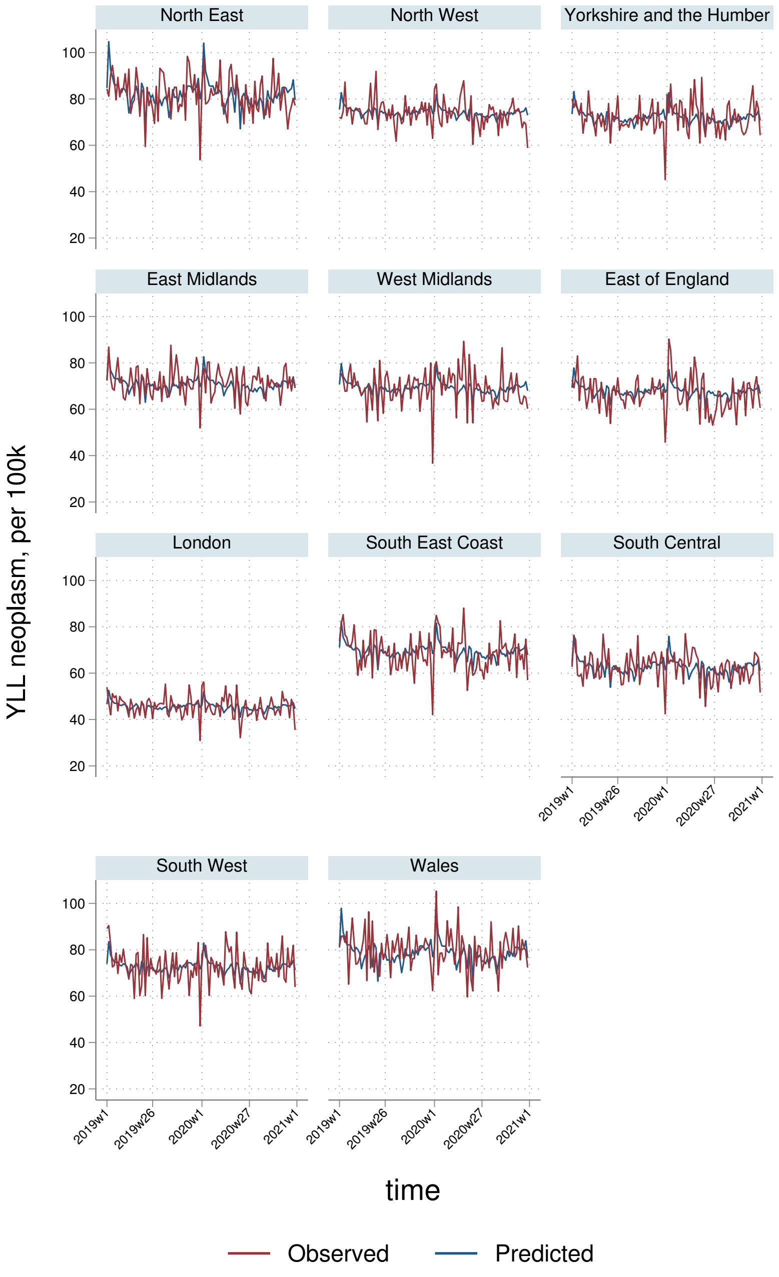

North East
North West
Yorkshire and the Humber
East Midlands
West Midlands
East of England
London
South East Coast
South Central
South West
Wales
YLL neoplasm, per 100k
100
80
60
40
20
2019w1
2019w26
2020w1
2020w27
2021w1
time
Observed
Predicted

### 9 All other indirect

#### 9.1 AASMRs

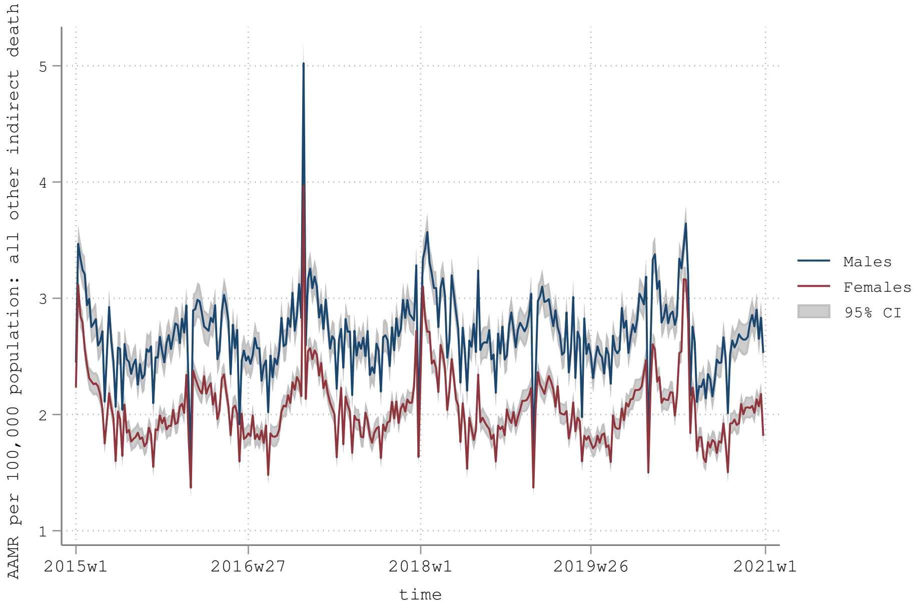


AAMR: age-adjusted mortality rate using the 2020-2025 WHO reference population (see https://www.who.int/healthinfo/paper31.pdf)

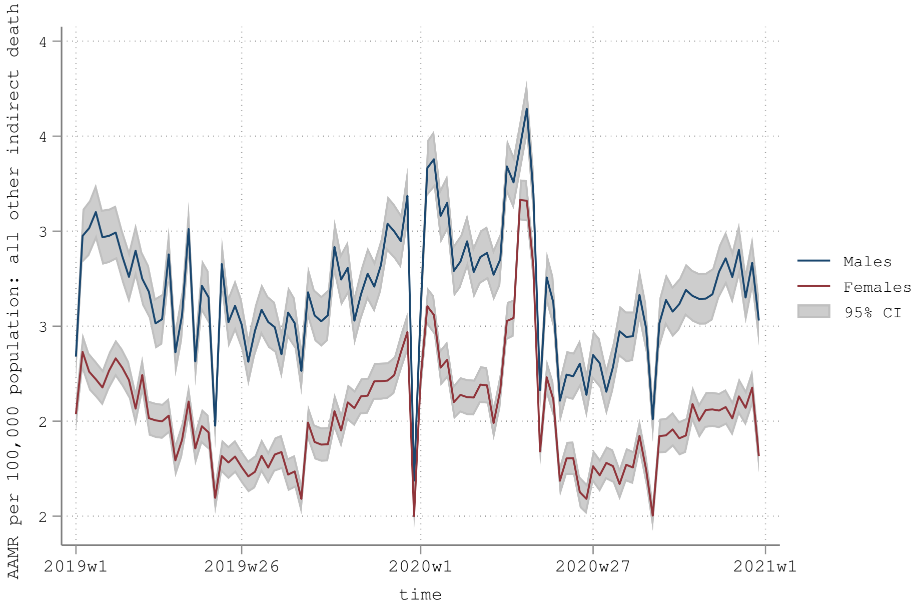


AAMR: age-adjusted mortality rate using the 2020-2025 WHO reference population (see https://www.who.int/healthinfo/paper31.pdf)

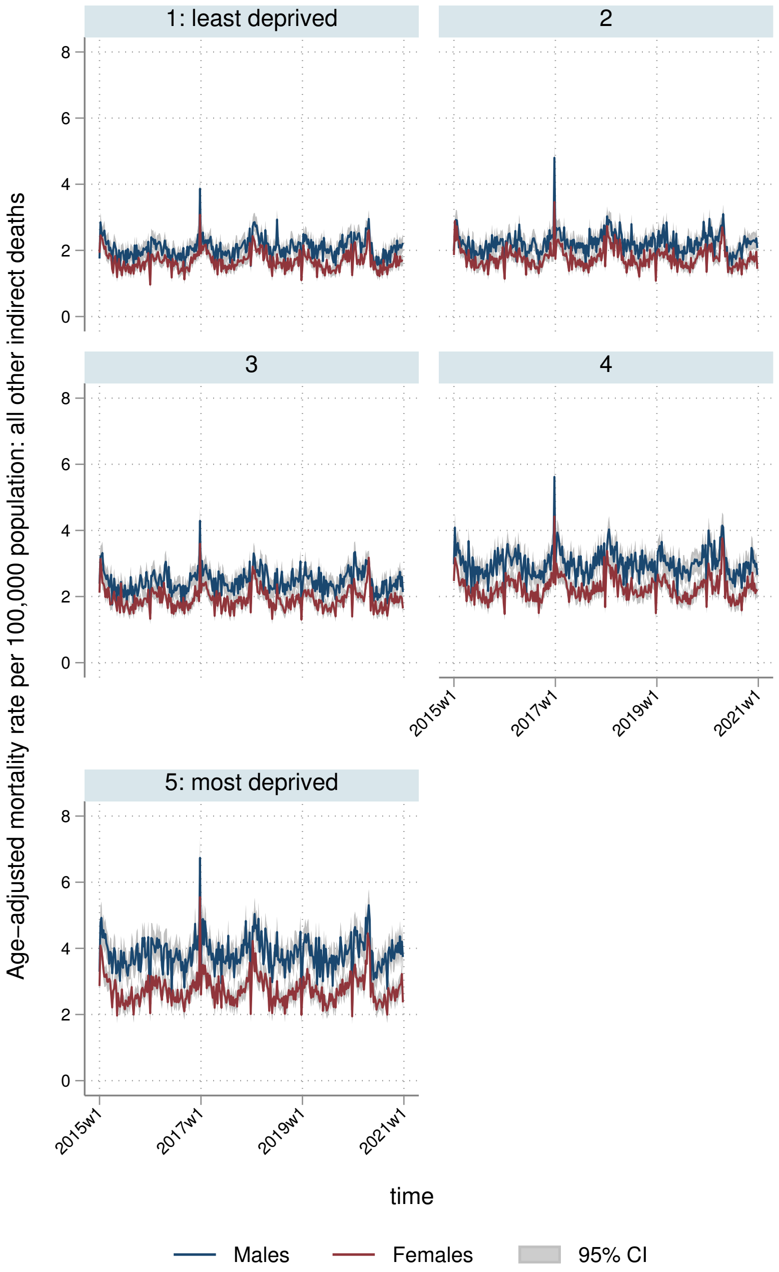
1: least deprived
2
3
4
5: most deprived
Age-adjusted mortality rate per 100,000 population: all other indirect deaths
0
2
4
6
8
2015w1
2017w1
2019w1
2021w1
time
Males
Females
95% CI

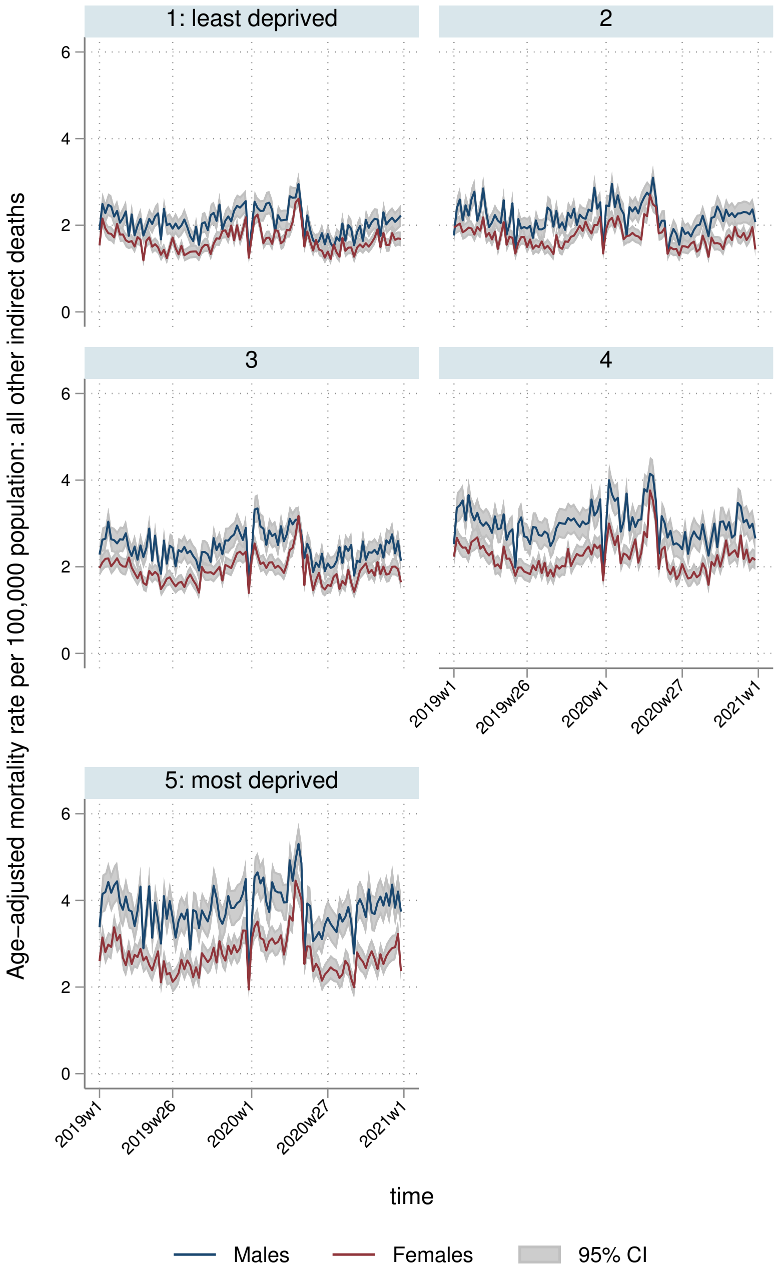
1: least deprived
2
3
4
5: most deprived
Age-adjusted mortality rate per 100,000 population: all other indirect deaths
0
2
4
6
2019w1
2019w26
2020w1
2020w27
2021w1
time
Males
Females
95% CI

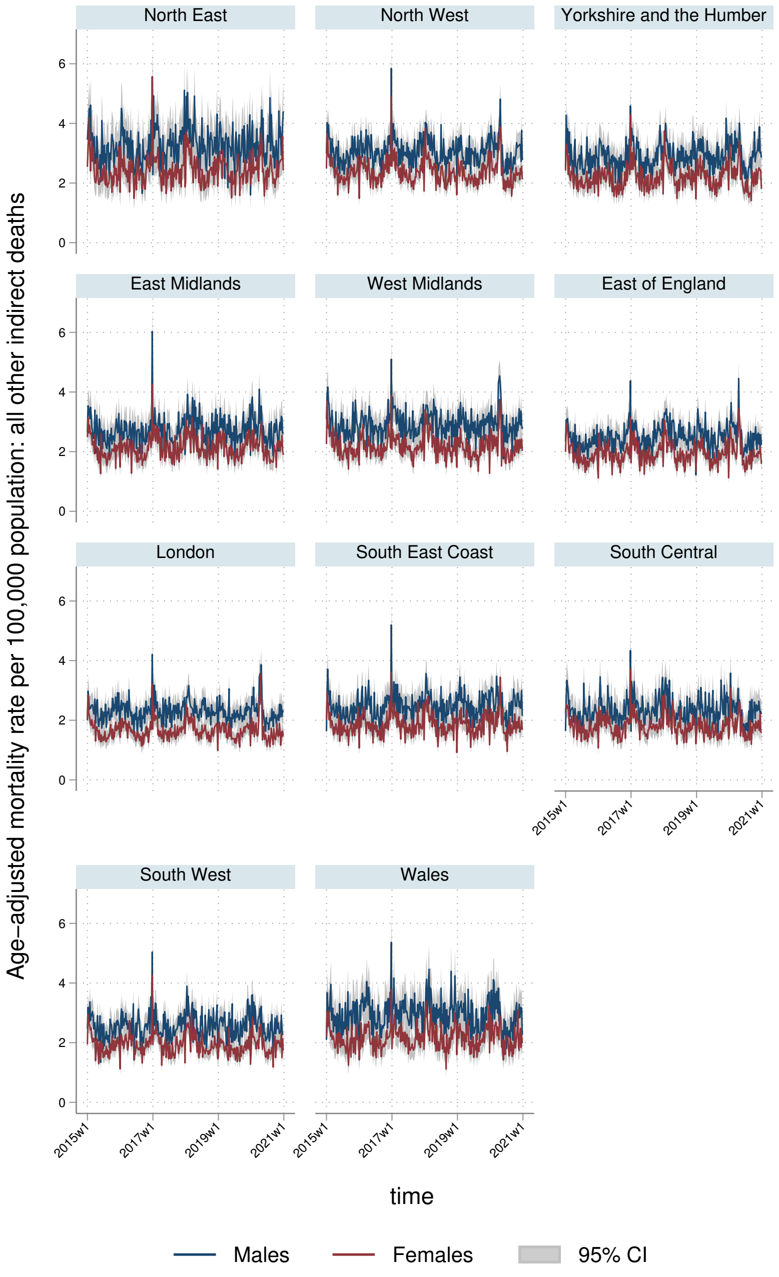
North East
North West
Yorkshire and the Humber
East Midlands
West Midlands
East of England
London
South East Coast
South Central
South West
Wales
Age-adjusted mortality rate per 100,000 population: all other indirect deaths
0
2
4
6
2015w1
2017w1
2019w1
2021w1
time
Males
Females
95% CI

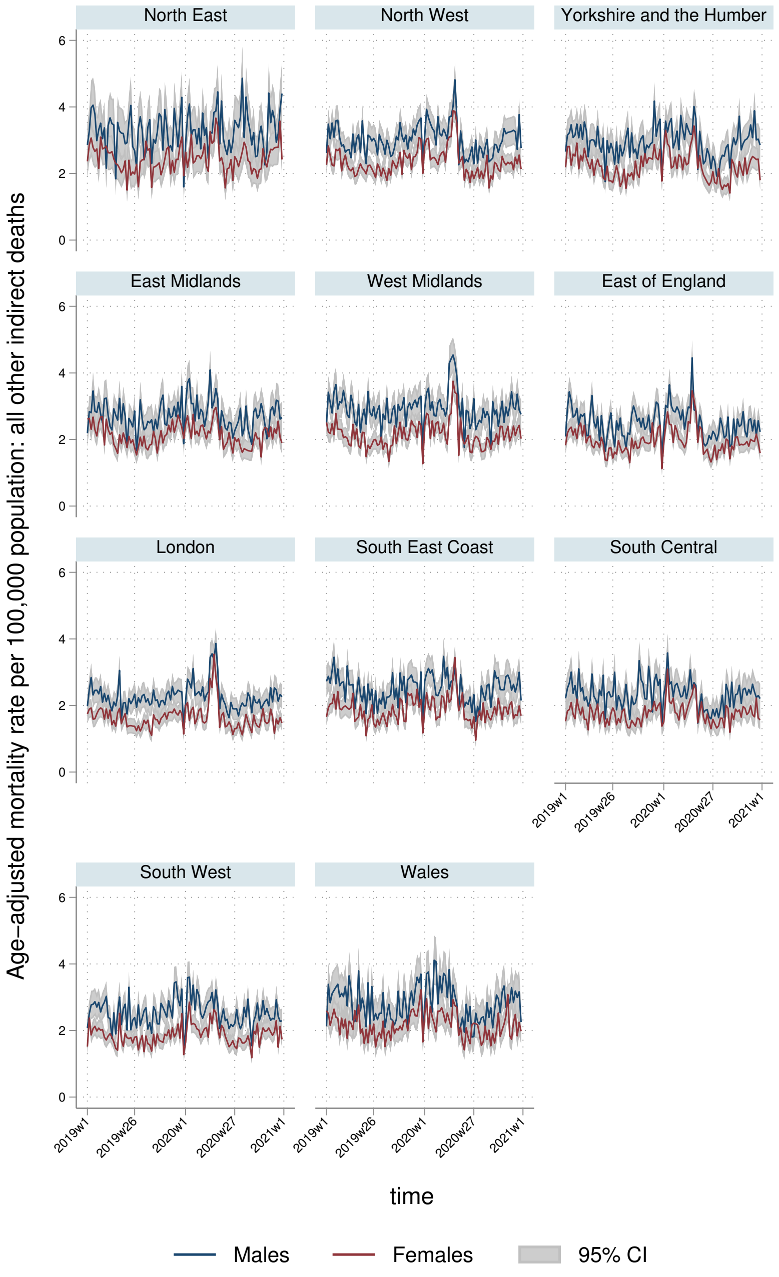
North East
North West
Yorkshire and the Humber
East Midlands
West Midlands
East of England
London
South East Coast
South Central
South West
Wales
Age-adjusted mortality rate per 100,000 population: all other indirect deaths
0
2
4
6
2019w1
2019w26
2020w1
2020w27
2021w1
time
Males
Females
95% CI

#### 9.2 YLLs

##### 9.2.1 England-Wales aggregate

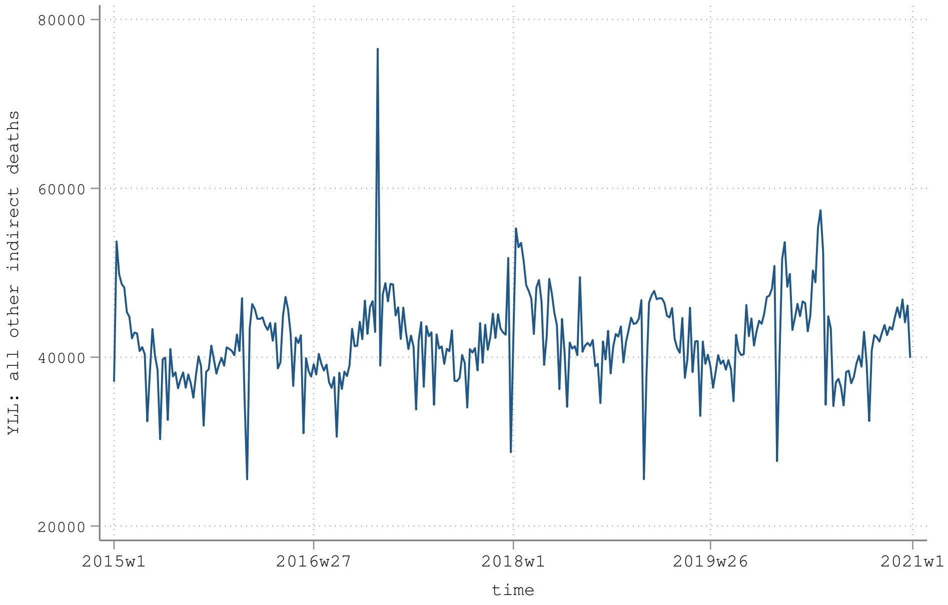


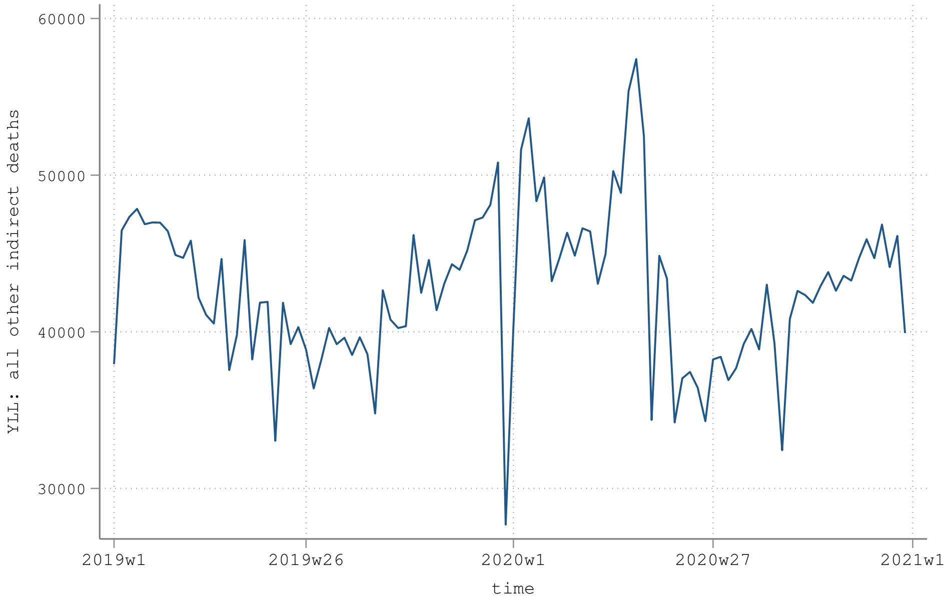

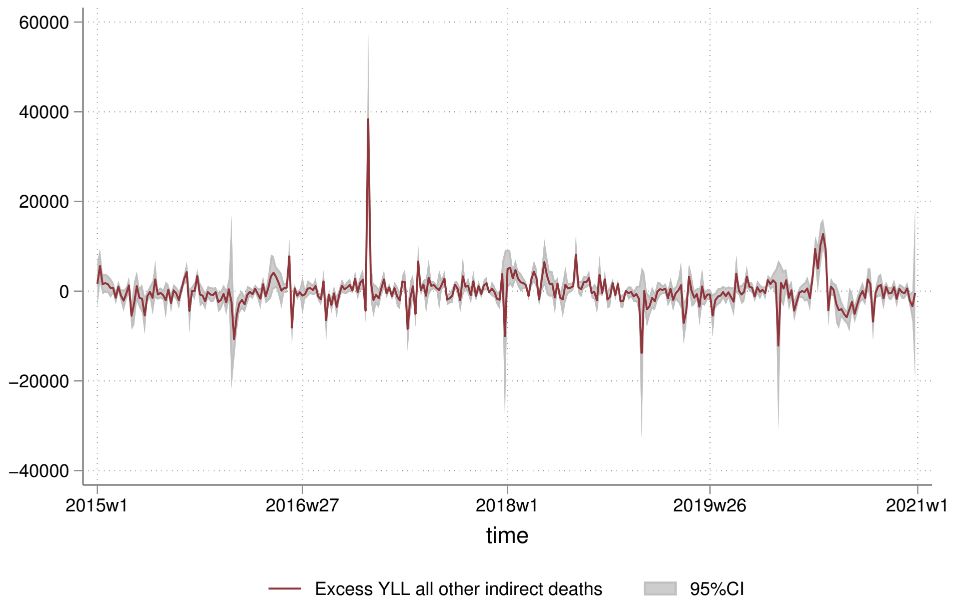
60000
40000
20000
0
−20000
−40000
2015w1
2016w27
2018w1
2019w26
2021w1
time
Excess YLL all other indirect deaths
95%CI

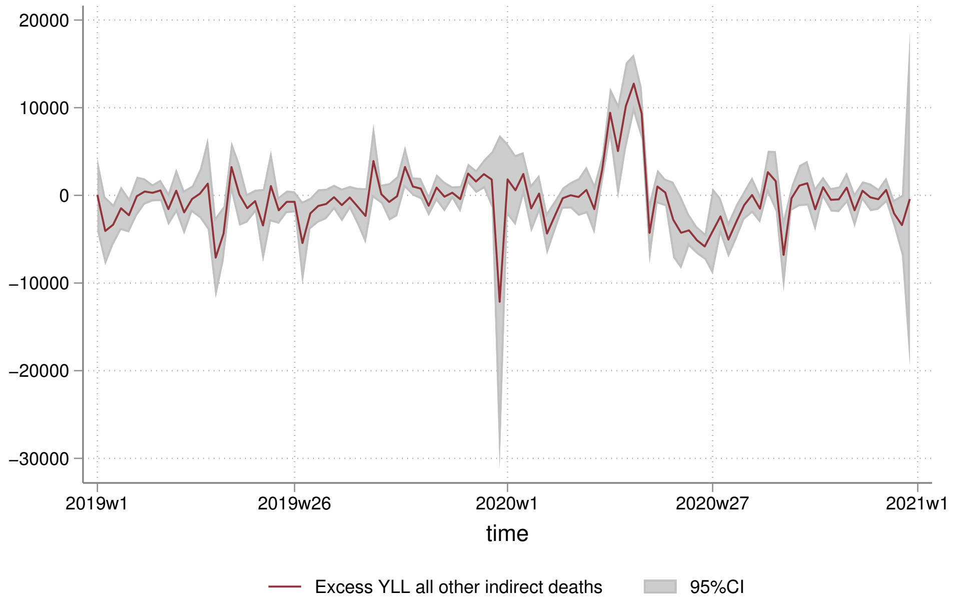
20000
10000
0
−10000
−20000
−30000
2019w1
2019w26
2020w1
2020w27
2021w1
time
Excess YLL all other indirect deaths
95%CI

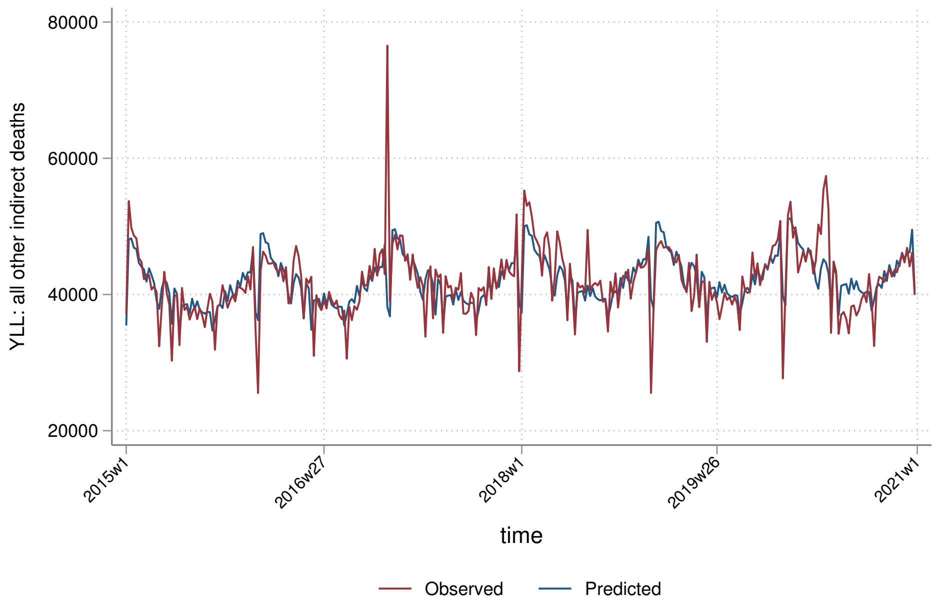
80000
60000
40000
20000
YLL: all other indirect deaths
2015w1
2016w27
2018w1
2019w26
2021w1
time
Observed
Predicted

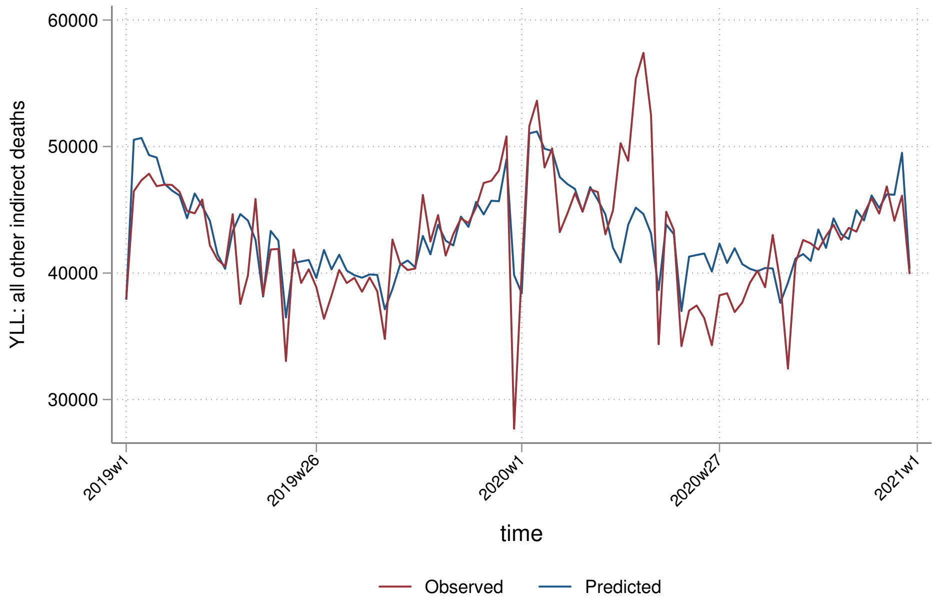
60000
50000
40000
30000
YLL: all other indirect deaths
2019w1
2019w26
2020w1
2020w27
2021w1
time
Observed
Predicted

##### 9.2.2 By sex

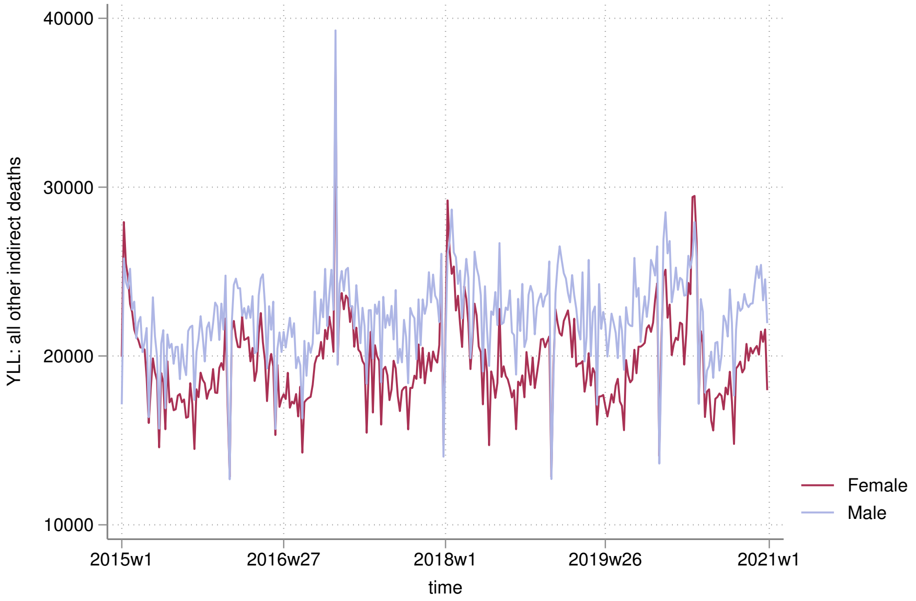

40000
30000
20000
10000
YLL: all other indirect deaths
2015w1
2016w27
2018w1
2019w26
2021w1
time
Female
Male


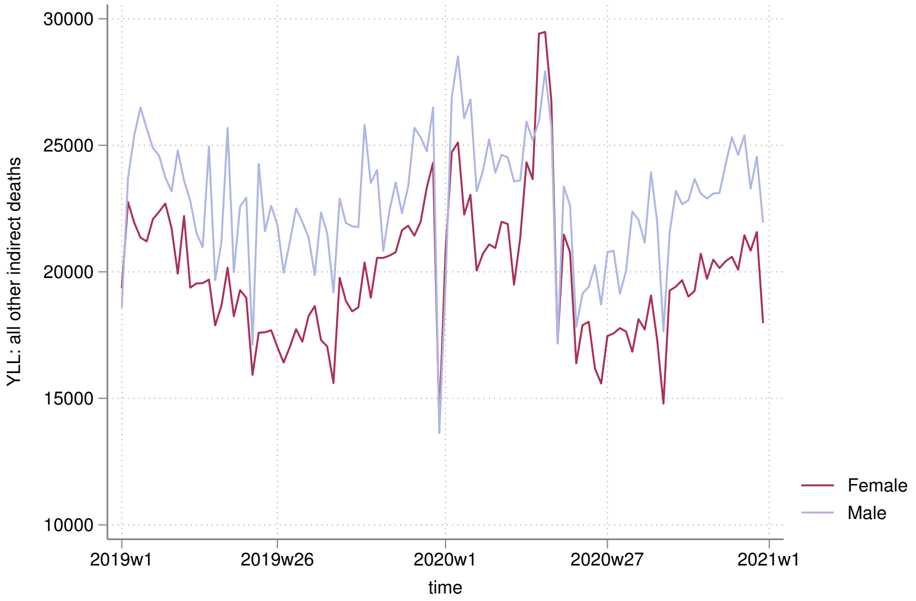

30000
25000
20000
15000
10000
YLL: all other indirect deaths
2019w1
2019w26
2020w1
2020w27
2021w1
time
Female
Male

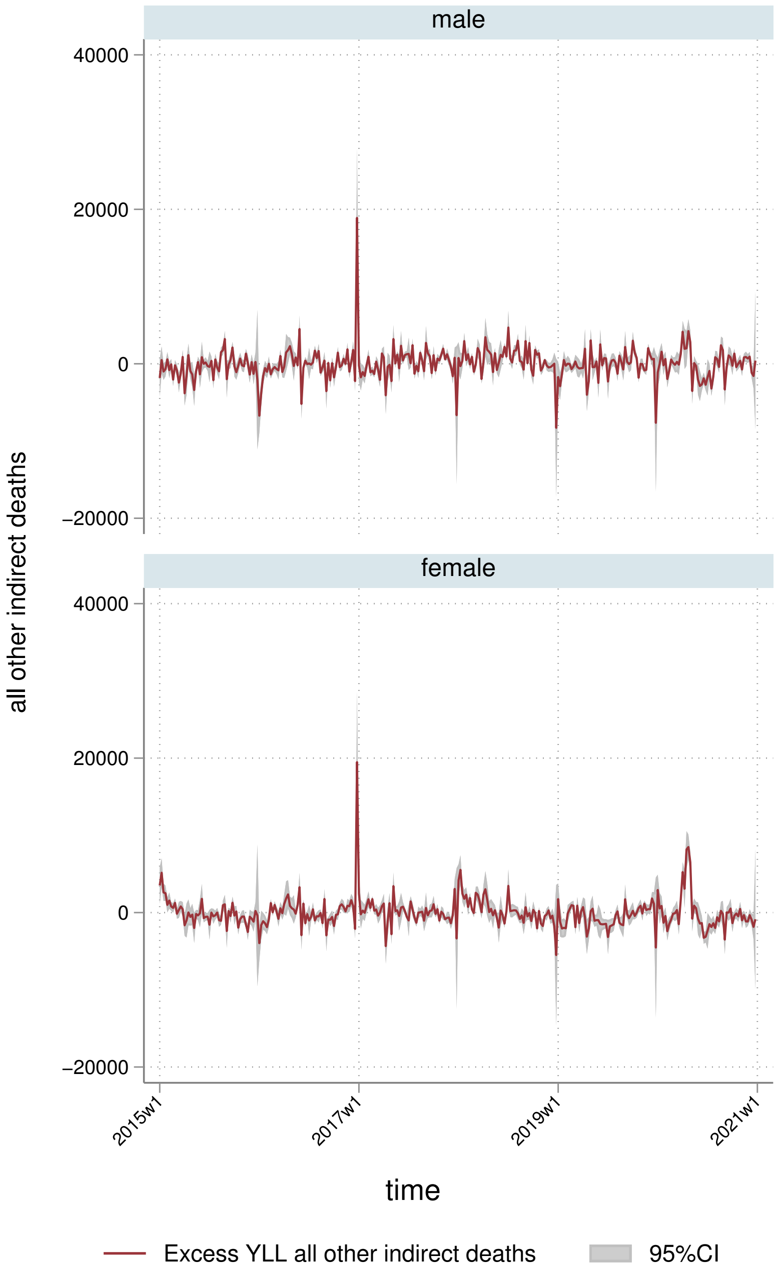
male
female
40000
20000
0
−20000
all other indirect deaths
2015w1
2017w1
2019w1
2021w1
time
Excess YLL all other indirect deaths
95%CI

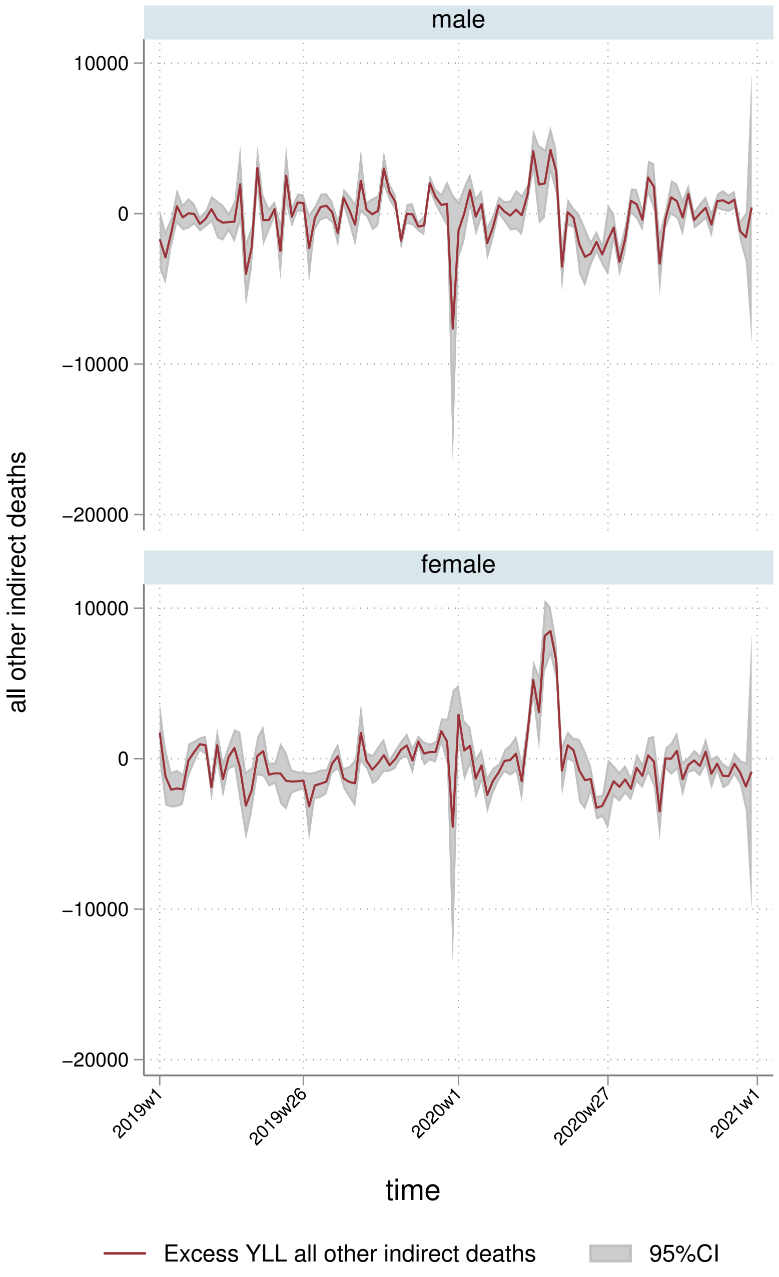
male
female
10000
0
−10000
−20000
all other indirect deaths
2019w1
2019w26
2020w1
2020w27
2021w1
time
Excess YLL all other indirect deaths
95%CI

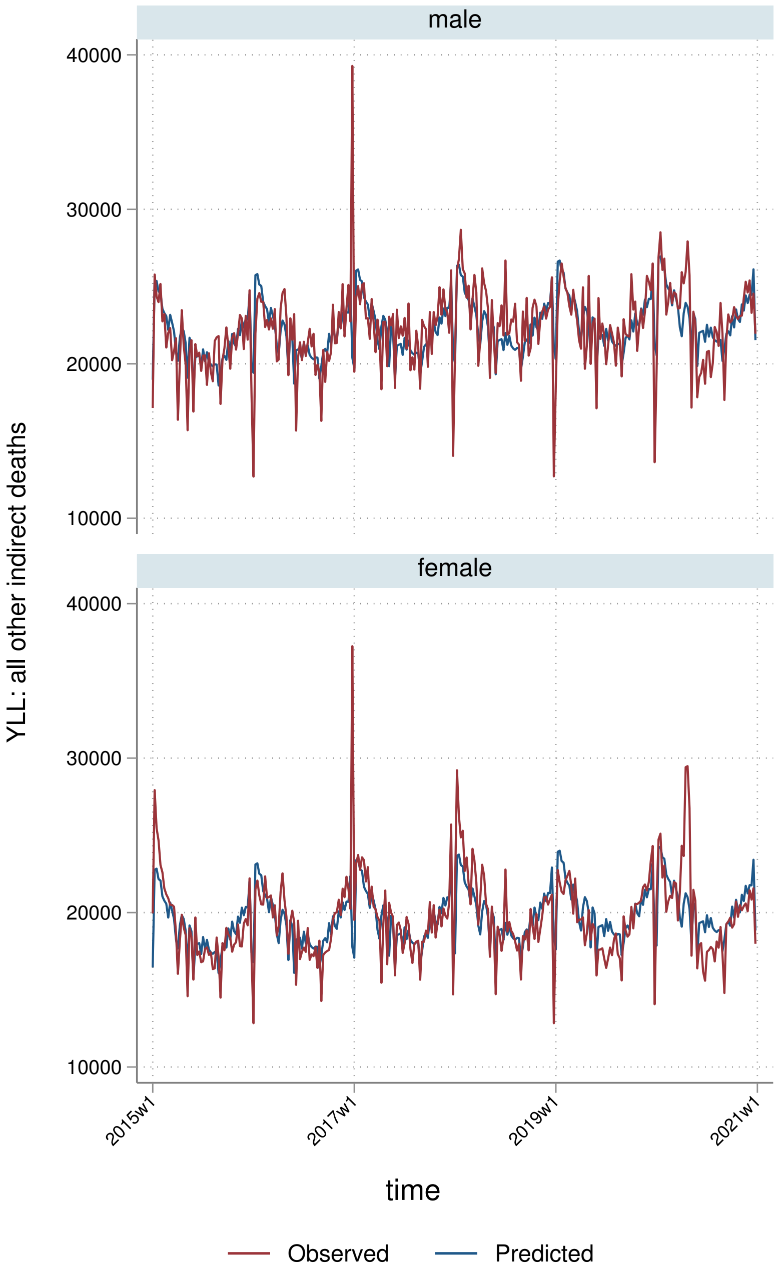

male
female
40000
30000
20000
10000
YLL: all other indirect deaths
2015w1
2017w1
2019w1
2021w1
time
Observed
Predicted

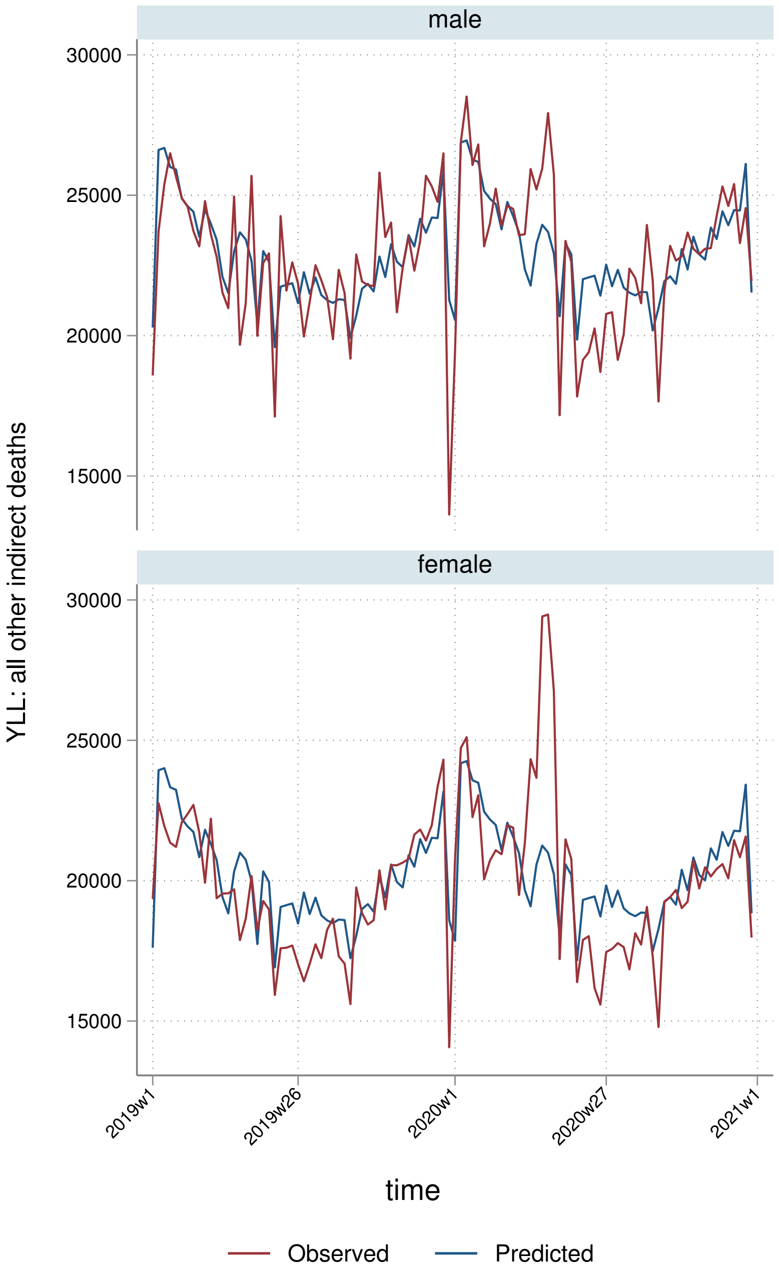
male
female
30000
25000
20000
15000
YLL: all other indirect deaths
2019w1
2019w26
2020w1
2020w27
2021w1
time
Observed
Predicted

##### 9.2.3 By deprivation quintile

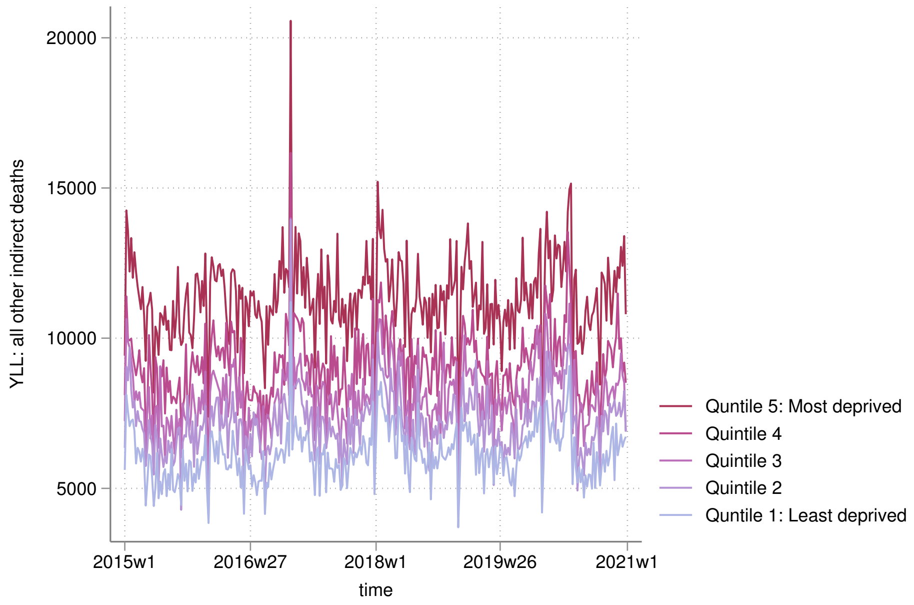


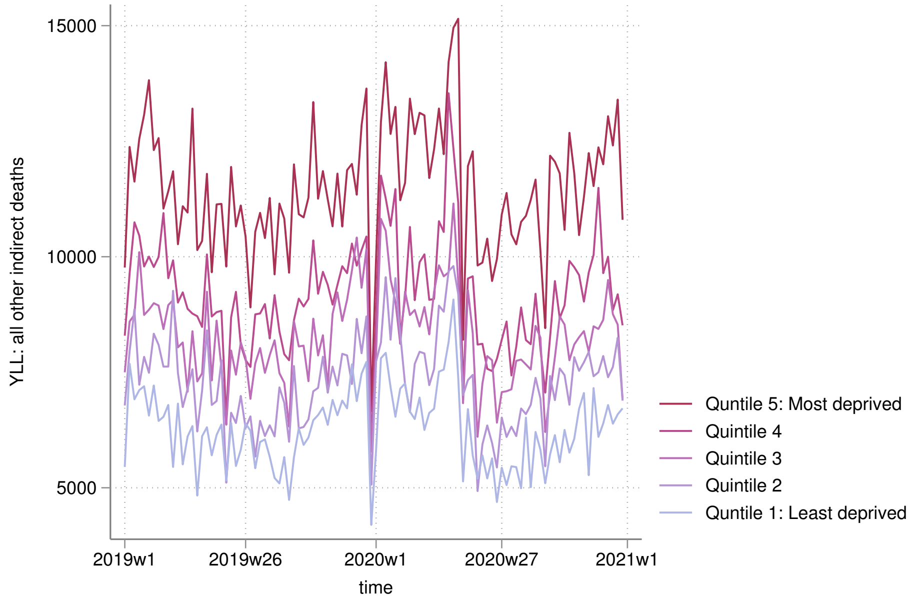

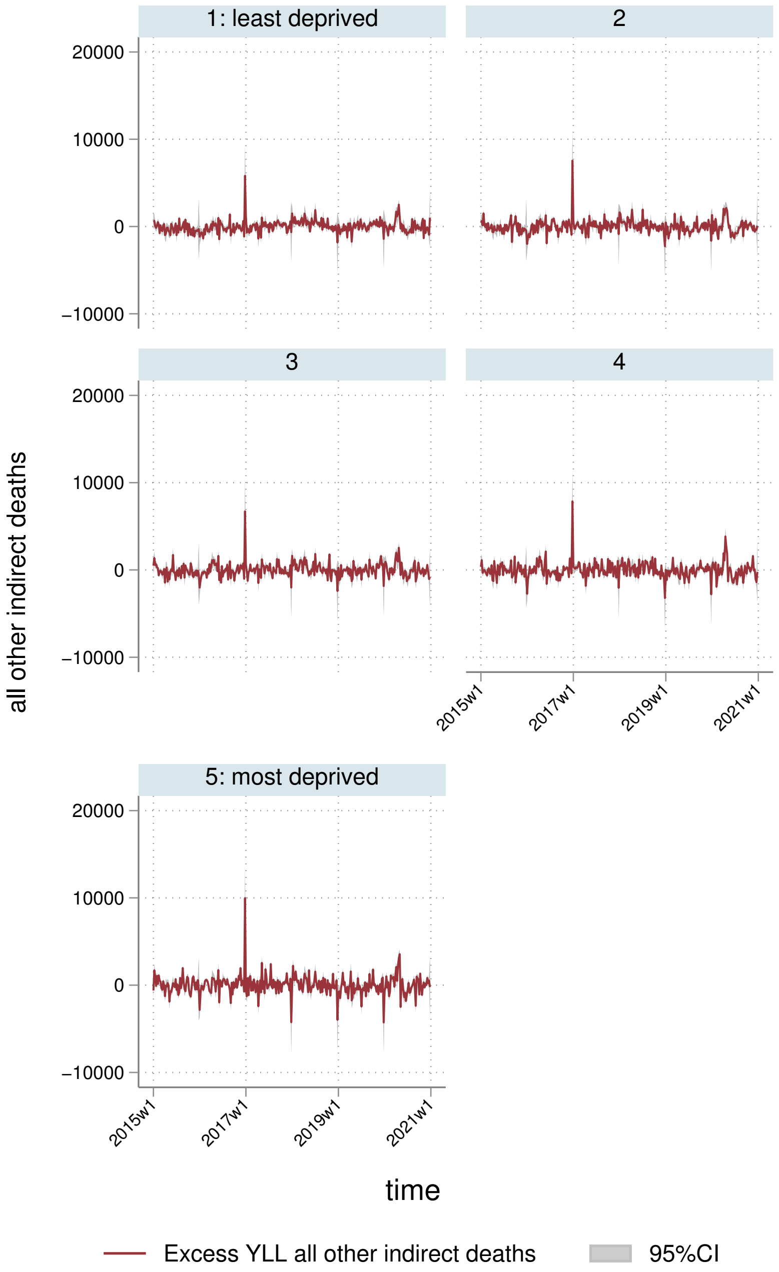
1: least deprived
2
3
4
5: most deprived
20000
10000
0
−10000
all other indirect deaths
2015w1
2017w1
2019w1
2021w1
time
Excess YLL all other indirect deaths
95%CI

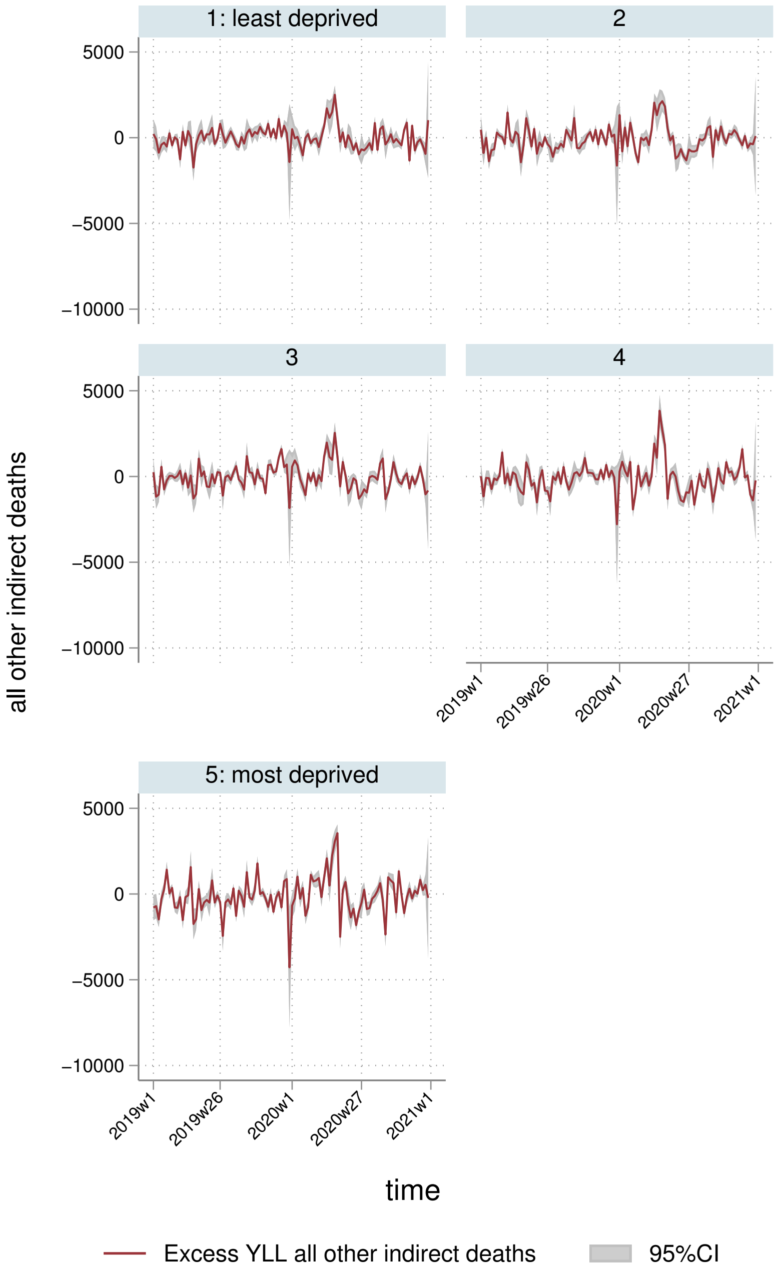

1: least deprived
2
3
4
5: most deprived
5000
0
−5000
−10000
all other indirect deaths
2019w1
2019w26
2020w1
2020w27
2021w1
time
Excess YLL all other indirect deaths
95%CI

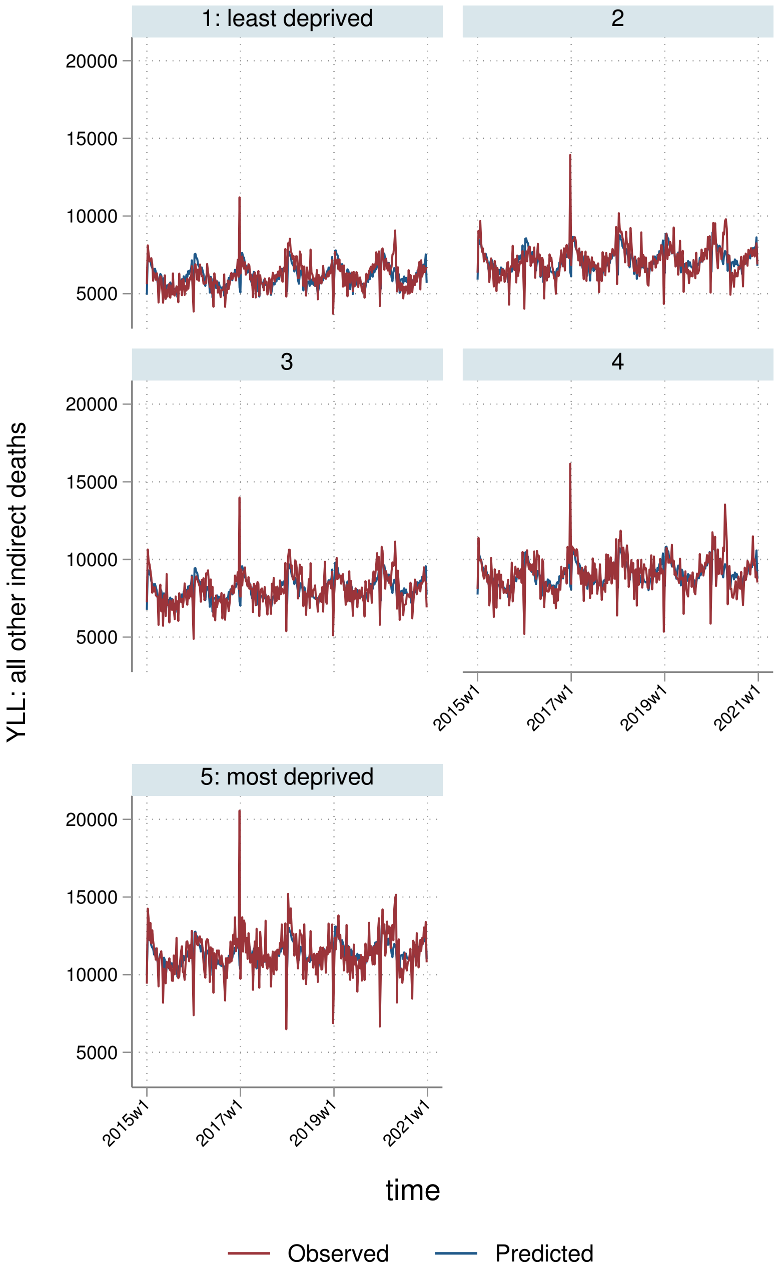

1: least deprived
2
3
4
5: most deprived
20000
15000
10000
5000
YLL: all other indirect deaths
2015w1
2017w1
2019w1
2021w1
time
Observed
Predicted

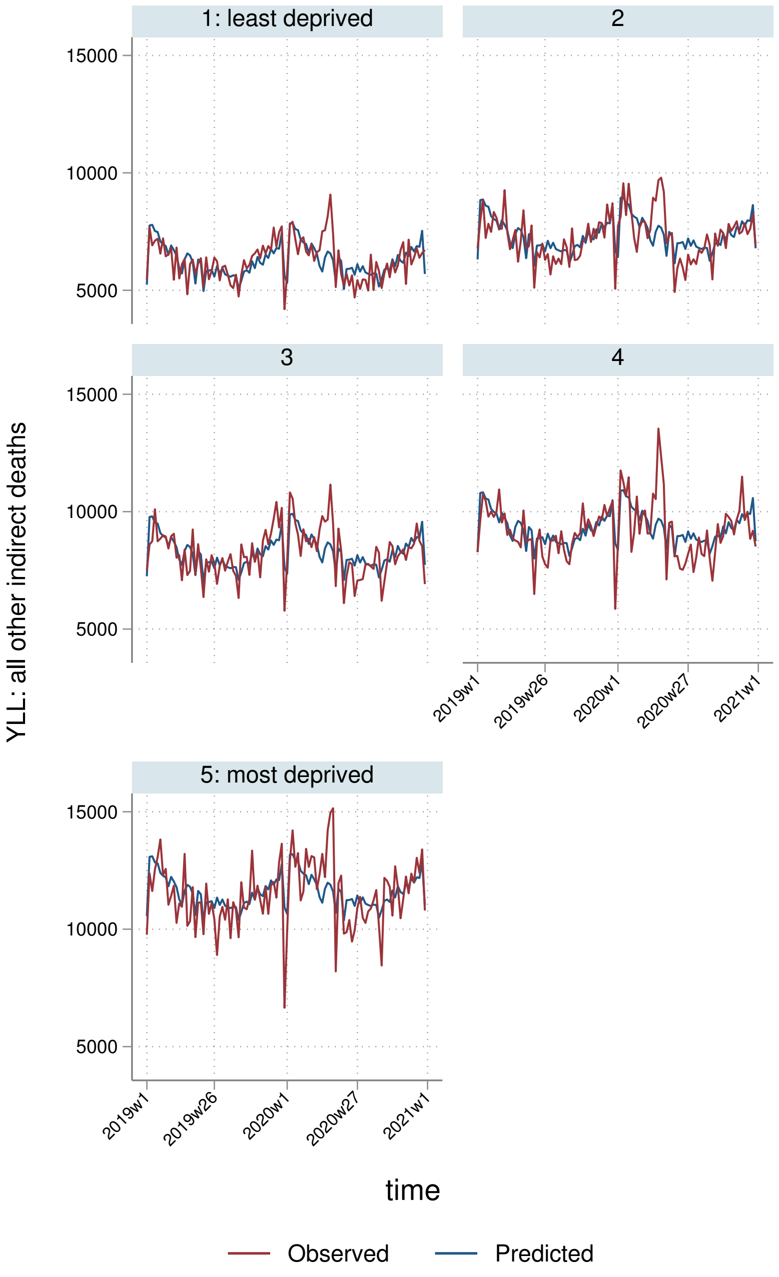
1: least deprived
2
3
4
5: most deprived
YLL: all other indirect deaths
15000
10000
5000
2019w1
2019w26
2020w1
2020w27
2021w1
time
Observed
Predicted

##### 9.2.4 By Strategic Health Authority

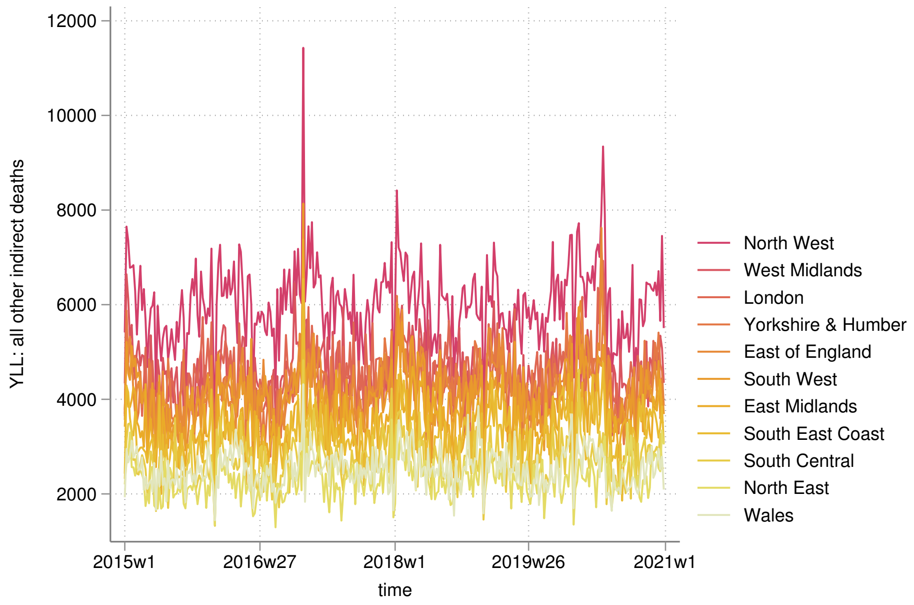


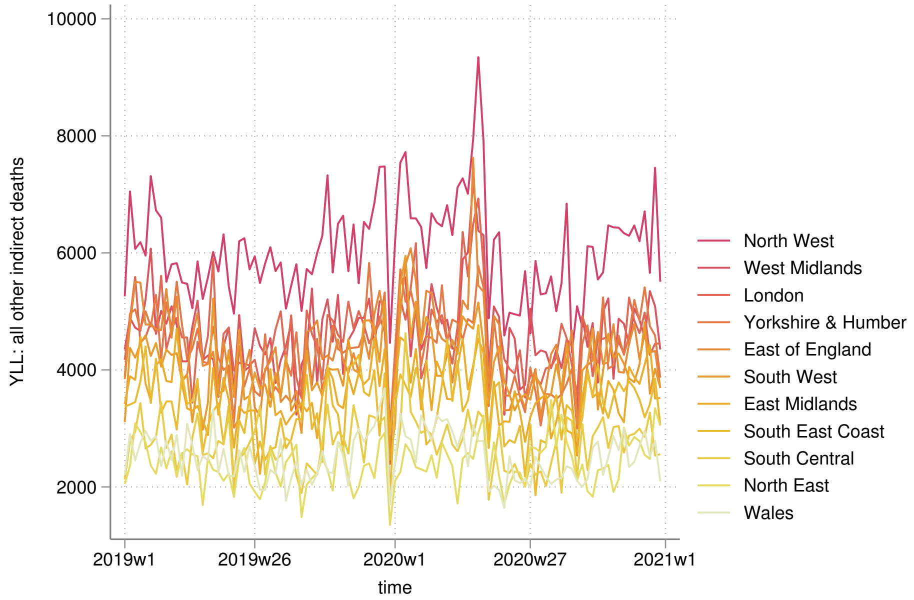

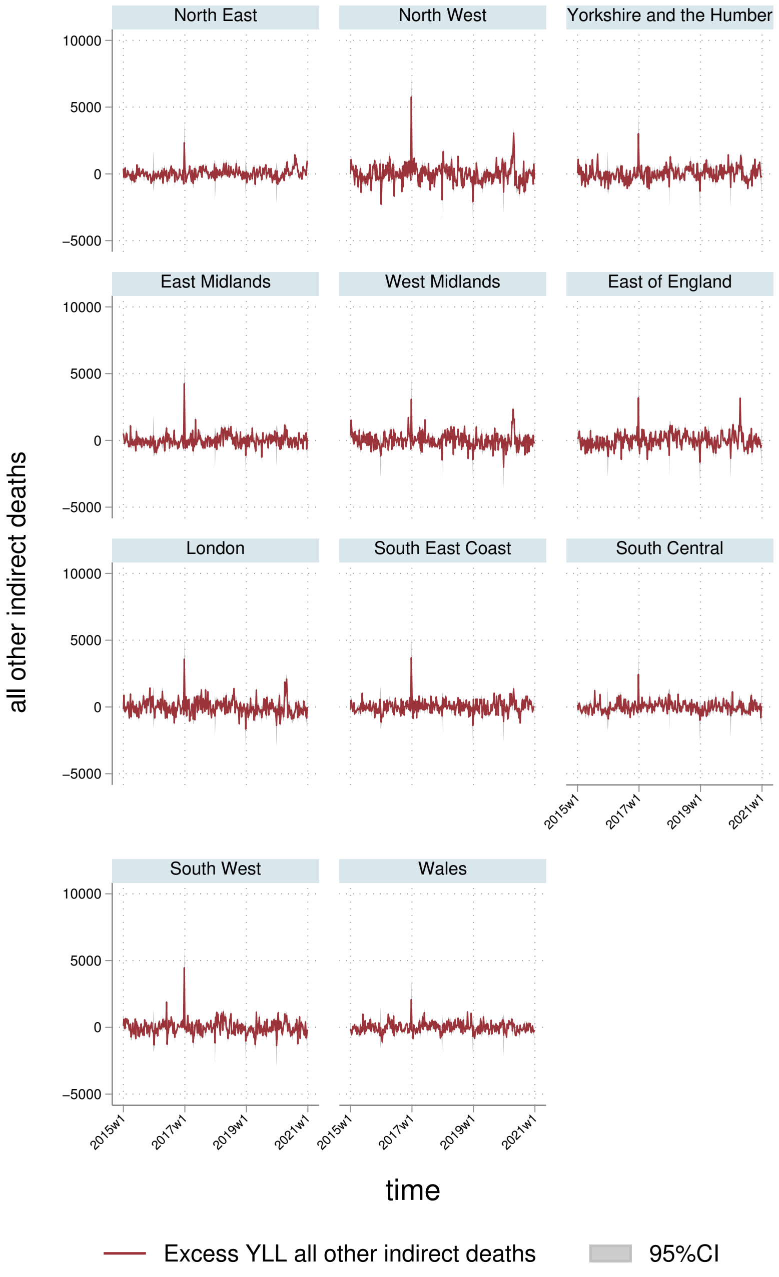
North East
North West
Yorkshire and the Humber
East Midlands
West Midlands
East of England
London
South East Coast
South Central
South West
Wales
10000
5000
0
−5000
2015w1
2017w1
2019w1
2021w1
all other indirect deaths
time
Excess YLL all other indirect deaths
95%CI

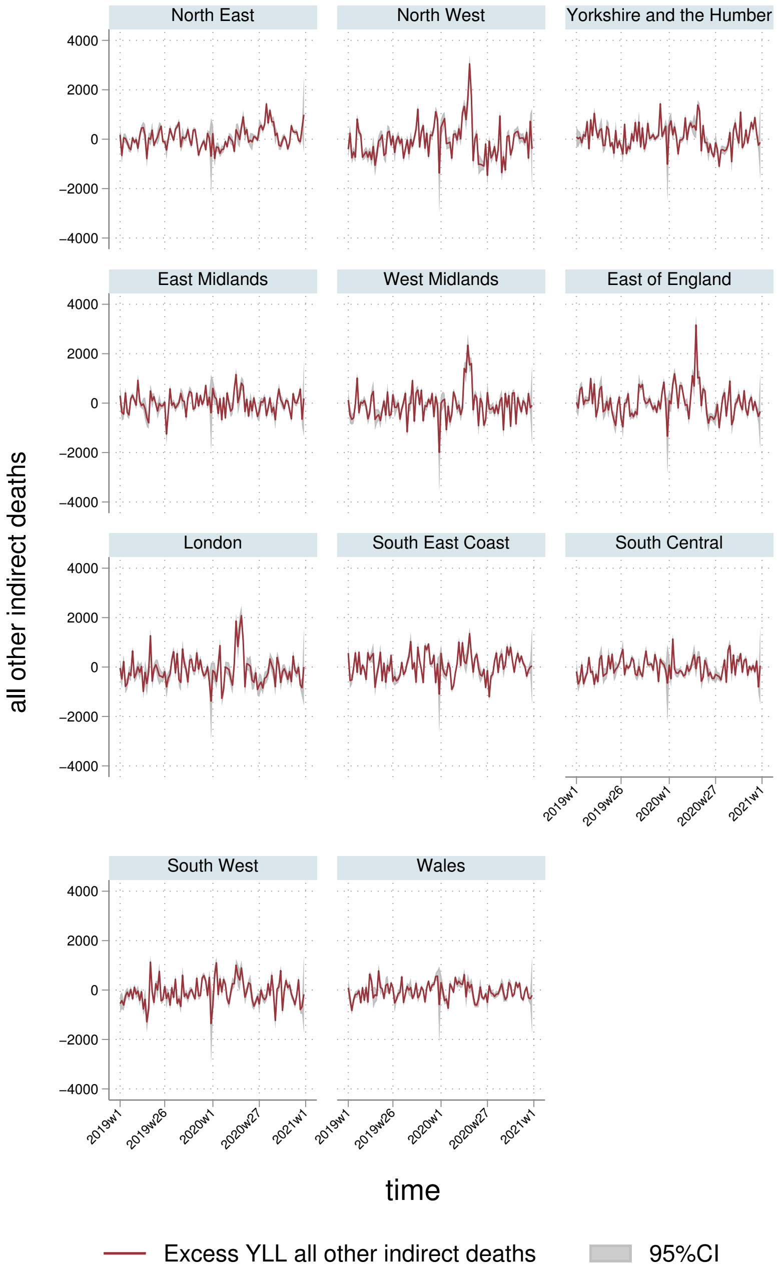

North East
North West
Yorkshire and the Humber
East Midlands
West Midlands
East of England
London
South East Coast
South Central
South West
Wales
4000
2000
0
−2000
−4000
all other indirect deaths
2019w1
2019w26
2020w1
2020w27
2021w1
time
Excess YLL all other indirect deaths
95%CI

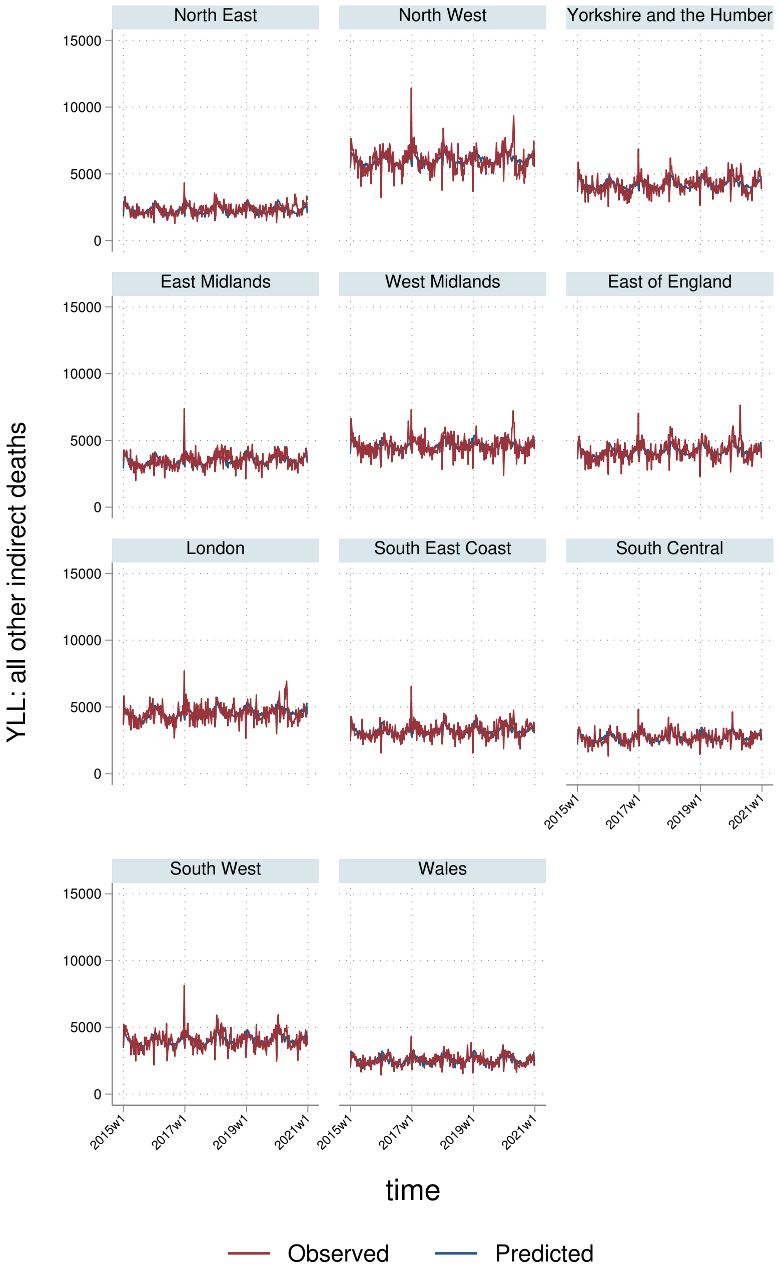
North East
North West
Yorkshire and the Humber
East Midlands
West Midlands
East of England
London
South East Coast
South Central
South West
Wales
YLL: all other indirect deaths
15000
10000
5000
0
2015w1
2017w1
2019w1
2021w1
time
Observed
Predicted

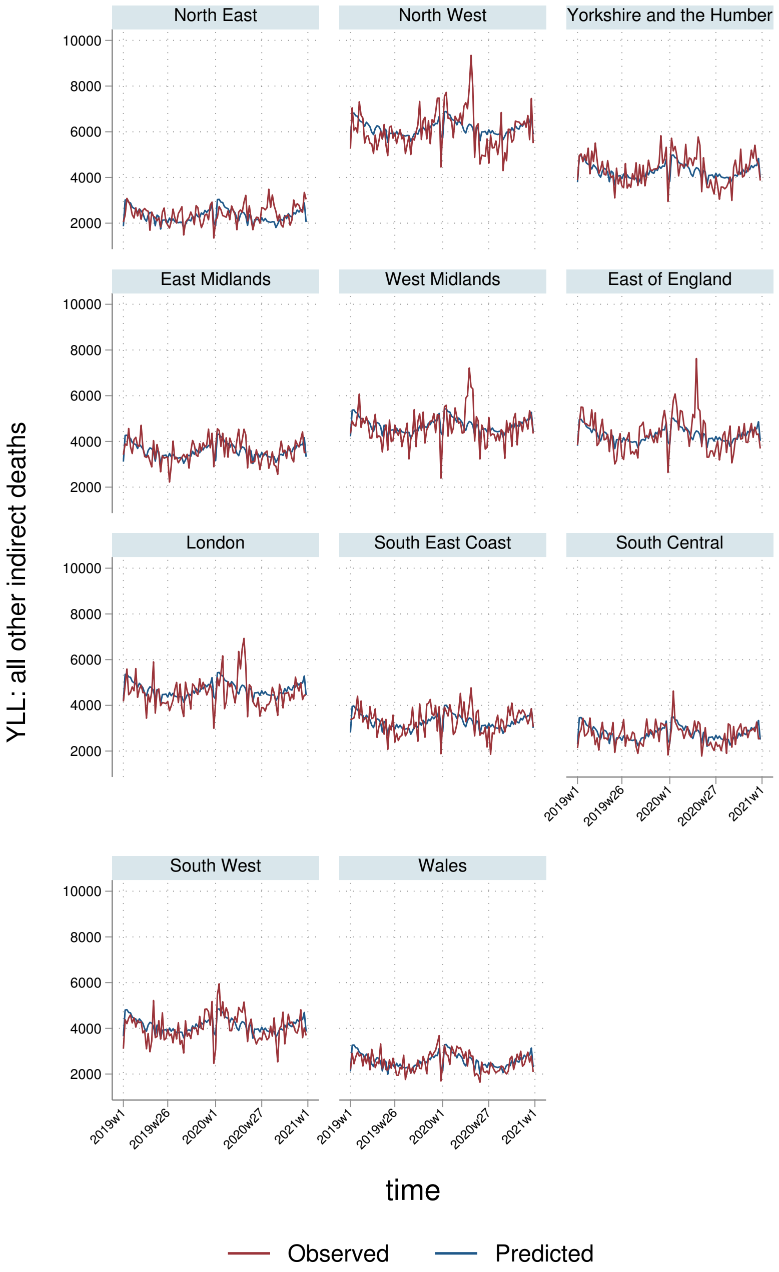

North East
North West
Yorkshire and the Humber
East Midlands
West Midlands
East of England
London
South East Coast
South Central
South West
Wales
10000
8000
6000
4000
2000
YLL: all other indirect deaths
2019w1
2019w26
2020w1
2020w27
2021w1
time
Observed
Predicted

#### 9.3 YLLs per $100,000$ population

##### 9.3.1 England-Wales aggregate

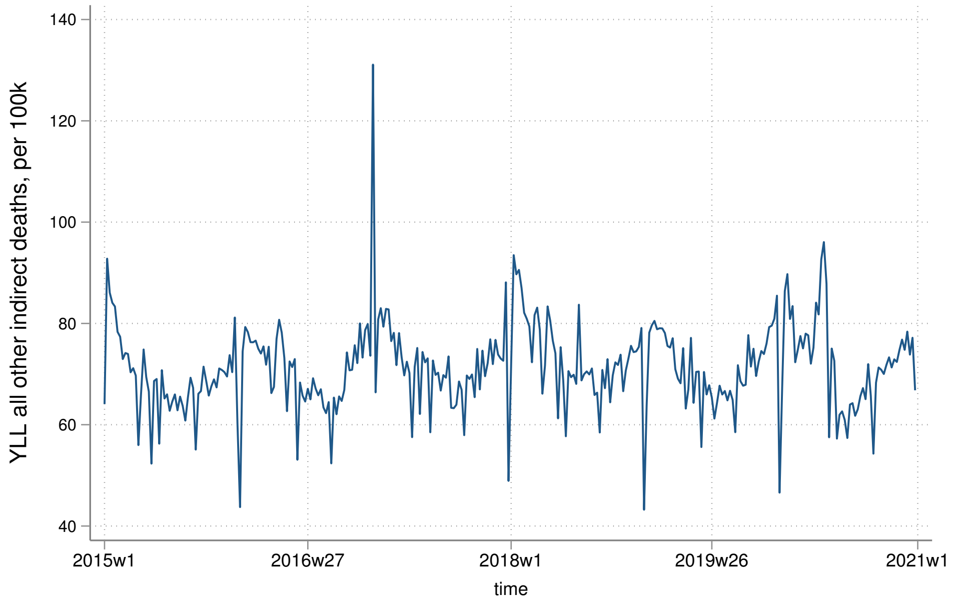


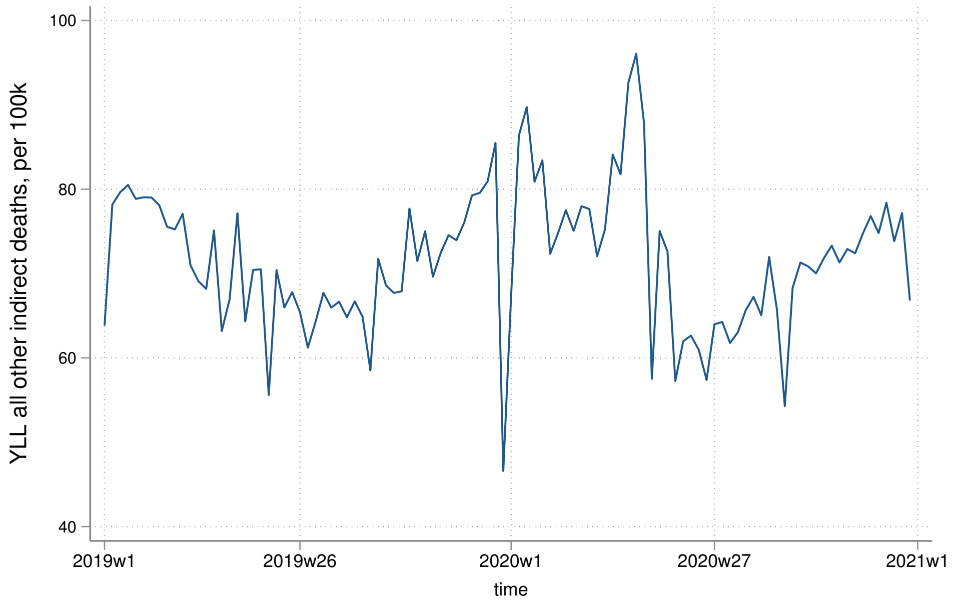

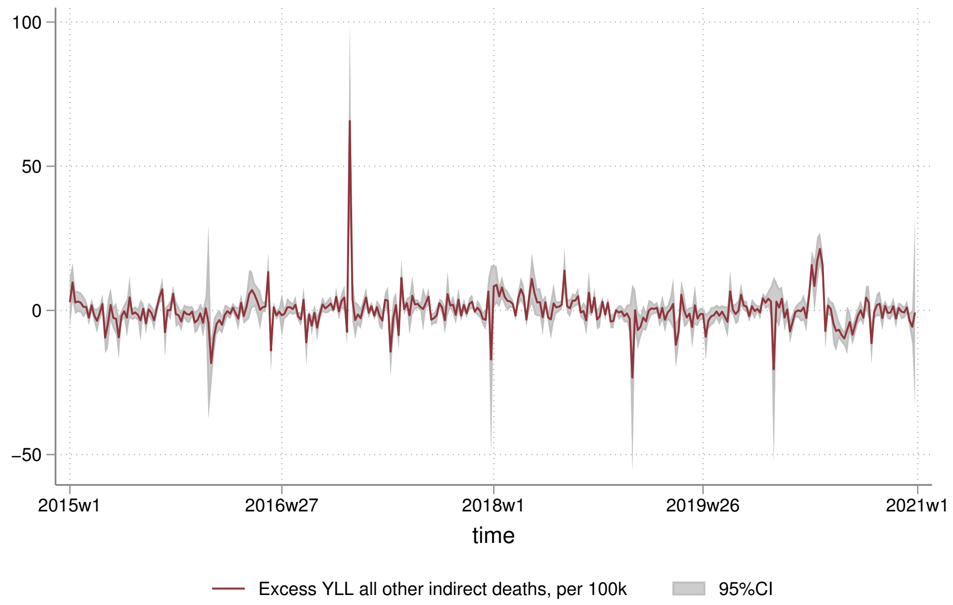
100
50
0
−50
2015w1
2016w27
2018w1
2019w26
2021w1
time
Excess YLL all other indirect deaths, per 100k
95%CI

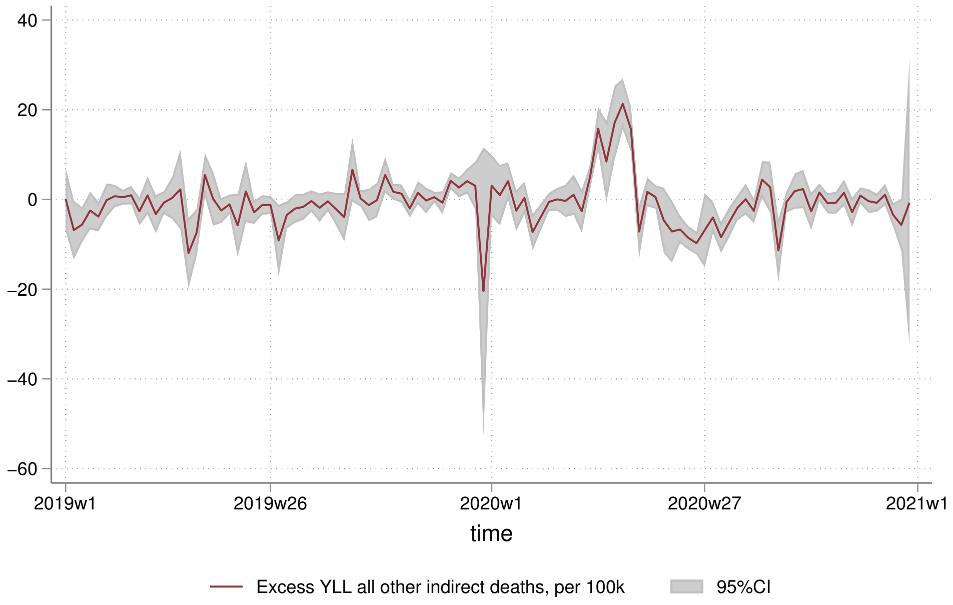
40
20
0
−20
−40
−60
2019w1
2019w26
2020w1
2020w27
2021w1
time
Excess YLL all other indirect deaths, per 100k
95%CI

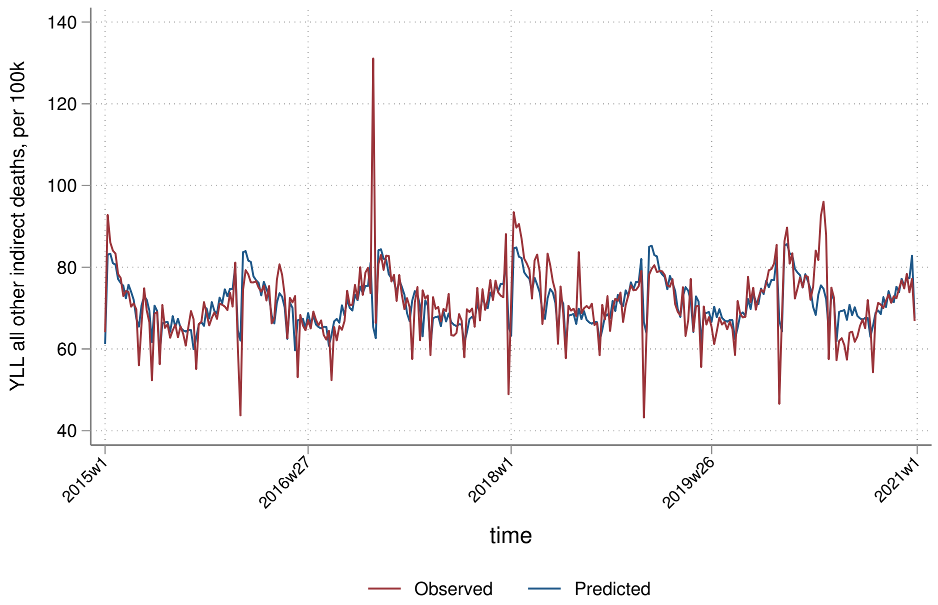

YLL all other indirect deaths, per 100k
140
120
100
80
60
40
2015w1
2016w27
2018w1
2019w26
2021w1
time
Observed
Predicted


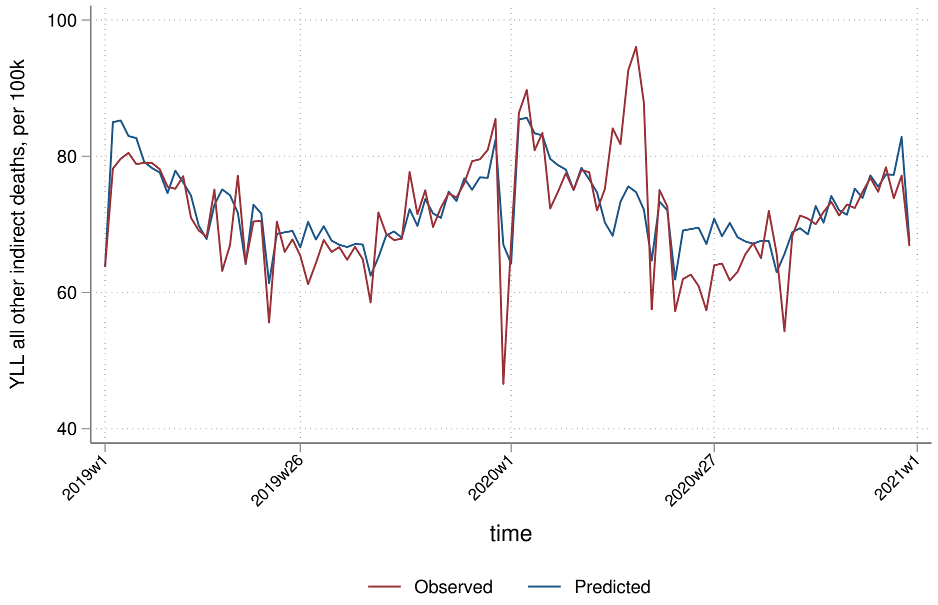

YLL all other indirect deaths, per 100k
100
80
60
40
2019w1
2019w26
2020w1
2020w27
2021w1
time
Observed
Predicted

##### 9.3.2 By sex

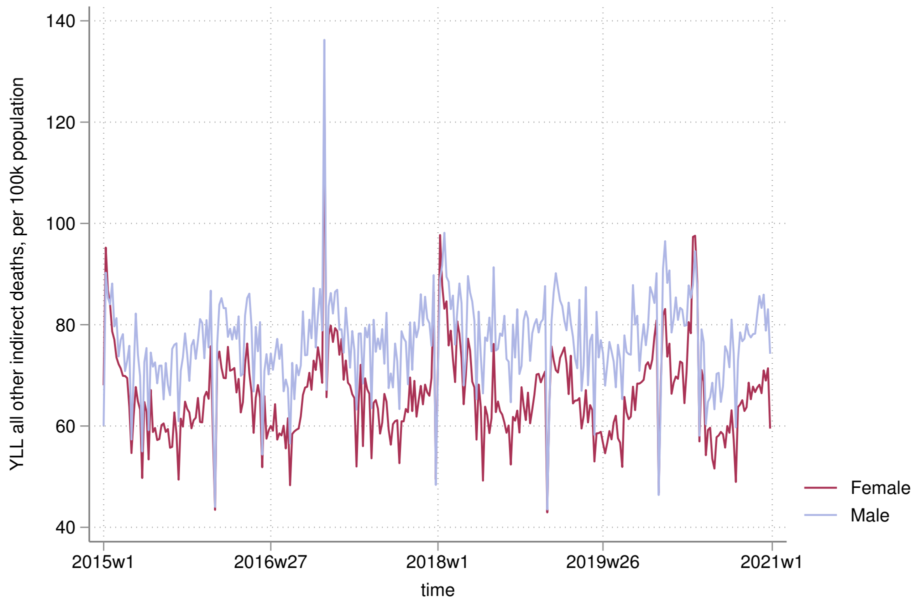
140
120
100
80
60
40
YLL all other indirect deaths, per 100k population
2015w1
2016w27
2018w1
2019w26
2021w1
time
Female
Male

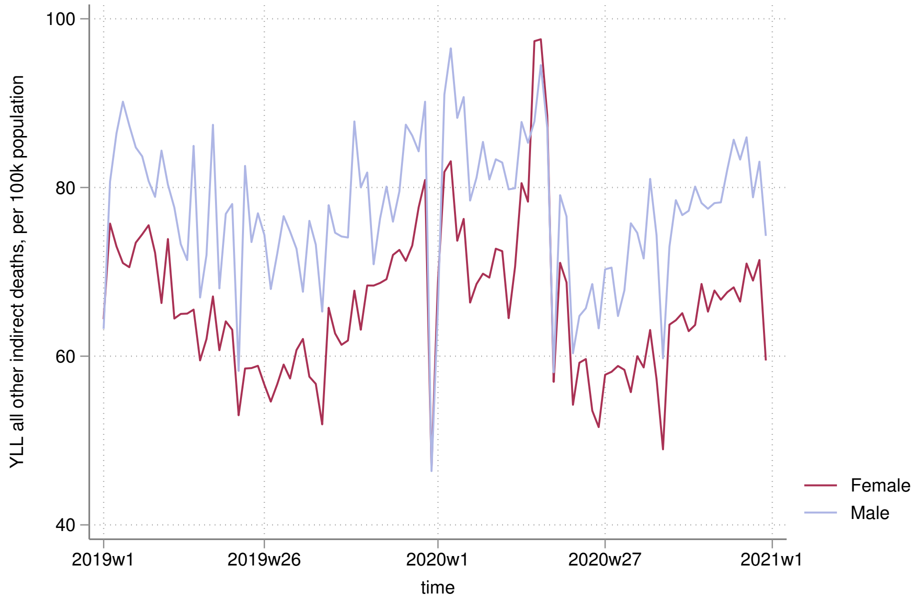
100
80
60
40
YLL all other indirect deaths, per 100k population
2019w1
2019w26
2020w1
2020w27
2021w1
time
Female
Male

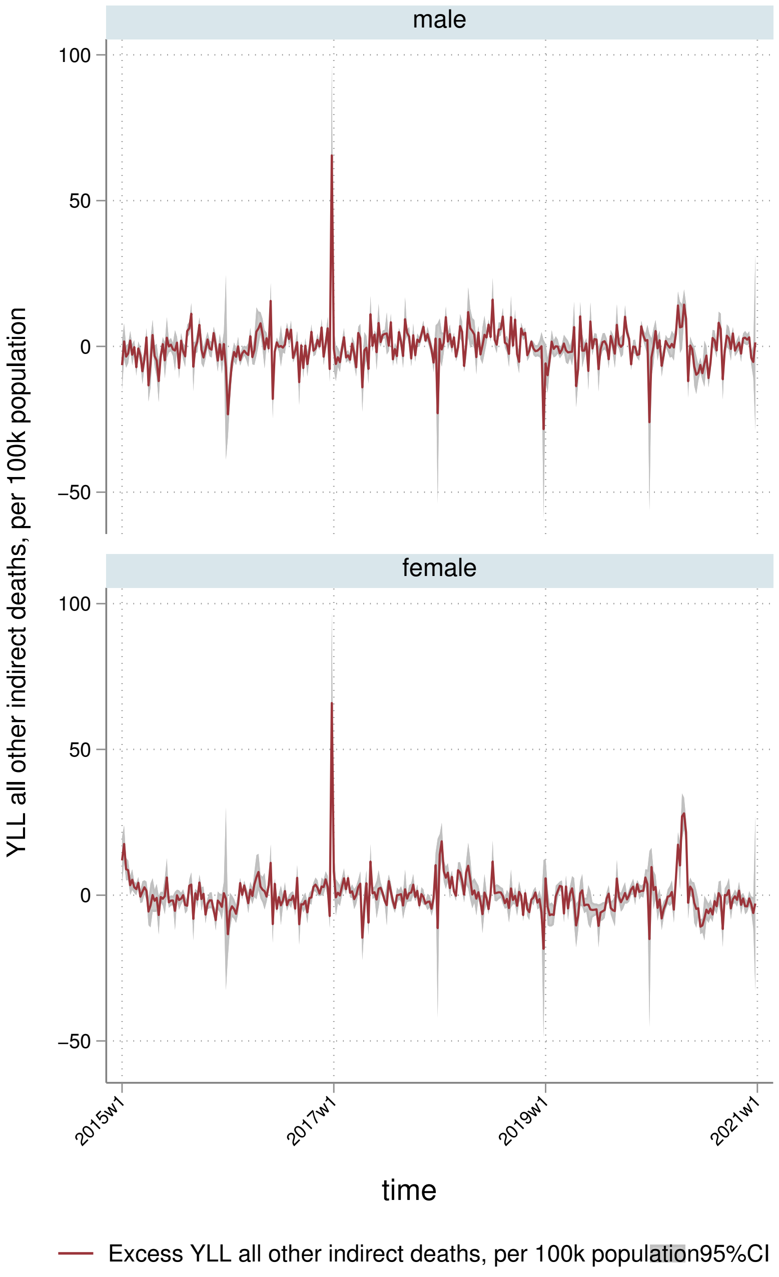
male
female
100
50
0
−50
YLL all other indirect deaths, per 100k population
2015w1
2017w1
2019w1
2021w1
time
Excess YLL all other indirect deaths, per 100k population
95%CI

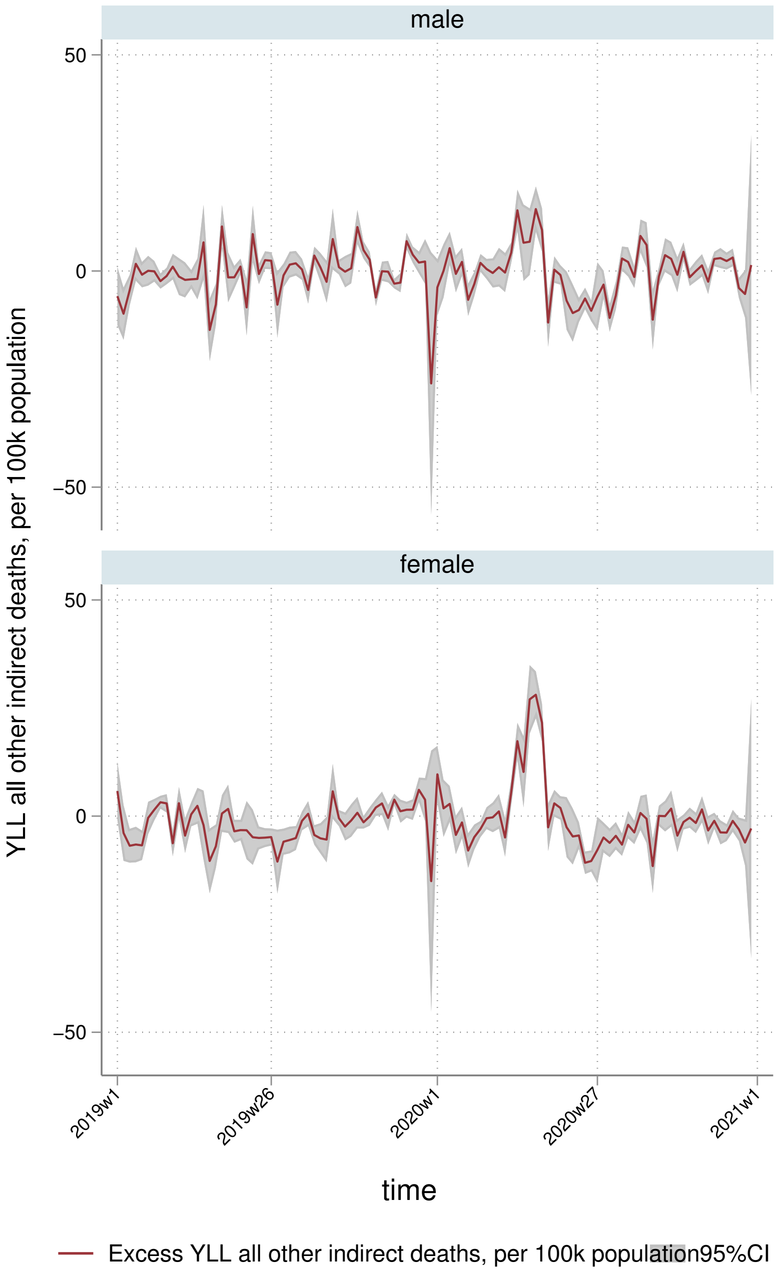
male
female
50
0
−50
50
0
−50
YLL all other indirect deaths, per 100k population
2019w1
2019w26
2020w1
2020w27
2021w1
time
Excess YLL all other indirect deaths, per 100k population
95%CI

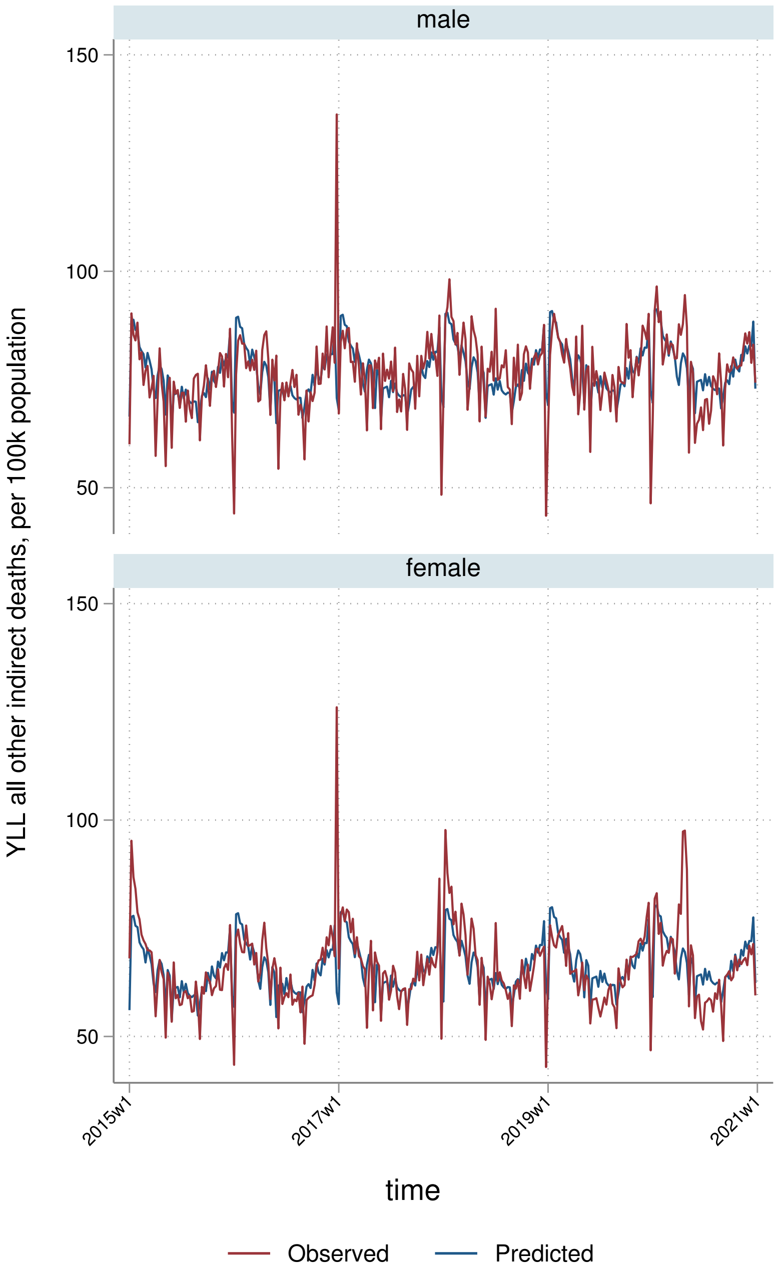

male
female
150
100
50
150
100
50
YLL all other indirect deaths, per 100k population
2015w1
2017w1
2019w1
2021w1
time
Observed
Predicted

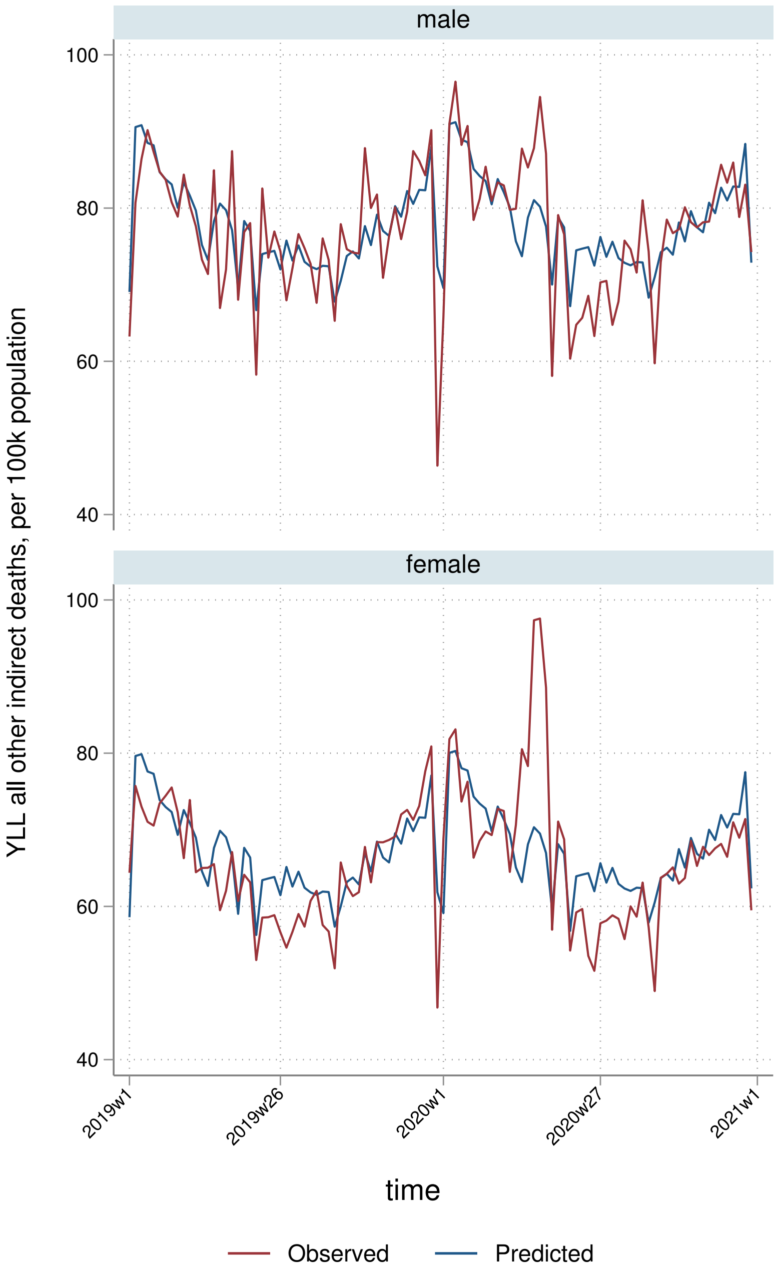

male
female
100
80
60
40
YLL all other indirect deaths, per 100k population
2019w1
2019w26
2020w1
2020w27
2021w1
time
Observed
Predicted

##### 9.3.3 By deprivation quintile

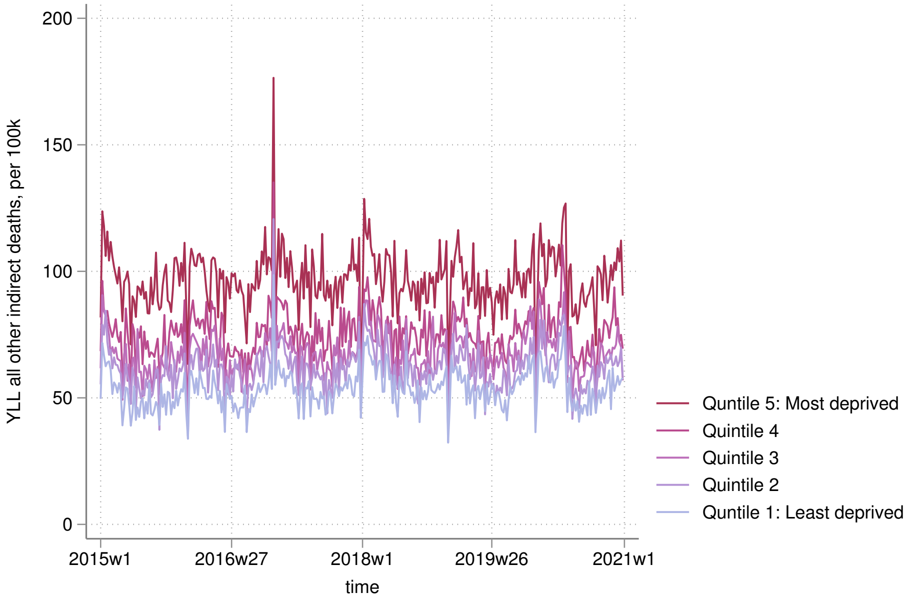


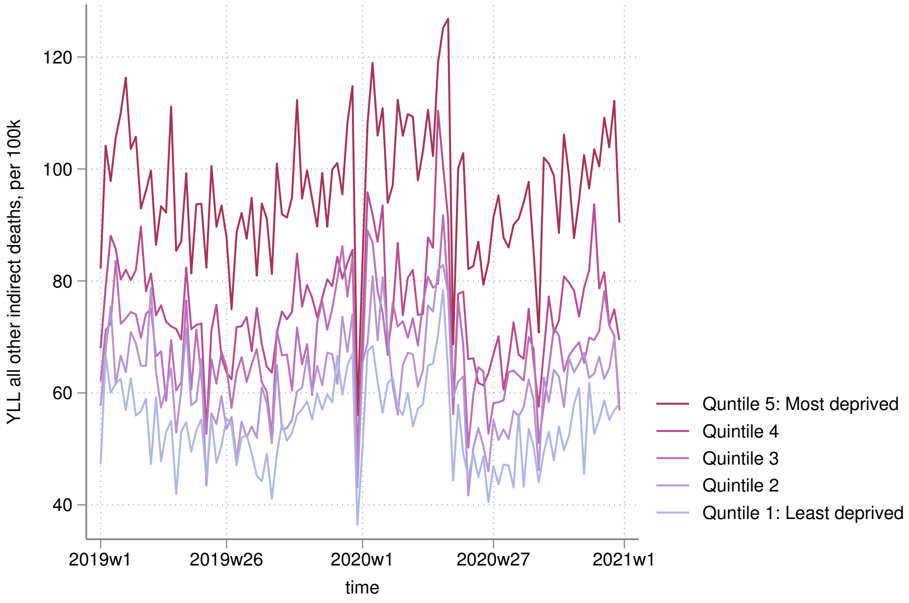

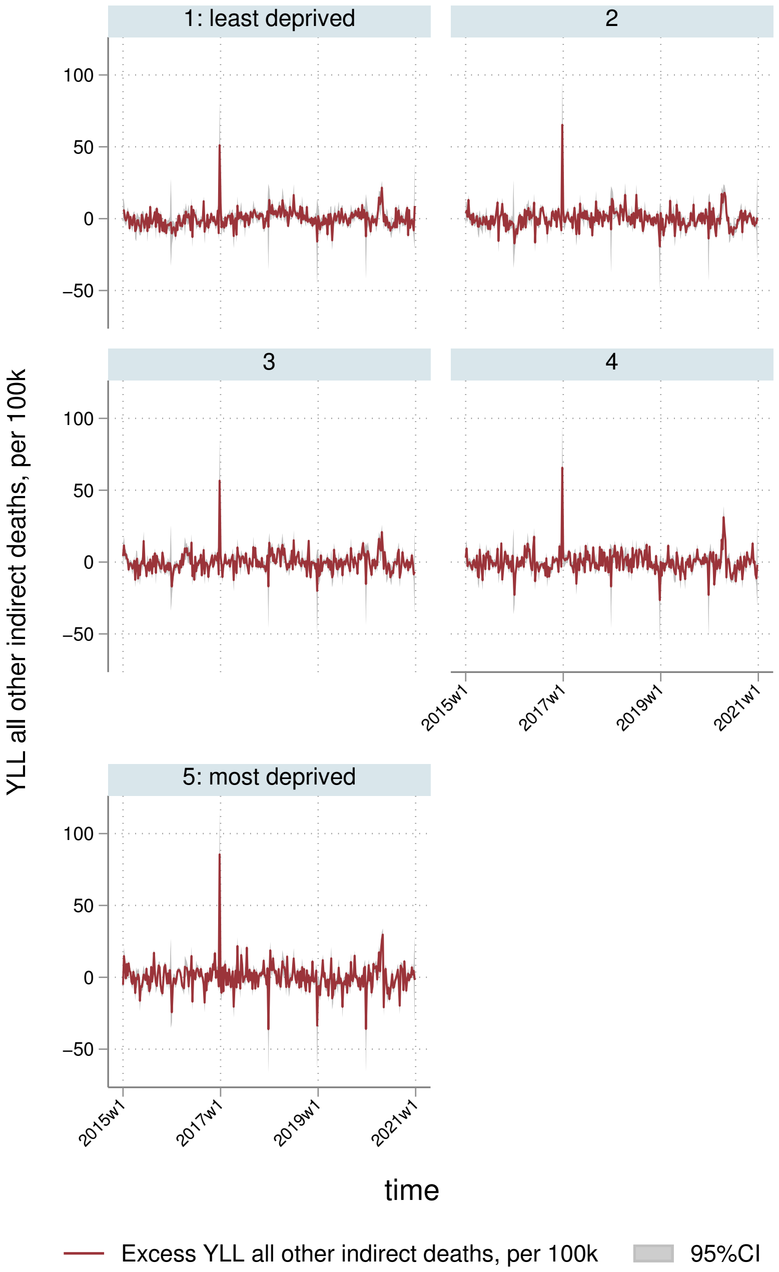

1: least deprived
2
3
4
5: most deprived
100
50
0
−50
2015w1
2017w1
2019w1
2021w1
YLL all other indirect deaths, per 100k
time
Excess YLL all other indirect deaths, per 100k
95%CI

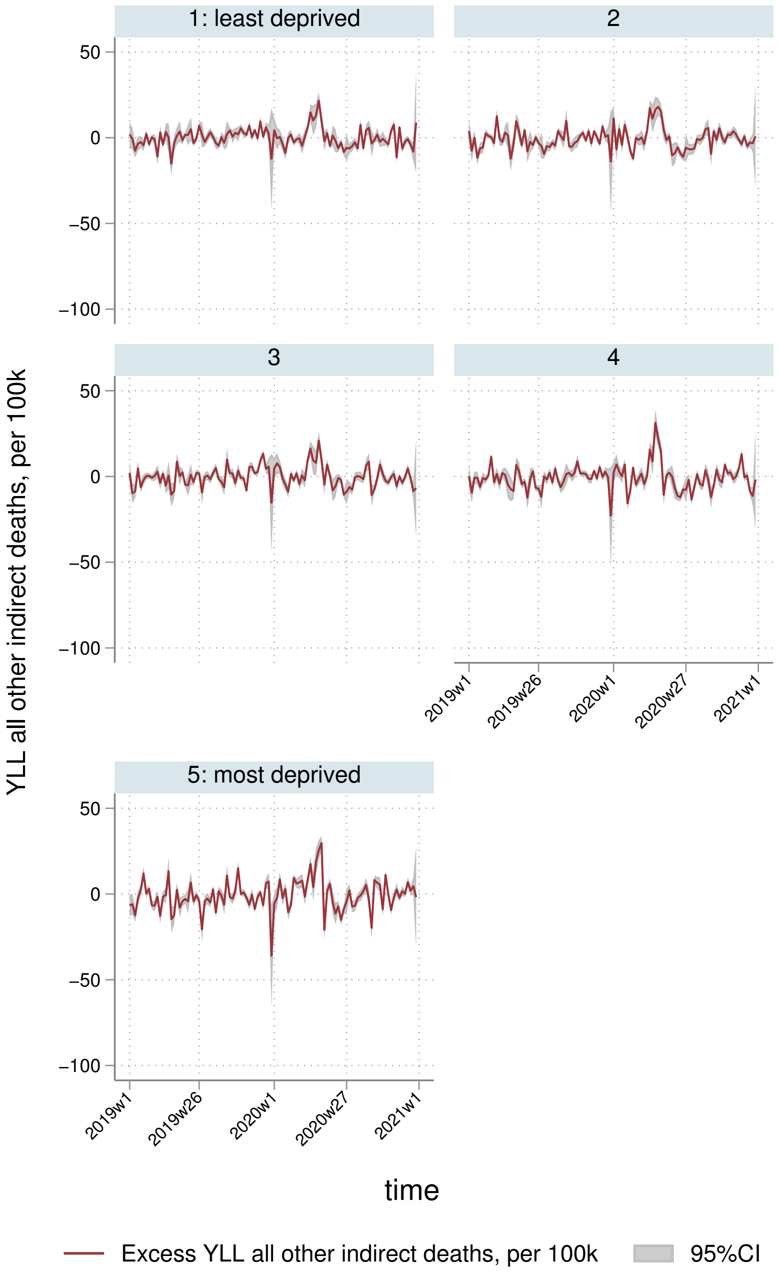
1: least deprived
2
3
4
5: most deprived
50
0
−50
−100
YLL all other indirect deaths, per 100k
2019w1
2019w26
2020w1
2020w27
2021w1
time
Excess YLL all other indirect deaths, per 100k
95%CI

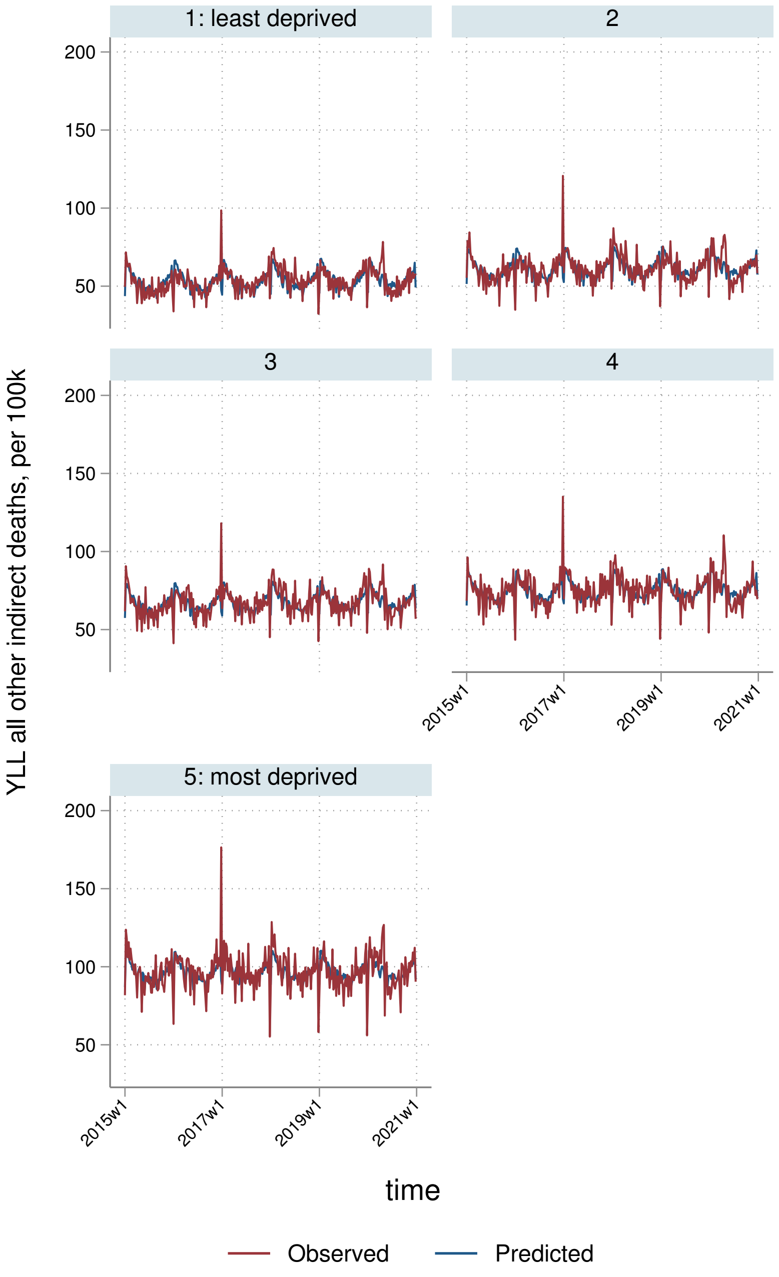

1: least deprived
2
3
4
5: most deprived
200
150
100
50
2015w1
2017w1
2019w1
2021w1
YLL all other indirect deaths, per 100k
time
Observed
Predicted

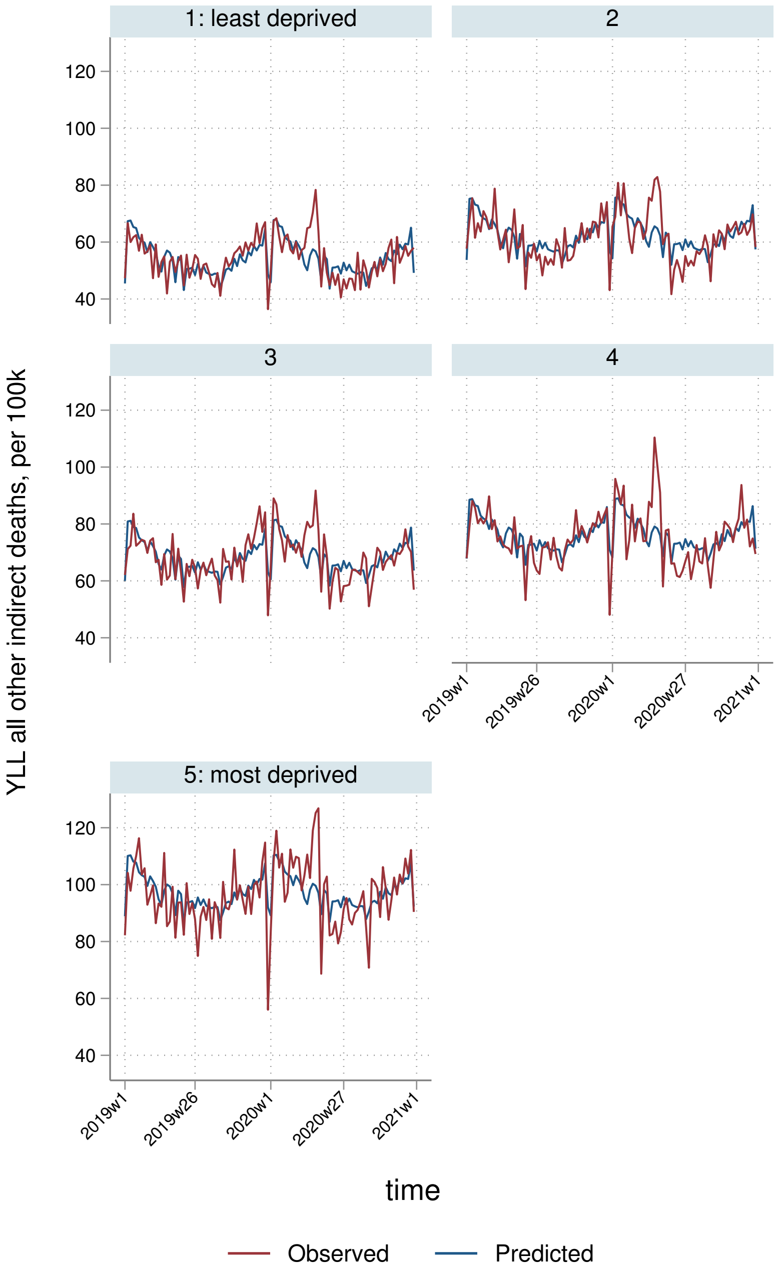
1: least deprived
2
3
4
5: most deprived
YLL all other indirect deaths, per 100k
120
100
80
60
40
2019w1
2019w26
2020w1
2020w27
2021w1
time
Observed
Predicted

##### 9.3.4 By Strategic Health Authority

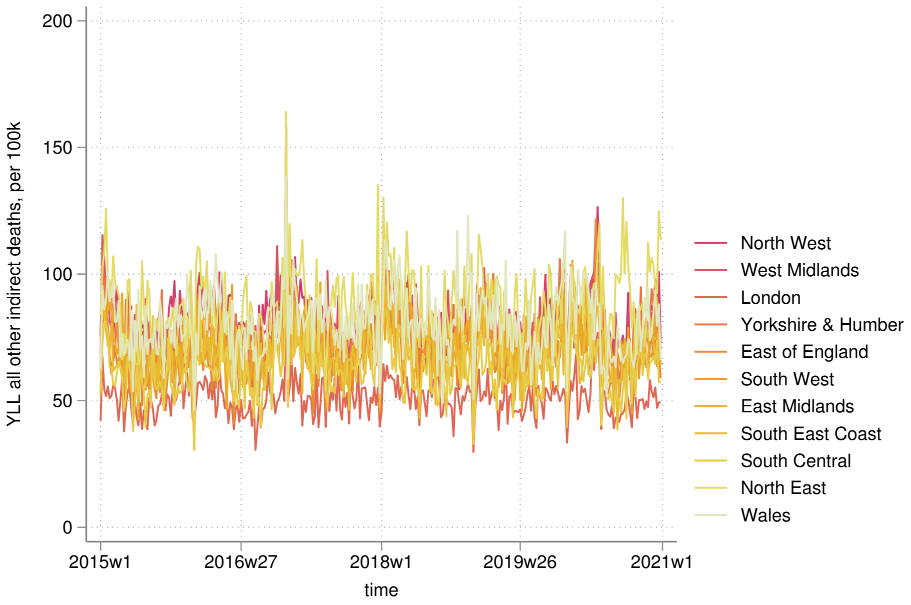


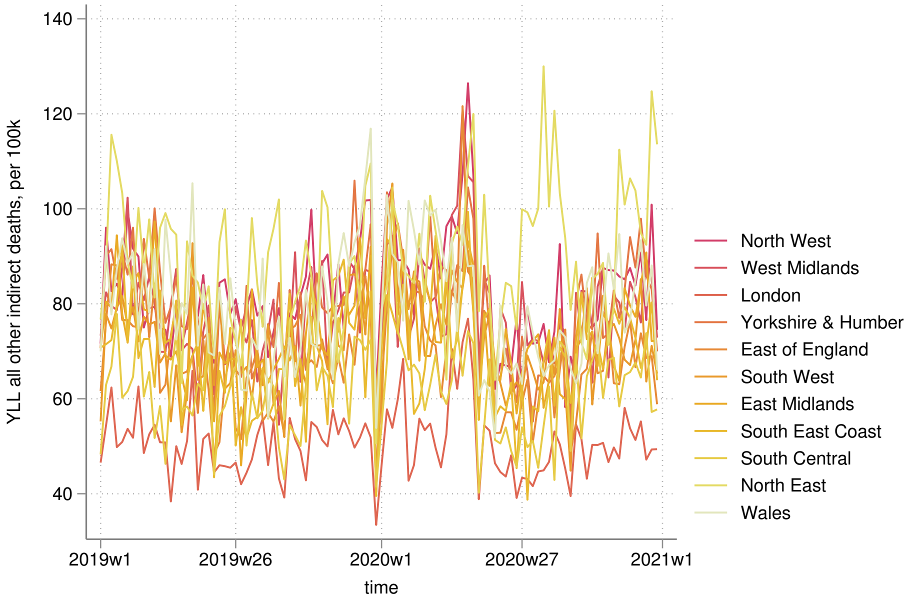

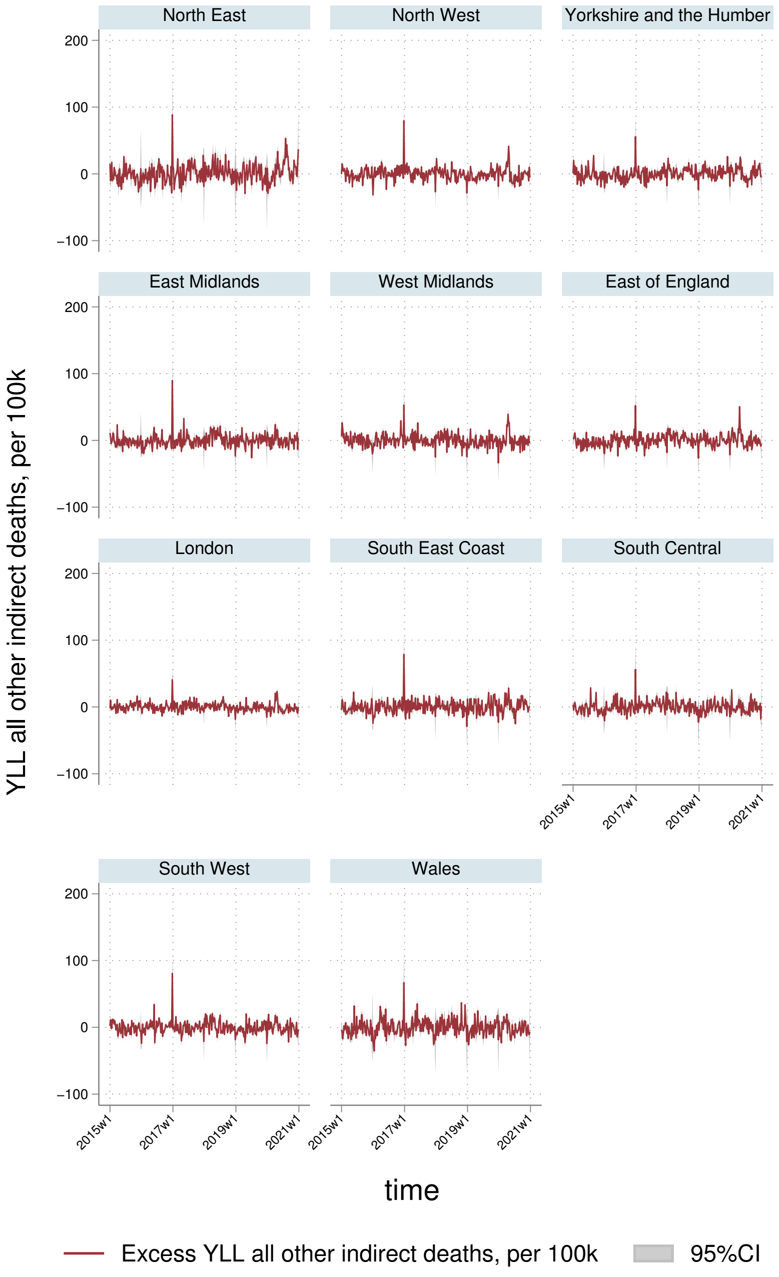
North East
North West
Yorkshire and the Humber
East Midlands
West Midlands
East of England
London
South East Coast
South Central
South West
Wales
200
100
0
−100
2015w1
2017w1
2019w1
2021w1
YLL all other indirect deaths, per 100k
time
Excess YLL all other indirect deaths, per 100k
95%CI

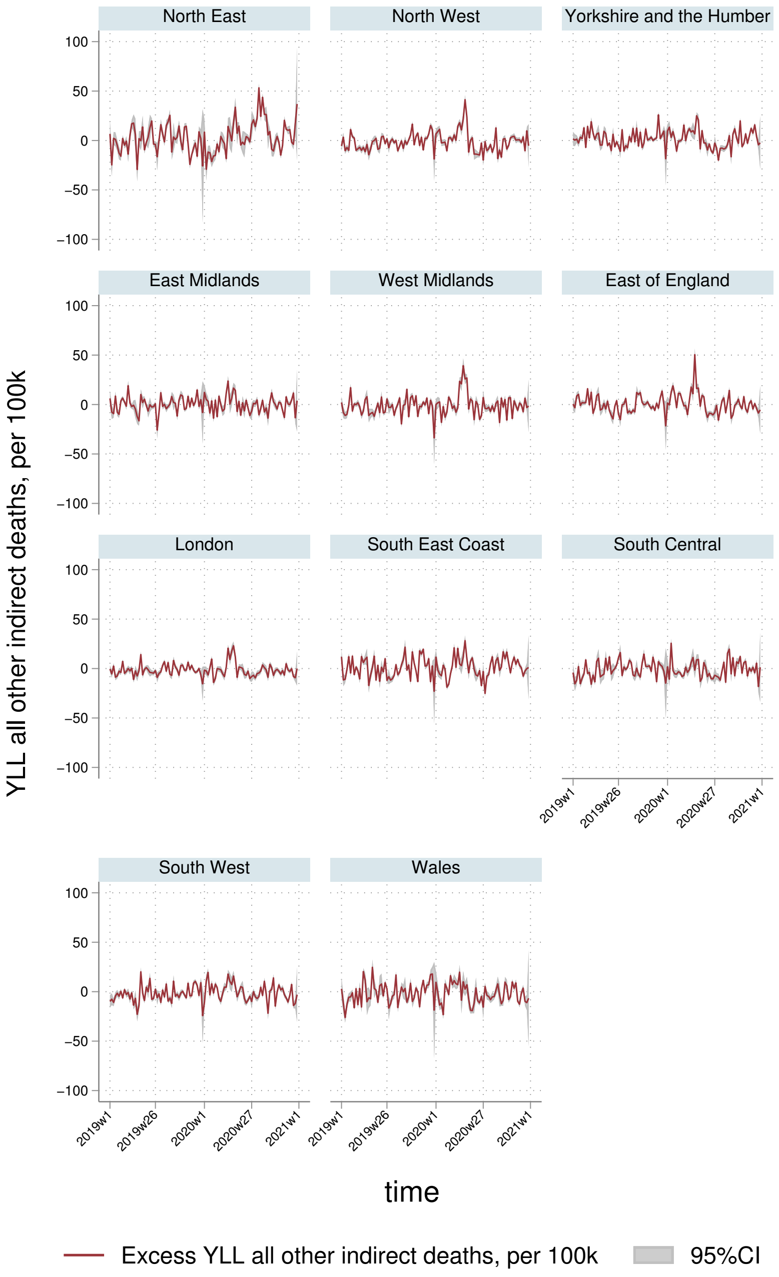

North East
North West
Yorkshire and the Humber
East Midlands
West Midlands
East of England
London
South East Coast
South Central
South West
Wales
YLL all other indirect deaths, per 100k
100
50
0
−50
−100
2019w1
2019w26
2020w1
2020w27
2021w1
time
Excess YLL all other indirect deaths, per 100k
95%CI

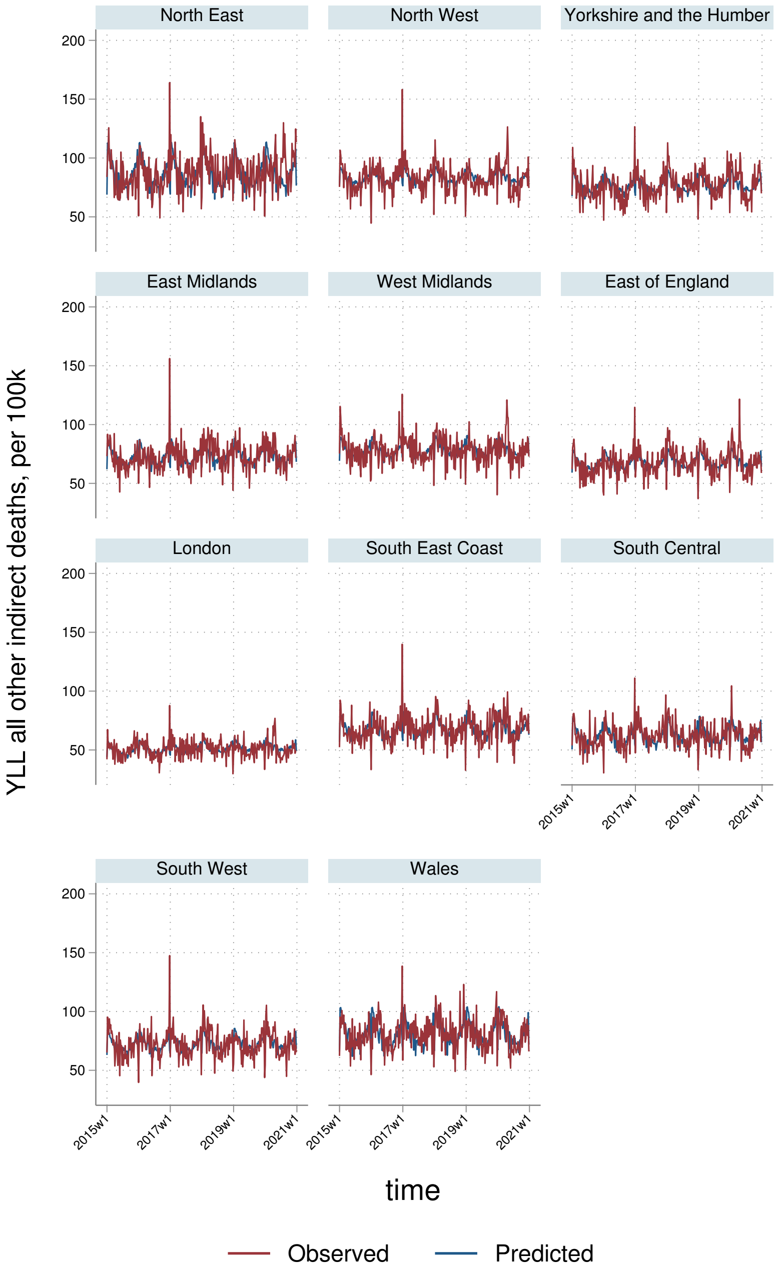
North East
North West
Yorkshire and the Humber
East Midlands
West Midlands
East of England
London
South East Coast
South Central
South West
Wales
YLL all other indirect deaths, per 100k
200
150
100
50
2015w1
2017w1
2019w1
2021w1
time
Observed
Predicted

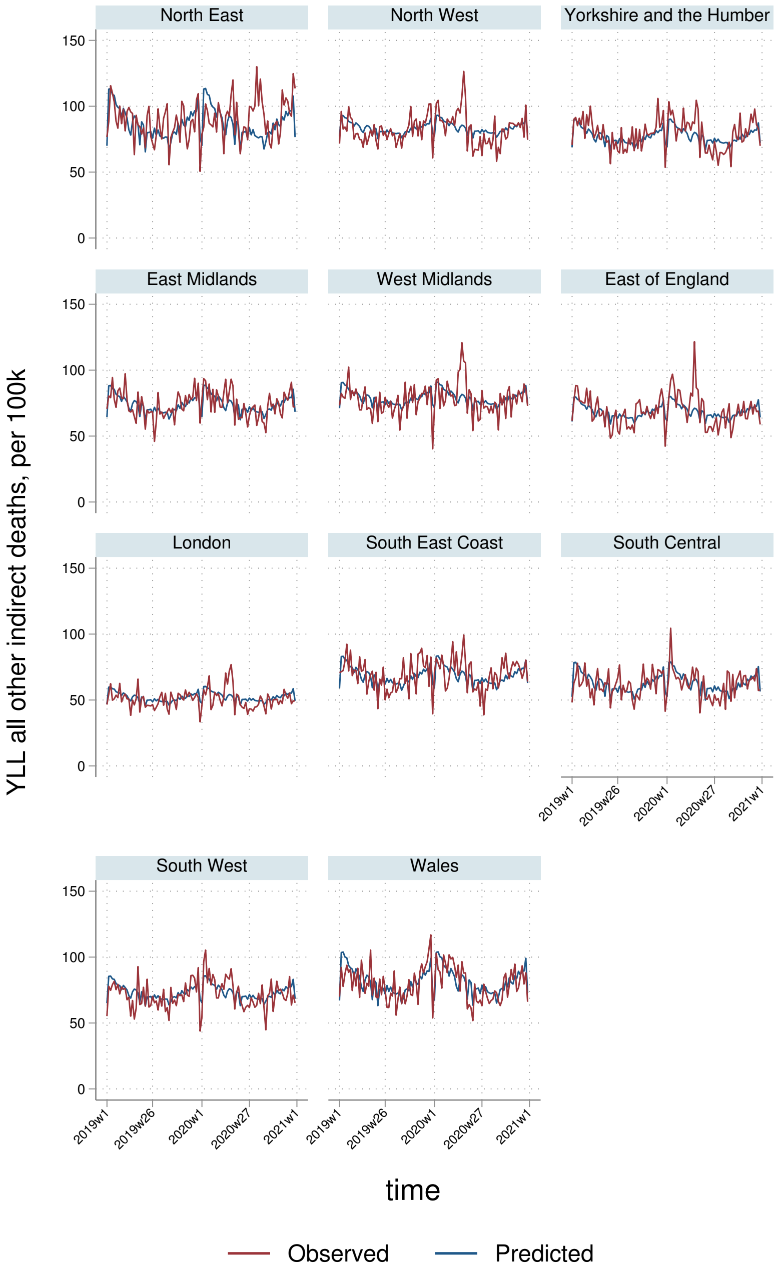
North East
North West
Yorkshire and the Humber
East Midlands
West Midlands
East of England
London
South East Coast
South Central
South West
Wales
YLL all other indirect deaths, per 100k
0
50
100
150
2019w1
2019w26
2020w1
2020w27
2021w1
time
Observed
Predicted
